# Striational Antibody-Associated Myositis – Bridging the Gap between Thymoma and Myasthenia Gravis: A Systematic Review

**DOI:** 10.64898/2026.01.27.25340404

**Authors:** Jie Luo, Jilei Lin, John Shymansky, Huixia Judy Wang

**Author notes:** Correspondence to: J Luo.

## Abstract

An overlap syndrome of myositis and/or myocarditis associated with myasthenia gravis (MG) has emerged as a life-threatening immune-related adverse event (irAE) in cancer patients treated with immune checkpoint inhibitors (ICIs). This syndrome closely resembles a rare form of idiopathic inflammatory myopathy (IIM) seen in a subset of MG patients. In this systematic review, we searched PubMed for reports of concurrent MG and IIM as well as ICI-related overlap syndromes. By integrating clinical, serological, and pathological observations, we delineated a previously unrecognized clinicopathological subtype of myositis that overlaps with MG. This entity is defined by a strong association with striational antibodies (StrAbs) and frequent co-occurrence with thymoma as a paraneoplastic process, and we classify it as StrAb-associated myositis. The idiopathic and ICI-induced forms share similar, though not identical, clinical, serological, and histopathological characteristics. We found that AChR antibody positivity, independent of established clinical risk factors such as respiratory or cardiac involvement, predicted more severe ICI-myotoxicity. Together with supporting evidence, our findings suggest a pathogenic model in which thymoma-driven cytotoxic T-cell responses trigger secondary AChR autoimmunity. These results highlight the potential utility of StrAbs and anti-AChR antibodies as practical biomarkers for diagnosis, risk stratification, and early intervention in patients at risk for severe neuromuscular irAEs.

## INTRODUCTION

Idiopathic inflammatory myopathies (IIMs) are a heterogeneous group of rare autoimmune disorders characterized by chronic muscle inflammation.^1^ Myositis-specific antibodies (MSAs), present in approximately 60 - 70% of patients, often correlate with unique clinical subgroups and seldom coexist in a single patient.^2^ Myasthenia gravis (MG) is another rare autoimmune disease, characterized by muscle weakness and abnormal fatigability. It results from autoantibody- mediated attack against components of the postsynaptic neuromuscular junction, most commonly the muscle nicotinic acetylcholine receptor (AChR), but also muscle-specific kinase (MuSK), low-density lipoprotein receptor-related protein 4 (LRP4), and agrin.^3^ Patients lacking detectable autoantibodies are referred to as seronegative MG.

The co-occurrence of MG and IIM is exceptionally uncommon. In a large single-center retrospective study, only 10 of 970 IIM patients (1%) developed MG.^4^ Similarly, two retrospective studies comprising 1,365 MG patients identified just 21 cases (1.5%) with concurrent IIM.^5,6^ Despite their rarity, these cases share striking clinical and serological features. Over 70% are associated with thymomas, contrasting sharply with the 15 - 20% thymoma prevalence in generalized MG.^7^ Systemic features characteristic of classic myositis are rarely observed. Serological profiles are characterized by dual positivity for anti-AChR antibodies and striational antibodies (StrAbs) targeting titin, ryanodine receptor (RyR), and Kv1.4 potassium channel. By contrast, conventional MSAs are rarely detected.

The overlap of MG and myositis has also emerged as a distinct immune-related adverse event (irAE) in patients treated with immune checkpoint inhibitors (ICIs).^8^ Clinical and serological features in these cases closely resemble the idiopathic cases, most notably the frequent presence of StrAbs and absence of conventional MSAs. Furthermore, patients with thymic epithelial tumors (TETs), particularly thymomas, appear to be at significantly higher risk of developing this syndrome during ICI therapy.^9^

In this study, we systematically reviewed the literature to identify both idiopathic and ICI-induced cases of concomitant MG and myositis. Our objectives were to delineate their clinical, serological, and histopathologic features, and to investigate the potential pathogenic mechanisms underlying their strong association with thymomas.

## Methods

### Search Strategy

We conducted comprehensive PubMed searches for eligible studies, without any language restrictions, from inception to 2024. Reference lists of identified publications and related articles were also reviewed to capture additional relevant studies. Although this review was not registered in PROSPERO, it adheres to the Preferred Reporting Items for Systematic Reviews and Meta-Analyses (PRISMA) guidelines.^10^ The detailed review protocol and complete search strategy are provided in Supplementary Appendix 1.

### Study selection

Published case reports and case series that reported original data (age, sex, primary symptoms, serological and neurophysiological findings, histological examinations, treatments, and clinical outcomes) were considered potentially eligible for inclusion. The database search was conducted by a single reviewer (J.S.). Articles identified through the search were imported into Covidence and independently screened by two reviewers (J.S. and J. Luo) based on titles and abstracts. Full- text articles were then reviewed for eligibility by the same two reviewers. Any discrepancies were resolved through discussion. Articles published in non-English languages were translated into English using ChatGPT (OpenAI, San Francisco, CA, USA), and the accuracy of critical information was verified using Google Translate.

### Inclusion and exclusion criteria

For the study on concomitant MG and IIM, we included patients with diagnosis of both conditions. Patients were excluded if they developed MG or inflammatory myopathies as an adverse effect of ICIs or were diagnosed with Lambert-Eaton myasthenic syndrome or congenital myasthenic syndrome. For the study on isolated IIM associated with TETs, we included patients diagnosed with myositis or myocarditis in the presence of TETs, but without associated MG. Patients with ICI-induced myositis/myocarditis were also excluded. For the study on ICI-induced inflammatory myopathies or MG, we included patients with diagnosis of either or both conditions as an adverse effect of ICI therapy. Cases presenting with clinical symptoms consistent with MG but not meeting full diagnostic criteria were classified as MG-like syndrome and were also included. Across these studies, individuals were eligible if they met criteria for definitive, probable, or possible myositis/myocarditis.

For the study on spontaneous regression of thymoma, we included patients with thymomas that regressed spontaneously without any relevant treatment, including tumor-directed therapy, corticosteroids, or other immunosuppressive agents. Given that many reports of spontaneous thymoma regression have been published in Japanese journals, we also performed a literature search using the J-STAGE and J-Global databases to identify additional eligible studies.

The diagnosis of MG was based on history and clinical signs of skeletal muscle weakness, along with one or more of the following four criteria: (1) presence of MG-specific serum antibodies (anti-AChR, anti-MuSK, or anti-LRP4); (2) clinical improvement following administration of acetylcholinesterase inhibitors; (3) a decremental response on RNS; or (4) increased jitter or blocking on SFEMG.^11^ Cases with detectable MG-specific serum antibodies in the absence of clinical manifestations of MG were classified as subclinical MG. Myositis was diagnosed according to the Bohan and Peter criteria^12,13^ with modifications, including clinical symptoms, elevated serum muscle enzyme levels, myogenic findings on EMG, inflammatory changes on muscle biopsy, edema-like signal abnormalities in affected muscles on MRI, and the presence of MSAs. Myocarditis was diagnosed based on the presence of cardiac symptoms without another identifiable cause, electrocardiographic abnormalities, new wall motion abnormalities on echocardiogram, cardiac MRI findings, elevated cardiac biomarkers, or histologic evidence of inflammation on endomyocardial biopsy.^14^ The diagnosis of TETs was based on imaging of an anterior mediastinal mass and confirmed by pathological examination, with classification according to the WHO histological classification system.^15^ For cases predating the WHO system, tumors were retrospectively classified into WHO categories by two reviewers (J.S. and J. Luo) when available histological descriptions (either textual or photographic) were deemed sufficient.

### Data extraction

Two reviewers (J.S. and J. Luo) independently extracted the following data into Microsoft Excel spreadsheets: study details (PubMed ID, publication year, country), patient characteristics (age, sex), diagnosis (disease subtype, diagnostic method, age at onset), pathological findings (immunological and histological features, muscular and cardiac enzyme levels), medical history, treatments, and clinical outcomes. Discrepancies in data collection between the two extractors were resolved through consensus discussions.

### Statistical analysis

All statistical analyses were performed using R version 4.5.1. The primary packages used included stats, quantreg, nnet, multcomp, and strucchange. A two-sided significance level of 5% was adopted.

Regression-based approaches were used to compare variables across groups while adjusting for potential confounders. For continuous outcomes, group means were compared using ordinary least squares (OLS) regression with t-tests for hypothesis testing, whereas group medians were compared using quantile regression, with inference based on the asymptotic normality of the estimator and kernel-based estimation of the conditional density. For non-continuous outcomes, we used generalized linear models (GLMs), selecting link functions appropriate to the outcome distribution: logistic regression for binary outcomes, multinomial regression for categorical outcomes with more than two levels, and Poisson regression for count data. Hypothesis testing in GLMs was performed using likelihood ratio tests. When the grouping variable had more than two levels, multiple comparison adjustments were applied. For OLS regression, Tukey’s Honest Significant Difference (HSD) procedure was used. For logistic regression, simultaneous inference procedures were implemented following the framework developed by Hothorn et al.^16^ In all other cases, Bonferroni correction was applied to control for multiple testing.

## Results

### Concomitant IIM and MG

We identified 797 records through a PubMed search and an additional 100 articles through reference screening, of which 155 studies involving 206 unique patients met the inclusion criteria (Fig. 1 and Supplementary Appendix 2A). Studies excluded after full-text review and corresponding reasons are provided in Supplementary Appendix 3A. To evaluate potential patient selection bias in case reports, we compared the demographic and clinical features of the 175 patients identified from case reports/series with those of 31 patients from three single-center retrospective studies (Supplementary Table 1). No significant differences were observed, supporting the validity of pooling data from all 206 patients.

**Fig. 1:**
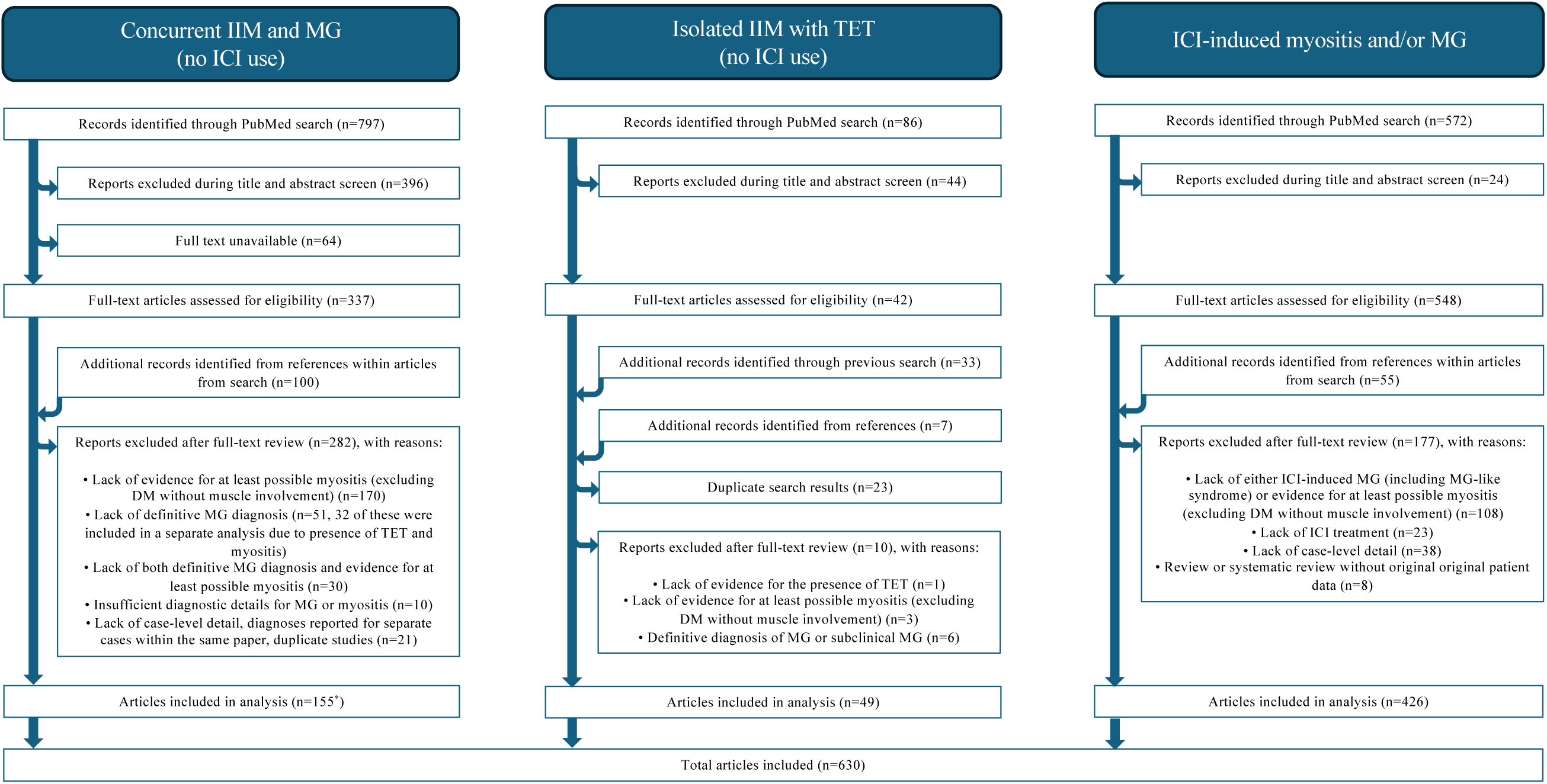
Flowchart illustrating the literature search and study selection process for reports of concomitant IIM and MG, TETs with isolated myositis, and ICI-induced myositis and/or MG. We conducted a comprehensive PubMed search from inception to September 30, 2024 for reports with concomitant IIM and MG; from inception to October 15, 2024 for reports with TETs with isolated myositis; and from January 1, 2010 to May 31, 2024 for reports with ICI-induced myositis and/or MG. Superscript symbol ‘*’ indicates that a case series included in the concomitant myositis and MG cohort also contained a case that contributed to the analysis of TET-associated isolated myositis.

Table 1 summarizes the key characteristics of the 206 cases, stratified by age at MG onset into early-onset (EOMG) and late-onset MG (LOMG). The demographic characteristics of this cohort closely resembled those of the general population with isolated MG or IIM (Fig. 2). Interestingly, patients with LOMG were more likely to develop MG and IIM in close temporal proximity, as indicated by shorter intervals between the two onsets. This overlap likely contributes to the reduced sensitivity of the acetylcholinesterase inhibitor test in LOMG, highlighting the diagnostic challenges arising from their intertwined clinical presentations.

**Fig. 2:**
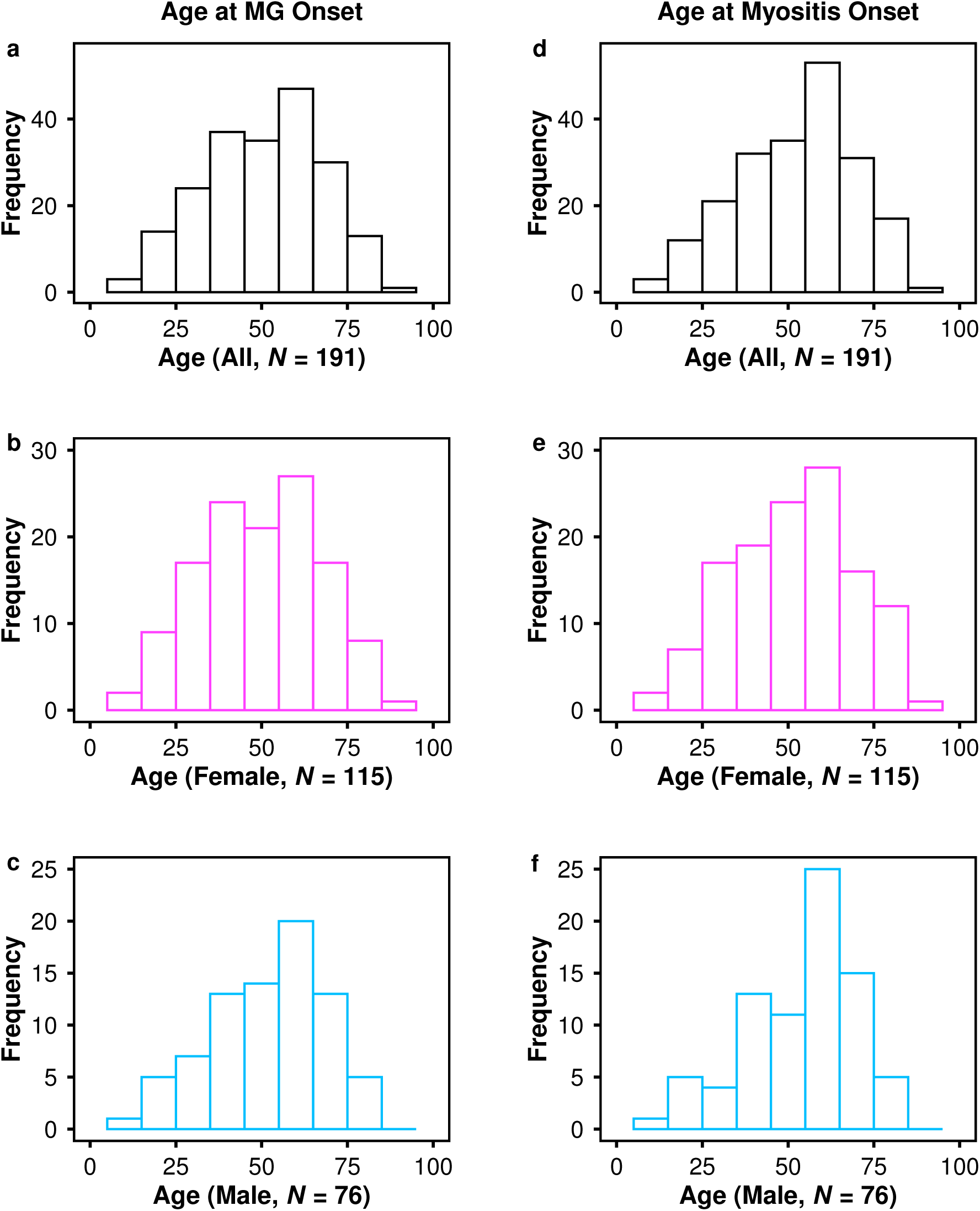
Histograms showing the distribution of age at symptom onset for MG (a-c) and IIM (d-f) in the cohort of 206 patients diagnosed with both conditions. In females, the age at onset of MG displays a bimodal distribution, with an early-onset peak in the third decade and a late-onset peak in the sixth decade (**b**), whereas males show a predominantly single late-onset peak (**c**). In contrast, the age at onset of IIM demonstrates a single late-onset peak in both sexes (**e** and **f**).

**Table 1.**
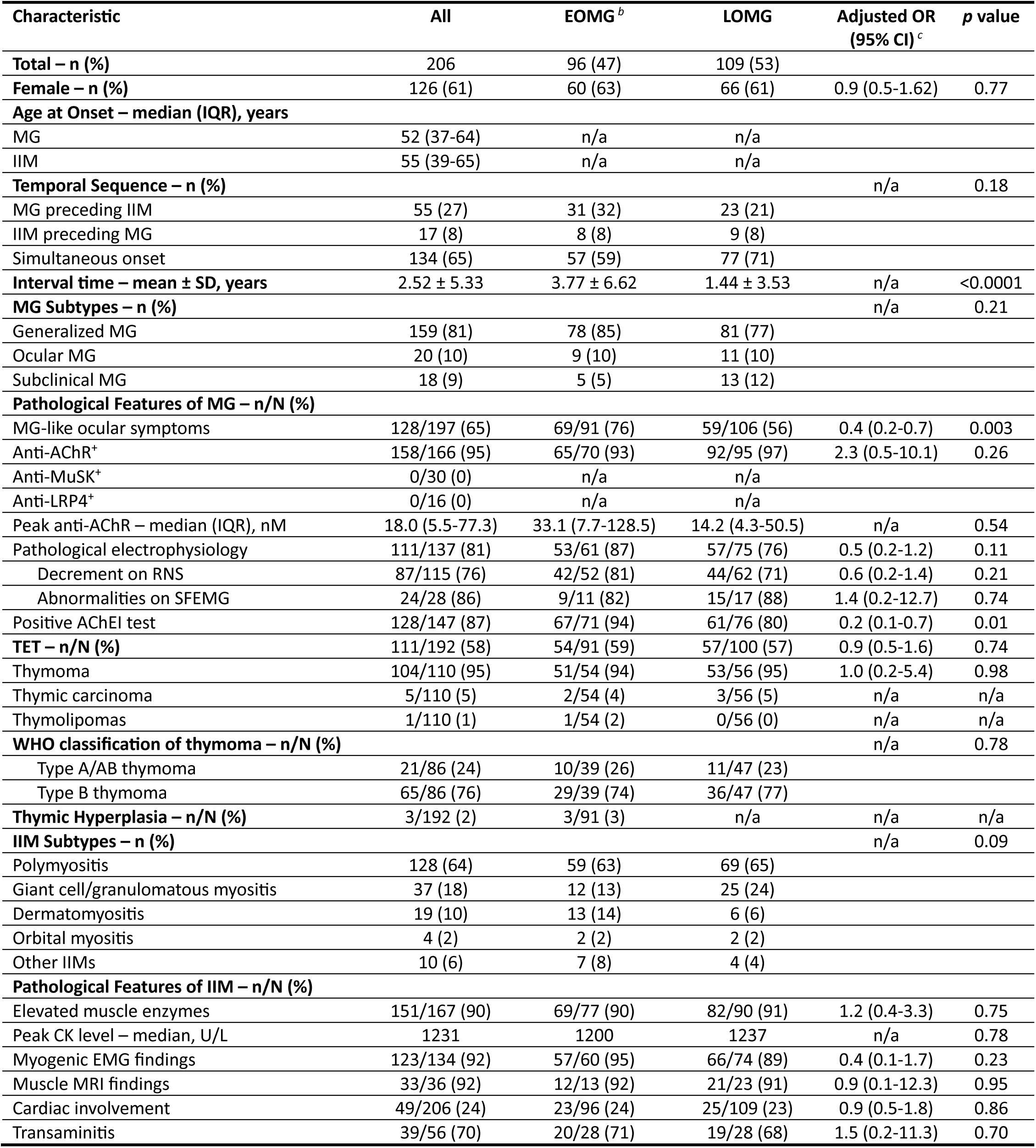
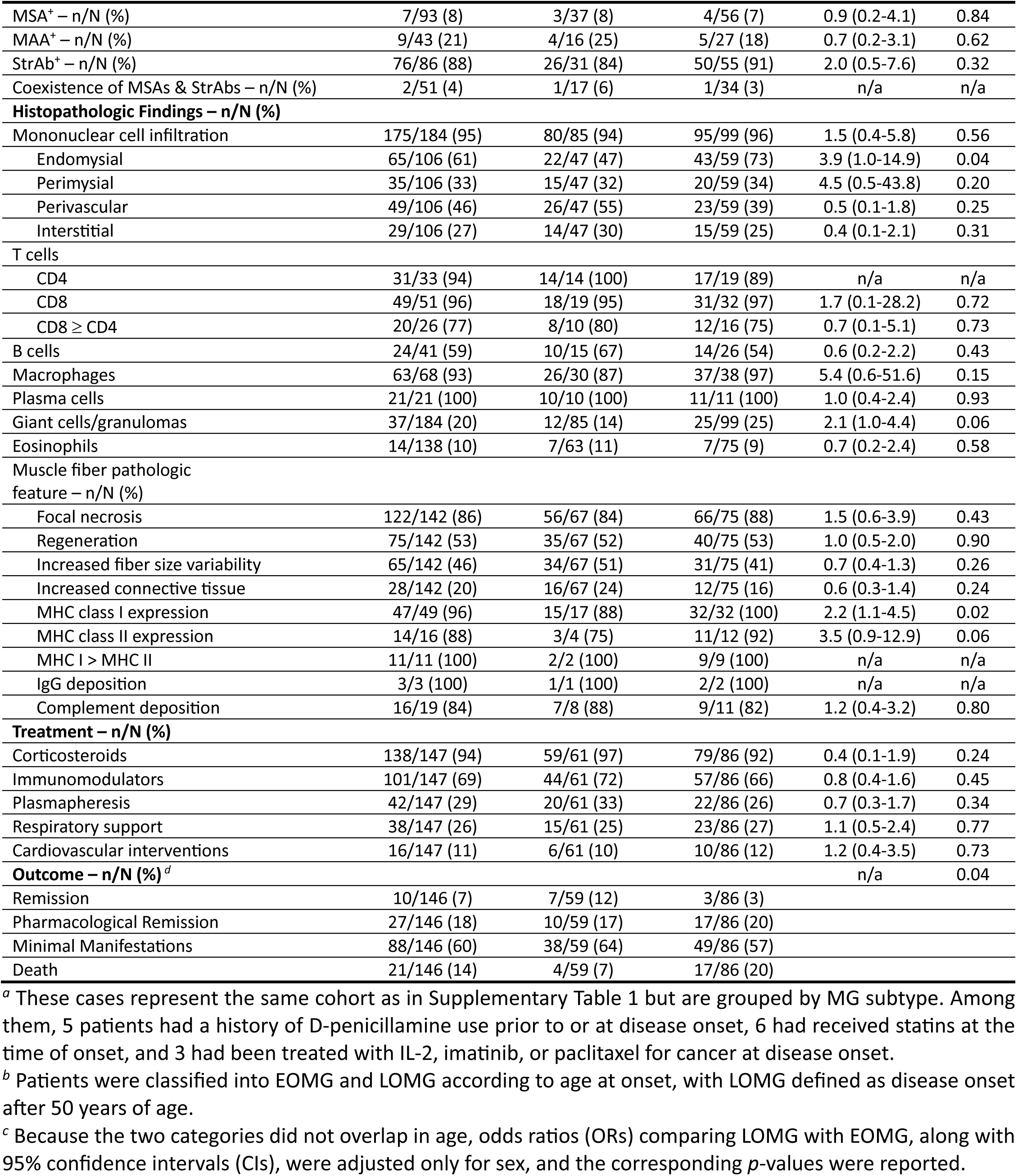
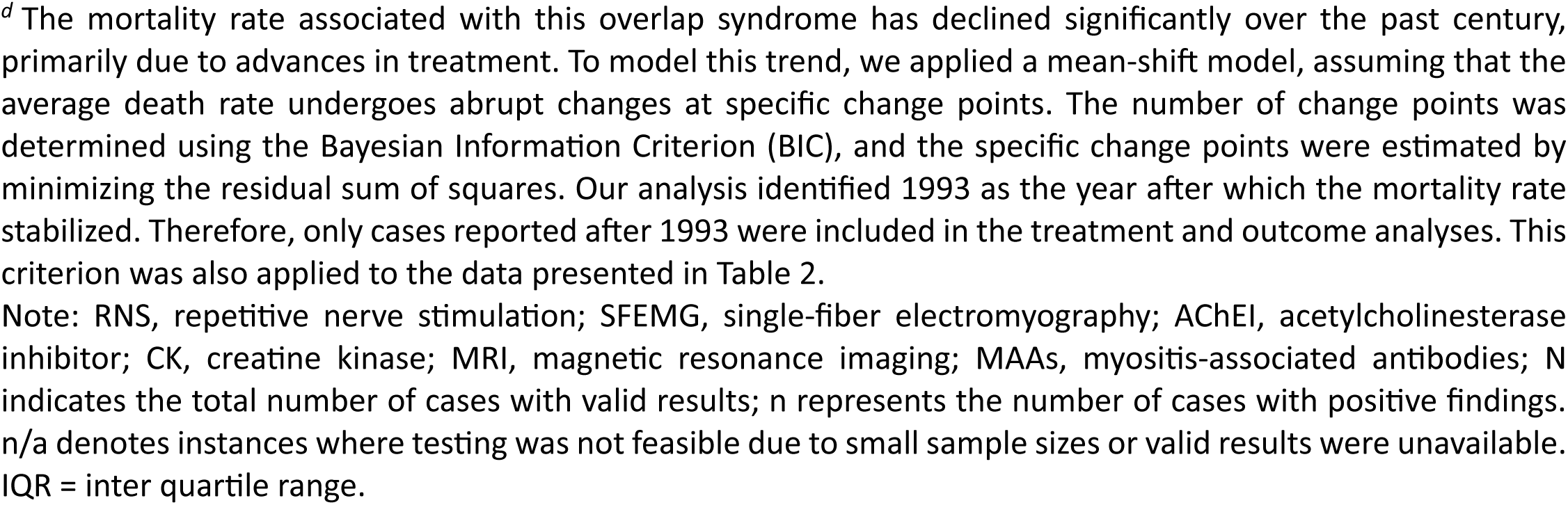
Demographic, pathological, treatment, and outcome characteristics of patients with concurrent MG and myositis, stratified by MG subtype (N = 206) *^a^*.

The vast majority of these cases had AChR-positive MG. None of the tested patients had anti- MuSK or anti-LRP4 antibodies. Nearly 90% examined patients tested positive for StrAbs while 8% were positive for MSAs, with only two individuals harboring both. Contrarily, StrAbs are seldom detected in classical myositis (Supplementary Table 2). This striking, nearly-exclusive presence of StrAbs strongly supports existence of a distinct IIM subtype, characterized by this novel group of MSAs, which we provisionally designate as StrAb-associated myositis.

LOMG cases showed worse outcomes than those with EOMG (mortality rate: 20% versus 7%, adjusted OR (adj-OR)=3.4, 95%CI:1.1-10.6, *p*=0.04), a difference that cannot be attributed solely to cardiac involvement or concomitant TETs (Table 1). Thymoma was also associated with worse outcomes (*p*<0.05). Nearly 60% of patients had an accompanying thymoma, over two-thirds of which were WHO type B, whereas concomitant thymic carcinoma and thymic hyperplasia were rare. The prevalence of thymoma did not differ by age or gender (Fig. 3a). Compared to patients without thymoma, those with thymoma exhibited a higher prevalence of StrAbs, granulomatous inflammation, and cardiac involvement, while none had detectable conventional MSAs.

**Fig. 3:**
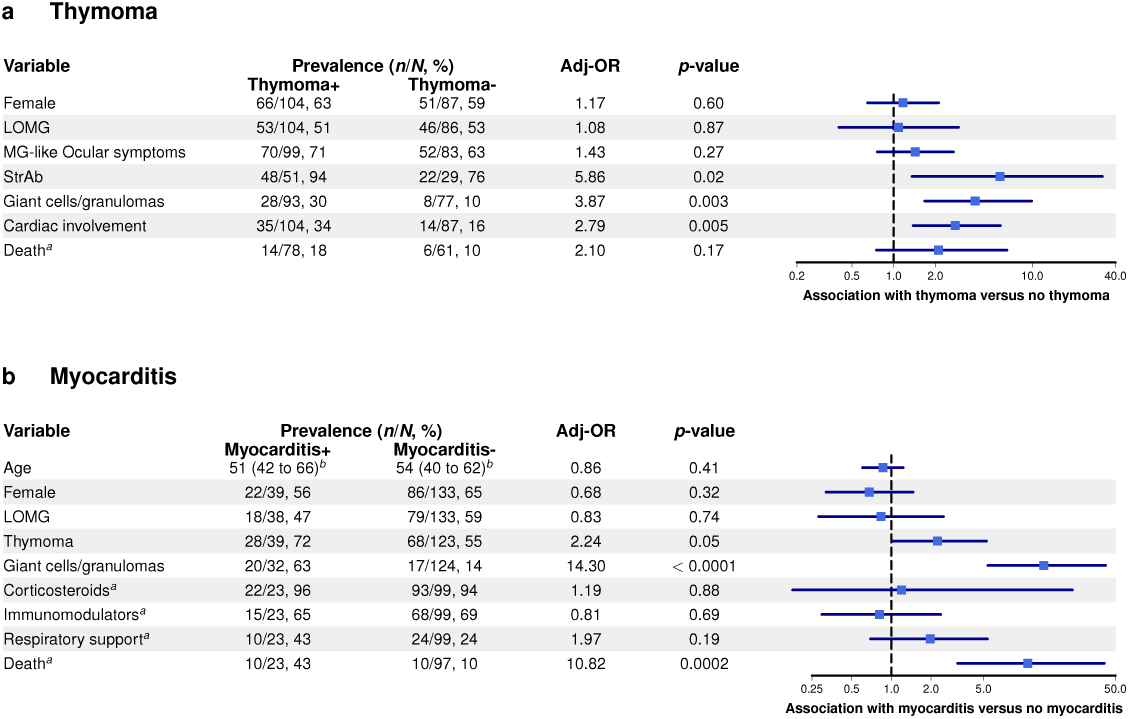
Forest plots of univariate analysis. **a**, Thymoma was significantly associated with StrAb positivity, giant cells and/or granulomas, and cardiac involvement (N = 192). Cases without available thymic pathology were excluded. **b**, Myocarditis was associated with thymoma and giant cells and/or granulomas and was linked to more severe outcomes (N = 172). Given the correlation between thymoma and granulomatous inflammation, interaction analyses were performed to examine their respective contributions to myocarditis risk. After adjusting for age, sex, and thymoma status, the presence of giant cells or granulomas remained significantly associated with an increased risk of myocarditis (*p*=0.03). In contrast, thymoma was not significantly associated with myocarditis risk after controlling for age, sex, and the presence of granulomatous inflammation (*p*=0.93). The interaction term between thymoma and granulomatous inflammation was not statistically significant (*p*=0.46), suggesting that the apparent contribution of thymoma to myocarditis risk may be secondary to its association with the development of granulomatous inflammation. Cases classified as “dermatomyositis,” “orbital myositis,” and “miscellaneous” in Table 2 were excluded from the primary analysis; sensitivity analyses that included these cases yielded similar results. Superscript letter ‘a’ indicates that only cases reported after 1993 were included in the treatment and outcome analyses. Superscript letter ‘b’ indicates that median ages with corresponding IQRs are shown. Adj-OR=odds ratio adjusted for age and sex.

**Table 2.**
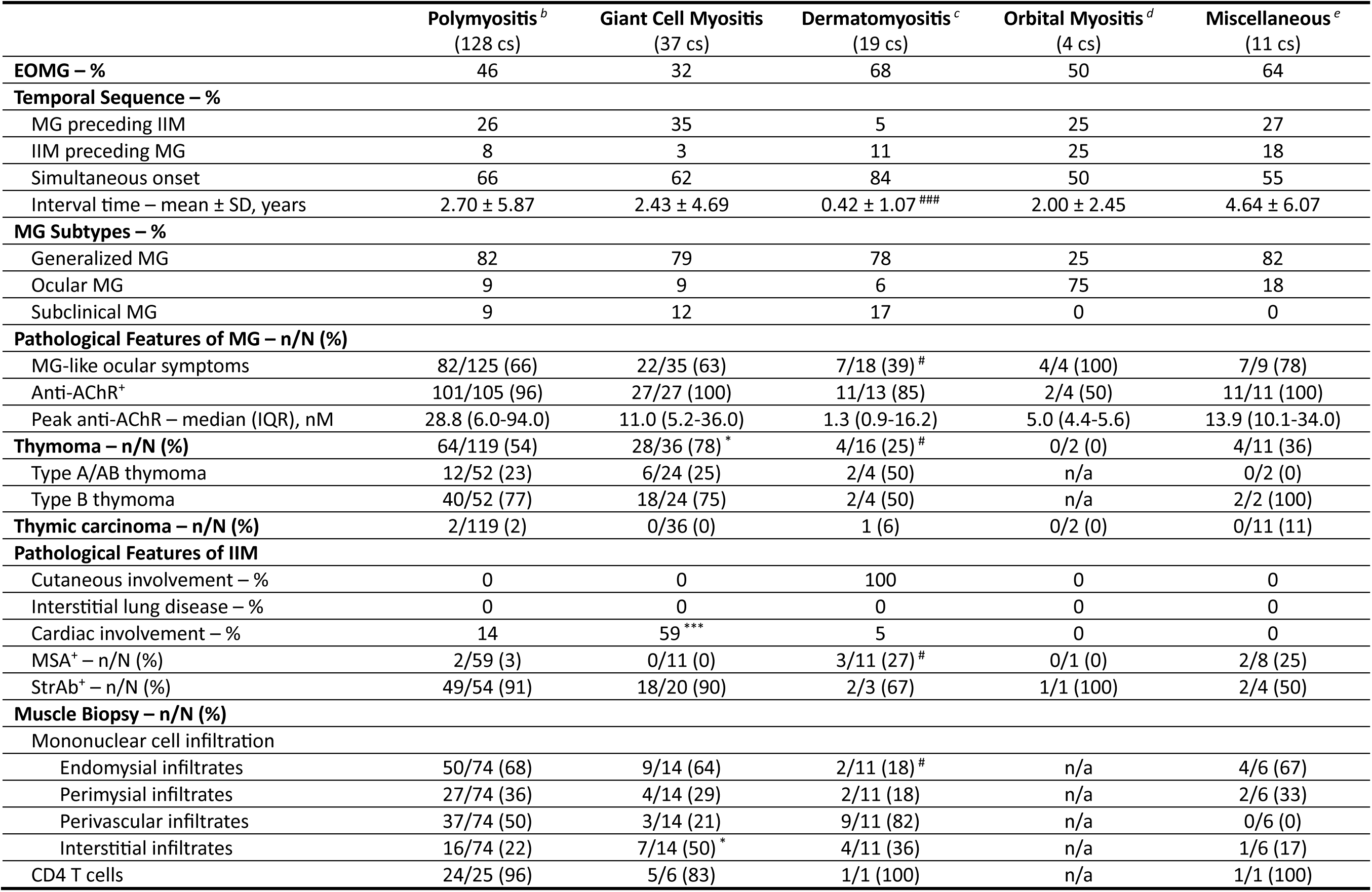
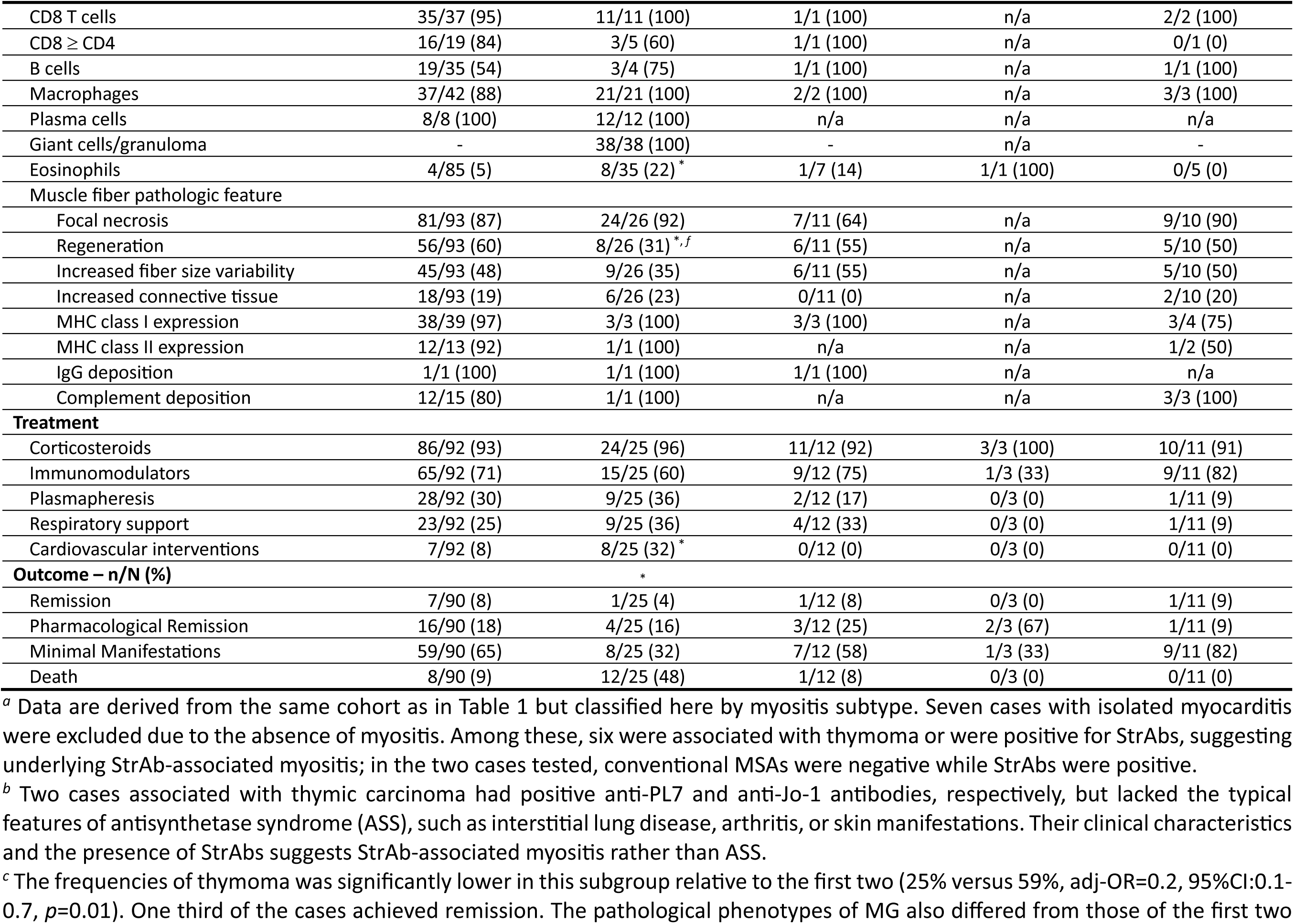
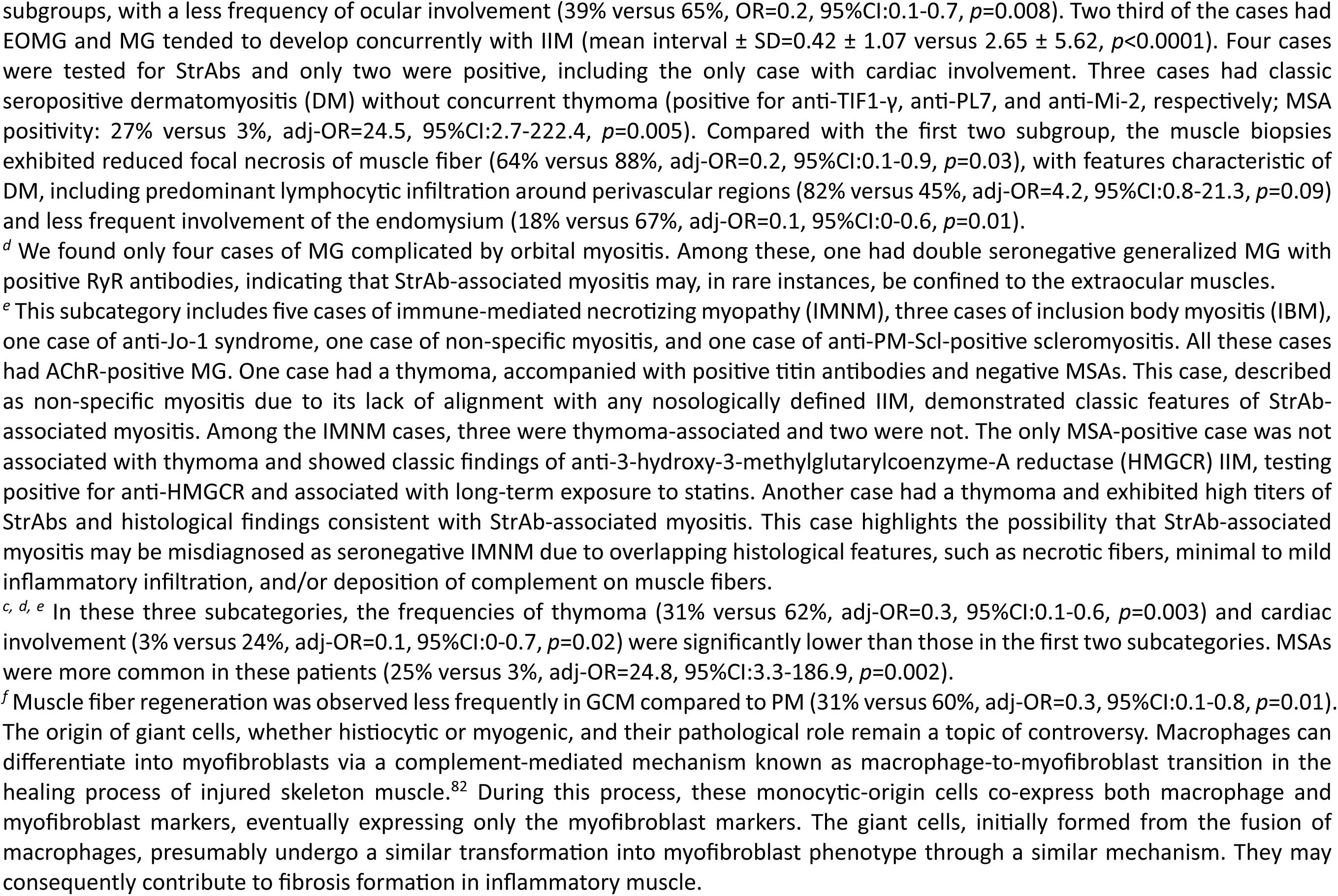
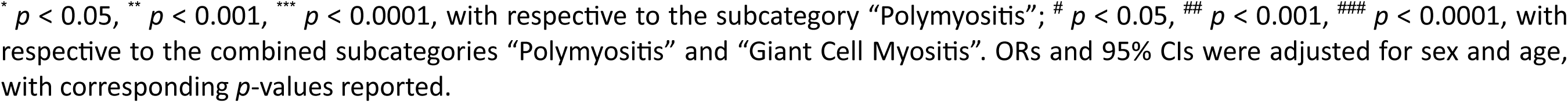
Clinicopathological features of the same cohort of patients with concurrent MG and myositis (N = 199), stratified by myositis subtype *^a^*.

### Classification Based on Myopathic Features

Based on the clinical and histopathological features, the same cohort presented in Table 1 was reclassified into five subgroups: polymyositis (PM), giant cell myositis (GCM), dermatomyositis (DM), orbital myositis, and a miscellaneous category (Table 2). Over 60% of cases exhibited a distinct phenotype characterized by chronic muscle weakness without prominent systemic manifestations such as skin lesions or interstitial lung disease. They rarely had MSAs, whereas StrAbs presented in 91% of tested patients, distinguishing them from any established myositis subtype. Accordingly, these cases were classified as PM.

Histopathological examination of this subgroup typically revealed mononuclear inflammatory cells surrounding and invading non-necrotic muscle fibers, accompanied by marked overexpression of major histocompatibility complex (MHC) class I molecules on the sarcolemma in almost all biopsies. MHC class II expression was also detected on muscle fibers in most specimens, albeit at lower levels. The inflammatory infiltrates, primarily located in the endomysium, were composed predominantly of CD8⁺ T cells and CD68⁺ macrophages, with fewer CD4⁺ T cells. B cells presented in > 50% of the examined cases, and plasma cells were consistently observed across all analyzed biopsies. IgG and complement deposits were frequently observed on muscle fiber membranes or within endomysial connective tissue adjacent to inflammatory foci, although these observations were based on a limited number of analyzed biopsies. Interestingly, lymphoid follicle-like structures resembling germinal centers were reported in muscle biopsies in two instances.^17,18^

Nonspecific histological changes included focal muscle fiber degeneration or necrosis, scattered muscle fiber regeneration, increased variability in fiber caliber, and enhanced perimysial and/or endomysial connective tissue. The lymphocytic infiltration was often extensive but varied from mild to severe. Occasionally, biopsies appeared normal or showed only non-specific alterations, likely reflecting sampling limitations due to the patchy distribution of necrosis and inflammation, or the confounding effects of prior steroid use.

The histological features of the second subcategory resembled those of the first, with the notable exception of multinucleated giant cells and/or granulomas within the lymphocytic infiltrates, an essential criterion for the diagnosis of GCM (Table 2). Nearly all examined patients were StrAb- positive, whereas none had MSAs. Another marked characteristic of this subgroup is its significantly higher risk of thymoma (78% versus 54% in PM, adj-OR=3.1, 95%CI:1.3-7.5, *p*=0.01) and cardiac involvement (59% versus 14% in PM, adj-OR=9.1, 95%CI:3.9-21.2, *p*<0.0001), contributing to substantially worse outcomes (mortality rate: 48% versus 9% in PM, adj-OR=6.3, 95%CI:2.8-14.2, *p*<0.0001). Collectively, the overlapping clinical, serological, and histological features support that both subgroups represent manifestations of the same disease entity, with GCM constituting a more severe form. Accordingly, the two subgroups were consolidated into a single category: StrAb-associated myositis.

The remaining three subcategories exhibited clinical characteristics that distinguish them from the first two. Although StrAb-associated myositis could not be definitively confirmed or excluded for most cases due to infrequent StrAb testing, the overall pattern suggests several possibilities: (1) these subcategories likely encompass more than one distinct subtype of myositis; (2) StrAb- associated myositis may occasionally exhibit atypical manifestations including orbital myositis and cutaneous involvement; and (3) such atypical symptoms are not characteristic of StrAb-associated myositis and appear to be rare in its isolated form.

Cardiac involvement is not uncommon in StrAb-associated myositis. Our analysis demonstrated that myocarditis risk is influenced by both thymoma status and the presence of granulomatous inflammation (Fig. 3b). Interaction analyses further suggest that the apparent effect of thymoma on myocarditis risk may be mediated through its association with granulomatous inflammation. The close association between myocarditis and myositis, along with the parallel histological features observed in myocardial and skeletal muscles, indicates that both tissues may harbor common autoimmune targets recognized by infiltrating inflammatory cells.

### StrAb-Associated Myositis without MG

To investigate whether StrAb-associated myositis can occur independently of MG, we conducted a PubMed search for reports of IIM in association with TETs, explicitly excluding cases with confirmed MG (Fig. 1). A total of 49 reports involving 49 unique patients met inclusion criteria (Supplementary Appendix 2C). We classified these cases into two subgroups based on TET type: thymic carcinoma and thymoma (Supplementary Table 3). Among patients with thymoma, 91% developed GCM or PM, of whom 71% tested positive for StrAbs and none for MSAs. In contrast, nearly 70% of patients with thymic carcinoma developed DM, with 63% of tested cases showing MSA positivity. These findings suggest that most thymoma-associated IIM cases likely represent StrAb-associated myositis, while thymic carcinoma is more commonly linked to classical myositis phenotypes, particularly DM.

Another markable pathological feature was that 62% patients with thymoma exhibited cardiac involvement, which was accompanied by markedly increased mortality (59% versus 7%, adj- OR=21.9, 95%CI:2.4-199.5, *p*=0.006). Because MG can develop several years after the onset of StrAb-associated myositis (Table 1), some patients who succumbed to severe disease may have eventually developed MG had they survived. Nonetheless, we cannot exclude the possibility that isolated StrAb-associated myositis may occur in rare instances.

### ICI-Induced Myositis and/or MG

To evaluate the presence of StrAb-associated myositis in ICI-treated cancer patients, we conducted a PubMed search for reports describing myositis, myocarditis, MG, or combinations of these conditions as adverse effects of ICI therapy (Fig. 1). Following full-text review, 426 studies involving 525 individual patients met inclusion criteria (Supplementary Appendix 2E). Studies excluded and corresponding reasons are detailed in Supplementary Appendix 3C. To assess potential patient selection bias inherent to case reports, we compared clinical characteristics of 300 patients with ICI-induced myocarditis from our dataset against 795 patients from an international ICI-Myocarditis Registry.^9^ Aside from the higher use of intravenous immunoglobulin in our cohort, no significant differences were observed (Supplementary Table 4), suggesting minimal selection bias in our assembled cohort.

Among 463 cases of ICI-induced myositis and/or myocarditis, 30 exhibited characteristic cutaneous manifestations consistent with DM (Supplementary Table 5). Nearly half of these patients had lung cancer, and approximately three-quarters were positive for MSAs, with anti- TIF1γ being the most prevalent (84%), supporting the likelihood of paraneoplastic DM. We identified only a single case of IBM following ICI therapy,^19^ and no ASS cases were found, suggesting that the PD1 and CTLA-4 pathways may play a limited role in the immunopathogenesis these conditions.

The remaining 432 cases did not align with any established myositis subtype and were classified as non-specific PM. Within this group, 45% of patients showed MG-like ocular symptoms, compared to 7% in the DM group. Additionally, 69% of patients had concurrent ICI-induced myocarditis, compared to 3% in the DM group. Patients in this category also exhibited poorer outcomes, with a trend toward higher rates of mortality or hospice care (31% versus 13%, adj- OR=2.7, 95%CI:0.9-8.1, *p*=0.07). Serologically, 41% and 72% of patients tested positive for anti- AChR and StrAbs, respectively, while only 6% were seropositive for MSAs. Together, these findings support the notion that this undefined myositis phenotype constitutes a distinct form of StrAb- associated myositis in the context of ICI therapy.

### Classification of ICI-Induced Myositis Based on the Predominant Muscles Affected

Compared to the idiopathic form, cases with ICI-induced StrAb-associated myositis exhibited similar but distinct clinical and serological features. While idiopathic StrAb-associated myositis is predominantly accompanied by MG, only 37% of ICI-related patients showed MG-like syndrome, and merely 26% had a confirmed diagnosis of MG. By contrast, cardiac involvement was significantly more frequent in the ICI-related group (71% versus 25%, adj-OR=8.1, 95%CI:5.1-12.8, *p*<0.0001). These patients also had a slightly lower prevalence of StrAbs (72% versus 91%, adj- OR=0.2, 95%CI:0.1-0.7, *p*=0.008). To further explore this distinct phenotype, we classified the 432 cases into three subgroups based on the primary muscles involved (Table 3).

**Table 3.**
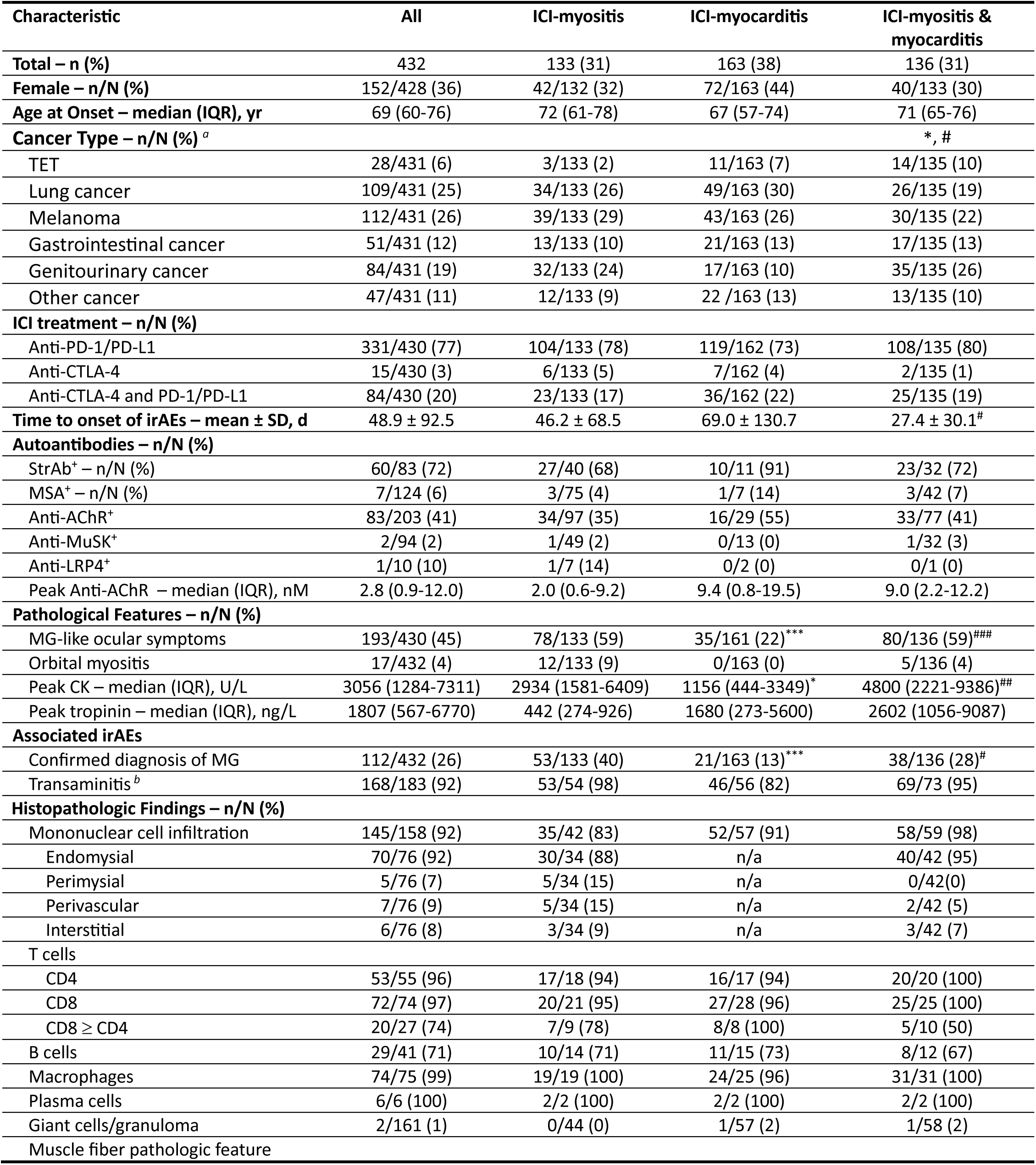
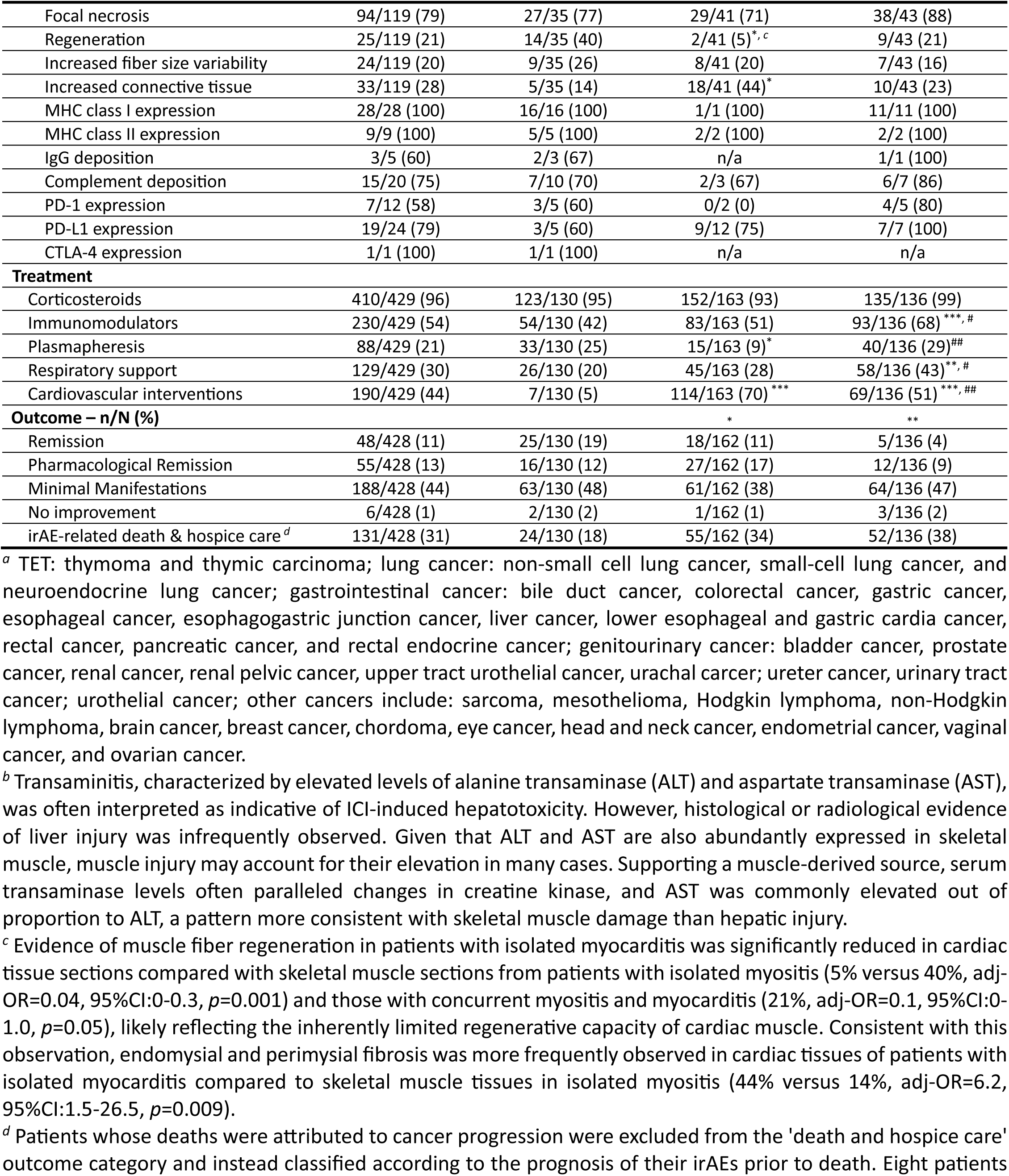
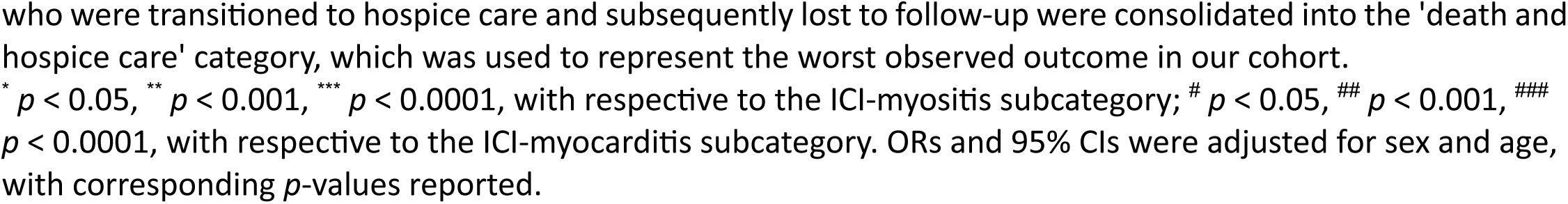
Subclassifications of ICI-PM cases from Supplementary Table 5 (N = 432), stratified into three phenotypic categories based on clinical manifestations.

Among these cases, 31% of patients developed isolated ICI-induced myositis, 38% developed isolated ICI-induced myocarditis, and the remaining 31% presented with concurrent ICI-induced myositis and myocarditis. Myocarditis, whether occurring alone or alongside myositis, was associated with significantly worse clinical outcomes, with higher rates of mortality or hospice care (34% in isolated myocarditis versus 18% in isolated myositis, adj-OR=2.7, 95%CI:1.4-5.5, *p*=0.002; 38% in concurrent myocarditis versus 18% in isolated myositis, adj-OR=2.8, 95%CI:1.4- 5.7, *p*=0.001).

MG-like ocular symptoms, including asymmetrical, painless ptosis and diplopia, are classic clinical signs of MG. Interestingly, such symptoms were less common in patients with isolated myocarditis (22% versus 59% in isolated myositis, adj-OR=0.2, 95%CI:0.1-0.4, *p*<0.0001; versus 59% in concurrent myositis and myocarditis, adj-OR=0.2, 95%CI:0.1-0.4, *p*<0.0001), who also demonstrated a significantly lower risk of developing concurrent MG (13% versus 40% in isolated myositis, adj-OR=0.2, 95%CI:0.1-0.5, *p*<0.0001; versus 28% in concurrent myositis and myocarditis, adj-OR=0.4, 95%CI:0.2-0.9, *p*=0.01). In cases of ICI-induced isolated myocarditis, histological and EMG analyses of skeletal muscle were infrequently performed due to the absence of clinical signs suggestive of myositis. However, among the cases where such assessments were conducted (n=5), ICI-induced myositis was ruled out in all instances, indicating that muscle inflammation was confined to cardiac tissue, with minimal or no skeletal muscle involvement.

This is further supported by the significantly lower peak creatine kinase (CK) levels observed in isolated myocarditis (median (IQR)=1156 (444-3349) U/L) compared to those in isolated myositis (2934 (1581-6409) U/L, *p*=0.02) and concurrent myositis and myocarditis (4800 (2221-9386) U/L, *p*=0.0005).

Histopathological analysis of skeletal muscle from patients with ICI-induced isolated myositis and those with concurrent myositis and myocarditis revealed mononuclear inflammatory infiltrates predominantly located in the endomysium, resembling the pattern of idiopathic StrAb-associated myositis. Infiltrates consisted primarily of CD8⁺ T cells and CD68⁺ macrophages, with fewer CD4⁺ T cells; B cells were present in 71% of cases, and plasma cells were found in all examined biopsies. Unlike idiopathic StrAb-associated myositis, multinucleated giant cells were extremely rare, presumably reflecting the typically acute or subacute onset of neuromuscular irAEs. MHC class I was consistently overexpressed on the sarcolemma, and MHC class II, though studied less frequently, was also upregulated in all tested samples. PD-1-positive inflammatory cells and sarcolemmal PD-L1 expression were common in inflammatory foci. Complement deposition on muscle fibers and/or within endomysial connective tissue was detected in most specimens, and germinal center-like structures were also identified in muscle biopsies.^20^ Parallel mononuclear cell infiltration and muscle fiber necrosis were also evident in endomyocardial biopsies from patients with isolated or concurrent myocarditis.

Across the 3 subgroups, 19% and 29% of all cases were assessed for StrAbs and conventional MSAs, respectively. Among those tested, 72% were positive for StrAbs, while only 6% tested positive for MSAs. Anti-AChR antibodies were assessed in 47% of cases, with a positivity rate of 41%, while anti-MuSK and anti-LRP4 antibodies were rarely detected. Although only one quarter of the cohort had a confirmed diagnosis of MG, the relatively high prevalence of anti-AChR antibodies raises the possibility of underdiagnosed MG.

### Impact of Testing Selection Bias on Autoantibody Seropositivity Rates

It is important to recognize that testing for anti-AChR antibodies and StrAbs (traditionally considered MG-related autoantibodies) is typically initiated by neurologists when characteristic MG symptoms are present. Therefore, in cohorts where only a subset of patients exhibits MG-like features, targeted testing may introduce selection bias that affects observed seropositivity rates.

To assess the extent of this bias, we evaluated the correlation between MG-like ocular symptoms and the likelihood of testing for anti-AChR and StrAbs. Testing for anti-AChR antibodies was strongly driven by the presence of MG-like ocular symptoms in both the isolated myositis and isolated myocarditis subgroups, whereas this effect was less pronounced in the concurrent myositis and myocarditis group (Fig. 4a). A similar trend was observed for StrAb testing, although the association did not reach statistical significance in the isolated myositis subgroup (Fig. 4b). Analysis of autoantibody positivity among patients with and without MG-like ocular symptoms showed no significant differences in either anti-AChR or StrAb positivity (Fig. 4c). Despite the presence of selection bias, these results indicate that its impact on overall seropositivity rates was likely limited.

**Fig. 4:**
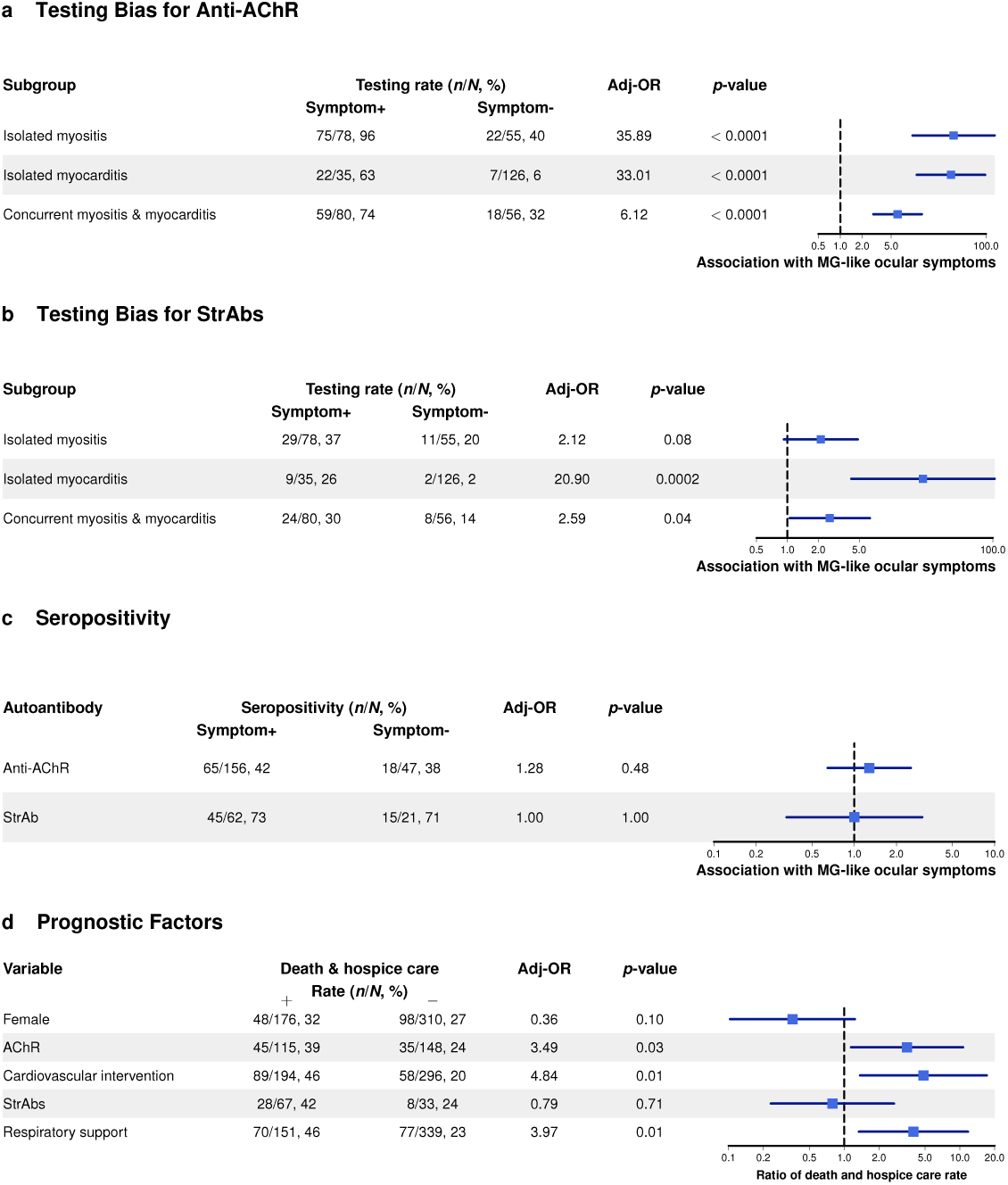
Selection bias from targeted testing based on MG-like ocular symptoms and prognostic biomarkers. **a**,**b**, Associations between MG-like ocular symptoms and the performance of serological testing for anti-AChR and StrAbs varied across clinical manifestations. **c**, Autoantibody positivity was not associated with the presence of MG-like ocular symptoms. **d**, Anti-AChR positivity, respiratory involvement (defined by the need for respiratory support), and cardiac involvement (defined by the need for cardiovascular interventions) each remained statistically significant after adjustment for confounders. By contrast, the presence of StrAbs was not significantly associated with increased risk of death or hospice care. ORs were adjusted for age and sex in **a**-**c**, and for age, sex, and additional covariates in **d**.

**Fig. 5:**
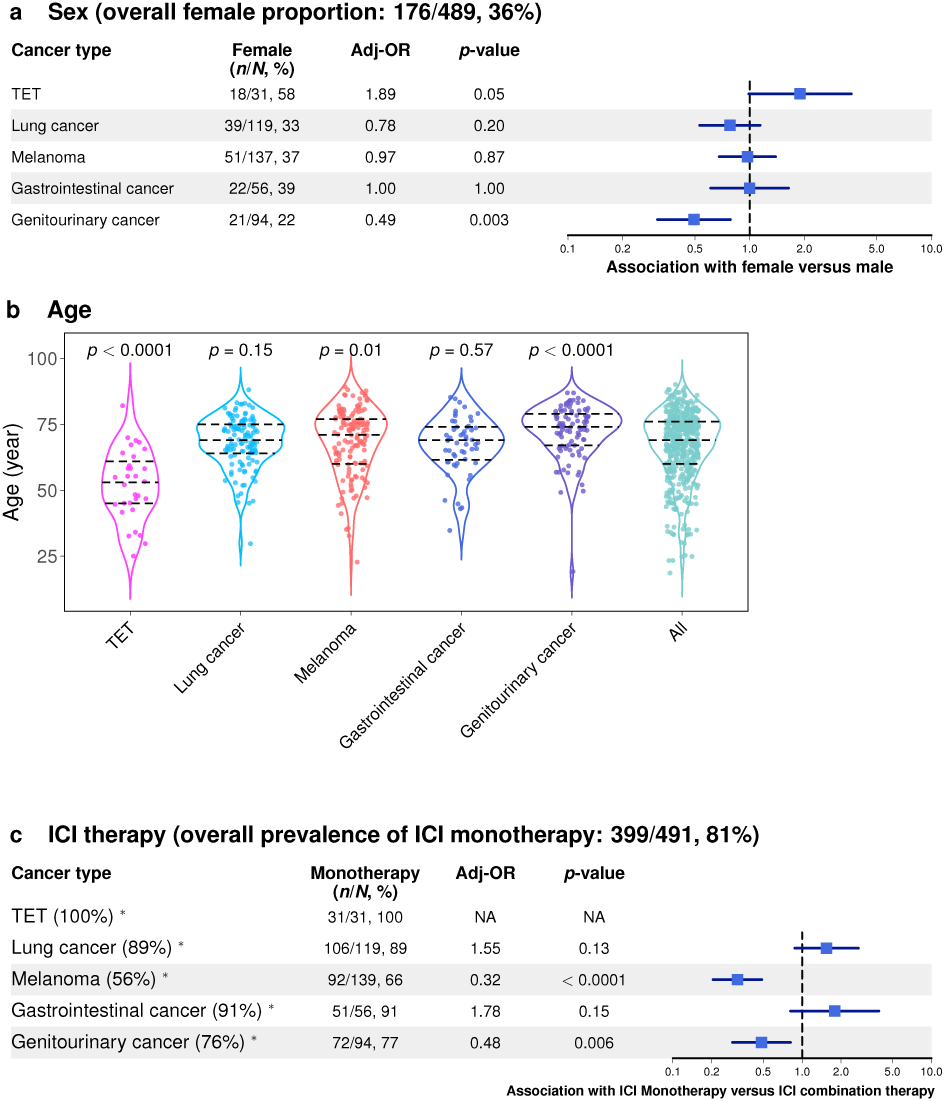
Variation in sex distribution, median age, and types of ICIs administered across different cancer types. Patients from Tables 3 and 4 were stratified into six groups according to their cancer type: TET (6%), lung cancer (24%), melanoma (29%), gastrointestinal cancer (11%), genitourinary cancer (19%), and an “other” category comprising all remaining cancer types (11%). The female proportion (**a**), median age (**b**), and prevalence of ICI monotherapy (**c**) within each cancer group were compared with their respective values in the overall cohort to assess cancer type-specific associations. Odds ratios were adjusted for age in analyses of sex distribution (**a**) and for age and sex in the analysis of ICI therapy (**c**). In the violin plot, dashed lines indicate median ages with corresponding IQRs (**b**). Valid OR and *p*-value for TET were not estimable because no patients in this group received ICI combination therapy (**c**). Percentages in parentheses (indicated by the superscript symbol ‘*’) represent the proportion of patients who received PD-1/PD-L1 monotherapy.

### ICI-induced MG

Beyond cases of ICI-induced MG occurring alongside StrAb-associated myositis, we identified an additional 62 instances of isolated ICI-induced MG, including 13 with clinical MG. Their demographic and clinical characteristics were compared with those of patients with concomitant MG shown in Table 3 (Table 4). In both groups, MG-like ocular symptoms were present in over 80% of cases. More than 70% of patients initially described as having MG-like syndromes ultimately received a confirmed MG diagnosis. Notably, 17% of patients with confirmed MG and coexisting myositis did not exhibit classical fluctuating muscle weakness, indicating that MG may manifest atypically in this overlap setting. Furthermore, all standard MG diagnostic tests, except SFEMG, exhibited reduced sensitivity in patients with overlapping myositis. In contrast, SFEMG demonstrated comparable sensitivity to that observed in isolated MG, underscoring its diagnostic value in this context.

**Table 4.**
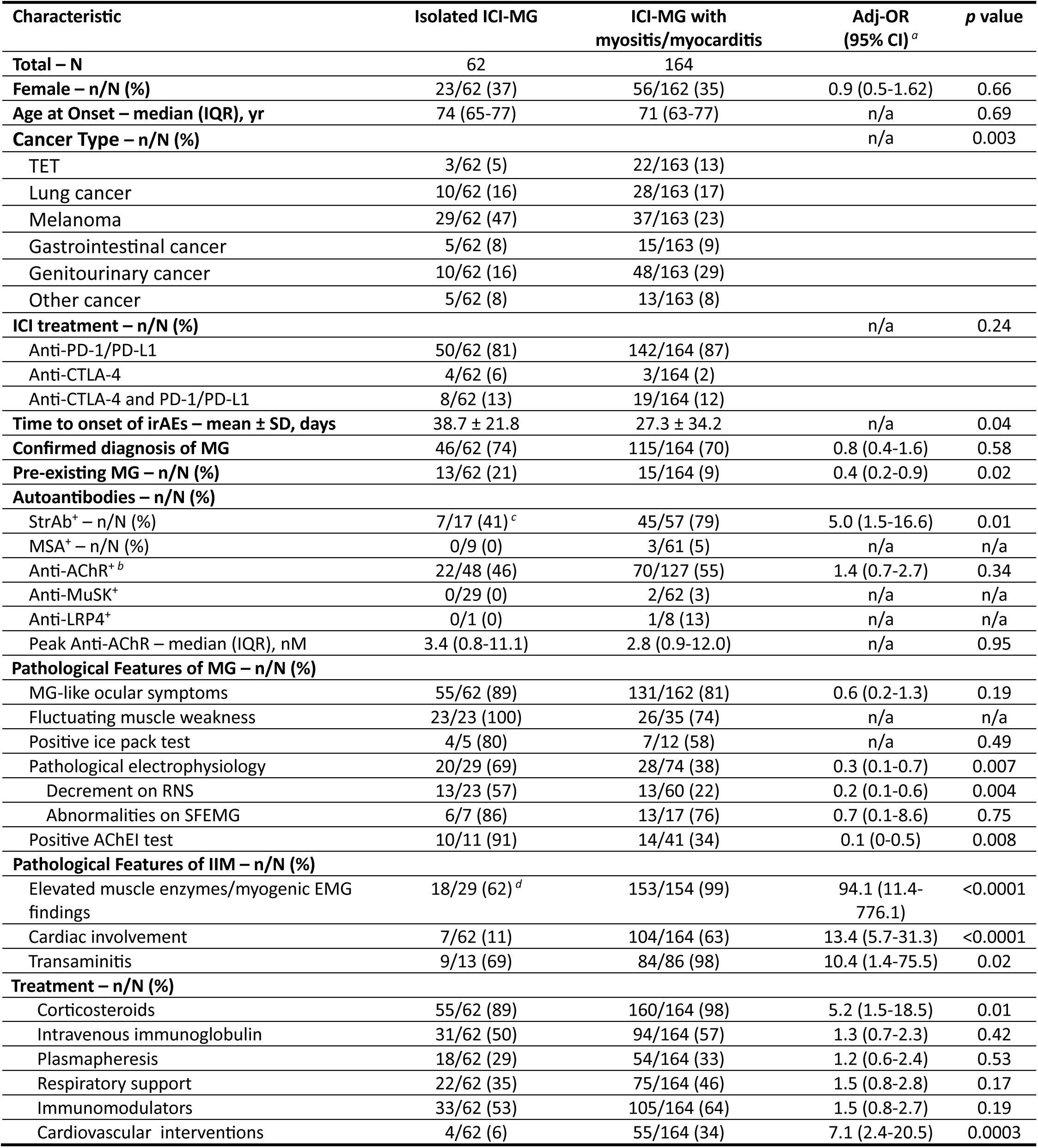
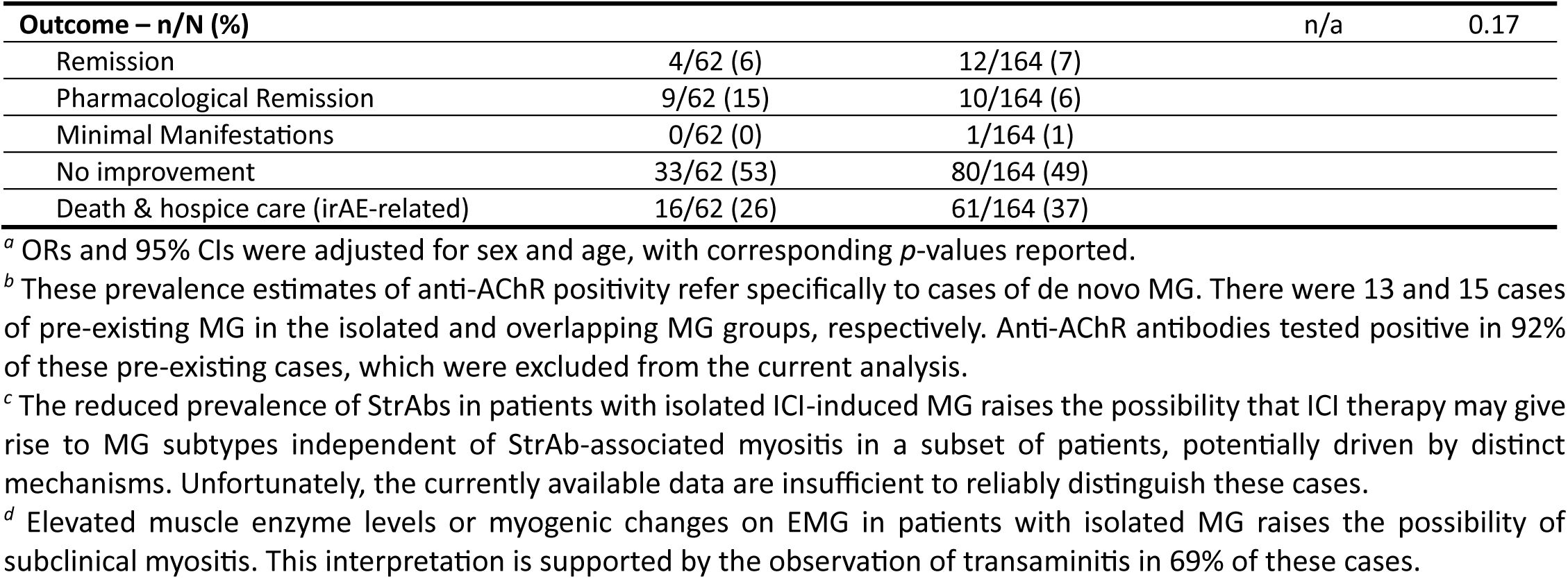
Comparison of ICI-induced MG-like syndrome cases without myositis or myocarditis (N = 62) and MG-like syndrome-associated cases (N = 164) drawn from the ICI-PM cohort in Supplementary Table 5. The latter group overlaps with those reported in Table 3.

It is well recognized that anti-AChR antibodies are more frequently negative in ICI-induced MG than in idiopathic MG.^21^ Even among seropositive cases, anti-AChR titers tended to be lower compared with idiopathic cases (median titer (IQR)=3.1 (0.8-11.8) versus 24.4 (5.9-78.0), *p*=0.06). A plausible explanation is that many patients with acute-onset ICI-induced MG were evaluated early in the disease course, before anti-AChR antibody levels had risen to detectable thresholds. Indeed, prior studies have shown that some idiopathic MG patients who initially test seronegative subsequently convert to seropositive status six months after onset.^22,23^ A similar mechanism may account for the slightly lower prevalence of StrAbs in ICI-induced StrAb-associated myositis compared with idiopathic cases. In our cohort, pre-treatment serum samples were available for six patients who underwent StrAb testing, five of whom exhibited elevated pre-existing StrAb levels. These findings suggest that ICI therapy may both unmask previously subclinical StrAb- associated myositis and, in some cases, trigger de novo disease.

### Prognostic Biomarker for Severe ICI-Induced Myotoxicity

We next investigated potential prognostic biomarkers. In the combined cohort from Tables 3 and 4, mortality or transition to hospice care occurred more frequently among patients with positive anti-AChR antibodies (39% versus 24% in seronegative patients, adj-OR=2.0, 95%CI:1.2-3.4, *p*=0.01), those with respiratory involvement (46% versus 23% without respiratory involvement, adj-OR=2.9, 95%CI:1.9-4.5, *p*<0.0001), and those with cardiac involvement (46% versus 20% without cardiac involvement, adj-OR=4.0, 95% CI: 2.6-6.1, *p*<0.0001). These associations remained significant when broader outcomes were considered (all *p*<0.05). In a multivariable model incorporating all candidate predictors, anti-AChR positivity, respiratory involvement, and cardiac involvement each retained statistical significance after mutual adjustment, underscoring their independent and robust contributions to poor prognosis (Fig. 4d).

Fenioux et al reported a higher prevalence of anti-AChR antibodies in patients with ICI-induced myocarditis.^9^ This association, however, was not replicated in our cohort (anti-AChR positivity: 46% versus 42%, adj-OR=1.2, 95%CI:0.7-1.9, *p*=0.55). Similarly, no association was observed between anti-AChR positivity and an increased risk of death or hospice care among patients with myocarditis (41% in AChR-positive versus 40% in AChR-negative patients, adj-OR=1.2, 95%CI:0.5- 2.6, *p*=0.72). By contrast, among patients with myositis, anti-AChR positivity was significantly associated with adverse outcomes (40% versus 23%, adj-OR=2.2, 95%CI:1.1-4.5, *p*=0.02). To further clarify these discrepancies, we performed interaction analyses between AChR positivity and myositis/myocarditis in relation to risk of death or hospice care. After adjustment for age, sex, and myocarditis, anti-AChR positivity remained significantly associated with worse outcomes (*p*=0.003). Myocarditis also remained an independent predictor of adverse outcomes after controlling for age, sex, and AChR positivity (*p*=0.0006). The interaction term between AChR positivity and myocarditis reached only marginal significance (*p*=0.08), supporting an additive rather than synergistic effect. In contrast, no significant interaction was observed between AChR positivity and myositis (*p*=0.60), and the association between AChR positivity and outcomes was attenuated after adjustment for myositis (*p*=0.30). Collectively, these findings suggest that the adverse prognostic impact of anti-AChR positivity is driven primarily by its association with myositis, rather than myocarditis.

Previous studies have identified thymic malignancies, especially thymoma, as major risk factors for ICI-induced neuromuscular irAEs.^9,24^ However, the contribution of other cancer types has remained less clear. In our analyses, patients with TET, of which 90% were thymoma, demonstrated higher frequencies of MG, myocarditis, and overlapping manifestations compared with their average prevalences across all cancer types, whereas the incidence of myositis in TETs was comparable to its overall average (Fig. 6). Myositis and concurrent MG and myositis/myocarditis were more common in patients with genitourinary cancer relative to their average incidences. In contrast, compared with overall averages, myocarditis was less common among patients with melanoma or genitourinary cancer, and MG were infrequent among those with lung or gastrointestinal cancers. Overlapping manifestations were also less frequent in patients with lung cancer.

**Fig. 6:**
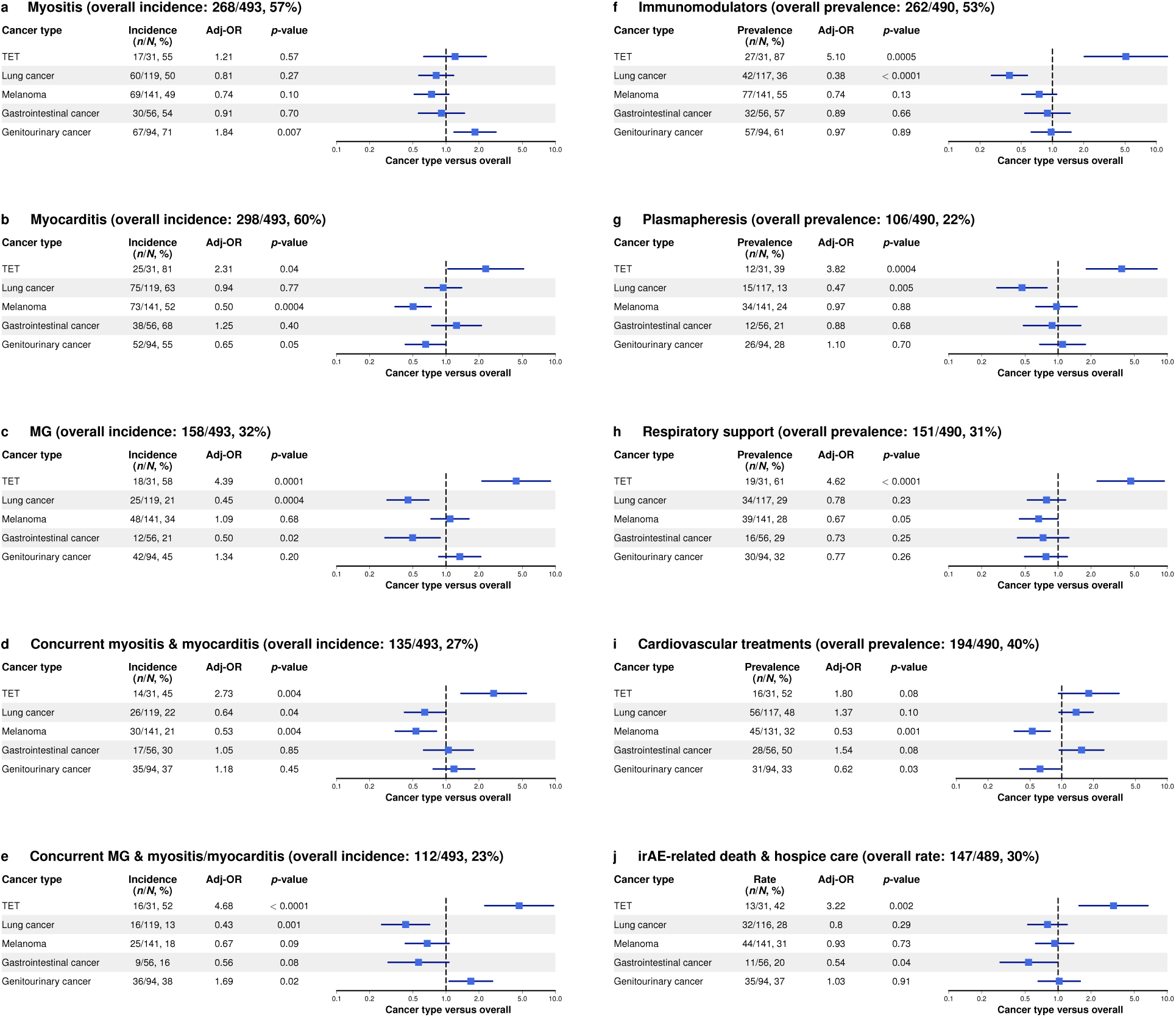
Associations between cancer type and neuromuscular irAE characteristics. Patients from Tables 3 and 4 were stratified into six groups according to their cancer type. The prevalences of MG, myositis, myocarditis, and their combinations within each cancer group were then compared with their respective prevalences in the overall cohort to assess cancer type-specific associations (**a**-**e**). Different cancer types showed distinct correlations with neuromuscular irAE manifestations. In accordance with these associations, patients with TET more often required escalation to second-line immunosuppressive therapy (**f**), plasmapheresis (**g**), and respiratory support (**h**). By contrast, the use of immunomodulators and plasmapheresis was less frequent among patients with lung cancer. Cardiovascular interventions were rarely employed in melanoma and genitourinary cancer (**i**). The rate of death and hospice care was significantly higher in patients with TET and significantly lower in those with gastrointestinal cancer compared with the overall cohort (**j**). Given that sex distribution, median age, and types of ICIs administered differed significantly across some groups (Fig. 5), all analyses were adjusted for sex, age, and ICI type.

## Discussion

In this systematic review, we characterized strikingly consistent clinical, serological, and histopathological features in patients with concomitant MG and IIM. These included a high prevalence of thymoma, the invariable presence of anti-AChR antibodies together with StrAbs, absence of conventional MSAs, and characteristic pathological findings showing both cellular infiltrates and humoral component deposition in striated muscle. Collectively, these observations delineate a distinct form of myositis defined by the presence of StrAbs, a newly recognized class of MSAs. Our analysis suggests that thymoma may increase the risk of myocarditis by promoting granulomatous inflammation. Contrarily, thymic carcinoma was more commonly associated with DM, DM-specific autoantibodies, and seldom with myocarditis.

By reviewing 525 cancer patients who developed neuromuscular toxicity following ICI therapy, we differentiated cases of StrAb-associated MG-myositis overlapping syndrome from those of DM and identified the former as the predominant form of myotoxic irAEs. Unlike ICI-induced DM, which showed a strong correlation with lung cancer, the StrAb-associated syndrome was strongly associated with TET, particularly thymoma, and to a less extent with genitourinary malignancies. Compared with idiopathic cases, ICI-induced cases showed less frequent MG but more frequent cardiac involvement. We also identified anti-AChR antibodies as a potential prognostic biomarker in ICI-induce myositis, contributing to severe outcomes through a mechanism that appears to be independent of established clinical risk factors such as respiratory or cardiac compromise.

Aside from rare cases involving concomitant MG and IIM, StrAbs are also found in many anti- AChR-positive MG patients without overt myositis, especially those with thymoma.^25^ These individuals frequently exhibit myopathic abnormalities on EMG and muscle inflammation with histological features resembling PM (so-called lymphorrhages).^26,27^ These parallel serological, electrophysiological, and histological findings support co-occurrence of a subclinical form of StrAb-associated myositis in this subgroup, as detailed in Supplementary Appendix 4. Building on these observations and existing evidence, we propose a novel hypothesis regarding the pathogenesis of this MG-myositis syndrome in the context of thymoma.

Previous studies have documented histopathological and functional abnormalities in thymomas. Neoplastic thymic epithelial cells (TECs) exhibit variable mixtures of cortical TEC-like (cTEC-like) and medullary TEC-like (mTEC-like) phenotypes, intermingled at the single-cell level or forming small clusters.^28^ Most neoplastic mTECs lack expression of the autoimmune regulator (AIRE),^29^ reshaping the thymic microenvironment by impairing AIRE-dependent tissue-specific antigen (TSA) expression, inducing abnormal CTLA-4 overexpression, and reducing chemokines required for dendritic-cell recruitment and indirect antigen presentation.^30–33^ Thymoma TECs express little MHC class II molecules,^34^ leading to inefficient positive selection of CD4^+^ T cells and skewed intratumoral maturation toward a CD8^+^ phenotype.^35–37^ RNA-seq studies demonstrate increased intratumoral expression of the AChR α1 subunit gene (CHRNA1) and several neuron-specific genes (e.g., NEFM, RYR3, KCNC2, GABRA5) in thymoma patients with MG compared to those without MG.^29,38^ The coded proteins often harbor cross-reactive linear T cell epitopes shared with striational antigens, arising either from sequence homology or from broader immunogenic similarities with these targets.^39–42^ Overexpression of this relatively limited antigen set in neoplastic mTECs may produce disproportionately high densities of the corresponding self- peptide-MHC complexes, thereby amplifying the overall strength of T cell receptor (TCR)-self- peptide-MHC interactions.

We hypothesize that neoplastic mTECs act as atypical antigen-presenting cells (APCs) in the autoimmunogenic thymoma microenvironment, directly presenting neuromuscular self-antigens to striated-muscle-specific CD8⁺ T cells (Fig. 7). This self-autonomous presentation drives intratumoral CD8⁺ T-cell activation. Several lines of evidence support this model: (1) thymoma cells from MG patients spontaneously produce autoantibodies against type I interferons (IFN-Is) and IL-12, with serum levels rising further at recurrence or metastasis, indicating ongoing autoimmunization within the tumor milieu;^43,44^ (2) neoplastic mTECs express MHC class I and the required costimulatory molecules for CD8⁺ T-cell engagement;^34,45^ (3) MG-associated thymoma TECs can present small peptides to specific T cells *in vitro* with efficiency comparable to professional APCs;^46^ (4) thymocytes from patients with thymoma-associated MG (TAMG) display TCR-independent cytotoxicity in the presence of low concentrations of IL-2;^47^ and (5) CD8⁺ T lymphocytes infiltrating skeletal muscle in TAMG patients without clinically IIM exhibit a naïve-like CD45RA^+^ phenotype closely resembling that of thymoma-exported T cells.^48^ Once activated, neuromuscular-specific CD8⁺ T cells emigrating from thymomas can traffic to peripheral tissues, where they differentiate into tissue-resident CD8⁺ memory T cells and persist for years, even after thymectomy.^35^ These CD8⁺CD45RA⁺ T cells have been detected adjacent to muscle fibers overexpressing MHC-I at sites of lymphocytic infiltration in patients lacking clinical myositis, whereas the emergence of overt myositis correlates with a shift toward CD45RA⁻ infiltrating lymphocytes.^4,48^ Therefore, thymoma-exported CD8⁺CD45RA⁺ cells likely represent antigen- experienced T cells with a naïve-like phenotype rather than bona fide naïve cells.

**Fig. 7:**
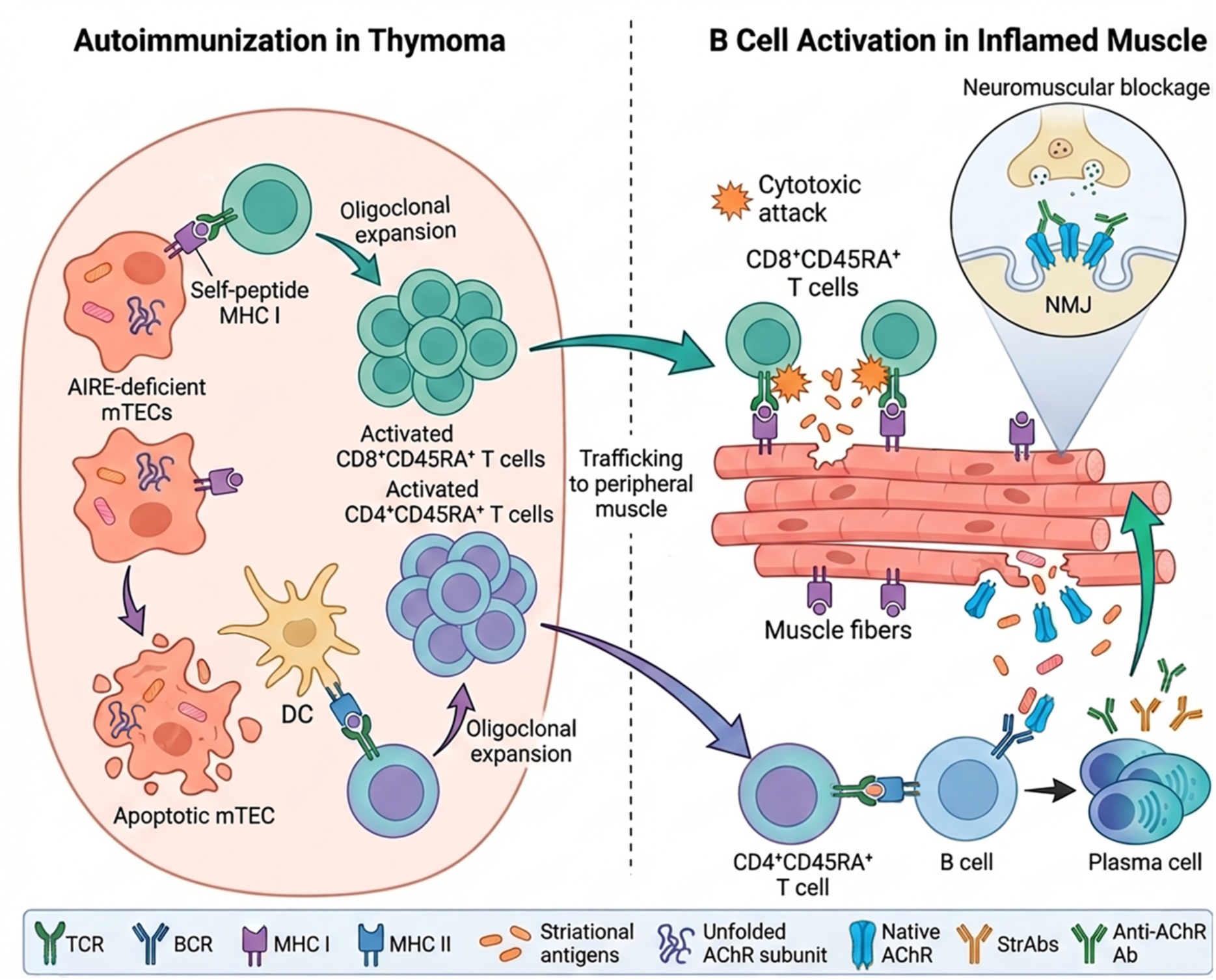
A proposed model created using FigureLabs.ai. Thymomas provide an autoimmunogenic niche that promotes the expansion of striated muscle-specific cytotoxic T cells. These T cells initiate myositis and, through local intramuscular autoimmunization, propagate a secondary humoral response against AChRs.

Interestingly, the humoral response in TAMG patients is narrowly focused on a limited set of muscle antigens that share linear epitopes with proteins expressed by neoplastic TECs, rather than broadly targeting the muscle proteome. This pattern suggests that humoral specificity is constrained by the availability of specific CD4⁺ T-cell clones primed and expanded within the thymoma microenvironment. Autoantibodies against AChR, cytokines, and striational antigens are largely restricted to Th1-biased IgG subclasses.^49–51^ This polarization likely reflects the cytokine milieu of thymomas, which display high levels of IFN-I and IL-12, canonical drivers of proinflammatory Th1 responses.^52^ Given the absence of MHC II-positive TECs, extrathymic plasmacytoid and conventional dendritic cells are likely recruited into the distorted thymic environment to cross-present self-antigens derived from apoptotic neoplastic TECs.^53^

Neuromuscular-specific T cells exported by the thymoma undergo further oligoclonal expansion driven by chronic inflammation in muscle and neuronal tissues.^54^ Over time, this progressive skewing of the T-cell repertoire reduces overall TCR diversity, as these expanded clones increasingly occupy peripheral immunological space. Together with the imbalanced reconstitution of CD4⁺ and CD8⁺ T-cell thymopoiesis, the resulting contracted repertoire may ultimately impair effective immunosurveillance of malignant cells. Clinically, this mechanistic framework offers a plausible explanation for the high prevalence of extrathymic cancers in patients with thymoma.^55^

The majority of anti-AChR autoantibodies in MG patients are highly conformation-dependent and show minimal cross-reactivity with linear AChR peptides.^56,57^ Because native AChR and AChR-expressing myoid cells are absent in thymomas, these tumors are unlikely to serve as the site of AChR-specific B-cell priming.^58,59^ Consistently, thymoma-derived cells seldom produce anti-AChR antibodies or StrAbs spontaneously,^60^ and antibody titers against these antigens do not consistently rise following thymoma recurrence or metastasis.^43,44^ Collectively, the activation of B cells specific for AChR or striational antigens likely occurs outside the thymoma microenvironment.

In MG, anti-AChR antibodies selectively target the skeletal muscle nicotinic AChRs and do not cross-react with heart muscle, owing to distinct AChR subtypes expressed in these tissues.^61^ Skeletal fibers predominantly express skeletal muscle nicotinic AChRs, whereas cardiac muscle expresses muscarinic and neuronal nicotinic AChRs.^62^ If the autoimmune response targeting skeletal muscle AChRs originates primarily from peripheral autoimmunization within inflamed skeletal muscle, patients whose inflammation is confined to the myocardium would be expected to have a lower risk of developing MG. This aligns with our finding that MG occurs far less frequently in isolated ICI-induced myocarditis than in cases with concomitant myositis.

The parallel histopathological features observed in both idiopathic and ICI-related cases, including muscle fiber degeneration, sarcolemmal MHC II expression, CD4⁺ T-cell-rich inflammatory foci, the presence of B cells and plasma cells, IgG and complement deposition on muscle fibers, and lymphoid follicle-like architectures, collectively indicate ongoing intramuscular autoimmune activation. Cytotoxic injury mediated by striated-muscle-specific CD8⁺ T cells drives muscle fiber degeneration and necrosis, which, if inadequately cleared, may create a reservoir of native AChRs and striational antigens. Macrophages and B cells located near endplates may capture, process, and present these antigens, thereby supporting the initiation and perpetuation of the autoreactivity.^63^ Demonstrating muscle-resident B cells capable of spontaneously producing AChR-specific antibodies or StrAbs would provide direct evidence of ongoing autoimmunization within skeletal muscle.

Despite the strong association between StrAb-associated MG-myositis overlap syndrome and thymoma, 40% of idiopathic cases and most ICI-related cases lacked detectable thymomas. This discrepancy raises a critical question: how do autoreactive T cells escape central tolerance and become activated in nonthymomatous individuals? One possibility is underdiagnosis of thymoma, as some patients may harbor microscopic lesions undetectable by computed tomography; others may have already rejected an occult thymoma prior to clinical evaluation. Supporting evidence is summarized in Supplementary Appendix 5.

Non-thymic factors may also contribute. StrAbs have been reported in extrathymic malignancies,^64–67^ suggesting that autoimmunity to striated muscle may arise as a paraneoplastic phenomenon driven by tumor-derived onconeural antigens expressing cryptic epitopes. Additional support comes from the development of StrAb-associated myositis in ICI-treated patients with extrathymic malignancies. In our analysis, in addition to TETs, genitourinary cancers showed a strong association with StrAb-associated myositis. Genetic predisposition may further facilitate the emergence of striated-muscle-specific autoreactive T cells. A loss-of-function variant in the gene PTPN22, which enhances TCR signaling in autoreactive T cells bearing low-avidity TCRs and promotes their activation and oligoclonal expansion,^68^ has been frequently found in a subgroup of nonthymomatous MG patients with anti-titin autoantibodies.^69^ Broader GWAS and integrative genomic studies will be instrumental in identifying additional risk alleles and elucidating the molecular basis of muscle autoimmunity in these patients.

The presence of StrAbs serves as the hallmark of StrAb-associated myositis, and their detection is central to establishing the diagnosis. Although pre-treatment StrAb testing has rarely been performed in ICI-treated patients, limited evidence suggests that subclinical StrAb-associated myositis may frequently pre-exist in this population. This raises the possibility that pre-treatment StrAb screening could help stratify the risk of myotoxic irAEs, though systematic prospective studies are needed to rigorously evaluate its predictive utility.

The co-concurrence of MG and StrAb-associated myositis pose significant diagnostic challenges due to overlapping muscular symptoms. Characteristic myalgia and proximal muscle weakness are not always present, and isolated bulbar and axial weakness is not uncommon. Severe complications such as respiratory failure from diaphragmatic myositis or cardiac insufficiency may mimic myasthenic crisis. When both conditions affect the same muscle groups, symptom fluctuation may be less pronounced, and acetylcholinesterase inhibitors may provide limited or no benefit. Our review identified cases in which MG was confirmed electrophysiologically but accompanied by fixed muscle weakness.^70–72^

Anti-AChR antibody positivity plays a pivotal role in diagnosing ICI-induced MG, and in many cases MG is confirmed solely on the basis of seropositivity. However, serology alone is insufficient: negative results do not exclude MG because of the substantial seronegative rate, and elevated titers do not reliably differentiate symptomatic from subclinical disease. A comprehensive diagnostic evaluation remains essential regardless of serostatus. In our analyses, all conventional MG diagnostic tests, except SFEMG, showed reduced sensitivity in patients with overlapping myositis, underscoring the unique diagnostic value of SFEMG in this setting.

Asymmetrical, painless ptosis and diplopia are classic features of MG and are virtually absent in classic IIM. Their occurrence in a patient with myositis strongly supports the diagnosis of StrAb- associated myositis. Autopsy studies in MG have documented lymphorrhages in extraocular muscles.^27^ In our cohort, orbital MRI has revealed inflammatory changes in 52% (11/21) of cases where imaging was available, indicating that extraocular muscles are commonly involved in ICI-induced StrAb-associated myositis. Notably, fluctuating ocular weakness and orbital MRI abnormalities may coexist in the same patient.^73^ Whether these ocular manifestations primarily reflect neuromuscular transmission defects, muscle inflammation, or both remains unclear.

The concurrence of StrAb-associated myositis in MG patients contributes to muscle weakness both through direct cellular injury and by fueling the autoimmune response against AChRs. Recognizing the involvement of cellular immune components is essential for accurate diagnosis and for guiding appropriate therapeutic strategies. The fact that StrAb-associated MG-myositis overlap syndrome represents the most common myotoxicity in ICI-treated patients raises the possibility that immune checkpoint agonists may have therapeutic value in dampening excessive immune activation, particularly in non-cancer settings.

Typically, the cellular response diminishes after discontinuation of immunotherapy and initiation of corticosteroids, as reflected by the rapid decline in CK levels and the absence of inflammatory infiltrates in muscle biopsies obtained after high-dose corticosteroids. Nevertheless, some patients continue to deteriorate clinically despite apparent suppression of the cellular inflammation, suggesting that secondary humoral mechanisms may underlie the later disease course. This notion is supported by complement deposition on muscle fibers in post-treatment biopsies lacking cellular infiltrates.^74^ Furthermore, the therapeutic efficacy of rituximab and complement inhibitors in ICI-induced myositis and myocarditis underscores the contribution of humoral components, which appear not only as downstream consequence of inflammation but also as direct pathogenic drivers.^74–80^ Rigorous prospective studies will be essential to validate these observations and to clarify the therapeutic potential of targeting humoral pathways in this setting.

A prior study identified anti-AChR antibody positivity as a predictor of adverse outcomes in ICI- induced myocarditis.^9^ In our cohort, however, this association was evident in ICI-induced myositis rather than myocarditis. This apparent discrepancy may reflect differences in case composition: whereas only 46% of myocarditis patients in our cohort had concomitant myositis, nearly all patients (87%) in the prior study did. Thus, the previously reported association may reflect a genuine relationship mediated primarily through myositis rather than myocarditis. Clinically, these findings highlight the potential value of anti-AChR antibody testing before and during the early cycles of ICI therapy. Notably, a recent study demonstrated that most patients who were anti-AChR-positive after ICI therapy had already been seropositive beforehand.^81^ Incorporating antibody testing into baseline assessments may therefore improve risk stratification and individualized management, though its role in routine practice requires validation in prospective studies.

Several limitations should be acknowledged. First, because our review is based entirely on case reports, publication bias is likely, and some patient data were incomplete or unverifiable. Second, the cases span nearly a century, during which diagnostic criteria, disease definitions, and treatment standards changed substantially. Although we addressed this by excluding cases that did not meet contemporary diagnostic criteria and by defining outcome windows based on stabilized mortality rates, variability remains. Third, antibody testing was performed only in selected patients, introducing measurable selection bias, though subgroup analyses suggested that its effect on estimated seropositivity rates was limited. Finally, many MSAs were only recently identified and therefore absent from older reports, underscoring the need for prospective studies to validate and refine these observations.

In conclusion, this systematic review characterizes a distinct clinicopathological subtype of myositis defined by StrAb positivity and strong linkage to thymoma. The data support a model in which thymoma-driven cytotoxic T-cell activity initiates muscle inflammation and promotes secondary AChR autoimmunity, producing the characteristic overlap of MG and myositis. The same pathophysiologic signature also appears in ICI-related neuromuscular toxicity, underscoring its translational relevance. Clinically, StrAbs and anti-AChR antibodies emerge as practical biomarkers for early diagnosis and risk assessment in patients susceptible to severe neuromuscular complications.

## Footnotes

### Data availability

All data needed to evaluate the conclusions of this study are included in the main text and/or the Supplementary Information. Data from primary studies are publicly available through the databases and search strategies described in Supplementary Appendix 1. A complete list of included studies is provided in Supplementary Appendix 2. Any additional materials related to this study are available from the corresponding author upon reasonable request.

### Code availability

The underlying R code for the present study can be accessed in a GitHub repository: https://github.com/jileil2/thymoma_case_reports.

## Acknowledgements

The authors thank Dr. Henry Kaminski (Department of Neurology & Rehabilitation Medicine, George Washington University), Dr. Andrew Mammen (National Institute of Arthritis and Musculoskeletal and Skin Diseases, National Institutes of Health), and Dr. Eric Lancaster (Department of Neurology, University of Pennsylvania Perelman School of Medicine) for their constructive comments on this study.

## Contributions

J.Luo takes full responsibility for the work as a whole, including study conception and design, manuscript drafting, and the decision to submit the manuscript. J.Luo and J.S. conducted study screening and data extraction. J.Lin and H.W. conducted the statistical analyses. All authors approved the final manuscript.

## References

1. Lundberg, I.E., et al. Idiopathic inflammatory myopathies. Nat Rev Dis Primers 7, 86 (2021).

2. McHugh, N.J. & Tansley, S.L. Autoantibodies in myositis. Nat Rev Rheumatol 14, 290–302 (2018).

3. Ludwig, R.J., et al. Mechanisms of Autoantibody-Induced Pathology. Front Immunol 8, 603 (2017).

4. Uchio, N., et al. Inflammatory myopathy with myasthenia gravis: Thymoma association and polymyositis pathology. Neurol Neuroimmunol Neuroinflamm 6, e535 (2019).

5. Suzuki, S., et al. Autoimmune targets of heart and skeletal muscles in myasthenia gravis. Arch Neurol 66, 1334–1338 (2009).

6. Garibaldi, M., et al. Muscle involvement in myasthenia gravis: Expanding the clinical spectrum of Myasthenia-Myositis association from a large cohort of patients. Autoimmun Rev 19, 102498 (2020).

7. Mao, Z.F., Mo, X.A., Qin, C., Lai, Y.R. & Hackett, M.L. Incidence of thymoma in myasthenia gravis: a systematic review. J Clin Neurol 8, 161–169 (2012).

8. Seki, M., et al. Inflammatory myopathy associated with PD-1 inhibitors. J Autoimmun 100, 105–113 (2019).

9. Fenioux, C., et al. Thymus alterations and susceptibility to immune checkpoint inhibitor myocarditis. Nat Med 29, 3100–3110 (2023).

10. Page, M.J., et al. The PRISMA 2020 statement: an updated guideline for reporting systematic reviews. BMJ 372, n71 (2021).

11. Nicolle, M.W. Myasthenia Gravis and Lambert-Eaton Myasthenic Syndrome. Continuum (Minneap Minn) 22, 1978–2005 (2016).

12. Bohan, A. & Peter, J.B. Polymyositis and dermatomyositis (second of two parts). N Engl J Med 292, 403–407 (1975).

13. Bohan, A. & Peter, J.B. Polymyositis and dermatomyositis (first of two parts). N Engl J Med 292, 344–347 (1975).

14. Bonaca, M.P., et al. Myocarditis in the Setting of Cancer Therapeutics: Proposed Case Definitions for Emerging Clinical Syndromes in Cardio-Oncology. Circulation 140, 80–91 (2019).

15. Board, W.C.o.T.E. *Thoracic tumours*, (International Agency for Research on Cancer, Lyon (France), 2021).

16. Hothorn, T., Bretz, F. & Westfall, P. Simultaneous inference in general parametric models. Biom J 50, 346–363 (2008).

17. Terayama, A., et al. A case of thymoma-associated myasthenia gravis accompanied with myositis showing the clusters of histiocyte along the fascicles in perimysium. Clin Neurol Neurosurg 229, 107715 (2023).

18. Carroll, G.J., Will, R.K., Peter, J.B., Garlepp, M.J. & Dawkins, R.L. Penicillamine induced polymyositis and dermatomyositis. J Rheumatol 14, 995–1001 (1987).

19. Uchio, N., et al. Pembrolizumab on pre-existing inclusion body myositis: a case report. BMC Rheumatol 4, 48 (2020).

20. Matsubara, S., Seki, M., Suzuki, S., Komori, T. & Takamori, M. Tertiary lymphoid organs in the inflammatory myopathy associated with PD-1 inhibitors. J Immunother Cancer 7, 256 (2019).

21. O’Hare, M. & Guidon, A.C. Peripheral nervous system immune-related adverse events due to checkpoint inhibition. Nat Rev Neurol 20, 509–525 (2024).

22. Chan, K.H., Lachance, D.H., Harper, C.M. & Lennon, V.A. Frequency of seronegativity in adult-acquired generalized myasthenia gravis. Muscle Nerve 36, 651–658 (2007).

23. Sanders, D.B., Andrew, P.I., Howard, J.F., Jr. & Massey, J.M. Seronegative myasthenia gravis. Neurology 48(Suppl 5), S40–S45 (1997).

24. Gougis, P., et al. Clinical spectrum and evolution of immune-checkpoint inhibitors toxicities over a decade-a worldwide perspective. EClinicalMedicine 70, 102536 (2024).

25. Romi, F., Skeie, G.O., Gilhus, N.E. & Aarli, J.A. Striational antibodies in myasthenia gravis: reactivity and possible clinical significance. Arch Neurol 62, 442–446 (2005).

26. Somnier, F.E., Skeie, G.O., Aarli, J.A. & Trojaborg, W. EMG evidence of myopathy and the occurrence of titin autoantibodies in patients with myasthenia gravis. Eur J Neurol 6, 555–563 (1999).

27. Russell, D.S. Histological changes in the striped muscles in myasthenia gravis. J Pathol Bacteriol 65, 279–289 (1953).

28. Strobel, P., et al. Corticomedullary differentiation and maturational arrest in thymomas. Histopathology 64, 557–566 (2014).

29. Yasumizu, Y., et al. Myasthenia gravis-specific aberrant neuromuscular gene expression by medullary thymic epithelial cells in thymoma. Nat Commun 13, 4230 (2022).

30. Morimoto, J., et al. Aire suppresses CTLA-4 expression from the thymic stroma to control autoimmunity. Cell Rep 38, 110384 (2022).

31. Arbour, K.C., et al. Expression of PD-L1 and other immunotherapeutic targets in thymic epithelial tumors. PLoS One 12, e0182665 (2017).

32. Santoni, G., et al. High CTLA-4 expression correlates with poor prognosis in thymoma patients. Oncotarget 9, 16665–16677 (2018).

33. Hubert, F.X., et al. Aire regulates the transfer of antigen from mTECs to dendritic cells for induction of thymic tolerance. Blood 118, 2462–2472 (2011).

34. Savino, W., et al. Thymoma epithelial cells secrete thymic hormone but do not express class II antigens of the major histocompatibility complex. J Clin Invest 76, 1140–1146 (1985).

35. Buckley, C., Douek, D., Newsom-Davis, J., Vincent, A. & Willcox, N. Mature, long-lived CD4+ and CD8+ T cells are generated by the thymoma in myasthenia gravis. Ann Neurol 50, 64–72 (2001).

36. Hoffacker, V., et al. Thymomas alter the T-cell subset composition in the blood: a potential mechanism for thymoma-associated autoimmune disease. Blood 96, 3872–3879 (2000).

37. Takeuchi, Y., et al. Accumulation of immature CD3-CD4+CD8- single-positive cells that lack CD69 in epithelial cell tumors of the human thymus. Cell Immunol 161, 181–187 (1995).

38. Radovich, M., et al. The Integrated Genomic Landscape of Thymic Epithelial Tumors. Cancer Cell 33, 244–258 e210 (2018).

39. Schultz, A., et al. Neurofilament is an autoantigenic determinant in myasthenia gravis. Ann Neurol 46, 167–175 (1999).

40. Marx, A., et al. Expression of neurofilaments and of a titin epitope in thymic epithelial tumors. Implications for the pathogenesis of myasthenia gravis. Am J Pathol 148, 1839–1850 (1996).

41. Mencarelli, C., Magi, B., Marzocchi, B., Armellini, D. & Pallini, V. Evolution of the "titin epitope" in neurofilament proteins. Comp Biochem Physiol B 100, 741–744 (1991).

42. Wilisch, A., et al. [Molecular mimicry between neurofilaments and titin as the basis for autoimmunity towards skeletal muscle in paraneoplastic myasthenia gravis]. Verh Dtsch Ges Pathol 80, 261–266 (1996).

43. Buckley, C., Newsom-Davis, J., Willcox, N. & Vincent, A. Do titin and cytokine antibodies in MG patients predict thymoma or thymoma recurrence? Neurology 57, 1579–1582 (2001).

44. Meager, A., Vincent, A., Newsom-Davis, J. & Willcox, N. Spontaneous neutralising antibodies to interferon--alpha and interleukin-12 in thymoma-associated autoimmune disease. Lancet 350, 1596–1597 (1997).

45. Romi, F., et al. Titin and ryanodine receptor epitopes are expressed in cortical thymoma along with costimulatory molecules. J Neuroimmunol 128, 82–89 (2002).

46. Gilhus, N.E., et al. Antigen presentation by thymoma epithelial cells from myasthenia gravis patients to potentially pathogenic T cells. J Neuroimmunol 56, 65–76 (1995).

47. Takahashi, K., Monden, Y., Saito, S., Kamamura, Y. & Uyama, T. Myasthenia gravis induces the activation and maturation of lymphocytes in thymoma. J Clin Immunol 16, 190–197 (1996).

48. Zamecnik, J., et al. Muscle lymphocytic infiltrates in thymoma-associated myasthenia gravis are phenotypically different from those in polymyositis. Neuromuscul Disord 17, 935–942 (2007).

49. Karner, J., et al. Anti-cytokine autoantibodies suggest pathogenetic links with autoimmune regulator deficiency in humans and mice. Clin Exp Immunol 171, 263–272 (2013).

50. Rodgaard, A., Nielsen, F.C., Djurup, R., Somnier, F. & Gammeltoft, S. Acetylcholine receptor antibody in myasthenia gravis: predominance of IgG subclasses 1 and 3. Clin Exp Immunol 67, 82–88 (1987).

51. Romi, F., et al. Complement activation by titin and ryanodine receptor autoantibodies in myasthenia gravis. A study of IgG subclasses and clinical correlations. J Neuroimmunol 111, 169–176 (2000).

52. Cufi, P., et al. Thymoma-associated myasthenia gravis: On the search for a pathogen signature. J Autoimmun 52, 29–35 (2014).

53. Song, Y., et al. Aberrant dendritic cell subsets in patients with myasthenia gravis and related clinical features. Neuroimmunomodulation (2023).

54. Tackenberg, B., et al. Expanded TCR Vbeta subsets of CD8(+) T-cells in late-onset myasthenia gravis: novel parallels with thymoma patients. J Neuroimmunol 216, 85–91 (2009).

55. Filosso, P.L., et al. Thymoma and the increased risk of developing extrathymic malignancies: a multicentre study. Eur J Cardiothorac Surg 44, 219–224; discussion 224 (2013).

56. Luo, J., et al. Main immunogenic region structure promotes binding of conformation- dependent myasthenia gravis autoantibodies, nicotinic acetylcholine receptor conformation maturation, and agonist sensitivity. J Neurosci 29, 13898–13908 (2009).

57. Loutrari, H., Kokla, A., Trakas, N. & Tzartos, S.J. Expression of human-Torpedo hybrid acetylcholine receptor (AChR) for analysing the subunit specificity of antibodies in sera from patients with myasthenia gravis (MG). Clin Exp Immunol 109, 538–546 (1997).

58. Wilisch, A., et al. Association of acetylcholine receptor alpha-subunit gene expression in mixed thymoma with myasthenia gravis. Neurology 52, 1460–1466 (1999).

59. Siara, J., Rudel, R. & Marx, A. Absence of acetylcholine-induced current in epithelial cells from thymus glands and thymomas of myasthenia gravis patients. Neurology 41, 128–131 (1991).

60. Shiono, H., et al. Spontaneous production of anti-IFN-alpha and anti-IL-12 autoantibodies by thymoma cells from myasthenia gravis patients suggests autoimmunization in the tumor. Int Immunol 15, 903–913 (2003).

61. Suzuki, S., et al. Cardiac involvements in myasthenia gravis associated with anti-Kv1.4 antibodies. Eur J Neurol 21, 223–230 (2014).

62. Xie, D., et al. An endogenous cholinergic system controls electrical conduction in the heart. Eur Heart J 46, 1232–1246 (2025).

63. Nakano, S. & Engel, A.G. Myasthenia gravis: quantitative immunocytochemical analysis of inflammatory cells and detection of complement membrane attack complex at the end-plate in 30 patients. Neurology 43, 1167–1172 (1993).

64. McKeon, A., Lennon, V.A., LaChance, D.H., Klein, C.J. & Pittock, S.J. Striational antibodies in a paraneoplastic context. Muscle Nerve 47, 585–587 (2013).

65. Shelly, S., et al. Clinical Utility of Striational Antibodies in Paraneoplastic and Myasthenia Gravis Paraneoplastic Panels. Neurology (2021).

66. Mikkelsen, A.W., Nilsson, A.C., Tenstad, H.B., Lillevang, S.T. & Asgari, N. Initial screening for neuronal autoantibodies and their putative impact on survival in patients with small- cell lung cancer. Thorac Cancer 15, 1350–1356 (2024).

67. Horta, E.S., et al. Neural autoantibody clusters aid diagnosis of cancer. Clin Cancer Res 20, 3862–3869 (2014).

68. Anderson, W., et al. PTPN22 R620W gene editing in T cells enhances low-avidity TCR responses. Elife 12 (2023).

69. Greve, B., et al. The autoimmunity-related polymorphism PTPN22 1858C/T is associated with anti-titin antibody-positive myasthenia gravis. Hum Immunol 70, 540–542 (2009).

70. Iwatsubo, T., Arahata, K., Motoyoshi, Y., Murayama, S. & Mannen, T. [Co-existence of clinical and immunohistochemical features of polymyositis and myasthenia gravis]. Rinsho Shinkeigaku 28, 913–918 (1988).

71. Noda, T., Kageyama, H., Miura, M., Tamura, T. & Ito, H. [A case of myasthenia gravis and myositis induced by pembrolizumab]. Rinsho Shinkeigaku 59, 502–508 (2019).

72. Wong, E.Y.T., et al. Immune checkpoint inhibitor-associated myositis and myasthenia gravis overlap: Understanding the diversity in a case series. Asia Pac J Clin Oncol 17, e262–e267 (2021).

73. Ozarczuk, T.R.A., Prentice, D.A., Kho, L.K. & vanHeerden, J. Checkpoint inhibitor myasthenia-like syndrome and myositis associated with extraocular muscle atrophy. J Clin Neurosci 71, 271–272 (2020).

74. Gosser, C., Al Bawaliz, A., Bahaj, W., Chesney, J. & Ranjan, S. Immune Checkpoint Inhibitor-Induced Myositis/Myasthenia Gravis Overlap. Cureus 15, e49007 (2023).

75. Nelke, C., et al. Immune Checkpoint Inhibition-Related Myasthenia-Myositis-Myocarditis Responsive to Complement Blockade. Neurol Neuroimmunol Neuroinflamm 11 (2024).

76. Zadeh, S., et al. Novel uses of complement inhibitors in myasthenia gravis-Two case reports. Muscle Nerve 69, 368–372 (2024).

77. Cuenca, J.A., et al. Management of respiratory failure in immune checkpoint inhibitors-induced overlap syndrome: a case series and review of the literature. BMC Anesthesiol 23, 310 (2023).

78. Jain, V., Remley, W., Bunag, C., Elfasi, A. & Chuquilin, M. Rituximab in Refractory Myositis and Acute Neuropathy Secondary to Checkpoint Inhibitor Therapy. Cureus 14, e25129 (2022).

79. Grewal, N.K.S., Maning, J., Gordon, L.I. & Akhter, N. Checkpoint inhibitor myocarditis with preceding immunosuppression and tolerance of sequential anthracycline therapy. BMJ Case Rep 17 (2024).

80. Aggarwal, N., Bianchini, D., Parkar, R. & Turner, J. Immunotherapy-Induced Overlap Syndrome: Myositis, Myasthenia Gravis, and Myocarditis-A Case Series. Case Rep Med 2024, 5399073 (2024).

81. Plomp, L., et al. Features of myositis and myasthenia gravis in patients treated with immune checkpoint inhibitors: a multicentric, retrospective cohort study. Lancet Reg Health Eur 50, 101192 (2025).

82. Qi, B., et al. Macrophage-Myofibroblast Transition as a Potential Origin for Skeletal Muscle Fibrosis After Injury via Complement System Activation. J Inflamm Res 17, 1083–1094 (2024).

