## Supplementary Data for "Striational Antibody-Associated Myositis – Bridging the Gap between Thymoma and Myasthenia Gravis: A Systematic Review"

### Appendix 1: Systematic Review Protocol

***Search Strategy***

We conducted comprehensive PubMed searches for eligible studies, without language restrictions, from database inception to 2024. The complete search strategy, including all terms and Boolean combinations, is provided below. Reference lists of included and related articles were manually screened to identify additional eligible studies. Because many early reports (particularly before the 1970s) are not indexed in PubMed, manual reference screening was emphasized.

This review followed the PRISMA 2020 reporting guidelines. The protocol was not prospectively registered in PROSPERO because the study was initially designed as a narrative review but later evolved into a systematic review after recognizing the feasibility of structured data synthesis. The finalized protocol is provided here to ensure transparency and reproducibility.

*Databases*

- PubMed
- J-STAGE
- J-Global

*Other Sources*

- Reference lists of included studies
- Manual citation tracking

***Search Terms***

1. Myasthenia gravis (MG) keywords:
   (myasthenia) OR (myasthenic)
2. Myositis keywords:
   (myopathy) OR (myopathies) OR (myositis) OR (myocarditis) OR (dermatomyositis) OR (polymyositis)
3. Thymic epithelial tumor (TET) keywords:
   (thymoma) OR (thymic carcinoma) OR (thymic epithelial tumor) OR (TET)
4. Immune checkpoint inhibitor (ICI) keywords:
   (immune checkpoint inhibitor) OR (ICI) OR (pembrolizumab) OR (ipilimumab) OR (nivolumab) OR (atezolizumab) OR (cemiplimab) OR (avelumab) OR (durvalumab) OR (tremelimumab) OR (immune checkpoint inhibitors) OR (ICIs)

***Boolean Combinations***

A | PubMed

1. myasthenia gravis keywords AND myositis keywords NOT immune checkpoint inhibitor keywords (inception–September 30, 2024)
2. thymic epithelial tumor keywords AND myositis keywords NOT myasthenia gravis keywords NOT immune checkpoint inhibitor keywords (inception–October 15, 2024)
3. myasthenia gravis keywords OR myositis keywords AND immune checkpoint inhibitor keywords (January 1, 2010–May 31, 2024)
4. (“thymoma” AND “regression” AND “spontaneous”) (inception–July 23, 2025)

B | J-STAGE

1. (“thymoma” AND “regression” AND “spontaneous”)
2. ("自然縮小" AND "胸腺腫")

C | J-Global

1. (“thymoma” AND “regression” AND “spontaneous”)
2. ("自然縮小" AND "胸腺腫")

***Eligibility Criteria***

*Inclusion:*
Published case reports or case series (including conference abstracts) providing original data on age, sex, symptoms, serological findings, neurophysiology, pathology, treatments, and outcomes. Eligible patients included those with:

- Concomitant idiopathic MG and idiopathic inflammatory myopathy (IIM).
- Isolated IIM associated with TETs (without MG).
- Spontaneous regression of thymoma.
- ICI-induced MG (including MG-like syndrome), myositis, or myocarditis.

*Exclusion:*

- Lambert-Eaton myasthenic syndrome.
- Congenital myasthenic syndromes.
- ICI-induced cases were excluded from analyses focused on idiopathic MG–IIM overlap.

***Diagnostic Definitions***

- *Myasthenia gravis (MG)*: Clinical weakness plus ≥1 of the following: MG-specific antibodies (anti-AChR, anti-MuSK, anti-LRP4), response to acetylcholinesterase inhibitors, decrement on repetitive nerve stimulation (RNS), or increased jitter on single-fiber electromyography (SFEMG).

- *Subclinical MG*: Antibody positivity without clinical manifestations.
- *Myositis*: Based on modified Bohan and Peter criteria—clinical, biochemical, electrophysiological, histological, and MRI evidence, with or without myositis-specific antibodies (MSAs).
- *Myocarditis*: Compatible symptoms with ECG, imaging, biopsy evidence, or elevated cardiac biomarkers.
- *TETs*: Diagnosed radiologically and confirmed histopathologically, classified according to the WHO histological system (retrospectively applied where necessary).

***Study Selection Process***

Searches were conducted by a single reviewer (J.S.). Records were imported into Covidence for screening. Two reviewers (J.S. and J. Luo) independently screened titles and abstracts, followed by full-text review for eligibility. Discrepancies were resolved by consensus.

Potential duplicate reports were carefully evaluated to avoid double counting of cases. Duplicates were identified based on (1) explicit author statements indicating overlapping patient cohorts, and (2) concordance in authorship, key demographic variables (e.g., age, sex), diagnostic features (e.g., autoantibody profiles, immune checkpoint inhibitor type, comorbidities such as cancer type or myocarditis), and quantitative data (e.g., antibody titers, serum creatine kinase levels). When duplicates were confirmed, data from all relevant reports were consolidated for analysis. When duplication remained suspected but could not be definitively confirmed, only the most comprehensive or most recent publication was retained for inclusion.

Non-English articles were translated using ChatGPT, and key data were cross-verified using Google Translate to ensure accuracy.

***Data Extraction***

Two reviewers (J.S. and J. Luo) independently extracted the following variables:

- Study identifiers (PubMed ID, year, country)
- Patient demographics (age, sex)
- Diagnosis and disease subtype
- Laboratory and pathological findings
- Antibody profiles (MSAs, StrAbs, MG antibodies)
- Treatments and clinical outcomes

Data were entered into a standardized Excel sheet. Discrepancies were resolved by discussion.

***Risk of Bias Assessment***

Formal risk-of-bias assessment was not applicable given the nature of the included evidence (case reports and case series). However, all extracted data underwent independent verification by both reviewers to ensure completeness and accuracy.

***Strategy for Data Synthesis***

1. Overview

Given the nature of the included evidence—individual case reports and small case series describing rare autoimmune neuromuscular syndromes—the data synthesis was primarily descriptive and exploratory rather than meta-analytic. The objective was to identify consistent clinical, serological, and pathological patterns and to explore associations with outcomes.

2. Descriptive Synthesis

All extracted variables were summarized narratively and tabulated:

- Categorical variables (e.g., sex, antibody status, thymoma presence, outcome categories) were summarized as frequencies and percentages.
- Continuous variables (e.g., age, enzyme levels) were summarized as medians and interquartile ranges (IQRs) or means and standard deviations.
- Clinical outcomes were categorized as:
    1. *Remission* – complete resolution of symptoms without ongoing therapy.
    2. *Pharmacological remission* – symptom resolution maintained under continued immunosuppressive therapy.
    3. *Minimal manifestations* – mild residual symptoms without functional impairment.
    4. *Death* – death related to disease or treatment.

These categories were used consistently across subgroups to enable comparison of disease severity and prognosis. For the ICI-related analyses, specific modifications were introduced to more accurately capture clinical outcomes.

3. Exploratory Quantitative Analyses

Where sufficient data were available, exploratory analyses assessed associations between clinical, serological, and pathological variables and outcomes.

- Continuous variables: Compared across groups using OLS regression (means) or quantile regression (medians).
- Categorical/ordinal variables:
    - Logistic regression for binary variables (e.g., death vs. survival).

    - Multinomial regression for variables with more than two categories (e.g., “Remission,” “Pharmacological remission,” “Minimal manifestations,” “Death”).
    - Covariates included age, sex, antibody status, thymoma presence, and cardiac involvement.
    - Adjusted odds ratios (ORs) and 95% confidence intervals (CIs) were reported when feasible.
- Multiple group comparisons: Adjustments were applied when comparing more than two disease categories (e.g., ICI-induced myositis vs. ICI-induced myocarditis vs. ICI-induced myositis-myocarditis overlap).

4. Handling of Missing or Incomplete Data

Given the variable reporting quality of case reports and small series, missing or incomplete data were anticipated. The following procedures were applied:

- Inclusion: Cases with missing variables were retained for qualitative synthesis but excluded from quantitative analyses requiring unavailable data.
- Age ranges: When age was reported as a range, the midpoint of the range was assigned as an approximation.
- No formal imputation: Missing values were not statistically imputed.
- Verification: Two reviewers independently checked data consistency and plausibility.

5. Integration of Findings

Findings from clinical, serological, and histopathological domains were integrated to:

- Characterize the phenotypic spectrum of MG-IIM overlap.
- Explore associations between striational antibody positivity, thymoma histology, cardiac involvement, and outcomes.
- Compare idiopathic and ICI-induced presentations to identify shared mechanisms.

The synthesis aimed to characterize a previously unrecognized clinicopathological subtype of myositis and to develop a conceptual framework linking thymoma-driven autoimmunity to myotoxic and neuromuscular pathology.

6. Subgroup Analyses

Analyses were stratified by:

- Age of onset: early-onset MG and late-onset MG.
- Myositis subtype: polymyositis, giant cell myositis, dermatomyositis, orbital myositis, and others.

- TET histological subtype: thymoma (A, AB, B1-B3) and thymic carcinoma.
- Antibody profile (StrAb, MSA, anti-AChR, anti-MuSK, anti-LRP4).
- Cardiac involvement (present vs. absent).
- Etiology (idiopathic vs. ICI-induced).

7. Presentation of Results

Findings were summarized in structured tables and visualized using bar charts and forest-style plots for odds ratios.

### Appendix 2: List of Included Studies

#### *A. Concomitant IIM and MG*

1. Ago, T., *et al.* Dermatomyositis associated with invasive thymoma. *Intern Med* **38**, 155-159 (1999).
2. Akatsuka, S. & Torigata, C. [Malignant thymoma associated with polymyositis--an autopsy case]. *Nihon Rinsho* **35**, 1788-1792 (1977).
3. Ansevin, C.F. & Agamanolis, D.P. Rippling muscles and myasthenia gravis with rippling muscles. *Arch Neurol* **53**, 197-199 (1996).
4. Antoine, J.C., Camdessanche, J.P., Absi, L., Lassabliere, F. & Feasson, L. Devic disease and thymoma with anti-central nervous system and antithymus antibodies. *Neurology* **62**, 978-980 (2004).
5. Avni, I., Sharabi, Y., Sadeh, M. & Buchman, A.S. Eosinophilia, myositis, and myasthenia gravis associated with a thymoma. *Muscle Nerve* **34**, 242-245 (2006).
6. Barraquerbordas, L., Peresserra, J. & Salisachsrowe, P. [Focal nodular chronic polymyositis with myasthenic syndrome]. *Rev Neurol (Paris)* **113**, 69-73 (1965).
7. Barton, F.E. & Branch, C.F. Myasthenia gravis: report of a case with necropsy. *J Ame Med Assoc* **109**, 2044-2048 (1937).
8. Behan, W.M., Behan, P.O. & Doyle, D. Association of myasthenia gravis and polymyositis with neoplasia, infection and autoimmune disorders. *Acta Neuropathol* **57**, 221-229 (1982).
9. Bonduelle, M. & Bouygues, P. [Myasthenia and polymyositis: a case of polymyositis with thymoma, myasthenic syndrome in myositis; myasthenia, a syndrome or disease]. *Presse Med (1893)* **63**, 1572-1575 (1955).
10. Bourgeois-Droin, C., *et al.* [Thymoma associated with myasthenia, erythroblastopenia, myositis and giant cell myocarditis. One case (author's transl)]. *Nouv Presse Med* **10**, 2097-2098, 2103-2094 (1981).
11. Burke, J.S., Medline, N.M. & Katz, A. Giant cell myocarditis and myositis. Associated with thymoma and myasthenia gravis. *Arch Pathol* **88**, 359-366 (1969).
12. Buscaino, G.A., Caruso, G. & Meletti, M. [Apropos of 2 cases with the clinical picture of myasthenia and histopathologic findings of polymyositic changes. Presence of thymoma in one of them deteriorating after thymectomy]. *Acta Neurol (Napoli)* **22**, 305-346 (1967).
13. Butte, M.J., Haines, C., Bonilla, F.A. & Puck, J. IL-7 receptor deficient SCID with a unique intronic mutation and post-transplant autoimmunity due to chronic GVHD. *Clin Immunol* **125**, 159-164 (2007).
14. Caquet, R., *et al.* [Myasthenia-like polymyositis, thymoma, smooth muscle degeneration]. *Nouv Presse Med* **3**, 743-746 (1974).
15. Carroll, G.J., Will, R.K., Peter, J.B., Garlepp, M.J. & Dawkins, R.L. Penicillamine induced polymyositis and dermatomyositis. *J Rheumatol* **14**, 995-1001 (1987).
16. Chiavistelli, P., Cei, M., Carmignani, G., Bartolomei, C. & Mumoli, N. Pseudoischemic electrocardiogram in myasthenia gravis with thymoma: reversibility after thymectomy. *Clin Cardiol* **32**, E75-78 (2009).
17. Chitasombat, M.N., Ratchatanawin, N. & Visessiri, Y. Disseminated extrapulmonary Legionella pneumophila infection presenting with panniculitis: case report and literature review. *BMC Infect Dis* **18**, 467 (2018).
18. Damato, V., *et al.* When myasthenia gravis is not all. *J Neurol* **261**, 835-836 (2014).
19. Davis, C.J. & Gallai, V. Myasthenia progressing to polymyositis with respiratory failure. *Acta Neurol (Napoli)* **1**, 365-370 (1979).

20. de Jongste, M.J., Oosterhuis, H.J. & Lie, K.I. Intractable ventricular tachycardia in a patient with giant cell myocarditis, thymoma and myasthenia gravis. *Int J Cardiol* **13**, 374-378 (1986).
21. de Lacroix-Szmania, I., Cacoub, P., Paugam, B., Grunenwald, D. & Godeau, P. [Association of polymyositis, myasthenia gravis and thymoma]. *Rev Med Interne* **17**, 344-345 (1996).
22. De Reuck, J., Thiery, E., De Coster, W. & Van Der Eecken, H. Myasthenic syndrome in polymyositis. *Eur Neurol* **14**, 275-284 (1976).
23. Demichelis, C., *et al.* Neuromuscular complications following targeted therapy in cancer patients: beyond the immune checkpoint inhibitors. Case reports and review of the literature. *Neurol Sci* **42**, 1405-1409 (2021).
24. Diaco, M., *et al.* Association of myasthenia gravis and antisynthetase syndrome: a case report. *Int J Immunopathol Pharmacol* **17**, 395-399 (2004).
25. Essigman, W.K. Multiple side effects of penicillamine therapy in one patient with rheumatoid arthritis. *Ann Rheum Dis* **41**, 617-620 (1982).
26. Ezzatian-Ahar, S., Pedersen, E.G., Schroder, H.D., Horn, H.C. & Gaist, D. [Paraneoplastic myasthenia gravis and polymyositis secondary to a thymoma in a young woman]. *Ugeskr Laeger* **178**(2016).
27. Finsterer, J., Stollberger, C. & Ho, C.Y. Respiratory insufficiency from myasthenia gravis and polymyositis due to malignant thymoma triggering Takotsubo syndrome. *Int J Neurosci* **128**, 1207-1210 (2018).
28. Fraenkel, P.G., *et al.* Induction of myasthenia gravis, myositis, and insulin-dependent diabetes mellitus by high-dose interleukin-2 in a patient with renal cell cancer. *J Immunother* **25**, 373-378 (2002).
29. Frasson, E., *et al.* Statin-associated necrotizing autoimmune myopathy with concurrent myasthenia gravis. *Clin Case Rep* **9**, e03925 (2021).
30. Funabashi, H., *et al.* [An autopsy case of a patient with myasthenia gravis who showed various symptoms of collagen diseases and complicated with malignant thymoma]. *Ryumachi* **29**, 185-191 (1989).
31. Furuta, C., *et al.* A case of thymoma-associated multiorgan autoimmunity including polymyositis and myocarditis. *Surg Case Rep* **7**, 226 (2021).
32. Fuse, K., *et al.* [A case of anti-acetylcholine receptor antibody-positive ocular myasthenia gravis with anti-titin antibody and anti-Kv1.4 antibody positive inflammatory myopathy]. *Rinsho Shinkeigaku* **63**, 830-835 (2023).
33. Garibaldi, M., *et al.* Muscle involvement in myasthenia gravis: Expanding the clinical spectrum of Myasthenia-Myositis association from a large cohort of patients. *Autoimmun Rev* **19**, 102498 (2020).
34. Ghiringhelli, P., Chelazzi, P., Chelazzi, G., Bellintani, C. & Rania, S. [Multiorgan autoimmune syndrome: case report]. *Ann Ital Med Int* **18**, 51-55 (2003).
35. Giallafos, E., Zouvelou, V., Maurogeni, S. & Stamboulis, E. Subclinical cardiac involvement in thymomatous Myasthenia Gravis. *Hellenic J Cardiol* **57**, 345-347 (2016).
36. Gidron, A., Quadrini, M., Dimov, N. & Argiris, A. Malignant thymoma associated with fatal myocarditis and polymyositis in a 32-year-old woman with a history of hairy cell leukemia. *Am J Clin Oncol* **29**, 213-214 (2006).
37. Greenberg, S.A. Acquired rippling muscle disease with myasthenia gravis. *Muscle Nerve* **29**, 143-146 (2004).
38. Harati, Y. & Patten, B.M. Prednisone use in concurrent autoimmune diseases. *Arch Neurol* **36**, 103-106 (1979).
39. Hasbani, G.E., *et al.* Eosinophilic Orbital Myositis Superseding Ocular Myasthenia. *Mediterr J Rheumatol* **35**, 192-194 (2024).

40. Hassel, B., Gilhus, N.E., Aarli, J.A. & Skogen, O.R. Fulminant myasthenia gravis and polymyositis after thymectomy for thymoma. *Acta Neurol Scand* **85**, 63-65 (1992).
41. Hatakeyama, S. & Inui, S. On a case of unusual triad: thymoma, giant-cell myocarditis and myasthenia gravis. *Bull Tokyo Med Dent Univ* **11**, 1-16 (1964).
42. Hausmanowa-Petrusewicz, I., Blaszczyk, M. & Jablonska, S. Coexistence of scleromyositis associated with PM-Scl antibody and myasthenia. *Neuromuscul Disord* **5**, 145-147 (1995).
43. Hengstman, G.J., Drost, G., Wagenaar, M. & van Engelen, B.G. Persistent increased risk for thymoma in myasthenia gravis associated with myositis. *Muscle Nerve* **34**, 251-252 (2006).
44. Hill, E.K., King, P.H. & Hughey, L.C. Dermatomyositis and concomitant overlap myasthenic syndrome: a rare presentation. *J Am Acad Dermatol* **65**, e150-152 (2011).
45. Hofstad, H.*, et al.* [Myasthenia gravis with myocarditis and sudden death]. *Tidsskr Nor Laegeforen* **107**, 1322-1323, 1345 (1987).
46. Hogg, E.J., Lewis, R.A., Bannykh, S. & Tagliati, M. Head drop in Parkinson's disease complicated by myasthenia gravis and myopathy. *J Neurol Sci* **376**, 216-218 (2017).
47. Huang, C.*, et al.* Case report: Multi-antibody-positive myasthenia gravis concomitant myositis associated with thymoma. *Front Immunol* **15**, 1423547 (2024).
48. Huang, G., Zhou, X. & Yao, D. Report of a case of necrotizing autoimmune myopathy with thymoma-associated myasthenia gravis. *Int J Neurosci* **130**, 1178-1181 (2020).
49. Huang, K.*, et al.* Concurrent inflammatory myopathy and myasthenia gravis with or without thymic pathology: A case series and literature review. *Semin Arthritis Rheum* **48**, 745-751 (2019).
50. Ikeda, Y.*, et al.* Giant cell myocarditis associated with multiple autoimmune disorders following highly active antiretroviral therapy for human immunodeficiency virus type 1 infection. *Int J Cardiol* **206**, 79-81 (2016).
51. Ikeda, Y., Tanaka, M., Mizushima, K. & Okamoto, K. A case of eosinophilic polymyositis complicated by myasthenia gravis. *Muscle Nerve* **21**, 1356-1358 (1998).
52. Illac, C., Boudat, A.M., Larrieu, J.M. & Delisle, M.B. [Giant cell myositis and myasthenia gravis: a case report]. *Ann Pathol* **33**, 53-56 (2013).
53. Inoue, M.*, et al.* [Concurrence of myasthenia gravis, polymyositis, thyroiditis and eosinophilia in a patient with type B1 thymoma]. *Rinsho Shinkeigaku* **47**, 423-428 (2007).
54. Iqbal, S.M., Burns, L. & Zhi, C. Granulomatous Myositis Associated with Myasthenia Gravis: A Rare Case. *Cureus* **11**, e5090 (2019).
55. Iwatsubo, T., Arahata, K., Motoyoshi, Y., Murayama, S. & Mannen, T. [Co-existence of clinical and immunohistochemical features of polymyositis and myasthenia gravis]. *Rinsho Shinkeigaku* **28**, 913-918 (1988).
56. Jan, V.*, et al.* [D-penicillamine-induced pemphigus, polymyositis and myasthenia]. *Ann Dermatol Venereol* **126**, 153-156 (1999).
57. Jasim, S. & Shaibani, A. Nonsarcoid granulomatous myopathy: two cases and a review of literature. *Int J Neurosci* **123**, 516-520 (2013).
58. Jesel, M., Stoebner, P., Zenglein, J.P. & Isch, F. [Myastheniform syndrome and polymyositis. Clinical, electromyographic and ultrastructural correlations in 2 cases]. *Rev Otoneuroophtalmol* **41**, 175-183 (1969).
59. Jimenez, J.C., Lucas, C.G., LaHue, S.C. & Sharpe, B.A. Giant cell myositis associated with metastatic thymoma and granulomatous hypercalcaemia. *BMJ Case Rep* **15**(2022).
60. Jin, J., Isfort, M.C., Ascherman, D.P. & Lacomis, D. Granulomatous Myositis Associated With Acetylcholine Receptor Antibodies Without Clinical Myasthenia. *J Clin Neuromuscul Dis* **23**, 49-52 (2021).

61. Jordan, B., Eger, K. & Zierz, S. [Polymyositis associated with thymoma]. *Nervenarzt* **80**, 708-711 (2009).
62. Kakutani, T. & Yoshizawa, M. Myasthenia gravis with inclusion body myositis: A case report. *Mod Rheumatol Case Rep* **8**, 83-85 (2023).
63. Kanatani, M., Adachi, T., Sakata, R., Watanabe, Y. & Hanajima, R. [A case of sporadic late-onset nemaline myopathy associated with myasthenia gravis positive for anti-titin antibody and anti-Kv1.4 antibody]. *Rinsho Shinkeigaku* **60**, 489-494 (2020).
64. Kanbayashi, T., *et al.* Myasthenia gravis with inflammatory myopathy without elevation of creatine kinase. *Neuromuscul Disord* **31**, 570-573 (2021).
65. Kanzato, N., Nakasone, I., Sunagawa, O., Komine, Y. & Fukiyama, K. [Giant-cell myocarditis without a symptom of heart failure seen in a patient with myasthenia gravis and concurrent Hashimoto's disease]. *Rinsho Shinkeigaku* **41**, 813-817 (2001).
66. Katabami, S., *et al.* Polymyositis associated with thymoma and the subsequent development of pure red cell aplasia. *Intern Med* **34**, 569-573 (1995).
67. Ketz, E., Fopp, M., Weissert, M. & Bekier, A. Polymyositis, myasthenic syndrome and thymoma in a patient with defective cell-mediated immunity. *Acta Neurol Belg* **79**, 469-474 (1979).
68. Kidher, E.S., Briceno, N., Taghi, A. & Chukwuemeka, A. An interesting collection of paraneoplastic syndromes in a patient with a malignant thymoma. *BMJ Case Rep* **2012**(2012).
69. Kim, J.H., Jang, H., Kwon, H.J., Suh, Y.L. & Min, J.H. Thymoma-Associated Paraneoplastic Myositis, Presenting with Rapidly Progressive Muscle Contractures. *J Clin Neurol* **17**, 496-498 (2021).
70. Klein, H.O. & Lennartz, K.J. [On syntrophy of myasthenia gravis, polymyositis, myocarditis and thymoma; at the same time a contribution to pathogenesis]. *Dtsch Med Wochenschr* **91**, 1727-1730 (1966).
71. Klein, J.J., Gottlieb, A.J., Mones, R.J., Appel, S.H. & Osserman, K.E. Thymoma and Polymyositis. Onset of Myasthenia Gravis after Thymectomy: Report of Two Cases. *Arch Intern Med* **113**, 142-152 (1964).
72. Ko, K.F., Ho, T. & Chan, K.W. Autoimmune chronic active hepatitis and polymyositis in a patient with myasthenia gravis and thymoma. *J Neurol Neurosurg Psychiatry* **59**, 558-559 (1995).
73. Kobayashi, T., *et al.* A patient with Graves' disease, myasthenia gravis, and polymyositis. *Thyroid* **7**, 631-632 (1997).
74. Kolsi, R., Bahloul, Z., Hachicha, J., Gouiaa, R. & Jarraya, A. [Dermatopolymyositis induced by D-penicillamine in rheumatoid polyarthritis. Apropos of 1 case with review of the literature]. *Rev Rhum Mal Osteoartic* **59**, 341-344 (1992).
75. Kon, T., *et al.* Giant cell polymyositis and myocarditis associated with myasthenia gravis and thymoma. *Neuropathology* **33**, 281-287 (2013).
76. Korner, F. & Regli, F. [On the syntropy of myasthenia and myositis]. *Dtsch Med Wochenschr* **90**, 1950-1954 (1965).
77. Kornizky, Y., Heller, I., Isakov, A., Shapira, I. & Topilsky, M. Dysphagia with multiple autoimmune disease. *Clin Rheumatol* **19**, 321-323 (2000).
78. Kosmorsky, G.S., Mehta, N., Mitsumoto, H. & Prayson, R. Intermittent esotropia associated with rippling muscle disease. *J Neuroophthalmol* **15**, 147-151 (1995).
79. Koul, D., *et al.* Fulminant giant cell myocarditis and cardiogenic shock: an unusual presentation of malignant thymoma. *Cardiol Res Pract* **2010**, 185896 (2010).
80. Lambrianides, S., *et al.* A novel case of inclusion body myositis and myasthenia gravis. *Neuromuscul Disord* **29**, 771-775 (2019).

81. Lane, R.J.M., *et al.* Thymectomy in polymyositis. *Lancet* **323**, 626-627 (1984).
82. Lauletta, A., *et al.* Distal upper limb involvement in myasthenia-myositis association. *Neurol Sci* **44**, 719-722 (2023).
83. Lee, A.B. & Thurston, R.S. Malignant thymoma and myasthenia gravis presenting as polymyositis: a case report. *J La State Med Soc* **160**, 286-288 (2008).
84. Limaye, K., Vallurupalli, S. & Lee, R.W. Myasthenia of the Heart. *Am J Med* **129**, e19-21 (2016).
85. Lin, J., *et al.* Giant cell polymyositis associated with myasthenia gravis and thymoma. *J Clin Neurosci* **21**, 2252-2254 (2014).
86. Lucchini, M., *et al.* Long-term Follow-up and Muscle Imaging Findings in Brachio-Cervical Inflammatory Myopathy. *Neurol Neuroimmunol Neuroinflamm* **8**(2021).
87. Maaroufi, A., Assoufi, N., Essaoudi, M.A. & Fatihi, J. Thymoma may explain the confusion: a case report. *J Med Case Rep* **15**, 616 (2021).
88. Maekawa, R., Shibuya, H., Hideyama, T. & Shiio, Y. [A case of myasthenia gravis with invasive thymoma associated with diffuse panbronchiolitis, alopecia, dysgeusia, cholangitis and myositis]. *Rinsho Shinkeigaku* **54**, 703-708 (2014).
89. Martyn, J.B., Wong, M.J. & Huang, S.H. Pulmonary and neuromuscular complications of mixed connective tissue disease: a report and review of the literature. *J Rheumatol* **15**, 703-705 (1988).
90. Mathis, S., *et al.* Simultaneous Combined Myositis, Inflammatory Polyneuropathy, and Overlap Myasthenic Syndrome. *Case Rep Neurol Med* **2016**, 6108234 (2016).
91. McCrea, P.C. & Jagoe, W.S. Myocarditis in Myasthenia Gravis with Thymoma. *Ir J Med Sci* **454**, 453-457 (1963).
92. Meegada, S., Akbar, H., Siddamreddy, S., Casement, D. & Verma, R. Granulomatous Myositis Associated with Extremely Elevated Anti-striated Muscle Antibodies in the Absence of Myasthenia Gravis. *Cureus* **12**, e6981 (2020).
93. Miller, H., Shenstone, B.D., Joffe, R. & Kannangara, S. A case of invasive thymoma associated with myasthenia gravis, myositis and demyelinating neuropathy. *Clin Exp Neurol* **22**, 13-18 (1986).
94. Mitsumune, S., *et al.* Autologous Bone Marrow Transplantation for Polymyositis Combined with Myasthenia Gravis and Aplastic Anemia: A Case Report. *Case Rep Neurol* **10**, 108-111 (2018).
95. Morrison, I., McEntegart, A. & Capell, H. Polymyositis with cardiac manifestations and unexpected immunology. *Ann Rheum Dis* **61**, 1110-1111 (2002).
96. Mrowiec, W. & Terminska-Mrowiec, K. [Myasthenic reaction in polymyositis]. *Przegl Lek* **47**, 420-421 (1990).
97. Nakano, T., Shindo, M., Oguchi, K., Yanagisawa, N. & Tsukagoshi, H. [Chronic polymyositis with electrophysiological characteristics of myasthenia gravis and myasthenic syndrome. A case report (author's transl)]. *Rinsho Shinkeigaku* **20**, 100-106 (1980).
98. Namba, T., Brunner, N.G. & Grob, D. Association of myasthenia gravis with pemphigus vulgaris, Candida albicans infection, polymyositis and myocarditis. *J Neurol Sci* **20**, 231-242 (1973).
99. Namba, T., Brunner, N.G. & Grob, D. Idiopathic giant cell polymyositis. Report of a case and review of the syndrome. *Arch Neurol* **31**, 27-30 (1974).
100. Otton, S.H., Standen, G.R. & Ormerod, I.E. T cell lymphocytosis associated with polymyositis, myasthenia gravis and thymoma. *Clin Lab Haematol* **22**, 307-308 (2000).
101. Paik, J.J., Corse, A.M. & Mammen, A.L. The co-existence of myasthenia gravis in patients with myositis: a case series. *Semin Arthritis Rheum* **43**, 792-796 (2014).

102. Pascuzzi, R.M., Roos, K.L. & Phillips, L.H., 2nd. Granulomatous inflammatory myopathy associated with myasthenia gravis. A case report and review of the literature. *Arch Neurol* **43**, 621-623 (1986).
103. Pestronk, A., Kos, K., Lopate, G. & Al-Lozi, M.T. Brachio-cervical inflammatory myopathies: clinical, immune, and myopathologic features. *Arthritis Rheum* **54**, 1687-1696 (2006).
104. Priemer, D.S., Davidson, D.D., Loehrer, P.J. & Badve, S.S. Giant Cell Polymyositis and Myocarditis in a Patient With Thymoma and Myasthenia Gravis: A Postviral Autoimmune Process? *J Neuropathol Exp Neurol* **77**, 661-664 (2018).
105. Qiao, L., *et al.* Muscular pathological features in patients with myasthenia gravis. *Clin Neuropathol* **40**, 319-327 (2021).
106. Rajan, A., Kotlyar, D. & Giaccone, G. Acute autoimmune hepatitis, myositis, and myasthenic crisis in a patient with thymoma. *J Thorac Oncol* **8**, e87-88 (2013).
107. Raschilas, F., *et al.* Concomitant polymyositis and myasthenia gravis reveal malignant thymoma. A case report and review of the literature. *Ann Med Interne (Paris)* **150**, 370-373 (1999).
108. Reznik, M. [Two cases of myasthenic syndrome with thymoma, polymyositis, myocarditis, and thyroiditis]. *J Neurol Sci* **22**, 341-351 (1974).
109. Rodolico, C., Messina, S., Toscano, A., Vita, G. & Gaeta, M. Axial myopathy in myasthenia: a misleading cause of dropped head. *Muscle Nerve* **29**, 329-330 (2004).
110. Rodriguez Prida, J., Trapiella Martinez, L. & Astudillo Gonzalez, A. Giant cell myositis associated with myasthenia gravis and thymoma. *Med Clin (Barc)* **151**, 169 (2018).
111. Roggenkamper, P. & Velho-Groneberg, P. [Ocular myositis/myasthenia (author's transl)]. *Klin Monbl Augenheilkd* **179**, 357-358 (1981).
112. Rowland, L.P. Prostigmine-responsiveness and the diagnosis of myasthenia gravis. *Neurology* **5**, 612-623 (1955).
113. Ruiz, J., *et al.* [Giant-cell myocarditis: a systemic disease? Apropos a case]. *Med Clin (Barc)* **101**, 459-461 (1993).
114. Saeki, S., Fukusako, T., Negoro, K., Nogaki, H. & Morimatsu, M. [Polymyositis followed by myasthenia gravis]. *Nihon Ronen Igakkai Zasshi* **33**, 532-534 (1996).
115. Sanguesa Gomez, C., *et al.* Dermatomyositis and myastenia gravis: An uncommon association with therapeutic implications. *Reumatol Clin* **11**, 244-246 (2015).
116. Santos, E., *et al.* Inflammatory myopathy associated with myasthenia gravis with and without thymic pathology: Report of four cases and literature review. *Autoimmun Rev* **16**, 644-649 (2017).
117. Sarwar, S., Oyewunmi, O., Bhola, K. & Heydari, B. Thymoma-Associated Myasthenia Gravis With Myocarditis. *Cureus* **15**, e42473 (2023).
118. Sasaki, H., Yano, M., Kawano, O., Hikosaka, Y. & Fujii, Y. Thymoma associated with fatal myocarditis and polymyositis in a 58-year-old man following treatment with carboplatin and paclitaxel: A case report. *Oncol Lett* **3**, 300-302 (2012).
119. Sato, H., *et al.* [A patient with giant cell myocarditis and myositis associated with thymoma and myasthenia gravis]. *Rinsho Shinkeigaku* **43**, 496-499 (2003).
120. Scangarello, F.A., *et al.* Giant cell myositis associated with concurrent myasthenia gravis: a case-based review of the literature. *Clin Rheumatol* **40**, 3841-3851 (2021).
121. Seton, M., Wu, C.C. & Louissaint, A., Jr. Case records of the Massachusetts General Hospital. Case 26-2013. A 46-year-old woman with muscle pain and swelling. *N Engl J Med* **369**, 764-773 (2013).
122. Shah, A., Pace, A., Hilton, D., Househam, E. & Weatherby, S. Giant cell myositis responsive to combined corticosteroids and immunoglobulin. *Pract Neurol* **15**, 456-459 (2015).

123. Shichijo, K., *et al.* Involvement of mitochondria in myasthenia gravis complicated with dermatomyositis and rheumatoid arthritis: a case report. *Acta Neuropathol* **109**, 539-542 (2005).
124. Shiihashi, G., *et al.* Enlargement of thymoma triggers overlapping autoimmune diseases. *Autoimmun Rev* **15**, 1200-1201 (2016).
125. Shimada, K., Koh, C.S., Tsukada, N., Shoji, S. & Yanagisawa, N. [Detection of immune complexes in the sera and around the muscle fibers in a case of myasthenia gravis and polymyositis]. *Rinsho Shinkeigaku* **29**, 432-435 (1989).
126. Soichot, P., Audry-Chaboud, D., Martin, F. & Bady, B. [Paraneoplastic neuromuscular manifestations--apropos of a case presenting successively a picture of myotonia, then myasthenia with myositis 4 years before the diagnosis of a colonic neoplasm]. *Rev Electroencephalogr Neurophysiol Clin* **12**, 147-152 (1982).
127. Souvannanorath, S., *et al.* NEUROMUSCULAR JUNCTION RELATED DISORDERS: EP.295 Brachio-cervical inflammatory myopathy associated with myasthenia gravis. *Neuromuscul Disord* **31**, S140 (2021).
128. Stefanou, M.I., *et al.* A case of late-onset, thymoma-associated myasthenia gravis with ryanodine receptor and titin antibodies and concomitant granulomatous myositis. *BMC Neurol* **16**, 172 (2016).
129. Suzuki, H., *et al.* Rare type myositis and myocarditis revealed in myasthenia gravis with malignant thymoma. *Saishin-igaku* **31**, 2417-2424 (1976).
130. Suzuki, S., *et al.* Autoimmune targets of heart and skeletal muscles in myasthenia gravis. *Arch Neurol* **66**, 1334-1338 (2009).
131. Szobor, A., Simon, K. & Szigeti, A. [Fulminating fatal dermatomyositis and myasthenia gravis after a bee sting (clinicopathological study)]. *Morphol Igazsagugyi Orv Sz* **25**, 207-212 (1985).
132. Tanahashi, N., *et al.* A case report of giant cell myocarditis and myositis observed during the clinical course of invasive thymoma associated with myasthenia gravis. *Keio J Med* **53**, 30-42 (2004).
133. Terayama, A., *et al.* A case of thymoma-associated myasthenia gravis accompanied with myositis showing the clusters of histiocyte along the fascicles in perimysium. *Clin Neurol Neurosurg* **229**, 107715 (2023).
134. Todorov, S.S., Deribas, V.J., Kazmin, A.S. & Todorov, S.S., Jr. [Morphological and immunogistochemical characteristics of myocardial damage in miastenia]. *Kardiologiia* **62**, 47-51 (2022).
135. Tomimoto, H., Akiguchi, I., Kameyama, M., Haibara, H. & Kitaichi, M. [Giant cell myositis and myocarditis associated with myasthenia gravis and thymoma--an autopsy case]. *Rinsho Shinkeigaku* **25**, 688-693 (1985).
136. Tsao, C.Y., Mendell, J.R., Lo, W.D., Luquette, M. & Rennebohm, R. Myasthenia gravis and associated autoimmune diseases in children. *J Child Neurol* **15**, 767-769 (2000).
137. Tse, S., *et al.* Myasthenia gravis and polymyositis as manifestations of chronic graft-versus-host-disease. *Bone Marrow Transplant* **23**, 397-399 (1999).
138. Turinese, A. & Pagnes, P. [Dermatomyositis and the myasthenic syndrome]. *Minerva Dermatol* **38**, 58-64 (1963).
139. Uchio, N., *et al.* Inflammatory myopathy with myasthenia gravis: Thymoma association and polymyositis pathology. *Neurol Neuroimmunol Neuroinflamm* **6**, e535 (2019).
140. van Boekel, V., Godoy, J.M. & Menezes, J.M. [An unusual association: polymyositis, myasthenia gravis and thymoma]. *Arq Neuropsiquiatr* **48**, 505-514 (1990).
141. van de Warrenburg, B.P., *et al.* Concomitant dermatomyositis and myasthenia gravis presenting with respiratory insufficiency. *Muscle Nerve* **25**, 293-296 (2002).

142. Vasilescu, C., Bucur, G., Petrovici, A. & Florescu, A. Myasthenia in patients with dermatomyositis: clinical, electrophysiological and ultrastructural studies. *J Neurol Sci* **38**, 129-144 (1978).
143. Venna, N., Gonzalez, R.G. & Zukerberg, L.R. Case records of the Massachusetts General Hospital. Case 39-2011. A woman in her 90s with unilateral ptosis. *N Engl J Med* **365**, 2413-2422 (2011).
144. Vernino, S., Auger, R.G., Emslie-Smith, A.M., Harper, C.M. & Lennon, V.A. Myasthenia, thymoma, presynaptic antibodies, and a continuum of neuromuscular hyperexcitability. *Neurology* **53**, 1233-1239 (1999).
145. Wang, Y., Zhao, N., Yang, J. & Wen, Y. Case Report: Orbital Myositis and Myasthenia Gravis as Symptoms of Immune Reconstitution Inflammatory Syndrome in a Patient With Human Immunodeficiency Virus Infection. *Front Immunol* **11**, 595068 (2020).
146. Warter, J.M., Stoebner, P., Morand, G., Bernhardt, C. & Isch, F. [Polymyositis, myasthenia and malignant thymoma. Apropos of a case]. *Rev Otoneuroophtalmol* **42**, 263-268 (1970).
147. Weiller, P.J., *et al.* [Association of polymyositis, myasthenia, and thymoma. A case and review of the literature]. *Ann Med Interne (Paris)* **135**, 299-304 (1984).
148. Yagi, N., Watanabe, T., Ikeda, Y. & Fukushima, N. Successful bridge to recovery in a patient with fulminant giant cell myocarditis that developed from multiple autoimmune disorders including myasthenia gravis: a case report. *Eur Heart J Case Rep* **6**, ytac046 (2022).
149. Yamaguchi, Y., Sakurai, Y., Mannen, T. & Shimizu, J. Rapidly progressive polymyositis with elevated antiacetylcholine receptor antibody activity. *Intern Med* **39**, 1108-1110 (2000).
150. Yamamoto, T., *et al.* Polymyositis and myocarditis after chemotherapy for advanced thymoma. *Cancer Treat Commun* **1**, 9-10 (2013).
151. Yanagihara, C., Nakaji, K., Tanaka, Y., Yabe, H. & Nishimura, Y. [A patient of chronic graft-versus-host disease presenting simultaneously with polymyositis and myasthenia gravis]. *Rinsho Shinkeigaku* **41**, 503-506 (2001).
152. Yildirim, F., Mutlu, M.Y., Icacan, O.C. & Bes, C. Dermatomyositis associated with thymoma: A case report and literature review. *Clin Ter* **174**, 115-120 (2023).
153. Yoshidome, Y., *et al.* A case of polymyositis complicated with myasthenic crisis. *Clin Rheumatol* **26**, 1569-1570 (2007).
154. Zamecnik, J., *et al.* [Granulomatous myopathy in patients with sarcoidosis and myasthenia gravis]. *Cesk Patol* **42**, 175-181 (2006).
155. Zhou, Z., Chen, X., Liu, G., Pu, J. & Wu, J. Presence of Multiple Autoimmune Antibodies Involved in Concurrent Myositis and Myocarditis and Myasthenia Gravis Without Thymoma: A Case Report. *Front Neurol* **10**, 770 (2019).

***B. StrAbs in Classic Myositis***

1. Strauss, A.J., van der Geld, H.W., Kemp, P.G., Jr., Exum, E.D. & Goodman, H.C. Immunological concomitants of myasthenia gravis. *Ann N Y Acad Sci* **124**, 744-766 (1965).
2. Wada, K., *et al.* Radioimmunoassay for antibodies to human skeletal muscle myosin in serum from patients with polymyositis. *Clin Exp Immunol* **52**, 297-304 (1983).
3. Cikes, N., *et al.* Striational autoantibodies: quantitative detection by enzyme immunoassay in myasthenia gravis, thymoma, and recipients of D-penicillamine or allogeneic bone marrow. *Mayo Clin Proc* **63**, 474-481 (1988).
4. Mygland, A., Aarli, J.A., Hofstad, H. & Gilhus, N.E. Heart muscle antibodies in myasthenia gravis. *Autoimmunity* **10**, 263-267 (1991).
5. Mygland, A., *et al.* Ryanodine receptor autoantibodies in myasthenia gravis patients with a thymoma. *Ann Neurol* **32**, 589-591 (1992).
6. Gautel, M., *et al.* Titin antibodies in myasthenia gravis: identification of a major immunogenic region of titin. *Neurology* **43**, 1581-1585 (1993).
7. Somnier, F.E., Skeie, G.O., Aarli, J.A. & Trojaborg, W. EMG evidence of myopathy and the occurrence of titin autoantibodies in patients with myasthenia gravis. *Eur J Neurol* **6**, 555-563 (1999).
8. Suzuki, S., *et al.* Novel autoantibodies to a voltage-gated potassium channel Kv1.4 in a severe form of myasthenia gravis. *J Neuroimmunol* **170**, 141-149 (2005).
9. Stergiou, C., *et al.* Titin antibodies in "seronegative" myasthenia gravis--A new role for an old antigen. *J Neuroimmunol* **292**, 108-115 (2016).
10. Kufukihara, K., *et al.* Cytometric cell-based assays for anti-striational antibodies in myasthenia gravis with myositis and/or myocarditis. *Sci Rep* **9**, 5284 (2019).

***C. Isolated IIM with TET***

1. Akimoto, N., *et al.* [A case of anti-155/140 antibodies-positive dermatomyositis associated with thymic carcinoma]. *Jpn J Clin Dermatol* **67**, 322-326 (2013).
2. Aksu, A., Sin, C. & Yilmaz, B. Adult Tiger Man: A Case of Dermatomyositis Associated With Thymoma. *Clin Nucl Med* **47**, e448-e449 (2022).
3. Azuma, Y., *et al.* Polymyositis with atypical pathological features associated with thymic carcinoma. *Intern Med* **48**, 163-168 (2009).
4. Barre, M., Delaporte, E., Berbis, P. & Benzaquen, M. Severe dermatomyositis revealing a thymic carcinoma: Did rituximab delay the diagnosis? *Dermatol Ther* **33**, e14016 (2020).
5. Bignami, A. & Calcara, S. Miocardite e miosite a cellule giganti associate a tumore del timo [Giant cell myocarditis and myositis associated with thymic tumor]. *Policlinico (Prat)* **69**, 857-866 (1962).
6. Bonduelle, M. & Bouygues, P. [Myasthenia and polymyositis: a case of polymyositis with thymoma, myasthenic syndrome in myositis; myasthenia, a syndrome or disease]. *Presse Med (1893)* **63**, 1572-1575 (1955).
7. Butany, J.W., McAuley, P., Bergeron, C. & MacLaughlin, P. Giant cell myocarditis and myositis associated with thymoma and leprosy. *Can J Cardiol* **7**, 141-145 (1991).
8. Cranney, A., Markman, S., Lach, B. & Karsh, J. Polymyositis in a patient with thymoma and T cell lymphocytosis. *J Rheumatol* **24**, 1413-1416 (1997).
9. D'Agostino, S., Avella, A. & Maddaluno, R. [Considerations on a case of the Fiedler type of acute interstitial myocarditis and of granulomatous myositis associated with a thvmic tumor]. *Rass Clin Ter* **61**, 118-126 (1962).
10. Dell'Amore, A., *et al.* Paraneoplastic dermatomyositis as presentation of thymic carcinoma. *Gen Thorac Cardiovasc Surg* **61**, 422-425 (2013).
11. Du, X., *et al.* Case Report: MDM4 Amplified in a Thymoma Patient With Autoimmune Enteropathy and Myocarditis. *Front Endocrinol (Lausanne)* **12**, 661316 (2021).
12. Fang, T.J., *et al.* Spontaneous retroperitoneal hemorrhage in a mediastinal tumor in a patient with polymyositis: a case report. *Kaohsiung J Med Sci* **24**, 436-440 (2008).
13. Fong, P.H., Wee, A., Chan, H.L. & Tan, Y.O. Primary thymic carcinoma and its association with dermatomyositis and pure red cell aplasia. *Int J Dermatol* **31**, 426-428 (1992).
14. Frith, J., Toller-Artis, E., Tcheurekdjian, H. & Hostoffer, R. Good syndrome and polymyositis. *Ann Allergy Asthma Immunol* **112**, 478 (2014).
15. Fu, Z., Chen, G., Chen, X. & Li, Q. 18F-FDG PET/CT in a Patient With Thymoma-Associated Paraneoplastic Polymyositis. *Clin Nucl Med* **45**, 148-150 (2020).
16. Funkhouser, J.W. Thymoma associated with myocarditis and the L.E.-cell phenomenon. Report of a case. *N Engl J Med* **264**, 34-36 (1961).
17. Giordano, A.S. & Haymond, J.L. Myasthenia gravis: a report of two cases with necropsy findings. *Am J Clin Pathol* **14**, 253-265 (1944).
18. Glennon, P.E., Petersen, M.E. & Sheppard, M.N. Fatal giant cell myocarditis after resection of thymoma. *Heart* **75**, 531-532 (1996).
19. Haen, S.P., *et al.* Choroidal metastases from thymic carcinoma during pregnancy: Case Report. *BMC Cancer* **15**, 972 (2015).
20. Herrmann, D.N., Blaivas, M., Wald, J.J. & Feldman, E.L. Granulomatous myositis, primary biliary cirrhosis, pancytopenia, and thymoma. *Muscle Nerve* **23**, 1133-1136 (2000).
21. Iacovelli, R., *et al.* Dermatomyositis as first clinical appearance for a thymic epidermoid cell carcinoma. *Acta Biomed* **81**, 68-71 (2010).
22. Inoue, Y., True, L.D. & Martins, R.G. Thymic carcinoma associated with paraneoplastic polymyositis. *J Clin Oncol* **27**, e33-34 (2009).

23. Isobe, K., *et al.* [An autopsied case of giant cell myocarditis and myositis associated with invasive thymoma]. *Nihon Kokyuki Gakkai Zasshi* **48**, 432-438 (2010).
24. Jin, Z.S., Tao, X.R. & Wang, Z.X. A case report of dermatomyositis mimicking myasthenia gravis. *Medicine (Baltimore)* **102**, e36234 (2023).
25. Karino, K., *et al.* Anti-TIF1gamma antibody predicted malignancy of thymic tumor with dermatomyositis as an "autoimmune tumor marker": A case report. *Medicine (Baltimore)* **97**, e13563 (2018).
26. Karippacheril, J.G., Shetty, R., Sagar, S.C. & Kamath, S.G. Myocarditis after thymoma resection, with left ventricular hypokinesia mimicking acute coronary syndrome. *J Anesth* **27**, 805-806 (2013).
27. Kilgallen, C.M., Jackson, E., Bankoff, M., Salomon, R.N. & Surks, H.K. A case of giant cell myocarditis and malignant thymoma: a postmortem diagnosis by needle biopsy. *Clin Cardiol* **21**, 48-51 (1998).
28. Koppula, B.R., Pipavath, S. & Lewis, D.H. Epstein-Barr virus (EBV) associated lymphoepithelioma-like thymic carcinoma associated with paraneoplastic syndrome of polymyositis: a rare tumor with rare association. *Clin Nucl Med* **34**, 686-688 (2009).
29. Langston, J.D., Wagman, G.F. & Dickenman, R.C. Granulomatous myocarditis and myositis associated with thymoma. *Arch Pathol* **68**, 367-373 (1959).
30. Le Marc'hadour, F., Martins Ramos, J., Pasquier, B., Pasquier, D. & Couderc, P. [Association of thymus carcinoma, Hashimoto's thyroiditis and polymyositis. Anatomoclinical case with autopsy findings]. *Ann Pathol* **9**, 355-359 (1989).
31. Maramao, F., Maramao, F.S., Monteso, L.S. & Marino, M. Biventricular Fatal Fulminant Myocarditis Infarct-Like Presentation in a Patient with Thymoma. *Arq Bras Cardiol* **121**, e20230868 (2024).
32. Mary, H., *et al.* [Nodular polymyositis and thymoma. Apropos of a case]. *Ann Chir* **40**, 583-584 (1986).
33. Meessen, H. [Clinico-pathological colloquy. Case 33]. *Dtsch Med Wochenschr* **87**, 2438-2441 (1962).
34. Miyata, R., *et al.* [A Case of Cancer-associated Dermatomyositis in a Patient with Thymic Carcinoma]. *Jpn J Lung Cancer* **56**, 189-193 (2016).
35. Munoz Malaga, A., Bautista Lorite, J., Lopez Dominguez, J.M. & Martinez Navarro, M.L. [Thymoma and muscular involvement. Differential diagnosis between polymyositis and myasthenia gravis]. *Med Clin (Barc)* **100**, 397 (1993).
36. Ohtsuki, A., Mayuzumi, N. & Ikeda, S. [A Case of Thymoma with Skin Metastasis, and Development into Dermatomyositis]. *The Nishinihon Journal of Dermatology* **70**, 43-45 (2008).
37. Pecoud, A., Essinger, A., Miklossy, J. & Ribaux, C. [Cachexia, pain and muscle weakness in a 69-year-old woman]. *Schweiz Med Wochenschr* **119**, 407-415 (1989).
38. Pentz, W.H. Advanced heart block as a manifestation of a paraneoplastic syndrome from malignant thymoma. *Chest* **116**, 1135-1136 (1999).
39. Rini, B.I. & Gajewski, T.F. Polymyositis with respiratory muscle weakness requiring mechanical ventilation in a patient with metastatic thymoma treated with octreotide. *Ann Oncol* **10**, 973-979 (1999).
40. Rowland, L.P., Lisak, R.P., Schotland, D.L., DeJesus, P.V. & Berg, P. Myasthenic myopathy and thymoma. *Neurology* **23**, 282-288 (1973).
41. Rundle, L.G. & Sparks, F.P. Thymoma and dermatomyositis. A disease entity. *Arch Pathol* **75**, 276-283 (1963).
42. Sakuma, H., Yoshida, H., Kasukawa, R., Satoh, N. & Yoshino, K. An autopsy case with Good's syndrome and dermatomyositis. *Clin Rheumatol* **4**, 196-201 (1985).

43. Schmid, K.O. [Granulomatous giant cell polymyositis and myocarditis in benign thymoma]. *Verh Dtsch Ges Pathol* **49**, 248-253 (1965).
44. Souadjian, J.V., Howell, L.P. & Lambert, E.H. Thymoma with myopathy. Report of a case. *Minn Med* **52**, 595-596 (1969).
45. Svahn, J.*, et al.* Immune-Mediated Rippling Muscle Disease Associated With Thymoma and Anti-MURC/Cavin-4 Autoantibodies. *Neurol Neuroimmunol Neuroinflamm* **10**(2023).
46. Takahashi, F.*, et al.* Successful resection of dermatomyositis associated with thymic carcinoma: report of a case. *Surg Today* **38**, 245-248 (2008).
47. Waller, J.V., Shapiro, M. & Paltauf, R. Congestive heart failure in postmenopausal muscular dystrophy: myositis, myocarditis, thymoma. *Am Heart J* **53**, 479-484 (1957).
48. Yang, X.*, et al.* Successful treatment of thymic carcinoma with dermatomyositis and interstitial pneumonia: A case report. *Thorac Cancer* **10**, 2031-2034 (2019).
49. Yildiz, C.*, et al.* Thymic carcinoma presenting with overlap polyarthritis and myositis: A rare paraneoplastic syndrome in childhood. *Int J Rheum Dis* **27**, e15187 (2024).

***D. Isolated IIM with StrAbs (without TET)***

1. Humbert, P., Faivre, R., Fellman, D., Bassand, J.P. & Dupond, J.L. Giant cell myocarditis: an autoimmune disease? *Am Heart J* **115**, 485-487 (1988).
2. Kuyama, N.*, et al*. Anti-Kv1.4 Antibody Without Myasthenia Gravis: A Rare Cause of Autoimmune Myocarditis and Myositis. *JACC Case Rep* **9**, 101734 (2023).
3. Singh, Y., Laskar, S., Mittal, M., Shirazi, N. & Gupta, S. Polymyositis Presenting with Respiratory Symptoms. *Neurol India* **69**, 1391-1393 (2021).
4. Sugiyama, A.*, et al*. Marked Respiratory Failure in an Ambulant Patient with Immune-mediated Necrotizing Myopathy and Anti-Kv1.4 and Anti-titin Antibodies. *Intern Med* **60**, 2671-2675 (2021).

***E. ICI-Induced Myositis and/or MG***

1. Abidoye, O., Kim, N. & Fombi, J. An Interesting Case Report of Myasthenia Gravis Exacerbation Induced by Durvalumab. *Cureus* **14**, e26985 (2022).
2. Abulnaja, R. Stage 4 Non-small Cell Lung Cancer With Human Epidermal Growth Factor Receptor 2 Alterations and Myocarditis Induced by Immune Checkpoint Inhibitors: A Case Report. *Cureus* **15**, e48859 (2023).
3. Adeoye, F.W., Jaffar, N., Surandran, S., Begum, G. & Islam, M.R. Durvalumab-Induced Triple-M Syndrome. *Eur J Case Rep Intern Med* **11**, 004729 (2024).
4. Aggarwal, N., Bianchini, D., Parkar, R. & Turner, J. Immunotherapy-Induced Overlap Syndrome: Myositis, Myasthenia Gravis, and Myocarditis-A Case Series. *Case Rep Med* **2024**, 5399073 (2024).
5. Aghel, N.*, et al.* Recurrent Myocarditis Induced by Immune-Checkpoint Inhibitor Treatment Is Accompanied by Persistent Inflammatory Markers Despite Immunosuppressive Treatment. *JCO Precis Oncol* **5**(2021).
6. Agrawal, N.*, et al.* Cardiac Toxicity Associated with Immune Checkpoint Inhibitors: Case Series and Review of the Literature. *Case Rep Oncol* **12**, 260-276 (2019).
7. Agrawal, Y.N., Howard Jr, J. & Collichio, F. Management of myasthenia gravis without significant exacerbation during nivolumab therapy for metastatic melanoma: a case report and review of literature. *Ann Clin Oncol* **2**(2019).
8. Ahdi, H.S., Abdulmujeeb, S. & Nabrinsky, E. Multiple Autoimmune Complications After a Single Dose of Pembrolizumab. *Cureus* **15**, e35871 (2023).
9. Ai, L.*, et al.* Nivolumab-associated DRESS in a genetic susceptible individual. *J Immunother Cancer* **9**(2021).
10. Akazawa, S.*, et al.* [A case of myopathy, myocarditis, and encephalitis with nonconvulsive status epileptics after immune checkpoint inhibitor therapy for ureter cancer]. *Rinsho Shinkeigaku* **62**, 395-398 (2022).
11. Al-Obaidi, A., Parker, N.A., Choucair, K., Alderson, J. & Deutsch, J.M. A Case of Acute Heart Failure Following Immunotherapy for Metastatic Lung Cancer. *Cureus* **12**, e8093 (2020).
12. Algaeed, M., Mukharesh, L., Heinzelmann, M. & Kaminski, H.J. Pearls & Oy-sters: Pembrolizumab-induced myasthenia gravis. *Neurology* **91**, e1365-e1367 (2018).
13. Alnahhas, I. & Wong, J. A case of new-onset antibody-positive myasthenia gravis in a patient treated with pembrolizumab for melanoma. *Muscle Nerve* **55**, E25-E26 (2017).
14. Alrayyashi, M., Uddin, M., Bdiwi, M. & Afonso, L. Immune checkpoint inhibitor-induced myopericarditis. *BMJ Case Rep* **17**(2024).
15. Ansari-Gilani, K.*, et al.* Myocarditis associated with immune checkpoint inhibitor therapy: a case report of three patients. *Emerg Radiol* **27**, 455-460 (2020).
16. Arangalage, D.*, et al.* Survival After Fulminant Myocarditis Induced by Immune-Checkpoint Inhibitors. *Ann Intern Med* **167**, 683-684 (2017).
17. Archibald, W.J.*, et al.* Brief Communication: Immune Checkpoint Inhibitor-induced Diaphragmatic Dysfunction: A Case Series. *J Immunother* **43**, 104-106 (2020).
18. Arman, C., Ibrahim, K., Elif, O.K., Hacer, D. & Yesim, P. Pembrolizumab-induced peripheral nervous system damage: A combination of myositis/ myasthenia overlap syndrome and motor axonal polyneuropathy. *Ideggyogy Sz* **76**, 422-426 (2023).
19. Arponen, O. & Skytta, T. Immune checkpoint inhibitor-induced myocarditis not visible with cardiac magnetic resonance imaging but detected with PET-CT: a case report. *Acta Oncol* **59**, 490-492 (2020).

20. Asano, R.*, et al.* Anti-TIF1gamma antibody-positive dermatomyositis in a 15-year-old boy with Epstein-Barr virus-related nasopharyngeal cancer receiving nivolumab. *Rheumatology (Oxford)* **60**, e197-e199 (2021).
21. Badovinac, S.*, et al.* Nivolumab-induced synchronous occurrence of myositis and hypothyroidism in a patient with squamous cell lung cancer. *Immunotherapy* **10**, 427-431 (2018).
22. Bae, S.*, et al.* Durvalumab-Associated Myocarditis Initially Presenting With Sinus Bradycardia Progressing Into Complete Heart Block. *Cureus* **15**, e40171 (2023).
23. Bai, J.*, et al.* Camrelizumab-Related Myocarditis and Myositis With Myasthenia Gravis: A Case Report and Literature Review. *Front Oncol* **11**, 778185 (2021).
24. Bai, J.S.*, et al.* Initial Elevated Myocardial Enzymes were Neglected in Lung Adenocarcinoma ICIS Associated Myocarditis: a Case Report. *Clin Lab* **68**(2022).
25. Baigi, T., Brown, E.N., De La Torre, R.M. & Abu-Shahin, F.I. Atezolizumab-associated myositis in a patient with unresectable hepatocellular carcinoma. *J Oncol Pharm Pract* **29**, 1757-1761 (2023).
26. Balanescu, D.V.*, et al.* Immunomodulatory treatment of immune checkpoint inhibitor-induced myocarditis: Pathway toward precision-based therapy. *Cardiovasc Pathol* **47**, 107211 (2020).
27. Baldessari, C.*, et al.* Myocarditis and diaphragmatic rhabdomyolysis with respiratory failure in a patient with metastatic melanoma treated with Nivolumab. *J Oncol Pharm Pract* **28**, 750-753 (2022).
28. Baldetti, L., Melillo, F., Beneduce, A. & Camici, P.G. Combined checkpoint inhibitor-associated myocarditis and pulmonary vasculitis mimicking acute pulmonary embolism. *Eur Heart J Cardiovasc Imaging* **20**, 243 (2019).
29. Barham, W.*, et al.* Case Report: Simultaneous Hyperprogression and Fulminant Myocarditis in a Patient With Advanced Melanoma Following Treatment With Immune Checkpoint Inhibitor Therapy. *Front Immunol* **11**, 561083 (2020).
30. Barry, T.*, et al.* Successful Treatment of Steroid-Refractory Checkpoint Inhibitor Myocarditis with Globulin Derived-Therapy: A Case Report and Literature Review. *Am J Med Sci* **362**, 424-432 (2021).
31. Bawek, S.J., Ton, R., McGovern-Poore, M., Khoncarly, B. & Narvel, R. Nivolumab-Induced Myasthenia Gravis Concomitant With Myocarditis, Myositis, and Hepatitis. *Cureus* **13**, e18040 (2021).
32. Becquart, O.*, et al.* Myasthenia Gravis Induced by Immune Checkpoint Inhibitors. *J Immunother* **42**, 309-312 (2019).
33. Behling, J., Kaes, J., Munzel, T., Grabbe, S. & Loquai, C. New-onset third-degree atrioventricular block because of autoimmune-induced myositis under treatment with anti-programmed cell death-1 (nivolumab) for metastatic melanoma. *Melanoma Res* **27**, 155-158 (2017).
34. Berg, D.D.*, et al.* Immune-related fulminant myocarditis in a patient receiving ipilimumab therapy for relapsed chronic myelomonocytic leukaemia. *Eur J Heart Fail* **19**, 682-685 (2017).
35. Berger, M.*, et al.* Pembrolizumab-induced dermatomyositis in a patient with metastatic melanoma. *Eur J Cancer* **104**, 227-230 (2018).
36. Berner, A.M., Sharma, A., Agarwal, S., Al-Sam, S. & Nathan, P. Fatal autoimmune myocarditis with anti-PD-L1 and tyrosine kinase inhibitor therapy for renal cell cancer. *Eur J Cancer* **101**, 287-290 (2018).
37. Bharathidasan, K.*, et al.* Nivolumab-induced fatal myocarditis: A case report. *Clin Case Rep* **11**, e7306 (2023).

38. Bi, H., Ren, D., Wang, Q., Ding, X. & Wang, H. Immune checkpoint inhibitor-induced myocarditis in lung cancer patients: a case report of sintilimab-induced myocarditis and a review of the literature. *Ann Palliat Med* **10**, 793-802 (2021).
39. Bilen, M.A.*, et al.* Acute rhabdomyolysis with severe polymyositis following ipilimumab-nivolumab treatment in a cancer patient with elevated anti-striated muscle antibody. *J Immunother Cancer* **4**, 36 (2016).
40. Botta, C.*, et al.* Myositis/Myasthenia after Pembrolizumab in a Bladder Cancer Patient with an Autoimmunity-Associated HLA: Immune-Biological Evaluation and Case Report. *Int J Mol Sci* **22**(2021).
41. Bourgeois-Vionnet, J.*, et al.* Nivolumab-induced myositis: A case report and a literature review. *J Neurol Sci* **387**, 51-53 (2018).
42. Brazel, D., Lee, S., Mahadevan, A., Warnecke, B. & Parajuli, R. Multiorgan Failure From Nivolumab and Ipilimumab: A Case Report and Literature Review. *Cureus* **15**, e41781 (2023).
43. Bukamur, H.S., Mezughi, H., Karem, E., Shahoub, I. & Shweihat, Y. Nivolumab-induced Third Degree Atrioventricular Block in a Patient with Stage IV Squamous Cell Lung Carcinoma. *Cureus* **11**, e4869 (2019).
44. Canino, F.*, et al.* Cemiplimab- and nivolumab-induced myasthenia gravis: two clinical cases. *Tumori* **107**, NP123-NP126 (2021).
45. Cao, J.*, et al.* Pembrolizumab-induced autoimmune Stevens-Johnson syndrome/toxic epidermal necrolysis with myositis and myocarditis in a patient with esophagogastric junction carcinoma: a case report. *Transl Cancer Res* **10**, 3870-3876 (2021).
46. Cardoso, I.*, et al.* Immune Checkpoint Inhibitor-Associated Myocarditis: A Rare Presentation With Atrioventricular Block and Sinus Node Dysfunction. *CJC Open* **5**, 829-832 (2023).
47. Carrera, W., Baartman, B.J. & Kosmorsky, G. A Case Report of Drug-Induced Myopathy Involving Extraocular Muscles after Combination Therapy with Tremelimumab and Durvalumab for Non-Small Cell Lung Cancer. *Neuroophthalmology* **41**, 140-143 (2017).
48. Chahine, J., Collier, P., Maroo, A., Tang, W.H.W. & Klein, A.L. Myocardial and Pericardial Toxicity Associated With Immune Checkpoint Inhibitors in Cancer Patients. *JACC Case Rep* **2**, 191-199 (2020).
49. Chaloulos-Iakovidis, P., Aicher, M.L. & Chilver-Stainer, L. [An Unusual Cause for a Bilateral Ptosis]. *Praxis (Bern 1994)* **110**, 643-646 (2021).
50. Cham, J., Ng, D. & Nicholson, L. Durvalumab-induced myocarditis, myositis, and myasthenia gravis: a case report. *J Med Case Rep* **15**, 278 (2021).
51. Chan, M.M., Kefford, R.F., Carlino, M., Clements, A. & Manolios, N. Arthritis and tenosynovitis associated with the anti-PD1 antibody pembrolizumab in metastatic melanoma. *J Immunother* **38**, 37-39 (2015).
52. Chang, A.*, et al.* Myocarditis With Radiotherapy and Immunotherapy in Multiple Myeloma. *J Oncol Pract* **14**, 561-564 (2018).
53. Chang, E., Sabichi, A.L. & Sada, Y.H. Myasthenia Gravis After Nivolumab Therapy for Squamous Cell Carcinoma of the Bladder. *J Immunother* **40**, 114-116 (2017).
54. Charles, J.*, et al.* Multi-organ failure induced by Nivolumab in the context of allo-stem cell transplantation. *Exp Hematol Oncol* **8**, 8 (2019).
55. Chatzantonis, G.*, et al.* Immune Checkpoint Inhibitor-Associated Myocarditis: A Run of Bad Luck or Rather Deficient-Monitoring Protocol? *JACC Case Rep* **2**, 630-635 (2020).
56. Chauhan, A., Burkeen, G., Houranieh, J., Arnold, S. & Anthony, L. Immune checkpoint-associated cardiotoxicity: case report with systematic review of literature. *Ann Oncol* **28**, 2034-2038 (2017).

57. Chauveheid, F., *et al.* [Life-threatening flare of an underlying <<paraneoplastic>> dermatomyositis in a patient with lung adenocarcinoma treated with anti-PD-1 pembrolizuma]. *Rev Med Liege* **77**, 462-467 (2022).
58. Chen, J.H., Lee, K.Y., Hu, C.J. & Chung, C.C. Coexisting myasthenia gravis, myositis, and polyneuropathy induced by ipilimumab and nivolumab in a patient with non-small-cell lung cancer: A case report and literature review. *Medicine (Baltimore)* **96**, e9262 (2017).
59. Chen, L., Zhang, S., Gong, L. & Zhang, Y. Case report: Regression after low-dose glucocorticoid therapy in a case of acute immune myocarditis induced by anti-PD-1 therapy for NSCLC. *Front Oncol* **14**, 1404045 (2024).
60. Chen, Q., *et al.* Fatal myocarditis and rhabdomyolysis induced by nivolumab during the treatment of type B3 thymoma. *Clin Toxicol (Phila)* **56**, 667-671 (2018).
61. Chen, Y., Chen, Y., Xie, J., Liu, D. & Hong, X. Multisystem immune-related adverse events due to toripalimab: Two cases-based review. *Front Cardiovasc Med* **9**, 1036603 (2022).
62. Chen, Y., Huang, A., Yang, Q., Yu, J. & Li, G. Case report: A successful re-challenge report of GLS-010 (Zimberelimab), a novel fully humanized mAb to PD-1, in a case of recurrent endometrial cancer. *Front Immunol* **13**, 987345 (2022).
63. Chen, Y., *et al.* Myocarditis related to immune checkpoint inhibitors treatment: two case reports and literature review. *Ann Palliat Med* **10**, 8512-8517 (2021).
64. Chen, Y.H., Liu, F.C., Hsu, C.H. & Chian, C.F. Nivolumab-induced myasthenia gravis in a patient with squamous cell lung carcinoma: Case report. *Medicine (Baltimore)* **96**, e7350 (2017).
65. Claus, J., Van Den Bergh, A., Verbeek, S., Wauters, E. & Nackaerts, K. Pembrolizumab-induced necrotizing myositis in a patient with metastatic non-small-cell lung cancer: a case report. *Lung Cancer Manag* **8**, LMT10 (2019).
66. Cohen, M., Mustafa, S., Elkherpitawy, I. & Meleka, M. A Fatal Case of Pembrolizumab-Induced Myocarditis in Non-Small Cell Lung Cancer. *JACC Case Rep* **2**, 426-430 (2020).
67. Compton, F., *et al.* Immune checkpoint inhibitor toxicity: A new indication for therapeutic plasma exchange? *J Clin Apher* **36**, 645-648 (2021).
68. Cooper, D.S., Meriggioli, M.N., Bonomi, P.D. & Malik, R. Severe Exacerbation of Myasthenia Gravis Associated with Checkpoint Inhibitor Immunotherapy. *J Neuromuscul Dis* **4**, 169-173 (2017).
69. Cortellini, A., *et al.* Immune Checkpoint Inhibitors and Myasthenic Syndromes: A Case Report of a Metastatic Renal Cell Carcinoma Patient Treated With Nivolumab. *J Clin Neuromuscul Dis* **20**, 99-100 (2018).
70. Coustal, C., Du Thanh, A., Roubille, F., Assenat, E. & Maria, A.T.J. Rare cutaneous toxicity of immune checkpoint inhibitors: A case of durvalumab-induced dermatomyositis. *Eur J Cancer* **155**, 25-27 (2021).
71. Crusz, S.M., *et al.* Rituximab in the treatment of pembrolizumab-induced myasthenia gravis. *Eur J Cancer* **102**, 49-51 (2018).
72. Cuenca, J.A., *et al.* Management of respiratory failure in immune checkpoint inhibitors-induced overlap syndrome: a case series and review of the literature. *BMC Anesthesiol* **23**, 310 (2023).
73. Dalal, F., Dalal, H. & Baltz, B. Pembrolizumab-Induced Myocarditis and Delayed Acute Inflammatory Demyelinating Polyradiculoneuropathy. *Cureus* **14**, e27112 (2022).
74. Dang, T., Macwan, S. & Dasanu, C.A. Late-onset double-seronegative myasthenia gravis syndrome and myasthenic crisis due to nivolumab use for Hodgkin's lymphoma. *J Oncol Pharm Pract* **27**, 1534-1538 (2021).

75. Davis, B.M., Fordjour, I., Chahin, M. & Guha, A. Immune checkpoint inhibitor-associated myocarditis and fulminant type I diabetes in a patient with metastatic non-small cell lung cancer. *BMJ Case Rep* **16**(2023).
76. de Chabot, G., *et al.* [Unexpected adverse events of immunotherapies in non-small cell lung cancer: About 2 cases]. *Rev Pneumol Clin* **73**, 326-330 (2017).
77. Deharo, F., *et al.* Immune Checkpoint Inhibitor-Induced Myositis/Myocarditis with Myasthenia Gravis-like Misleading Presentation: A Case Series in Intensive Care Unit. *J Clin Med* **11**(2022).
78. Delgado-Lazo, V., Abdelmottaleb, W. & Popescu-Martinez, A. Pembrolizumab-Induced Myocarditis and Pancreatitis in a Patient With Colon Cancer: A Case Report. *Cureus* **14**, e26034 (2022).
79. Delombaerde, D., *et al.* Ipilimumab- and nivolumab-induced myocarditis in a patient with metastatic cholangiocarcinoma: a case report. *J Med Case Rep* **16**, 275 (2022).
80. Delyon, J., *et al.* Immune checkpoint inhibitor rechallenge in patients with immune-related myositis. *Ann Rheum Dis* **78**, e129 (2019).
81. Deng, C., *et al.* Immune-Related Multiple-Organs Injuries Following ICI Treatment With Tislelizumab in an Advanced Non-Small Cell Lung Cancer Patient: A Case Report. *Front Oncol* **11**, 664809 (2021).
82. Derle, E. & Benli, S. Ipilimumab treatment associated with myasthenic crises and unfavorable disease course. *Neurol Sci* **39**, 1773-1774 (2018).
83. Dhenin, A., Samartzi, V., Lejeune, S. & Seront, E. Cascade of immunologic adverse events related to pembrolizumab treatment. *BMJ Case Rep* **12**(2019).
84. Diamantopoulos, P.T., Tsatsou, K., Benopoulou, O., Anastasopoulou, A. & Gogas, H. Inflammatory Myopathy and Axonal Neuropathy in a Patient With Melanoma Following Pembrolizumab Treatment. *J Immunother* **40**, 221-223 (2017).
85. Diamantopoulos, P.T., *et al.* Concomitant development of neurologic and cardiac immune-related adverse effects in patients treated with immune checkpoint inhibitors for melanoma. *Melanoma Res* **30**, 484-491 (2020).
86. Diaz-Rodriguez, P.E., *et al.* An Uncommon Case of Myocarditis Secondary to Durvalumab Plus Tremelimumab. *Cureus* **15**, e43628 (2023).
87. Doms, J., Prior, J.O., Peters, S. & Obeid, M. Tocilizumab for refractory severe immune checkpoint inhibitor-associated myocarditis. *Ann Oncol* **31**, 1273-1275 (2020).
88. Duarte, T., *et al.* A case of lymphocytic myocarditis in a patient treated with an immune checkpoint inhibitor, a recent class of chemotherapy agents. *Rev Port Cardiol* **41**, 1047-1051 (2022).
89. Dulgar, O., Saha, A., Elleson, K.M. & Markowitz, J. Successful treatment with carboplatin and paclitaxel in melanoma progression after immune-related adverse events. *Immunotherapy* **15**, 993-999 (2023).
90. Duminuco, A., *et al.* Previous therapy with immune checkpoint inhibitor as a cause of hypothyroidism, myositis, and renal insufficiency in a candidate for allogeneic hematopoietic transplantation. *Transpl Immunol* **75**, 101705 (2022).
91. Dumortier, J., Simon, M. & Bouhour, F. Fatal myositis and myasthenia induced by atezolizumab for the treatment of hepatocellular carcinoma. *Clin Res Hepatol Gastroenterol* **46**, 101854 (2022).
92. Earl, D.E., Loochtan, A.I. & Bedlack, R.S. Refractory myasthenia gravis exacerbation triggered By pembrolizumab. *Muscle Nerve* **57**, E120-E121 (2018).
93. Edahiro, R., *et al.* Severe myocarditis with slight lymphocytic infiltration after nivolumab treatment. *Lung Cancer* **140**, 116-117 (2020).

94. Ederhy, S.*, et al.* Immune Checkpoint Inhibitor Myocarditis With Normal Cardiac Magnetic Resonance Imaging: Importance of Cardiac Biopsy and Early Diagnosis. *Can J Cardiol* **37**, 1654-1656 (2021).
95. Eglenen Polat, B., Safi, D., Hafez, M. & Kamran, A. Pembrolizumab-Induced Myasthenia Gravis: A Case Report and Review of the Literature. *Cureus* **15**, e41087 (2023).
96. Elder, C.T., Davis, E.C., Jaipal, S. & Wight, C.E. Immune-checkpoint inhibitor toxicity during a pandemic: Overcoming patient fears to provide care. A case report. *J Oncol Pharm Pract* **27**, 2035-2040 (2021).
97. Erritzoe-Jervild, M., Scheie, D. & Stenor, C. Checkpoint inhibitor induced myositis - The value of MRI STIR. *eNeurologicalSci* **30**, 100442 (2023).
98. Esfahani, K.*, et al.* Alemtuzumab for Immune-Related Myocarditis Due to PD-1 Therapy. *N Engl J Med* **380**, 2375-2376 (2019).
99. Eslinger, C.*, et al.* Rechallenge With Switching Immune Checkpoint Inhibitors Following Autoimmune Myocarditis in a Patient With Lynch Syndrome. *J Natl Compr Canc Netw* **21**, 894-899 (2023).
100. Estenaga, A.*, et al.* Immunotherapy-intensified paraneoplastic dermatomyositis. *Indian J Dermatol Venereol Leprol* **88**, 93-96 (2021).
101. Fazal, M., Prentice, D.A., Kho, L.K. & Fysh, E. Nivolumab-associated myositis myocarditis and myasthenia and anti-striated muscle antibodies. *Intern Med J* **50**, 1003-1006 (2020).
102. Fazel, M. & Jedlowski, P.M. Severe Myositis, Myocarditis, and Myasthenia Gravis with Elevated Anti-Striated Muscle Antibody following Single Dose of Ipilimumab-Nivolumab Therapy in a Patient with Metastatic Melanoma. *Case Reports Immunol* **2019**, 2539493 (2019).
103. Feng, Y.*, et al.* Immune checkpoint inhibitor myocarditis in thymic epithelial tumors: a case report and literature review. *Transl Cancer Res* **13**, 1208-1218 (2024).
104. Figueroa-Perez, N., Kashyap, R., Bal, D., Anjum Khan, S. & Pattan, V. Autoimmune Myasthenia, Primary Adrenal Insufficiency, and Progressive Hypothyroidism Due to Pembrolizumab and Axitinib Combination Regimen. *Cureus* **13**, e16933 (2021).
105. Fionda, L.*, et al.* Eculizumab for myasthenic exacerbation during treatment with immune-checkpoint inhibitors. *Neurol Sci* **45**, 1243-1247 (2024).
106. Fox, B. & Backes, F. Varying presentations of immune checkpoint inhibitor-associated myocarditis: A case report of the clinical characteristics and outcomes of three patients. *Gynecol Oncol Rep* **49**, 101271 (2023).
107. Fox, E., Dabrow, M. & Ochsner, G. A Case of Nivolumab-Induced Myositis. *Oncologist* **21**, e3 (2016).
108. Frigeri, M.*, et al.* Immune Checkpoint Inhibitor-Associated Myocarditis: A New Challenge for Cardiologists. *Can J Cardiol* **34**, 92 e91-92 e93 (2018).
109. Fuentes-Antras, J.*, et al.* Fatal Autoimmune Storm After a Single Cycle of Anti-PD-1 Therapy: A Case of Lethal Toxicity but Pathological Complete Response in Metastatic Lung Adenocarcinoma. *Hematol Oncol Stem Cell Ther* **15**, 63-67 (2022).
110. Fukasawa, Y.*, et al.* Nivolumab-Induced Myocarditis Concomitant with Myasthenia Gravis. *Case Rep Oncol* **10**, 809-812 (2017).
111. Fukazawa, R.*, et al.* [A case of myasthenia gravis developed during pembrolizumab administration, suggesting an excitation-contraction connection disorder]. *Rinsho Shinkeigaku* **60**, 37-40 (2020).
112. Fukumitsu, M.*, et al.* Myocarditis associated with immune-checkpoint inhibitors diagnosed by cardiac magnetic resonance imaging. *Int Cancer Conf J* **12**, 109-114 (2023).

113. Gallegos, C., Rottmann, D., Nguyen, V.Q. & Baldassarre, L.A. Myocarditis with checkpoint inhibitor immunotherapy: case report of late gadolinium enhancement on cardiac magnetic resonance with pathology correlate. *Eur Heart J Case Rep* **3**, yty149 (2019).
114. Ganatra, S. & Neilan, T.G. Immune Checkpoint Inhibitor-Associated Myocarditis. *Oncologist* **23**, 879-886 (2018).
115. Gandiga, P.C., Wang, A.R., Gonzalez-Rivera, T. & Sreih, A.G. Pembrolizumab-associated inflammatory myopathy. *Rheumatology (Oxford)* **57**, 397-398 (2018).
116. Gao, L.*, et al.* Immune checkpoint inhibitor-induced myocarditis with myasthenia gravis overlap syndrome: A case report and literature review. *Medicine (Baltimore)* **101**, e32240 (2022).
117. Gao, P.*, et al.* Lethal Immune Myocarditis and Myasthenia Gravis Due to Anti-PD-1 Treatment for a Bladder Cancer Patient: A Case Report and Possible Treatment Inspiration. *Int Med Case Rep J* **17**, 359-365 (2024).
118. Garcez, D., Clara, A.I., Moraes-Fontes, M.F. & Marques, J.B. A Challenging Case of Eyelid Ptosis and Diplopia Induced by Pembrolizumab. *Cureus* **14**, e28330 (2022).
119. Garibaldi, M.*, et al.* Immune checkpoint inhibitors (ICIs)-related ocular myositis. *Neuromuscul Disord* **30**, 420-423 (2020).
120. Giancaterino, S.*, et al.* Complete heart block and subsequent sudden cardiac death from immune checkpoint inhibitor-associated myocarditis. *HeartRhythm Case Rep* **6**, 761-764 (2020).
121. Giblin, G.T.*, et al.* Subclinical Myocarditis After Combination Immune Checkpoint Inhibitor Therapy. *Circ Heart Fail* **14**, e007524 (2021).
122. Gibson, R., Delaune, J., Szady, A. & Markham, M. Suspected autoimmune myocarditis and cardiac conduction abnormalities with nivolumab therapy for non-small cell lung cancer. *BMJ Case Rep* **2016**(2016).
123. Giglio, D., Berntsson, H., Fred, A. & Ny, L. Immune Checkpoint Inhibitor-Induced Polymyositis and Myasthenia Gravis with Fatal Outcome. *Case Rep Oncol* **13**, 1252-1257 (2020).
124. Giovannini, E.*, et al.* Pembrolizumab-Induced Fatal Myasthenia, Myocarditis, and Myositis in a Patient with Metastatic Melanoma: Autopsy, Histological, and Immunohistochemical Findings-A Case Report and Literature Review. *Int J Mol Sci* **24**(2023).
125. Gonzalez, N.L., Puwanant, A., Lu, A., Marks, S.M. & Zivkovic, S.A. Myasthenia triggered by immune checkpoint inhibitors: New case and literature review. *Neuromuscul Disord* **27**, 266-268 (2017).
126. Gonzalez-Ferrero, T., Vargas-Osorio, K. & Gonzalez-Juanatey, J.R. Fulminant myocarditis with myositis after treatment with immune checkpoint inhibitors. *Med Clin (Barc)* **158**, 140-141 (2022).
127. Gonzalez-Velez, M., Suero-Abreu, G., Duma, N., Gutierrez, M. & Proverbs-Singh, T. Immune-related myocarditis and conduction abnormalities secondary to combination immunotherapy treatment with ipilimumab/nivolumab in a patient with Merkel cell carcinoma. *J Am Coll Cardiol* **71**(2018).
128. Gosser, C., Al Bawaliz, A., Bahaj, W., Chesney, J. & Ranjan, S. Immune Checkpoint Inhibitor-Induced Myositis/Myasthenia Gravis Overlap. *Cureus* **15**, e49007 (2023).
129. Grewal, N.K.S., Maning, J., Gordon, L.I. & Akhter, N. Checkpoint inhibitor myocarditis with preceding immunosuppression and tolerance of sequential anthracycline therapy. *BMJ Case Rep* **17**(2024).
130. Guiney, T.E.*, et al.* Case 30-2019: A 65-Year-Old Woman with Lung Cancer and Chest Pain. *N Engl J Med* **381**, 1268-1277 (2019).

131. Gullapalli, M., Arulprakash, N., Safar, M. & Kocurek, E. Case of Immune Checkpoint Inhibitor Induced Myasthenia Gravis. *Cureus* **16**, e58651 (2024).
132. Gupta, R., *et al.* Atezolizumab Induced Myocarditis on a Background of Cardiac Amyloidosis. *Am J Ther* **26**, e795-e797 (2019).
133. Haddox, C.L., *et al.* Pembrolizumab induced bulbar myopathy and respiratory failure with necrotizing myositis of the diaphragm. *Ann Oncol* **28**, 673-675 (2017).
134. Hajihossainlou, B., Vasileva, A., Manthri, S. & Chakraborty, K. Myasthenia gravis induced or exacerbated by immune checkpoint inhibitors: a rising concern. *BMJ Case Rep* **14**(2021).
135. Hamada, S., Fuseya, Y. & Tsukino, M. Pembrolizumab-Induced Rhabdomyolysis With Myositis in a Patient With Lung Adenocarcinoma. *Arch Bronconeumol (Engl Ed)* **54**, 346-348 (2018).
136. Hardy, T., *et al.* Acute fatal myocarditis after a single dose of anti-PD-1 immunotherapy, autopsy findings: a case report. *Cardiovasc Pathol* **46**, 107202 (2020).
137. Hasegawa, Y., Kawai, S., Ota, T., Tsukuda, H. & Fukuoka, M. Myasthenia gravis induced by nivolumab in patients with non-small-cell lung cancer: a case report and literature review. *Immunotherapy* **9**, 701-707 (2017).
138. Hassan, M.A., Batta, Y. & Afzal, M.A. A Case of Immune Checkpoint Inhibitor-Induced Probable Myocarditis and Treatment Response. *Cureus* **15**, e39692 (2023).
139. Hayakawa, N., Kikuchi, E., Suzuki, S. & Oya, M. Myasthenia gravis with myositis induced by pembrolizumab therapy in a patient with metastatic urothelial carcinoma. *Int Cancer Conf J* **9**, 123-126 (2020).
140. Hayashi, H., *et al.* A Successful Case of Hepatocellular Carcinoma Treated with Atezolizumab Plus Bevacizumab with Multisystem Immune-related Adverse Events. *Intern Med* **61**, 3497-3502 (2022).
141. Heinzerling, L., *et al.* Cardiotoxicity associated with CTLA4 and PD1 blocking immunotherapy. *J Immunother Cancer* **4**, 50 (2016).
142. Heleno, C.T., Mustafa, A., Gotera, N.A. & Tesar, A. Myasthenia Gravis as an Immune-Mediated Side Effect of Checkpoint Inhibitors. *Cureus* **13**, e16316 (2021).
143. Hellman, J.B., Traynis, I. & Lin, L.K. Pembrolizumab and epacadostat induced fatal myocarditis and myositis presenting as a case of ptosis and ophthalmoplegia. *Orbit* **38**, 244-247 (2019).
144. Hibino, M., Maeda, K., Horiuchi, S., Fukuda, M. & Kondo, T. Pembrolizumab-induced myasthenia gravis with myositis in a patient with lung cancer. *Respirol Case Rep* **6**, e00355 (2018).
145. Hinogami, H., *et al.* Case of dermatomyositis during treatment with pembrolizumab for lung cancer. *J Dermatol* **46**, e430-e432 (2019).
146. Ho, A.K. & Cooksley, T. Immune Checkpoint Inhibitor-Mediated Myasthenia Gravis. *J Emerg Med* **59**, 561-562 (2020).
147. Hong, G., *et al.* Sintilimab-induced inflammatory myopathy in a patient with esophageal cancer: a case report. *Front Immunol* **14**, 1253463 (2023).
148. Hu, X., Wei, Y. & Shuai, X. Case Report: Glucocorticoid Effect Observation in a Ureteral Urothelial Cancer Patient With ICI-Associated Myocarditis and Multiple Organ Injuries. *Front Immunol* **12**, 799077 (2021).
149. Hu, Y., *et al.* A case of subclinical immune checkpoint inhibitor-associated myocarditis in non-small cell lung cancer. *BMC Pulm Med* **23**, 119 (2023).
150. Huh, S.Y., *et al.* Emergence of Myasthenia Gravis with Myositis in a Patient Treated with Pembrolizumab for Thymic Cancer. *J Clin Neurol* **14**, 115-117 (2018).

151. Hunter, G., Voll, C. & Robinson, C.A. Autoimmune inflammatory myopathy after treatment with ipilimumab. *Can J Neurol Sci* **36**, 518-520 (2009).
152. Hyun, J.W.*, et al.* Fatal Simultaneous Multi-organ Failure Following Pembrolizumab Treatment for Refractory Thymoma. *Clin Lung Cancer* **21**, e74-e77 (2020).
153. Imai, R., Ikemura, S. & Jinta, T. Anti-TIF1gamma antibody-positive dermatomyositis associated with durvalumab administration in a patient with lung and oesophageal cancers. *Respirol Case Rep* **9**, e00736 (2021).
154. Imai, R.*, et al.* Fulminant Myocarditis Caused by an Immune Checkpoint Inhibitor: A Case Report With Pathologic Findings. *J Thorac Oncol* **14**, e36-e38 (2019).
155. Inayat, F., Masab, M., Gupta, S. & Ullah, W. New drugs and new toxicities: pembrolizumab-induced myocarditis. *BMJ Case Rep* **2018**(2018).
156. Isami, A.*, et al.* [A case of anti-titin antibody positive nivolumab-related necrotizing myopathy with myasthenia gravis]. *Rinsho Shinkeigaku* **59**, 431-435 (2019).
157. Islam, M.M. Pembrolizumab-induced diffuse myositis in a patient of metastatic colon cancer: a case report. *Clinical Medicine* **23**(2023).
158. Iwasaki, S.*, et al.* A case of immune checkpoint inhibitor-associated myocarditis after initiation of atezolizumab plus bevacizumab therapy for advanced hepatocellular carcinoma. *Clin J Gastroenterol* **14**, 1233-1239 (2021).
159. Jain, V.*, et al.* Autoimmune Myocarditis Caused by Immune Checkpoint Inhibitors Treated With Antithymocyte Globulin. *J Immunother* **41**, 332-335 (2018).
160. Jain, V., Remley, W., Bunag, C., Elfasi, A. & Chuquilin, M. Rituximab in Refractory Myositis and Acute Neuropathy Secondary to Checkpoint Inhibitor Therapy. *Cureus* **14**, e25129 (2022).
161. Jang, S.Y., Lee, S.Y., Lee, H.L. & Choi, J. Early development of pembrolizumab-induced fulminant myositis and cardiotoxicity in a patient with metastatic thymoma. *Respirol Case Rep* **10**, e01025 (2022).
162. Jazieh, K., Kottschade, L. & Dimou, A. Keeping an Eye Out for Immunotherapy Toxicity: A Case of Unilateral Ptosis Caused by Ipilimumab/Nivolumab Therapy. *J Immunother Precis Oncol* **7**, 126-129 (2024).
163. Jebaraj, A.P., Etheridge, T.J., Winegar, B.A. & Marx, D.P. Ipilimumab-related orbitopathy: a case report. *Orbit* **43**, 100-104 (2024).
164. Jenkins, J.D., Camara-Lemarroy, C., Joseph, J.T. & Brust, T. Multiple Immune-Related Adverse Event Overlap in Two Patients Treated with Pembrolizumab. *Can J Neurol Sci* **49**, 133-135 (2022).
165. Jespersen, M.S., Fano, S., Stenor, C. & Moller, A.K. A case report of immune checkpoint inhibitor-related steroid-refractory myocarditis and myasthenia gravis-like myositis treated with abatacept and mycophenolate mofetil. *Eur Heart J Case Rep* **5**, ytab342 (2021).
166. Jeyakumar, N.*, et al.* The Terrible Triad of Checkpoint Inhibition: A Case Report of Myasthenia Gravis, Myocarditis, and Myositis Induced by Cemiplimab in a Patient with Metastatic Cutaneous Squamous Cell Carcinoma. *Case Reports Immunol* **2020**, 5126717 (2020).
167. Ji, H.*, et al.* Sintilimab induced ICIAM in the treatment of advanced HCC: A case report and analysis of research progress. *Front Immunol* **13**, 995121 (2022).
168. Johansen, A., Christensen, S.J., Scheie, D., Hojgaard, J.L.S. & Kondziella, D. Neuromuscular adverse events associated with anti-PD-1 monoclonal antibodies: Systematic review. *Neurology* **92**, 663-674 (2019).
169. John, S.*, et al.* Progressive hypoventilation due to mixed CD8(+) and CD4(+) lymphocytic polymyositis following tremelimumab - durvalumab treatment. *J Immunother Cancer* **5**, 54 (2017).

170. Johnson, D.B., *et al.* Fulminant Myocarditis with Combination Immune Checkpoint Blockade. *N Engl J Med* **375**, 1749-1755 (2016).
171. Johnson, D.B., *et al.* Myasthenia Gravis Induced by Ipilimumab in Patients With Metastatic Melanoma. *J Clin Oncol* **33**, e122-124 (2015).
172. Jyothi Ramachandran Nair, D.P., Zachariah, S., Scollan, D. & Shaikh, A. Myocarditis: A Rare Complication of Immune Checkpoint Inhibitor Therapy. *Cureus* **16**, e60459 (2024).
173. Kadota, H., *et al.* Immune Checkpoint Inhibitor-Induced Myositis: a Case Report and Literature Review. *Curr Rheumatol Rep* **21**, 10 (2019).
174. Kamien, A., Knuth, A. & Santhosh-Kumar, C. Reactivation of myasthenia gravis secondary to nivolumab: case report and literature review. *J Hematol Oncol Pharm* **9**, 24-29 (2019).
175. Kamo, H., *et al.* Pembrolizumab-related systemic myositis involving ocular and hindneck muscles resembling myasthenic gravis: a case report. *BMC Neurol* **19**, 184 (2019).
176. Kang, K.H., *et al.* Immune checkpoint-mediated myositis and myasthenia gravis: A case report and review of evaluation and management. *Am J Otolaryngol* **39**, 642-645 (2018).
177. Kao, J.C., *et al.* Neurological Complications Associated With Anti-Programmed Death 1 (PD-1) Antibodies. *JAMA Neurol* **74**, 1216-1222 (2017).
178. Karlsen, T.L.S., Karlsen, J., Mo, R. & Hammer, T. [A woman in her eighties with chest pain during immunotherapy for melanoma]. *Tidsskr Nor Laegeforen* **141**(2021).
179. Kartolo, A., Towheed, T. & Mates, M. A case of successful pembrolizumab rechallenge in a patient with non-small-cell lung cancer and grade 3 dermatomyositis. *Immunotherapy* **13**, 477-481 (2021).
180. Kato, S., *et al.* Acute Myocarditis by Immune Checkpoint Inhibitor Identified by Quantitative Pixel-Wise Analysis of Native T1 Mapping. *Circ Cardiovasc Imaging* **14**, e012177 (2021).
181. Katsume, Y., *et al.* Complete Atrioventricular Block Associated with Pembrolizumab-induced Acute Myocarditis: The Need for Close Cardiac Monitoring. *Intern Med* **57**, 3157-3162 (2018).
182. Katyal, N., Katsumoto, T.R., Ramachandran, K.J., Yunce, M. & Muppidi, S. Plasma Exchange in Patients With Myositis due to Immune Checkpoint Inhibitor Therapy. *J Clin Neuromuscul Dis* **25**, 89-93 (2023).
183. Kee, W., Ng, K.Y.Y., Lee, J.J.X. & Tan, D.S.W. Myasthenia Gravis and Myocarditis After Administration of Pembrolizumab in a Patient With Metastatic Non-small Cell Lung Cancer and Resected Thymoma. *Clin Lung Cancer* **23**, e293-e295 (2022).
184. Khetan, V., Blake, E.A., Ciccone, M.A. & Matsuo, K. Rhabdomyolysis following single administration of pembrolizumab: Is severe immune-reaction a marker for durable treatment response? *Gynecol Oncol Rep* **35**, 100700 (2021).
185. Khoo, A., Zhuang, Y., Boundy, K. & Frasca, J. Immune checkpoint inhibitor-related myositis associated with atezolizumab therapy. *Neurol Clin Pract* **9**, e25-e26 (2019).
186. Khoury, Z.H., *et al.* Combination Nivolumab/Ipilimumab Immunotherapy For Melanoma With Subsequent Unexpected Cardiac Arrest: A Case Report and Review of Literature. *J Immunother* **42**, 313-317 (2019).
187. Khreisat, A., Bartosek, N., Amal, T. & Dalal, B. Durvalumab-Induced Myocarditis and Dilated Cardiomyopathy in a Patient With Non-small Cell Lung Cancer: A Diagnostic Conundrum. *Cureus* **16**, e51456 (2024).
188. Kim, J.S., *et al.* Myasthenia gravis and myopathy after nivolumab treatment for non-small cell lung carcinoma: A case report. *Thorac Cancer* **10**, 2045-2049 (2019).
189. Kimura, T., *et al.* Myasthenic crisis and polymyositis induced by one dose of nivolumab. *Cancer Sci* **107**, 1055-1058 (2016).

190. Kitazaki, Y., *et al.* Anti-Kv1.4 Antibody-positive Nivolumab-induced Myasthenia Gravis and Myositis Presenting with Bilateral Ptosis and Demonstrating Different Pathophysiologies. *Intern Med* **62**, 3013-3020 (2023).
191. Kobayashi, M., *et al.* Myositis induced by durvalumab in a patient with non-small cell lung cancer: A case report. *Thorac Cancer* **11**, 3614-3617 (2020).
192. Kobayashi, T., *et al.* Relationship between clinical course of nivolumab-related myositis and immune status in a patient with Hodgkin's lymphoma after allogeneic hematopoietic stem cell transplantation. *Int J Hematol* **109**, 356-360 (2019).
193. Koh, B., Tuite, K., Khattak, A. & Dyke, J.M. Lymphocyte involvement in nivolumab-induced autoimmune myositis. *Pathology* **51**, 555-557 (2019).
194. Komatsu, M., *et al.* A rare case of nivolumab-related myasthenia gravis and myocarditis in a patient with metastatic gastric cancer. *BMC Gastroenterol* **21**, 333 (2021).
195. Kondo, H., Kirigaya, J., Matsuzawa, Y. & Hibi, K. Two Cases of Immune Checkpoint Inhibitor-Induced Myocarditis With Complete Atrioventricular Block. *Cureus* **15**, e36446 (2023).
196. Konoeda, F., Suzuki, S., Nishimoto, Y., Hoshino, H. & Takagi, M. [A case of myasthenia gravis and myositis induced by nivolumab]. *Rinsho Shinkeigaku* **57**, 373-377 (2017).
197. Konstantina, T., *et al.* Fatal adverse events in two thymoma patients treated with anti-PD-1 immune check point inhibitor and literature review. *Lung Cancer* **135**, 29-32 (2019).
198. Kosche, C., Stout, M., Sosman, J., Lukas, R.V. & Choi, J.N. Dermatomyositis in a patient undergoing nivolumab therapy for metastatic melanoma: a case report and review of the literature. *Melanoma Res* **30**, 313-316 (2020).
199. Kosick, T.I., Patel, K., Jasinski, J. & Dada, B. A Case of Pembrolizumab-Induced Myasthenia Gravis. *Cureus* **15**, e45455 (2023).
200. Kowata, S., *et al.* Association of CD8 + T cells expressing nivolumab-free PD-1 with clinical status in a patient with relapsed refractory classical Hodgkin lymphoma. *Int J Hematol* **118**, 751-757 (2023).
201. Kudo, F., *et al.* Advanced Lung Adenocarcinoma with Nivolumab-associated Dermatomyositis. *Intern Med* **57**, 2217-2221 (2018).
202. Kuniyoshi, J., *et al.* Immune Checkpoint Inhibitor-Induced Myocarditis With Concurrent Myasthenia Gravis. *Can J Cardiol* **39**, 1646-1648 (2023).
203. Kurokawa, M., *et al.* [Re-Administration of Pembrolizumab for Urothelial Carcinoma after immune-Related Myasthenia Gravis : A Case Report]. *Hinyokika Kiyo* **68**, 295-300 (2022).
204. Kushnareva, E., *et al.* Case Report: Multiple Causes of Cardiac Death After the First Infusion of Atezolizumab: Histopathological and Immunohistochemical Findings. *Front Immunol* **13**, 871542 (2022).
205. Lara, M.S., *et al.* Immune Checkpoint Inhibitor-Induced Myasthenia Gravis in a Patient with Advanced NSCLC and Remote History of Thymoma. *Clin Lung Cancer* **20**, e489-e491 (2019).
206. Lau, K.H., Kumar, A., Yang, I.H. & Nowak, R.J. Exacerbation of myasthenia gravis in a patient with melanoma treated with pembrolizumab. *Muscle Nerve* **54**, 157-161 (2016).
207. Laubli, H., *et al.* Acute heart failure due to autoimmune myocarditis under pembrolizumab treatment for metastatic melanoma. *J Immunother Cancer* **3**, 11 (2015).
208. Leaver, P.J., Jang, H.S., Vernon, S.T. & Fernando, S.L. Immune checkpoint inhibitor-mediated myasthenia gravis with focal subclinical myocarditis progressing to symptomatic cardiac disease. *BMJ Case Rep* **13**(2020).
209. Lecouflet, M., *et al.* [Orbital myositis associated with ipilimumab]. *Ann Dermatol Venereol* **140**, 448-451 (2013).
210. Lee, D.H., *et al.* Case of pembrolizumab-induced myocarditis presenting as torsades de pointes with safe re-challenge. *J Oncol Pharm Pract* **26**, 1544-1548 (2020).

211. Lewis, R.I., *et al.* Case Report: Sudden very late-onset near fatal PD1 inhibitor-associated myocarditis with out-of-hospital cardiac arrest after >2.5 years of pembrolizumab treatment. *Front Cardiovasc Med* **11**, 1328378 (2024).
212. Li, W.L., *et al.* Long-term survival after immunotherapy and targeted therapy without chemotherapy in an elderly patient with HER2-positive gastroesophageal junction cancer: A case report. *Hum Vaccin Immunother* **18**, 2121109 (2022).
213. Liang, S., *et al.* Immune Myocarditis Overlapping With Myasthenia Gravis Due to Anti-PD-1 Treatment for a Chordoma Patient: A Case Report and Literature Review. *Front Immunol* **12**, 682262 (2021).
214. Liao, B., Shroff, S., Kamiya-Matsuoka, C. & Tummala, S. Atypical neurological complications of ipilimumab therapy in patients with metastatic melanoma. *Neuro Oncol* **16**, 589-593 (2014).
215. Liao, H.L., Chen, D.D., Pan, Y. & Liu, Z.J. [A case report of immune checkpoint inhibitor-induced myocarditis]. *Zhonghua Xin Xue Guan Bing Za Zhi* **50**, 710-712 (2022).
216. Lie, G., *et al.* Nivolumab resulting in persistently elevated troponin levels despite clinical remission of myocarditis and myositis in a patient with malignant pleural mesothelioma: case report. *Transl Lung Cancer Res* **9**, 360-365 (2020).
217. Liewluck, T., Kao, J.C. & Mauermann, M.L. PD-1 Inhibitor-associated Myopathies: Emerging Immune-mediated Myopathies. *J Immunother* **41**, 208-211 (2018).
218. Lin, Y., Yuan, X. & Chen, L. Immune myocarditis related to sintilimab treatment in a patient with advanced lung adenocarcinoma: A case report. *Front Cardiovasc Med* **9**, 955527 (2022).
219. Liu, Q., Ayyappan, S., Broad, A. & Narita, A. Pembrolizumab-associated ocular myasthenia gravis. *Clin Exp Ophthalmol* **47**, 796-798 (2019).
220. Liu, S., *et al.* Severe cardiotoxicity in 2 patients with thymoma receiving immune checkpoint inhibitor therapy: A case report. *Medicine (Baltimore)* **101**, e31873 (2022).
221. Liu, W.K., Naban, N., Kaul, A., Patel, N. & Fusi, A. Life-threatening polymyositis with spontaneous hematoma induced by nivolumab in a patient with previously resected melanoma. *Melanoma Res* **31**, 85-87 (2021).
222. Liu, X., *et al.* Sintilimab-Induced Myocarditis in a Patient with Gastric Cancer: A Case Report and Literature Review. *J Cardiovasc Dev Dis* **10**(2023).
223. Liu, Y., *et al.* Fatal myositis and spontaneous haematoma induced by combined immune checkpoint inhibitor treatment in a patient with pancreatic adenocarcinoma. *BMC Cancer* **19**, 1193 (2019).
224. Liu, Z., Fan, Y., Guo, J., Bian, N. & Chen, D. Fulminant myocarditis caused by immune checkpoint inhibitor: a case report and possible treatment inspiration. *ESC Heart Fail* **9**, 2020-2026 (2022).
225. Long, H.D., *et al.* Successful management of camrelizumab-induced immune-checkpoint-inhibitors-related myocarditis. *J Oncol Pharm Pract* **30**, 597-604 (2024).
226. Loochtan, A.I., Nickolich, M.S. & Hobson-Webb, L.D. Myasthenia gravis associated with ipilimumab and nivolumab in the treatment of small cell lung cancer. *Muscle Nerve* **52**, 307-308 (2015).
227. Lopez, D., Calvo, A. & Fershko, A. Myasthenia gravis and rhabdomyolysis in a patient with advanced renal cell cancer treated with nivolumab- a case report and review of the literature. *Br J Med Health Res* **2**, 11-16 (2015).
228. Lorente-Ros, A., *et al.* Checkpoint Immunotherapy-Induced Myocarditis and Encephalitis Complicated With Complete AV Block: Not All Hope Is Lost. *JACC Case Rep* **4**, 1032-1036 (2022).

229. Lorenzo, C.J., Fitzpatrick, H., Campdesuner, V., George, J. & Lattanzio, N. Pembrolizumab-Induced Ocular Myasthenic Crisis. *Cureus* **12**, e9192 (2020).
230. Luecke, E., *et al.* Immune Checkpoint Inhibitor-induced Fatal Myositis in a Patient With Squamous Cell Carcinoma and a History of Thymoma. *Clin Lung Cancer* **21**, e246-e249 (2020).
231. Luo, Y.B., *et al.* Case Report: The Neuromusclar Triad of Immune Checkpoint Inhibitors: A Case Report of Myositis, Myocarditis, and Myasthenia Gravis Overlap Following Toripalimab Treatment. *Front Cardiovasc Med* **8**, 714460 (2021).
232. Maeda, O., *et al.* Nivolumab for the treatment of malignant melanoma in a patient with pre-existing myasthenia gravis. *Nagoya J Med Sci* **78**, 119-122 (2016).
233. Maetani, T., Hamaguchi, T., Nishimura, T., Marumo, S. & Fukui, M. Durvalumab-associated Late-onset Myocarditis Successfully Treated with Corticosteroid Therapy. *Intern Med* **61**, 527-531 (2022).
234. Mahmood, S.S., *et al.* Myocarditis with tremelimumab plus durvalumab combination therapy for endometrial cancer: A case report. *Gynecol Oncol Rep* **25**, 74-77 (2018).
235. Makarious, D., Horwood, K. & Coward, J.I.G. Myasthenia gravis: An emerging toxicity of immune checkpoint inhibitors. *Eur J Cancer* **82**, 128-136 (2017).
236. Marano, A.L., *et al.* Subacute cutaneous lupus erythematosus and dermatomyositis associated with anti-programmed cell death 1 therapy. *Br J Dermatol* **181**, 580-583 (2019).
237. March, K.L., Samarin, M.J., Sodhi, A. & Owens, R.E. Pembrolizumab-induced myasthenia gravis: A fatal case report. *J Oncol Pharm Pract* **24**, 146-149 (2018).
238. Martinez-Calle, N., *et al.* Anti-PD1 associated fulminant myocarditis after a single pembrolizumab dose: the role of occult pre-existing autoimmunity. *Haematologica* **103**, e318-e321 (2018).
239. Mathews, E.P. & Romito, J.W. Management of immune checkpoint inhibitor-related acute hypoxic neuromuscular respiratory failure using high-flow nasal cannula. *Proc (Bayl Univ Med Cent)* **33**, 407-408 (2020).
240. Matson, D.R., Accola, M.A., Rehrauer, W.M. & Corliss, R.F. Fatal Myocarditis Following Treatment with the PD-1 Inhibitor Nivolumab. *J Forensic Sci* **63**, 954-957 (2018).
241. Matsubara, S., Seki, M., Suzuki, S., Komori, T. & Takamori, M. Tertiary lymphoid organs in the inflammatory myopathy associated with PD-1 inhibitors. *J Immunother Cancer* **7**, 256 (2019).
242. Matsui, H., *et al.* A Fatal Case of Myocarditis Following Myositis Induced by Pembrolizumab Treatment for Metastatic Upper Urinary Tract Urothelial Carcinoma. *Int Heart J* **61**, 1070-1074 (2020).
243. Matsuo, K., *et al.* Nivolumab-induced Myocarditis Successfully Treated with Corticosteroid Therapy: A Case Report and Review of the Literature. *Intern Med* **58**, 2367-2372 (2019).
244. McDowall, L.M., Fernando, S.L., Ange, N., Yun, J. & Chia, K.K.M. Immune checkpoint inhibitor-mediated myocarditis and ventricular tachycardia storm. *HeartRhythm Case Rep* **5**, 497-500 (2019).
245. Mehta, A., Gupta, A., Hannallah, F., Koshy, T. & Reimold, S. Myocarditis as an immune-related adverse event with ipilimumab/nivolumab combination therapy for metastatic melanoma. *Melanoma Res* **26**, 319-320 (2016).
246. Mehta, J.J., Maloney, E., Srinivasan, S., Seitz, P. & Cannon, M. Myasthenia Gravis Induced by Nivolumab: A Case Report. *Cureus* **9**, e1702 (2017).
247. Mei, H., *et al.* Immune checkpoint inhibitor-induced myocarditis and myositis in liver cancer patients: A case report and literature review. *Front Oncol* **12**, 1088659 (2022).

248. Menachery, S.M., Hang, Y., Pritchard, L., Poklepovic, A. & Bottinor, W. Immune Checkpoint Inhibitor Rechallenge in a Patient With Previous Fulminant Myocarditis. *Am J Cardiol* **199**, 33-36 (2023).
249. Messer, A., Drozd, B., Glitza, I.C., Lu, H. & Patel, A.B. Dermatomyositis associated with nivolumab therapy for melanoma: a case report and review of the literature. *Dermatol Online J* **26**(2020).
250. Minon-Fernandez, B., Losada-Domingo, J.M., Sanchez-Horvath, M.T. & Barcena-Llona, J. [Myasthenia gravis associated with nivolumab]. *Rev Neurol* **70**, 72-73 (2020).
251. Mitsune, A., *et al.* Relapsed Myasthenia Gravis after Nivolumab Treatment. *Intern Med* **57**, 1893-1897 (2018).
252. Miyashita, K., *et al.* [Dermatomyositis with squamous cell carcinoma of the lungs secondary to nivolumab treatment: a case report]. *Rinsho Shinkeigaku* **60**, 768-772 (2020).
253. Miyauchi, Y., *et al.* Myocarditis as an immune-related adverse event following treatment with ipilimumab and nivolumab combination therapy for metastatic renal cell carcinoma: a case report. *J Med Case Rep* **15**, 508 (2021).
254. Mizumaki, K., *et al.* Case of pembrolizumab-induced dermatomyositis with anti-transcription intermediary factor 1-gamma antibody. *J Dermatol* **49**, e311-e312 (2022).
255. Mohn, N., *et al.* Acute progressive neuropathy-myositis-myasthenia-like syndrome associated with immune-checkpoint inhibitor therapy in patients with metastatic melanoma. *Melanoma Res* **29**, 435-440 (2019).
256. Monge, C., *et al.* Myocarditis in a patient treated with Nivolumab and PROSTVAC: a case report. *J Immunother Cancer* **6**, 150 (2018).
257. Montes, V., Sousa, S., Pita, F., Guerreiro, R. & Carmona, C. Myasthenia Gravis Induced by Ipilimumab in a Patient With Metastatic Melanoma. *Front Neurol* **9**, 150 (2018).
258. Naganuma, K., *et al.* An Autopsy Case of Late-onset Fulminant Myocarditis Induced by Nivolumab in Gastric Cancer. *Intern Med* **61**, 2867-2871 (2022).
259. Nakagomi, Y., *et al.* Immune Checkpoint Inhibitor-Related Myositis Overlapping With Myocarditis: An Institutional Case Series and a Systematic Review of Literature. *Front Pharmacol* **13**, 884776 (2022).
260. Nakanishi, S., Nishida, S., Miyazato, M., Goya, M. & Saito, S. A case report of nivolumab-induced myasthenia gravis and myositis in a metastatic renal cell carcinoma patient. *Urol Case Rep* **29**, 101105 (2020).
261. Nasr, F., *et al.* Severe ophthalmoplegia and myocarditis following the administration of pembrolizumab. *Eur J Cancer* **91**, 171-173 (2018).
262. Nelke, C., *et al.* Immune Checkpoint Inhibition-Related Myasthenia-Myositis-Myocarditis Responsive to Complement Blockade. *Neurol Neuroimmunol Neuroinflamm* **11**(2024).
263. Ng, A.H., Molinares, D.M., Ngo-Huang, A.T. & Bruera, E. Immunotherapy-related skeletal muscle weakness in cancer patients: a case series. *Ann Palliat Med* **10**, 2359-2365 (2021).
264. Nguyen, B.H., Kuo, J., Budiman, A., Christie, H. & Ali, S. Two cases of clinical myasthenia gravis associated with pembrolizumab use in responding melanoma patients. *Melanoma Res* **27**, 152-154 (2017).
265. Nguyen, L.S., *et al.* Reversal of immune-checkpoint inhibitor fulminant myocarditis using personalized-dose-adjusted abatacept and ruxolitinib: proof of concept. *J Immunother Cancer* **10**(2022).
266. Nierstedt, R.T., Yeahia, R. & Barnett, K.M. Unanticipated Myocarditis in a Surgical Patient Treated With Pembrolizumab: A Case Report. *A A Pract* **14**, e01177 (2020).
267. Nishikawa, T., *et al.* Sinus Node Dysfunction Co-occurring with Immune Checkpoint Inhibitor-associated Myocarditis. *Intern Med* **61**, 2161-2165 (2022).

268. Nishikawa, T., *et al.* A Case of Lung Cancer with Very-Late-Onset Immune Checkpoint Inhibitor-Related Myocarditis. *CJC Open* **4**, 651-655 (2022).
269. Nishimura, T., *et al.* Fulminant Myocarditis for Non-small-cell Carcinoma of the Lung with Nivolumab and Ipilimumab Plus Chemotherapy. *Intern Med* **62**, 1319-1322 (2023).
270. Nishiyama, K., *et al.* Case report: Electrocardiographic changes in pembrolizumab-induced fatal myocarditis. *Front Immunol* **14**, 1078838 (2023).
271. Noda, T., Kageyama, H., Miura, M., Tamura, T. & Ito, H. [A case of myasthenia gravis and myositis induced by pembrolizumab]. *Rinsho Shinkeigaku* **59**, 502-508 (2019).
272. Norwood, T.G., *et al.* Evolution of Immune Checkpoint Blockade-Induced Myocarditis Over 2 Years. *JACC Case Rep* **2**, 203-209 (2020).
273. Norwood, T.G., *et al.* Smoldering myocarditis following immune checkpoint blockade. *J Immunother Cancer* **5**, 91 (2017).
274. Nosaki, Y., *et al.* Paraspinal muscle involvement in pembrolizumab-associated myositis. *Oxf Med Case Reports* **2020**, omaa003 (2020).
275. Ogawa, T., *et al.* Polymyositis induced by PD-1 blockade in a patient in hepatitis B remission. *J Neurol Sci* **381**, 22-24 (2017).
276. Ohira, J., Kawamoto, M., Sugino, Y. & Kohara, N. A case report of fulminant cytokine release syndrome complicated by dermatomyositis after the combination therapy with immune checkpoint inhibitors. *Medicine (Baltimore)* **99**, e19741 (2020).
277. Okubo, N., Kijima, T., Nukui, A. & Kamai, T. Immune-related myositis resulting from combination therapy of ipilimumab and nivolumab in patient with metastatic renal cell carcinoma. *BMJ Case Rep* **13**(2020).
278. Onda, A., *et al.* Pembrolizumab-induced Ocular Myasthenia Gravis with Anti-titin Antibody and Necrotizing Myopathy. *Intern Med* **58**, 1635-1638 (2019).
279. Onderko, L.L., Heinrich, R., Gosling, K., Downs, T. & Afari, M.E. Myocarditis Following Immune Checkpoint Inhibition With Pembrolizumab: Management in a Context of Steroid Intolerance. *CJC Open* **4**, 854-857 (2022).
280. Ono, R., *et al.* Nivolumab-induced Myositis and Myocarditis with Positive Anti-titin Antibody and Anti-voltage-gated Potassium Channel Kv1.4 Antibody. *Intern Med* **61**, 2973-2979 (2022).
281. Ono, S., Nakamura, M., Morise, S., Kunieda, T. & Yakushiji, Y. [A case of atezolizumab- and bevacizumab-induced myositis showing high intensity in the pterygoid muscles, soft palate, and tongue on STIR-MRI]. *Rinsho Shinkeigaku* **63**, 582-587 (2023).
282. Osaki, M., Tachikawa, R., Ohira, J., Hara, S. & Tomii, K. Anti-transcriptional intermediary factor 1-gamma antibody-positive dermatomyositis induced by nivolumab for lung adenocarcinoma: A case report. *Invest New Drugs* **39**, 251-255 (2021).
283. Oshima, Y., Fujii, S. & Horiuchi, K. Pembrolizumab-induced Myasthenia Gravis Relapse after Immunosuppressive Therapy. *Intern Med* **61**, 3281-3285 (2022).
284. Otsuka, S., Horiuchi, K., Nagano, Y., Kimura, N. & Hiraoka, K. Myasthenia Gravis Associated With Pembrolizumab for Relapsed Lung Cancer After Thymoma Resection. *Cureus* **15**, e49767 (2023).
285. Ozarczuk, T.R.A., Prentice, D.A., Kho, L.K. & vanHeerden, J. Checkpoint inhibitor myasthenia-like syndrome and myositis associated with extraocular muscle atrophy. *J Clin Neurosci* **71**, 271-272 (2020).
286. Pandya, S.K., Ulrickson, M., Dong, J., Willen, R. & Pandya, A. Pembrolizumab-Associated Seronegative Myasthenia Gravis in a Patient With Metastatic Renal Cell Carcinoma. *Cureus* **13**, e15174 (2021).
287. Peters, N., Greally, M., Breen, K., Fabre, A. & Blazkova, S. Immunotherapy- A double Edged Sword; A case of Fatal Myocarditis and Complete Response. *Ir Med J* **112**, 937 (2019).

288. Peverelli, L., *et al.* Severe inflammatory myopathy in a pulmonary carcinoma patient treated with Pembrolizumab: An alert for myologists. *J Neuromuscul Dis* **7**, 511-514 (2020).
289. Phadke, S.D., *et al.* Pembrolizumab Therapy Triggering an Exacerbation of Preexisting Autoimmune Disease: A Report of 2 Patient Cases. *J Investig Med High Impact Case Rep* **4**, 2324709616674316 (2016).
290. Pilia, A.M., *et al.* Pembrolizumab-associated anti-MDA5 dermatomyositis in a patient with lung cancer: a first case report. *Swiss Med Wkly* **154**, 3513 (2024).
291. Polat, P. & Donofrio, P.D. Myasthenia gravis induced by nivolumab therapy in a patient with non-small-cell lung cancer. *Muscle Nerve* **54**, 507 (2016).
292. Portoles Hernandez, A., *et al.* Checkpoint inhibitor-induced fulminant myocarditis, complete atrioventricular block and myasthenia gravis-a case report. *Cardiovasc Diagn Ther* **11**, 1013-1019 (2021).
293. Prevel, R., Colin, G., Cales, V., Renault, P.A. & Mazieres, J. [Third degree atrio-ventricular blockade during a myocarditis occurring under anti-PD1 : Case report and literature review]. *Rev Med Interne* **41**, 284-288 (2020).
294. Punekar, S.R., Castillo, R., Sandigursky, S. & Cho, D.C. Role of IVIG in the Treatment of Autoimmune Conditions With Concurrent Immune Checkpoint Inhibitors for Metastatic Cancer. *J Immunother* **44**, 335-337 (2021).
295. Pushkarevskaya, A., Neuberger, U., Dimitrakopoulou-Strauss, A., Enk, A. & Hassel, J.C. Severe Ocular Myositis After Ipilimumab Treatment for Melanoma: A Report of 2 Cases. *J Immunother* **40**, 282-285 (2017).
296. Reddy, N., *et al.* Progressive and Reversible Conduction Disease With Checkpoint Inhibitors. *Can J Cardiol* **33**, 1335 e1313-1335 e1315 (2017).
297. Reyes-Bueno, J.A., Rodriguez-Santos, L. & Serrano-Castro, P.J. [Myasthenia gravis induced by inmuno checkpoints inhibitors: first case report secondary to avelumab therapy and review of published cases]. *Rev Neurol* **68**, 333-338 (2019).
298. Reynolds, K.L. & Guidon, A.C. Diagnosis and Management of Immune Checkpoint Inhibitor-Associated Neurologic Toxicity: Illustrative Case and Review of the Literature. *Oncologist* **24**, 435-443 (2019).
299. Roberts, J.H., Smylie, M., Oswald, A., Cusnir, I. & Ye, C. Hepatitis is the new myositis: immune checkpoint inhibitor-induced myositis. *Melanoma Res* **28**, 484-485 (2018).
300. Robinson, S.D., Lai, C., Hotton, G. & Anand, G. Life threatening pembrolizumabinduced myositis in a patient treated for advanced adenocarcinoma of the lung. *Acute Med* **18**, 197-199 (2019).
301. Rossi, V.A., *et al.* Value of troponin T versus I in the diagnosis of immune checkpoint inhibitor-related myocarditis and myositis: rechallenge? *ESC Heart Fail* **10**, 2680-2685 (2023).
302. Rota, E., *et al.* Concomitant myasthenia gravis, myositis, myocarditis and polyneuropathy, induced by immune-checkpoint inhibitors: A life-threatening continuum of neuromuscular and cardiac toxicity. *eNeurologicalSci* **14**, 4-5 (2019).
303. Ruperti-Repilado, F.J., *et al.* Case report of elevation of high-sensitivity cardiac troponin T in the absence of cardiac involvement in immune checkpoint inhibitor-associated myositis. *Eur Heart J Case Rep* **6**, ytac353 (2022).
304. Saad, R., Ghaddar, A. & Zeenny, R.M. Pembrolizumab-induced myocarditis with complete atrioventricular block and concomitant myositis in a metastatic bladder cancer patient: a case report and review of the literature. *J Med Case Rep* **18**, 107 (2024).
305. Sahin, E., Cabuk, D., Tuncer Kuru, F. & Yazici, A. Atezolizumab-induced myositis in a patient with small-cell lung cancer. *J Oncol Pharm Pract* **30**, 220-224 (2024).

306. Saibil, S.D., *et al.* Fatal myocarditis and rhabdomyositis in a patient with stage IV melanoma treated with combined ipilimumab and nivolumab. *Curr Oncol* **26**, e418-e421 (2019).
307. Saini, L. & Chua, N. Severe inflammatory myositis in a patient receiving concurrent nivolumab and azacitidine. *Leuk Lymphoma* **58**, 2011-2013 (2017).
308. Saishu, Y., Yoshida, T., Seino, Y. & Nomura, T. Nivolumab-related myasthenia gravis with myositis requiring prolonged mechanical ventilation: a case report. *J Med Case Rep* **16**, 61 (2022).
309. Sakai, S., Tajiri, K., Li, S. & Ieda, M. Fatal cerebral haemorrhagic infarction due to left ventricular thrombus after healing of immune checkpoint inhibitor-associated myocarditis. *Eur Heart J Case Rep* **4**, 1-2 (2020).
310. Sakai, T., *et al.* Acute myocarditis and pericarditis after nivolumab treatment in patients with non-small cell lung cancer. *Annals of Oncology* **28**(2017).
311. Sakurai, T., Takahashi, J., Komatsu, T., Mitsumura, H. & Iguchi, Y. Anti-TIF1gamma antibody-positive dermatomyositis associated with nivolumab administration in a patient with advanced oesophageal squamous cell carcinoma: A case report and literature review. *Mod Rheumatol Case Rep* **7**, 416-421 (2023).
312. Salem, J.E., *et al.* Abatacept for Severe Immune Checkpoint Inhibitor-Associated Myocarditis. *N Engl J Med* **380**, 2377-2379 (2019).
313. Salido Iniesta, M., Lopez Lopez, L., Carreras Costa, F. & Sionis, A. A different type of acute myocarditis: a case report of acute autoimmune myocarditis mediated by anti-PD-1 T lymphocyte receptor (pembrolizumab). *Eur Heart J Case Rep* **4**, 1-6 (2020).
314. Samara, Y., Yu, C.L. & Dasanu, C.A. Acute autoimmune myocarditis and hepatitis due to ipilimumab monotherapy for malignant melanoma. *J Oncol Pharm Pract* **25**, 966-968 (2019).
315. Sanchez-Sancho, P., Selva-O'Callaghan, A., Trallero-Araguas, E., Ros, J. & Montoro, B. Myositis and myasteniform syndrome related to pembrolizumab. *BMJ Case Rep* **14**(2021).
316. Sato, T., *et al.* Monitoring of the Evolution of Immune Checkpoint Inhibitor Myocarditis With Cardiovascular Magnetic Resonance. *Circ Cardiovasc Imaging* **13**, e010633 (2020).
317. Sauer, R., *et al.* [Lymphocytic myocarditis in a patient with metastatic clear cell renal cell carcinoma treated with Nivolumab]. *Pathologe* **38**, 535-539 (2017).
318. Sawai, T., *et al.* [An autopsy case of nivolumab-induced myasthenia gravis and myositis]. *Rinsho Shinkeigaku* **59**, 360-364 (2019).
319. Schiopu, S.R.I., *et al.* Pembrolizumab-induced myocarditis in a patient with malignant mesothelioma: plasma exchange as a successful emerging therapy-case report. *Transl Lung Cancer Res* **10**, 1039-1046 (2021).
320. Schouwenburg, J.J. Nivolumab-induced Diaphragm Dysfunction: A Case Report. *Indian J Crit Care Med* **27**, 147-148 (2023).
321. Schwab, A., *et al.* Pembrolizumab-Induced Myasthenia Gravis and Myositis: Literature Review on Neurological Toxicities of Programmed Death Protein 1 Inhibitors. *J Med Cases* **13**, 530-535 (2022).
322. Sciacca, G., *et al.* Benign form of myasthenia gravis after nivolumab treatment. *Muscle Nerve* **54**, 507-509 (2016).
323. Seki, M., *et al.* Inflammatory myopathy associated with PD-1 inhibitors. *J Autoimmun* **100**, 105-113 (2019).
324. Sekiguchi, K., *et al.* Diaphragm involvement in immune checkpoint inhibitor-related myositis. *Muscle Nerve* **60**, E23-E25 (2019).
325. Semper, H., Muehlberg, F., Schulz-Menger, J., Allewelt, M. & Grohe, C. Drug-induced myocarditis after nivolumab treatment in a patient with PDL1- negative squamous cell carcinoma of the lung. *Lung Cancer* **99**, 117-119 (2016).

326. Sessums, M., Yarrarapu, S., Guru, P.K. & Sanghavi, D.K. Atezolizumab-induced myositis and myocarditis in a patient with metastatic urothelial carcinoma. *BMJ Case Rep* **13**(2020).
327. Shah, D. & Young, K. Exploring Pembrolizumab-Induced Myocarditis, Myositis, and Myasthenia Gravis: A Comprehensive Literature Review and Case Presentation on Bladder Cancer. *Cureus* **15**, e49867 (2023).
328. Shah, M., Tayar, J.H., Abdel-Wahab, N. & Suarez-Almazor, M.E. Myositis as an adverse event of immune checkpoint blockade for cancer therapy. *Semin Arthritis Rheum* **48**, 736-740 (2019).
329. Shalata, W.*, et al.* Tolerated Re-Challenge of Immunotherapy in a Patient with ICI Associated Myocarditis: A Case Report and Literature Review. *Medicina (Kaunas)* **59**(2023).
330. Shalata, W.*, et al.* Associated Myocarditis: A Predictive Factor for Response? *Case Rep Oncol* **13**, 550-557 (2020).
331. Shalata, W.*, et al.* Perimyocarditis Associated with Immune Checkpoint Inhibitors: A Case Report and Review of the Literature. *Medicina (Kaunas)* **60**(2024).
332. Sheik Ali, S.*, et al.* Drug-associated dermatomyositis following ipilimumab therapy: a novel immune-mediated adverse event associated with cytotoxic T-lymphocyte antigen 4 blockade. *JAMA Dermatol* **151**, 195-199 (2015).
333. Shen, L., Chen, H. & Wei, Q. Immune-Therapy-Related Toxicity Events and Dramatic Remission After a Single Dose of Pembrolizumab Treatment in Metastatic Thymoma: A Case Report. *Front Immunol* **12**, 621858 (2021).
334. Shibata, C.*, et al.* Paraneoplastic dermatomyositis appearing after nivolumab therapy for gastric cancer: a case report. *J Med Case Rep* **13**, 168 (2019).
335. Shikano, K.*, et al.* Nivolumab-induced anti-aminoacyl-tRNA synthetase antibody-positive polymyositis complicated by interstitial pneumonia in a patient with lung adenocarcinoma. *Scand J Rheumatol* **49**, 82-83 (2020).
336. Shindo, A., Yamasaki, M., Uchino, K. & Yamasaki, M. Asymptomatic Myocarditis with Mild Cardiac Marker Elevation Following Nivolumab-Induced Myositis. *Int Heart J* **63**, 180-183 (2022).
337. Shirai, T.*, et al.* Presence of antibodies to striated muscle and acetylcholine receptor in association with occurrence of myasthenia gravis with myositis and myocarditis in a patient with melanoma treated with an anti-programmed death 1 antibody. *Eur J Cancer* **106**, 193-195 (2019).
338. Shirai, T.*, et al.* Acetylcholine receptor binding antibody-associated myasthenia gravis and rhabdomyolysis induced by nivolumab in a patient with melanoma. *Jpn J Clin Oncol* **46**, 86-88 (2016).
339. Sibille, A.*, et al.* Granulomatosis With Polyangiitis in a Patient on Programmed Death-1 Inhibitor for Advanced Non-small-cell Lung Cancer. *Front Oncol* **9**, 478 (2019).
340. So, H.*, et al.* PD-1 inhibitor-associated severe myasthenia gravis with necrotizing myopathy and myocarditis. *J Neurol Sci* **399**, 97-100 (2019).
341. Soman, B., Dias, M.C., Rizvi, S.A.J. & Kardos, A. Myasthenia gravis, myositis and myocarditis: a fatal triad of immune-related adverse effect of immune checkpoint inhibitor treatment. *BMJ Case Rep* **15**(2022).
342. Stein-Merlob, A.F.*, et al.* Keeping immune checkpoint inhibitor myocarditis in check: advanced circulatory mechanical support as a bridge to recovery. *ESC Heart Fail* **8**, 4301-4306 (2021).
343. Su, L.*, et al.* Successful Therapy for Myocarditis Concomitant With Complete Heart Block After Pembrolizumab Treatment for Head and Neck Squamous Cell Carcinoma: A Case Report With Literature Review. *Front Cardiovasc Med* **9**, 898756 (2022).

344. Sugiyama, Y., *et al.* [Immune checkpoint inhibitor-induced anti-striational antibodies in myasthenia gravis and myositis: a case report]. *Rinsho Shinkeigaku* **61**, 630-634 (2021).
345. Sun, R., Shah, V., Tummala, S. & Chen, M. Nivolumab-induced myasthenia gravis with myositis in patients with genitourinary cancer (P4.2-005). *Neurology* **92**, P4.2-005 (2019).
346. Swali, R. Pembrolizumab-induced Myositis in the Setting of Metastatic Melanoma: An Increasingly Common Phenomenon. *J Clin Aesthet Dermatol* **13**, 44-45 (2020).
347. Szuchan, C., *et al.* Checkpoint inhibitor-induced myocarditis and myasthenia gravis in a recurrent/metastatic thymic carcinoma patient: a case report. *Eur Heart J Case Rep* **4**, 1-8 (2020).
348. Tadokoro, T., *et al.* Acute Lymphocytic Myocarditis With Anti-PD-1 Antibody Nivolumab. *Circ Heart Fail* **9**(2016).
349. Tahir, N., *et al.* Nivolumab, a Double-Edged Sword: A Case Report of Nivolumab-Induced Myasthenia Gravis. *J Med Cases* **12**, 424-428 (2021).
350. Tajmir-Riahi, A., *et al.* Life-threatening Autoimmune Cardiomyopathy Reproducibly Induced in a Patient by Checkpoint Inhibitor Therapy. *J Immunother* **41**, 35-38 (2018).
351. Takahashi, S., Mukohara, S., Hatachi, S., Yamashita, M. & Kumagai, S. A case of myositis with dropped head syndrome and anti-titin antibody positivity induced by pembrolizumab. *Scand J Rheumatol* **49**, 509-511 (2020).
352. Takai, M., *et al.* Simultaneous pembrolizumab-induced myasthenia gravis and myocarditis in a patient with metastatic bladder cancer: A case report. *Urol Case Rep* **31**, 101145 (2020).
353. Takatsuki, K., *et al.* A Rare Case of Pembrolizumab-Induced Dermatomyositis in a Patient with Cancer of Unknown Primary Origin. *Am J Case Rep* **22**, e930286 (2021).
354. Tan, J.L., Mugwagwa, A.N., Cieslik, L. & Joshi, R. Nivolumab-induced myocarditis complicated by complete atrioventricular block in a patient with metastatic non-small cell lung cancer. *BMJ Case Rep* **12**(2019).
355. Tan, K.X., *et al.* [Fulminant myocarditis caused by nivolumab treatment for non-small cell lung cancer (NSCLC): a case report]. *Zhonghua Zhong Liu Za Zhi* **42**, 1047-1048 (2020).
356. Tan, N.Y.L., Anavekar, N.S. & Wiley, B.M. Concomitant myopericarditis and takotsubo syndrome following immune checkpoint inhibitor therapy. *BMJ Case Rep* **13**(2020).
357. Tan, R.Y.C., Toh, C.K. & Takano, A. Continued Response to One Dose of Nivolumab Complicated by Myasthenic Crisis and Myositis. *J Thorac Oncol* **12**, e90-e91 (2017).
358. Tanabe, J., *et al.* Asymptomatic Immune Checkpoint Inhibitor-associated Myocarditis. *Intern Med* **60**, 569-573 (2021).
359. Tanaka, R., Sunada, Y. & Fujimoto, W. Successful reinstitution of nivolumab in combination with corticosteroids for metastatic malignant melanoma with myasthenia gravis as an immune-related adverse event. *Kawasaki Medical Journal* **43**, 59-61 (2017).
360. Tauber, M., *et al.* Severe necrotizing myositis associated with long term anti-neoplastic efficacy following nivolumab plus ipilimumab combination therapy. *Clin Rheumatol* **38**, 601-602 (2019).
361. Tay, R.Y., *et al.* Successful use of equine anti-thymocyte globulin (ATGAM) for fulminant myocarditis secondary to nivolumab therapy. *Br J Cancer* **117**, 921-924 (2017).
362. Tay, S.H., Wong, A.S. & Jeyasekharan, A.D. A patient with pembrolizumab-induced fatal polymyositis. *Eur J Cancer* **91**, 180-182 (2018).
363. Tedbirt, B., *et al.* Rechallenge of immune checkpoint inhibitor after pembrolizumab-induced myasthenia gravis. *Eur J Cancer* **113**, 72-74 (2019).
364. Thakolwiboon, S., Karukote, A. & Wilms, H. De Novo Myasthenia Gravis Induced by Atezolizumab in a Patient with Urothelial Carcinoma. *Cureus* **11**, e5002 (2019).

365. Thibault, C., Vano, Y., Soulat, G. & Mirabel, M. Immune checkpoint inhibitors myocarditis: not all cases are clinically patent. *Eur Heart J* **39**, 3553 (2018).
366. Thomas, R., Patel, H. & Scott, J. Dermatomyositis Flare With Immune Checkpoint Inhibitor Therapy for Melanoma. *Cureus* **13**, e14387 (2021).
367. Tian, C.Y., Ou, Y.H., Chang, S.L. & Lin, C.M. Pembrolizumab-induced myasthenia gravis-like disorder, ocular myositis, and hepatitis: a case report. *J Med Case Rep* **15**, 244 (2021).
368. Todo, M., *et al.* Pembrolizumab-induced myasthenia gravis with myositis and presumable myocarditis in a patient with bladder cancer. *IJU Case Rep* **3**, 17-20 (2020).
369. Tokunaga, T., *et al.* Fulminant myocarditis during postoperative adjuvant chemotherapy for lung cancer with atezolizumab: a case report. *J Med Case Rep* **18**, 162 (2024).
370. Tomoaia, R., Beyer, R.S., Pop, D., Minciuna, I.A. & Dadarlat-Pop, A. Fatal association of fulminant myocarditis and rhabdomyolysis after immune checkpoint blockade. *Eur J Cancer* **132**, 224-227 (2020).
371. Touat, M., *et al.* Immune checkpoint inhibitor-related myositis and myocarditis in patients with cancer. *Neurology* **91**, e985-e994 (2018).
372. Tozuka, T., *et al.* Pembrolizumab-induced agranulocytosis in a pulmonary pleomorphic carcinoma patient who developed interstitial lung disease and ocular myasthenia gravis. *Oxf Med Case Reports* **2018**, omy094 (2018).
373. Uchio, N., *et al.* Pembrolizumab on pre-existing inclusion body myositis: a case report. *BMC Rheumatol* **4**, 48 (2020).
374. Unluturk, Z., Karagulmez, A.M., Hayti, B. & Erdogan, C. Myocarditis-myositis-myasthenia gravis overlap syndrome depending on immune checkpoint inhibitor. *J Neurosci Rural Pract* **14**, 143-144 (2023).
375. Valenti-Azcarate, R., Esparragosa Vazquez, I., Toledano Illan, C., Idoate Gastearena, M.A. & Gallego Perez-Larraya, J. Nivolumab and Ipilimumab-induced myositis and myocarditis mimicking a myasthenia gravis presentation. *Neuromuscul Disord* **30**, 67-69 (2020).
376. Vallet, H., *et al.* Pembrolizumab-induced necrotic myositis in a patient with metastatic melanoma. *Ann Oncol* **27**, 1352-1353 (2016).
377. Veccia, A., *et al.* Myositis and myasthenia during nivolumab administration for advanced lung cancer: a case report and review of the literature. *Anticancer Drugs* **31**, 540-544 (2020).
378. Verma, N., Jaffer, M., Pina, Y., Peguero, E. & Mokhtari, S. Rituximab for Immune Checkpoint Inhibitor Myasthenia Gravis. *Cureus* **13**, e16337 (2021).
379. Vermeulen, L., *et al.* Myositis as a neuromuscular complication of immune checkpoint inhibitors. *Acta Neurol Belg* **120**, 355-364 (2020).
380. Vivas, A.J., Chaudhry, U., Punchayil Narayanankutty, N., Lopez, R. & Lamarche, J. Myasthenia Gravis-Like Syndrome Resulting From Immune Checkpoint Inhibitors in a Patient With Urothelial Carcinoma. *Cureus* **16**, e60003 (2024).
381. von Itzstein, M.S., *et al.* Statin Intolerance, Anti-HMGCR Antibodies, and Immune Checkpoint Inhibitor-Associated Myositis: A "Two-Hit" Autoimmune Toxicity or Clinical Predisposition? *Oncologist* **25**, e1242-e1245 (2020).
382. Wai Siu, D.H., *et al.* Immune checkpoint inhibitor-induced myocarditis, myositis, myasthenia gravis and transaminitis: a case series and review. *Immunotherapy* **14**, 511-520 (2022).
383. Wakefield, C., Shultz, C., Patel, B. & Malla, M. Life-threatening immune checkpoint inhibitor-induced myocarditis and myasthenia gravis overlap syndrome treated with abatacept: a case report. *BMJ Case Rep* **14**(2021).
384. Wang, C., Zhong, B., He, J. & Liao, X. Immune checkpoint inhibitor sintilimab-induced lethal myocarditis overlapping with myasthenia gravis in thymoma patient: A case report. *Medicine (Baltimore)* **102**, e33550 (2023).

385. Wang, F., Gong, X.L., Geng, H.Y., Cheng, Y. & Chen, X.N. [A case of asymptomatic immune checkpoint inhibitor associated myocarditis and myositis]. *Zhonghua Xin Xue Guan Bing Za Zhi* **50**, 1103-1105 (2022).
386. Wang, F.*, et al.* Fulminant myocarditis induced by immune checkpoint inhibitor nivolumab: a case report and review of the literature. *J Med Case Rep* **15**, 336 (2021).
387. Wang, F.*, et al.* A retrospective study of immune checkpoint inhibitor-associated myocarditis in a single center in China. *Chin Clin Oncol* **9**, 16 (2020).
388. Wang, H., Tian, R., Gao, P., Wang, Q. & Zhang, L. Tocilizumab for Fulminant Programmed Death 1 Inhibitor-Associated Myocarditis. *J Thorac Oncol* **15**, e31-e32 (2020).
389. Wang, Q. & Hu, B. Successful therapy for autoimmune myocarditis with pembrolizumab treatment for nasopharyngeal carcinoma. *Ann Transl Med* **7**, 247 (2019).
390. Wang, S.*, et al.* Acetylcholine receptor binding antibody-associated myasthenia gravis, myocarditis, and rhabdomyolysis induced by tislelizumab in a patient with colon cancer: A case report and literature review. *Front Oncol* **12**, 1053370 (2022).
391. Watson, R.A.*, et al.* Severe acute myositis and myocarditis on initiation of 6-weekly pembrolizumab post-COVID-19 mRNA vaccination. *J Immunother Cancer* **12**(2024).
392. Weaver, J.M.*, et al.* Improved outcomes with early immunosuppression in patients with immune-checkpoint inhibitor induced myasthenia gravis, myocarditis and myositis: a case series. *Support Care Cancer* **31**, 518 (2023).
393. Werner, J.M.*, et al.* Successful Treatment of Myasthenia Gravis Following PD-1/CTLA-4 Combination Checkpoint Blockade in a Patient With Metastatic Melanoma. *Front Oncol* **9**, 84 (2019).
394. Wiggins, C.J. & Chon, S.Y. Dermatomyositis, pembrolizumab, and squamous cell carcinoma of the lung. *Proc (Bayl Univ Med Cent)* **34**, 120-121 (2020).
395. Wintersperger, B.J.*, et al.* Immune checkpoint inhibitor-related myocarditis: an illustrative case series of applying the updated Cardiovascular Magnetic Resonance Lake Louise Criteria. *Eur Heart J Case Rep* **6**, ytab478 (2022).
396. Wong, E.Y.T.*, et al.* Immune checkpoint inhibitor-associated myositis and myasthenia gravis overlap: Understanding the diversity in a case series. *Asia Pac J Clin Oncol* **17**, e262-e267 (2021).
397. Wu, S., Shi, J., Guan, Y., Zhang, L. & Wang, H. Successful Management of Generalized Myasthenia Gravis Induced by Atezolizumab in a Patient With Extensive-Stage SCLC: A Case Report. *JTO Clin Res Rep* **3**, 100354 (2022).
398. Wu, S.J.*, et al.* [Tocilizumab therapy for immune checkpoint inhibitor associated myocarditis: a case report]. *Zhonghua Xin Xue Guan Bing Za Zhi* **50**, 397-400 (2022).
399. Xie, X.*, et al.* Case Report: Fatal Multiorgan Failure and Heterochronous Pneumonitis Following Pembrolizumab Treatment in a Patient With Large-Cell Neuroendocrine Carcinoma of Lung. *Front Pharmacol* **11**, 569466 (2020).
400. Xing, Q.*, et al.* Case Report: Treatment for steroid-refractory immune-related myocarditis with tofacitinib. *Front Immunol* **13**, 944013 (2022).
401. Xing, Q.*, et al.* Myositis-myasthenia gravis overlap syndrome complicated with myasthenia crisis and myocarditis associated with anti-programmed cell death-1 (sintilimab) therapy for lung adenocarcinoma. *Ann Transl Med* **8**, 250 (2020).
402. Yamaguchi, S.*, et al.* Late-Onset Fulminant Myocarditis With Immune Checkpoint Inhibitor Nivolumab. *Can J Cardiol* **34**, 812 e811-812 e813 (2018).
403. Yamaguchi, Y., Abe, R., Haga, N. & Shimizu, H. A case of drug-associated dermatomyositis following ipilimumab therapy. *Eur J Dermatol* **26**, 320-321 (2016).

404. Yamaguchi, Y., Aso, M., Nagasawa, H. & Wada, M. Atezolizumab-associated Dermatomyositis in Advanced Small-cell Lung Carcinoma. *Intern Med* **60**, 3025-3029 (2021).
405. Yanase, T.*, et al.* Myocarditis and myasthenia gravis by combined nivolumab and ipilimumab immunotherapy for renal cell carcinoma: A case report of successful management. *Urol Case Rep* **34**, 101508 (2021).
406. Yang, Y., Wu, Q., Chen, L., Qian, K. & Xu, X. Severe immune-related hepatitis and myocarditis caused by PD-1 inhibitors in the treatment of triple-negative breast cancer: a case report. *Ann Transl Med* **10**, 424 (2022).
407. Yang, Y.*, et al.* Anti-PD-1 and regorafenib induce severe multisystem adverse events in microsatellite stability metastatic colorectal cancer: a case report. *Immunotherapy* **13**, 1317-1323 (2021).
408. Yang, Z.X., Chen, X., Tang, S.Q. & Zhang, Q. Sintilimab-Induced Myocarditis Overlapping Myositis in a Patient With Metastatic Thymoma: A Case Report. *Front Cardiovasc Med* **8**, 797009 (2021).
409. Ye, Y., Li, Y., Zhang, S. & Han, G. Teriprizumab-induced myocarditis in a patient with cholangiocarcinoma: a case report. *J Int Med Res* **50**, 3000605221133259 (2022).
410. Yin, B.*, et al.* Myocarditis and myositis/myasthenia gravis overlap syndrome induced by immune checkpoint inhibitor followed by esophageal hiatal hernia: A case report and review of the literature. *Front Med (Lausanne)* **9**, 950801 (2022).
411. Yin, N.*, et al.* PD-1 inhibitor therapy causes multisystem immune adverse reactions: a case report and literature review. *Front Oncol* **12**, 961266 (2022).
412. Yogasundaram, H.*, et al.* Plasma Exchange for Immune Checkpoint Inhibitor-Induced Myocarditis. *CJC Open* **3**, 379-382 (2021).
413. Yoshioka, M., Kambe, N., Yamamoto, Y., Suehiro, K. & Matsue, H. Case of respiratory discomfort due to myositis after administration of nivolumab. *J Dermatol* **42**, 1008-1009 (2015).
414. Yuen, C., Fleming, G., Meyers, M., Soliven, B. & Rezania, K. Myasthenia gravis induced by avelumab. *Immunotherapy* **11**, 1181-1185 (2019).
415. Zadeh, S.*, et al.* Novel uses of complement inhibitors in myasthenia gravis-Two case reports. *Muscle Nerve* **69**, 368-372 (2024).
416. Zarkavelis, G.*, et al.* The cancer immunotherapy environment may confound the utility of anti-TIF-1gamma in differentiating between paraneoplastic and treatment-related dermatomyositis. Report of a case and review of the literature. *Contemp Oncol (Pozn)* **24**, 75-78 (2020).
417. Zhang, B., Gyawali, L., Liu, Z., Du, H. & Yin, Y. Camrelizumab-Related Lethal Arrhythmias and Myasthenic Crisis in a Patient with Metastatic Thymoma. *Case Rep Cardiol* **2022**, 4042909 (2022).
418. Zhang, C., Qin, S. & Zuo, Z. Immune-related myocarditis in two patients receiving camrelizumab therapy and document analysis. *J Oncol Pharm Pract* **28**, 1350-1356 (2022).
419. Zhang, J., Li, J., Zhai, L. & Lin, L. Coexisting of myasthenia gravis and fulminant myocarditis induced by nivolumab in a patient with ureteral epithelial cancer. *Neuro Endocrinol Lett* **42**, 383-386 (2021).
420. Zheng, S.*, et al.* A case of acute myocarditis induced by PD-1 inhibitor (sintilimab) in the treatment of large cell neuroendocrine carcinoma. *Heliyon* **9**, e16874 (2023).
421. Zhong, P.*, et al.* Myocarditis and myasthenia gravis induced by immune checkpoint inhibitor in a patient with relapsed thymoma: A case report. *Clin Case Rep* **11**, e7039 (2023).

422. Zhou, B., Li, M., Chen, T. & She, J. Case Report: Acute Myocarditis Due to PD-L1 Inhibitor Durvalumab Monotherapy in a Patient With Lung Squamous Cell Carcinoma. *Front Med (Lausanne)* **9**, 866068 (2022).
423. Zhu, J. & Li, Y. Myasthenia gravis exacerbation associated with pembrolizumab. *Muscle Nerve* **54**, 506-507 (2016).
424. Zimmer, L., *et al.* Neurological, respiratory, musculoskeletal, cardiac and ocular side-effects of anti-PD-1 therapy. *Eur J Cancer* **60**, 210-225 (2016).
425. Ziobro, A.S., *et al.* Myasthenia Gravis Associated With Programmed Death-1 (PD-1) Receptor Inhibitor Pembrolizumab: A 40-day Case Report. *J Pharm Pract* **34**, 166-170 (2021).
426. Zomborska, E., *et al.* Fatal myocarditis after the first dose of nivolumab. *Klin Onkol* **35**, 486-492 (2022).

***F. Thymoma Spontaneous Regression***

1. Alnassar, A. Spontaneous Regression of a Thymoma with Mild Pleural Effusion: A Case Report. *Int J Innov Res Med Sci* **8**, 17-19 (2023).
2. Chiyotanda, T.*, et al.* [Preoperative Spontaneous Regression of Thymoma]. *Kyobu Geka* **73**, 358-361 (2020).
3. Doutsu, Y.*, et al.* [A case of Good's syndrome with thymoma that regressed spontaneously--a review of clinical feature in 14 cases of Good's syndrome reported in Japan]. *Nihon Kyobu Shikkan Gakkai Zasshi* **26**, 770-777 (1988).
4. Fukui, T., Taniguchi, T., Kawaguchi, K. & Yokoi, K. Spontaneous regression of thymic epithelial tumours. *Interact Cardiovasc Thorac Surg* **18**, 399-401 (2014).
5. Furuya, K.*, et al.* Thymoma exhibiting spontaneous regression in size, pleural effusion and serum cytokeratin fragment level: A case report. *Mol Clin Oncol* **3**, 1058-1062 (2015).
6. Hayakawa, M., Oda, K. & Tomita, E. [Thymoma showing enlargement and reduction in size with inflammatory events;report of a case]. *Kyobu Geka* **67**, 508-511 (2014).
7. Hayakawa, T. 経過中に腫瘍内壊死により自然縮小した胸腺腫の 1 例 [A case of thymoma that spontaneously regressed due to intratumoral necrosis during the clinical course]. *J Jpn Assoc Chest Surg* **28**, S15 (2014).
8. Hori, D., Endo, S., Tsubochi, H., Nokubi, M. & Sohara, Y. Spontaneous regression of symptomatic thymoma caused by infarction. *Gen Thorac Cardiovasc Surg* **56**, 468-471 (2008).
9. Hosono, Y., Watanabe, S., Watanabe, S. & Mitsui, M. 胸部痛・発熱を契機に発見され, 術前に自然縮小を来した重症筋無力症合併胸腺腫の 1 例 [A case of thymoma with myasthenia gravis detected due to chest pain and fever, showing spontaneous regression before surgery]. *Jpn J Lung Cancer* **57**, 509 (2017).
10. Huang, T.W.*, et al.* Spontaneous regression of a mediastinal thymoma. *J Thorac Cardiovasc Surg* **137**, 1277-1278 (2009).
11. Hyogotani, A.*, et al.* 経過中に一過性の胸水貯留と自然縮小を認めた胸腺腫の 1 例 [A case of thymoma with transient pleural effusion and spontaneous regression during the clinical course]. *J Jpn Assoc Chest Surg* **24**, 613 (2010).
12. Iijima, Y.*, et al.* Sclerosing thymoma followed up for eight years as mediastinal goiter: A case report. *Int J Surg Case Rep* **68**, 115-118 (2020).
13. Imamura, N.*, et al.* 一過性の自然縮小と胸水貯留を認めた胸腺腫の 1 切除例 [A resected case of thymoma with transient spontaneous regression and pleural effusion]. *J Jpn Assoc Chest Surg* **25**, S12 (2011).
14. Inoue, H., Takahashi, M., Matsui, H. & Sato, Y. 自然退縮を認めた胸腺腫の 2 例 [Two cases of thymoma with spontaneous regression]. *J Jpn Assoc Chest Surg* **23**, 456 (2009).
15. Ishibashi, H.*, et al.* [Spontaneous regression of thymoma; report of a case]. *Kyobu Geka* **56**, 801-805 (2003).
16. Itoh, H.*, et al.* [A case of thymoma with spontaneous regression after rapid growth]. *J Jpn Assoc Chest Surg* **20**, 974-979 (2006).
17. Kadomatsu, Y., Kawasumi, Y., Ueno, H., Uchiyama, M. & Mori, S. 妊娠中に胸痛を契機に発見され,経過中に自然縮小を認めた胸腺腫の 1 手術例 [A surgical case of thymoma detected by chest pain during pregnancy that showed spontaneous regression during follow-up]. *Jpn J Lung Cancer* **57**, 508 (2017).
18. Kaneshiro, T., Abe, K., Kajiwara, K., Yamamoto, A. & Sato, M. 自然縮小を示した胸腺腫の一例 [A case of thymoma that showed spontaneous regression]. *J. Kanagawa Med. Ass* **33**, 75 (2006).

19. Kasai, Y. & Masuya, D. [A case of thymoma that showed repeated growth and regression]. *J Jpn Assoc Chest Surg* **30**, 36-39 (2016).
20. Kataoka, K., Seno, N. & Matsuura, M. [Thymoma regressing spontaneously; a case report]. *J Jpn Assoc Chest Surg* **8**, 49-53 (1994).
21. Kato, M.*, et al.* 自然縮小した胸腺腫の 1 例 [A case of spontaneously regressed thymoma]. *Jpn J Lung Cancer* **49**, 802 (2009).
22. Kikuchi, N.*, et al.* Spontaneous Regression of Type B3 Thymoma With Mesothelial Cyst: A Case Report. *J Thorac Imaging* **35**, W123-W126 (2020).
23. Kim, Y.H., Kim, J.J., Jeong, S.C. & Kim, I.S. Complete excision of acute necrotic regression of thymoma mimicking an infected mediastinal cyst with mediastinitis using video-assisted thoracoscopic technique. *J Thorac Dis* **10**, E364-E367 (2018).
24. Maeda, A., Nojima, Y., Saisho, S., Shimizu, K. & Nakata, M. [A case of spontaneously regressed thymoma]. *Kawasaki Med J* **51**, 1-6 (2025).
25. Makiuchi, A. & Hikita, H. 自然縮小を認めた胸腺腫の 1 例 [A case of thymoma with spontaneous regression]. *J Jpn Assoc Chest Surg* **22**, 489 (2008).
26. Matsuura, S., Kiyoshima, M., Kitahara, M., Suzuki, H. & Asato, Y. 経過中に自然縮小した胸腺腫の 1 例 [A case of thymoma that spontaneously regressed during the clinical course]. *Jpn J Lung Cancer* **55**, 121 (2015).
27. Miyazaki, H.*, et al.* 胸腺梗塞による自然縮小を認めた胸腺腫の 2 例 [Two cases of thymoma that showed spontaneous regression due to thymic infarction]. *J Jpn Assoc Chest Surg* **24**, 612 (2010).
28. Nakazono, T.*, et al.* Magnetic resonance imaging features of spontaneously regressed thymoma: report of 2 cases. *J Thorac Imaging* **24**, 62-65 (2009).
29. Nishina, K.*, et al.* Thymoma exhibiting spontaneous regression with developing myasthenia gravis: A case report. *Thorac Cancer* **13**, 1533-1536 (2022).
30. Nojima, T.*, et al.* 胸膜炎症状で発症し自然縮小した胸腺腫の 2 例 [Two cases of thymoma that presented with pleuritic symptoms and underwent spontaneous regression]. *Jpn J Lung Cancer* **56**, 1082 (2016).
31. Okagawa, T., Uchida, T. & Suyama, M. Thymoma with spontaneous regression and disappearance of pleural effusion. *Gen Thorac Cardiovasc Surg* **55**, 515-517 (2007).
32. Okamoto, T.*, et al.* 自然縮小を示した胸腺腫の２例 [Two cases of thymoma that showed spontaneous regression]. *J Jpn Assoc Chest Surg* **23**, 456 (2009).
33. Omori, T.*, et al.* 自然縮小を来した胸腺腫の 1 例 [A case of spontaneously regressed thymoma]. *Jpn J Lung Cancer* **57**, 132 (2017).
34. Ouchi, M., Inoue, S., Ozaki, Y. & Fujita, T. 自然縮小を認めた胸腺腫の１切除例 [A resected case of thymoma with spontaneous regression]. *Jpn J Lung Cancer* **49**, 803 (2009).
35. Sakaguchi, Y., Komatsu, T., Takubo, Y. & Terada, Y. Resected case of giant cystic thymoma with spontaneous intracystic hemorrhage. *Surg Case Rep* **5**, 30 (2019).
36. Shimomura, A.*, et al.* 2 ケ月の経過で縮小傾向を認めた胸腺腫の 1 手術例 [A surgical case of a thymoma showing a tendency to shrink over a two-month course]. *Jpn J Lung Cancer* **49**, 803 (2009).
37. Suzuki, M., Shimizu, R., Harada, M., Hishima, T. & Horio, H. Thymoma Exhibiting Spontaneous Regression With Cystic Change Due to Acute Infarction: A Case Report and Literature Review. *Cureus* **16**, e56240 (2024).
38. Toishi, M.*, et al.* 炎症所見を伴う急速増大後に自然縮小を示した胸腺腫 1 例 [A case of thymoma that showed spontaneous regression after rapid enlargement with inflammatory findings]. *Jpn J Lung Cancer* **51**, 509 (2011).

39. Tokunaga, T., Ose, N., Nagata, H., Morii, E. & Shintani, Y. Necrotic Thymoma Discovered Due to Subjective Symptoms: A Report of Three Cases. *Surg Case Rep* **11**(2025).
40. Tomiyama, K., Ishida, H., Miyake, M. & Taki, T. 自然縮小を示した胸腺腫の 1 手術例 [A surgically treated case of thymoma that showed spontaneous regression]. *Jpn J Lung Cancer* **41**, 537 (2001).
41. Toyokawa, G., *et al.* Regression of thymoma associated with a multilocular thymic cyst: report of a case. *Surg Today* **44**, 577-580 (2014).
42. Toyooka, S., *et al.* 画像上自然縮小を示し重症筋無力症の発症とともに再び増大した胸腺腫の一例 [A case of thymoma that showed radiologic spontaneous regression and then regrew with the onset of myasthenia gravis]. *J Jpn Assoc Chest Surg* **17**, 329 (2003).
43. Tsunooka, N., Hirayama, K., Matsuda, F. & Inazawa, K. [Preoperative spontaneous regression of type B3 thymoma;report of a case]. *Kyobu Geka* **68**, 153-156 (2015).
44. Yamashita, M., *et al.* 術前に自然縮小した胸腺腫の 2 手術例 [Two surgically treated cases of thymoma that spontaneously regressed preoperatively]. *J Jpn Assoc Chest Surg* **30**, S24 (2016).
45. Yasukawa, M., Kawaguchi, T., Kawai, N. & Tojo, T. [Spontaneous regression of thymona: Report of two cases]. *J Jpn Assoc Chest Surg* **31**, 36-41 (2017).
46. Yoshida, H., *et al.* 自然縮小を示した胸腺腫の 1 例 [A case of thymoma that showed spontaneous regression]. *Jpn J Lung Cancer* **42**, 481 (2002).
47. Yutaka, Y., Omasa, M., Shikuma, K., Okuda, M. & Taki, T. Spontaneous regression of an invasive thymoma. *Gen Thorac Cardiovasc Surg* **57**, 272-274 (2009).

### Appendix 3: Excluded Studies and Reasons for Exclusion After Full-Text Screening

#### *A. Concomitant IIM and MG*

• <u>*Lack of evidence for at least possible myositis (excluding DM without muscle involvement)*</u>:

1. Agarwal, P., Chapagain, U., Deewan, K.R. & Rana, P.V. Early onset myasthenia gravis with atypical features. *Kathmandu Univ Med J (KUMJ)* **6**, 231-234 (2008).
2. Alaama, T., Basharat, P. & Nicolle, M.W. Unusual case of recurrent falls: myasthenia gravis in an elderly patient. *Can Fam Physician* **58**, 1231-1232 (2012).
3. Alboini, P.E., Damato, V., Iorio, R., Luigetti, M. & Evoli, A. Myasthenia gravis with presynaptic neurophysiological signs: Two case reports and literature review. *Neuromuscul Disord* **25**, 646-650 (2015).
4. Alevizos, B., Gatzonis, S. & Anagnostara, C. Myasthenia gravis disclosed by lithium carbonate. *J Neuropsychiatry Clin Neurosci* **18**, 427-429 (2006).
5. Antelli, A., *et al.* Neuromyotonia and myasthenia gravis in the absence of thymoma. *Eur J Neurol* **13**, e5 (2006).
6. Asadollahi, M., Rezaiyan, B. & Amjadi, H. A rare case of facioscapulohumeral muscular dystrophy and myasthenia gravis. *Iran J Neurol* **11**, 28-29 (2012).
7. Asghar, H., Sheikh, F.N., Dev, H., Lazarevic, M.B. & Hassan, S.A. An Atypical Presentation of Myasthenia Gravis: A Case Report. *Cureus* **11**, e4563 (2019).
8. Baker, S.K. & Tarnopolsky, M.A. Sporadic rippling muscle disease unmasked by simvastatin. *Muscle Nerve* **34**, 478-481 (2006).
9. Barnes, P.R., *et al.* Recurrent congenital arthrogryposis leading to a diagnosis of myasthenia gravis in an initially asymptomatic mother. *Neuromuscul Disord* **5**, 59-65 (1995).
10. Beauchamp, M.L., Kemink, J.L. & Donofrio, P.D. Myasthenia gravis: onset following neurotologic surgery. *Am J Otol* **7**, 302-304 (1986).
11. Behbehani, R., Sharfuddin, K. & Anim, J.T. Mitochondrial ophthalmoplegia with fatigable weakness and elevated acetylcholine receptor antibody. *J Neuroophthalmol* **27**, 41-44 (2007).
12. Benito-Leon, J., Porta-Etessam, J. & Diaz de Bustamante, A. [Myasthemia gravis and myotonic dystrophy in the same patient]. *Rev Neurol* **32**, 498 (2001).
13. Benjamin, L.A., Chipolombwe, J. & Domargard, K. Case report: A young man with progressive weakness, double vision and breathlessness. *Malawi Med J* **23**, 94-95 (2011).
14. Bettini, M., *et al.* Immune-mediated rippling muscle disease and myasthenia gravis. *J Neuroimmunol* **299**, 59-61 (2016).
15. Breker, D.A., Little, A.A. & Trobe, J.D. Autoimmune acquired rippling muscle disease and myasthenia gravis. *J Neuroophthalmol* **35**, 98-99 (2015).
16. Briani, C., Cagnin, A., Blandamura, S. & Altavilla, G. Multiple paraneoplastic diseases occurring in the same patient after thymomectomy. *J Neurooncol* **99**, 287-288 (2010).
17. Calin, C., *et al.* Cardiac involvement in myasthenia gravis--is there a specific pattern? *Rom J Intern Med* **47**, 179-189 (2009).
18. Cantagrel, S., *et al.* Akinesia, arthrogryposis, craniosynostosis: a presentation of neonatal myasthenia with fetal onset. *Am J Perinatol* **19**, 297-301 (2002).
19. Cao, L., *et al.* Adult-onset Nemaline Myopathy Coexisting With Myasthenia Gravis: A Case Report. *Medicine (Baltimore)* **95**, e2527 (2016).
20. Cao, Y., Gui, M., Ji, S. & Bu, B. Guillain-Barre syndrome associated with myasthenia gravis: Three cases report and a literature review. *Medicine (Baltimore)* **98**, e18104 (2019).
21. Carvalho, M.S., *et al.* [Myopathies associated with tubular aggregates]. *Arq Neuropsiquiatr* **51**, 363-370 (1993).

22. Casasnovas, C., Povedano, M., Jauma, S., Montero, J. & Martinez-Matos, J.A. Musk-antibody positive myasthenia gravis presenting with isolated neck extensor weakness. *Neuromuscul Disord* **17**, 544-546 (2007).
23. Chakraborty, P.P., Mandal, S.K., Chowdhury, S.R., Bandyopadhyay, D. & Bhattacharjee, R. Mitochondrial myopathy associated with myasthenia gravis in a young man. *J Clin Neurosci* **14**, 705-708 (2007).
24. Chatterjee, T., Senthil Kumaran, S. & Roy, M. A Case Report and Literature Review of New-Onset Myasthenia Gravis After COVID-19 Infection. *Cureus* **14**, e33048 (2022).
25. Chieza, J.T., Fleming, I., Parry, N. & Skelton, V.A. Maternal myasthenia gravis complicated by fetal arthrogryposis multiplex congenita. *Int J Obstet Anesth* **20**, 79-82 (2011).
26. Ciaccio, M., Parodi, A. & Rebora, A. Myasthenia gravis and lupus erythematosus. *Int J Dermatol* **28**, 317-320 (1989).
27. Cosi, V., Faggi, L. & Piccolo, G. [A case of paraneoplastic myasthenia-like syndrome: electrophysiological and clinical peculiarities]. *Riv Neurol* **47**, 148-156 (1977).
28. Cowan, J., Moenning, J.E. & Bussard, D.A. Glucocorticoid therapy for myasthenia gravis resulting in resorption of the mandibular condyles. *J Oral Maxillofac Surg* **53**, 1091-1096 (1995).
29. Cucurachi, L., Cattaneo, L., Gemignani, F. & Pavesi, G. Late onset generalized myasthenia gravis presenting with facial weakness and bulbar signs without extraocular muscle involvement. *Neurol Sci* **30**, 343-344 (2009).
30. D'Amelio, M.*, et al.* Dropped head as an unusual presenting sign of myasthenia gravis. *Neurol Sci* **28**, 104-106 (2007).
31. de Carvalho, M. & Geraldes, R. Longstanding right-hand weakness in a patient with myasthenia gravis. *Muscle Nerve* **34**, 670-671 (2006).
32. de la Espriella, J.*, et al.* [Pure cutaneous dermatomyositis and myasthenia]. *Ann Dermatol Venereol* **119**, 840-842 (1992).
33. de los Angeles Avaria, M., Kleinsteuber, K., Novoa, F., Faundez, P. & Carvallo, P. Myotonic dystrophy in a female with myasthenia gravis. *Pediatr Neurol* **36**, 421-423 (2007).
34. Deguchi, K.*, et al.* Anti-MuSK Antibody-positive Myasthenia Gravis Successfully Treated with Outpatient Periodic Weekly Blood Purification Therapy. *Intern Med* **57**, 1455-1458 (2018).
35. Devic, P., Choumert, A., Vukusic, S., Confavreux, C. & Petiot, P. Myopathic camptocormia associated with myasthenia gravis. *Clin Neurol Neurosurg* **115**, 1488-1489 (2013).
36. Diaz-Manera, J.*, et al.* Antibodies to AChR, MuSK and VGKC in a patient with myasthenia gravis and Morvan's syndrome. *Nat Clin Pract Neurol* **3**, 405-410 (2007).
37. Domingo, C.A., Landau, M.E. & Campbell, W.W. Selective Triceps Muscle Weakness in Myasthenia Gravis is Under-Recognized. *J Clin Neuromuscul Dis* **18**, 103-104 (2016).
38. Evoli, A.*, et al.* Multiple paraneoplastic diseases associated with thymoma. *Neuromuscul Disord* **9**, 601-603 (1999).
39. Fang, J.*, et al.* Myasthenia gravis coexisting with HINT1-related motor axonal neuropathy without neuromyotonia: a case report. *BMC Neurol* **22**, 168 (2022).
40. Filippelli, E., Barone, S., Granata, A., Nistico, R. & Valentino, P. A case of facioscapulohumeral muscular dystrophy and myasthenia gravis with positivity of anti-Ach receptor antibody: a fortuitous association? *Neurol Sci* **40**, 195-197 (2019).
41. Fitzgerald, M.G. & Shafritz, A.B. Distal myasthenia gravis. *J Hand Surg Am* **39**, 1419-1420 (2014).
42. Fleisher, J.*, et al.* Acquired neuromyotonia heralding recurrent thymoma in myasthenia gravis. *JAMA Neurol* **70**, 1311-1314 (2013).

43. Foster, E., McLean, C. & White, O. Glioneuronal brainstem tumor - It's all in the eyes. *J Clin Neurosci* **60**, 151-153 (2019).
44. Fricke, J., Neugebauer, A., Kirsch, A. & Russmann, W. Ocular neuromyotonia: a case report. *Strabismus* **10**, 119-124 (2002).
45. Fujita, N., *et al.* [Rippling muscle disease with myasthenia gravis]. *Rinsho Shinkeigaku* **62**, 563-566 (2022).
46. Galassi, G., Rispoli, V., Iori, E., Ariatti, A. & Marchioni, A. Coincidental Onset of Ocular Myasthenia Gravis Following ChAdOx1 n-CoV-19 Vaccine against Severe Acute Respiratory Syndrome Coronavirus 2 (SARS-CoV-2). *Isr Med Assoc J* **24**, 9-10 (2022).
47. George, J.S., Harikrishnan, S., Ali, I., Baresi, R. & Hanemann, C.O. Acquired rippling muscle disease in association with myasthenia gravis. *J Neurol Neurosurg Psychiatry* **81**, 125-126 (2010).
48. Gherarducci, D. & Giannini, A. [Clinical and therapeutic findings on 4 cases of myasthenia]. *Riv Patol Nerv Ment* **77**, 573-578 (1956).
49. Gluck, J., Rymarczyk, B., Paluch, U., Rogala, B. & Brzoza, Z. A case of Good's syndrome diagnosed after more than 20 years since onset of myasthenia in a patient with psoriasis. *Neurol Sci* **37**, 1179-1180 (2016).
50. Golden, S.K., Reiff, C.J., Painter, C.J. & Repplinger, M.D. Myasthenia Gravis Presenting as Persistent Unilateral Ptosis with Facial Droop. *J Emerg Med* **49**, e23-25 (2015).
51. Gong, P.H., *et al.* Acute severe asthma with thyroid crisis and myasthenia: a case report and literature review. *Clin Respir J* **11**, 671-676 (2017).
52. Gorthi, S.P., Shankar, S., Johri, S., Mishra, A. & Chaudhary, N.R. HIV infection with myasthenia gravis. *J Assoc Physicians India* **53**, 995-996 (2005).
53. Hara, K., *et al.* Vocal cord paralysis in myasthenia gravis with anti-MuSK antibodies. *Neurology* **68**, 621-622 (2007).
54. Haran, M., *et al.* Can a rare form of myasthenia gravis shed additional light on disease mechanisms? *Clin Neurol Neurosurg* **115**, 562-566 (2013).
55. Hayat, G.R., Kulkantrakorn, K., Campbell, W.W. & Giuliani, M.J. Neuromyotonia: autoimmune pathogenesis and response to immune modulating therapy. *J Neurol Sci* **181**, 38-43 (2000).
56. Howard, J.F., Jr., *et al.* Long-term efficacy of eculizumab in refractory generalized myasthenia gravis: responder analyses. *Ann Clin Transl Neurol* **8**, 1398-1407 (2021).
57. Huang, N.H., Lien, L.M. & Chen, W.H. Macrophage Activation Syndrome in a Case of Myasthenia Gravis with Concurrent Cytomegalovirus Infection. *Acta Neurol Taiwan* **29(4)**, 114-118 (2020).
58. Humbert, P. & Dupond, J.L. [Multiple autoimmune syndromes]. *Ann Med Interne (Paris)* **139**, 159-168 (1988).
59. Husain, F., Ryan, N.J. & Hogan, G.R. Concurrence of limb girdle muscular dystrophy and myasthenia gravis. *Arch Neurol* **46**, 101-102 (1989).
60. Ishiguro, T., *et al.* Development of myasthenia gravis 8 years after interstitial lung disease associated with antisynthetase (anti-EJ antibody) syndrome. *Clin Case Rep* **5**, 61-65 (2017).
61. Isik, K., Morkavuk, G. & Odabasi, Z. Dropped Head Syndrome As a Presenting Sign of Different Diseases: Report of Three Cases. *Noro Psikiyatr Ars* **60**, 185-187 (2023).
62. Iwase, T. & Iwase, C. Systemic effect of local and small-dose botulinum toxin injection to unmask subclinical myasthenia gravis. *Graefes Arch Clin Exp Ophthalmol* **244**, 415-416 (2006).
63. Iyah, G.S. & Misra, P. Successful outcome of peripartum cardiomyopathy in a myasthenia gravis patient. *Congest Heart Fail* **14**, 48-51 (2008).

64. Jakubikova, M., Pitha, J., Latta, J., Ehler, E. & Schutzner, J. Myasthenia gravis, Castleman disease, pemphigus, and anti-phospholipid syndrome. *Muscle Nerve* **47**, 447-451 (2013).
65. Jang, J., Chang, M. & Kyung, S. Ocular Neuromyotonia and Myasthenia Gravis. *J Pediatr Ophthalmol Strabismus* **52**, 190-191 (2015).
66. Javitt, N.B. & Daniels, R.A. Myasthenia gravis with sarcoidosis; a case report. *J Mt Sinai Hosp N Y* **26**, 177-187 (1959).
67. Ji, K.H. & Bae, J.S. CPAP therapy reverses weakness of myasthenia gravis: role of obstructive sleep apnea in paradoxical weakness of myasthenia gravis. *J Clin Sleep Med* **10**, 441-442 (2014).
68. Jokela, M., Udd, B. & Paivarinta, M. Double trouble: spinal muscular atrophy type II and seropositive myasthenia gravis in the same patient. *Neuromuscul Disord* **22**, 129-130 (2012).
69. Kapica-Topczewska, K.*, et al.* Excessive daytime sleepiness in a patient with coexisting myotonic dystrophy type 1, myasthenia gravis and Graves' disease. *Neurol Neurochir Pol* **51**, 190-193 (2017).
70. Kaplan, F. & Topal, E. Successful desensitization protocol for pyridostigmine in a 12 year old patient with myasthenia gravis. *Turk J Pediatr* **65**, 326-329 (2023).
71. Karacostas, D., Mavromatis, I., Georgakoudas, G., Artemis, N. & Milonas, I. Isolated distal hand weakness as the only presenting symptom of myasthenia gravis. *Eur J Neurol* **9**, 429-430 (2002).
72. Karam, C. & Scelsa, S.N. Clinical Reasoning: A 48-year-old woman with generalized weakness. *Neurology* **74**, e76-80 (2010).
73. Kass, R. Mindful Breathing Offers Relief for Myasthenia Gravis: A Case Report. *Adv Mind Body Med* **33**, 22-25 (2019).
74. Khan, F.G. & Namran, S. Coexistence of myasthenia gravis with hypokalemic periodic paralysis: a rare presentation. *BMJ Case Rep* **12**(2019).
75. Khartade, H.K., Meshram, V.P., Tumram, N.K., Parchake, M.B. & Pathak, H.M. Fatal Aspiration of Barium Sulfate in a Case of Myasthenia Gravis: A Case Report and Review of Literature. *J Forensic Sci* **65**, 1350-1353 (2020).
76. Kilpatrick, C., Braund, W. & Burns, R. Myopathy with myasthenia features possibly induced by codeine linctus. *Med J Aust* **2**, 410 (1982).
77. Kim, J.B. & Ballow, M. Progressive muscle weakness in a 4-year-old girl. *Ann Allergy Asthma Immunol* **92**, 19-24 (2004).
78. Kinoshita, M., Nakazato, H., Wakata, N. & Satoyoshi, E. Myasthenic neuromyopathy. An unusual neuromuscular disorder. *Eur Neurol* **21**, 52-58 (1982).
79. Kirzinger, L.*, et al.* Myopathy in Childhood Muscle-Specific Kinase Myasthenia Gravis. *Pediatr Neurol* **65**, 90-92 (2016).
80. Koge, J.*, et al.* Morvan's syndrome and myasthenia gravis related to familial Mediterranean fever gene mutations. *J Neuroinflammation* **13**, 68 (2016).
81. Kopp, C.R., Jandial, A., Mishra, K., Sandal, R. & Malhotra, P. Myasthenia gravis unmasked by imatinib. *Br J Haematol* **184**, 321 (2019).
82. Lakhal, K., Blel, Y., Fysekidis, M., Mohammedi, K. & Bouadma, L. Concurrent Graves disease thyrotoxicosis and myasthenia gravis: the treatment of the former may dangerously reveal the latter. *Anaesthesia* **63**, 876-879 (2008).
83. Lane, R.J., Roncaroli, F., Charles, P., McGonagle, D.G. & Orrell, R.W. Acetylcholine receptor antibodies in patients with genetic myopathies: clinical and biological significance. *Neuromuscul Disord* **22**, 122-128 (2012).
84. Lapeer, G.L. Myasthenia gravis: a case report. *Cranio* **3**, 392-395 (1985).

85. Le Forestier, N., Gherardi, R.K. & Meyrignac, C. [Association of myasthenia and mitochondrial ocular myopathy revealed by bilateral ptosis]. *Presse Med* **22**, 1975 (1993).
86. Le Forestier, N.*, et al.* Myasthenic symptoms in patients with mitochondrial myopathies. *Muscle Nerve* **18**, 1338-1340 (1995).
87. Matsuda, Y., Sakata, C., Sunohara, N., Nonaka, I. & Satoyoshi, E. [Two cases of mitochondrial myopathy (focal cytochrome c oxidase deficiency), long-term follow-up on a diagnosis of ocular type myasthenia gravis]. *Rinsho Shinkeigaku* **29**, 1180-1182 (1989).
88. Matsumoto, H., Sugiyama, T., Ito, M. & Yachi, A. Occurrence of myasthenia gravis in a patient with congenital myotonia. *J Neurol Sci* **57**, 83-88 (1982).
89. Maytal, J., Spiro, A.J., Sinnar, S. & Moshe, S.L. The coexistence of myasthenia gravis and myotonic dystrophy in one family. *Neuropediatrics* **18**, 8-10 (1987).
90. McGonigal, A., Thomas, A.M. & Petty, R.K. Facioscapulohumeral muscular dystrophy and myasthenia gravis co-existing in the same patient: a case report. *J Neurol* **249**, 219-220 (2002).
91. Mehta, M.P. & Sokol, L.L. The case of a 30-year-old man with subacute gait instability, weakness, and muscle spasms. *Ann Clin Transl Neurol* **7**, 2535-2537 (2020).
92. Menkes, C.J., Chouraki, L., Lemaire, V., Guiraudon, C. & Delbarre, F. [Sarcoidosis, myasthenia and chronic polyarthritis. A case]. *Ann Med Interne (Paris)* **122**, 1015-1022 (1971).
93. Miyabe, M., Dohi, S., Iwasaki, H., Omote, K. & Takahashi, T. [Anesthetic managements of a patient with myasthenia gravis and myotonia congenita]. *Masui* **31**, 650-654 (1982).
94. Morise, S.*, et al.* Thymoma-associated Progressive Encephalomyelitis with Rigidity and Myoclonus (PERM) with Myasthenia Gravis. *Intern Med* **56**, 1733-1737 (2017).
95. Muller-Felber, W.*, et al.* Immunosuppressive treatment of rippling muscles in patients with myasthenia gravis. *Neuromuscul Disord* **9**, 604-607 (1999).
96. Murotani, Y., Kuroda, Y., Goto, K., Kawai, T. & Matsuda, S. Unexpected dislocation following accurate total hip arthroplasty caused by excessive hip joint laxity during myasthenic crisis: a case report. *J Med Case Rep* **12**, 331 (2018).
97. Natera-de Benito, D.*, et al.* KLHL40-related nemaline myopathy with a sustained, positive response to treatment with acetylcholinesterase inhibitors. *J Neurol* **263**, 517-523 (2016).
98. Nedelcu, L. & Dumitrescu, T. Gastrointestinal stromal tumor associated with myasthenia gravis: a case report. *Iran Red Crescent Med J* **19**, e58682 (2017).
99. Ngeh, J.K. & McElligott, G. Myasthenia gravis: an elusive diagnosis in older people. *J Am Geriatr Soc* **49**, 683-684 (2001).
100. Niks, E.H.*, et al.* Pre- and postsynaptic neuromuscular junction abnormalities in musk myasthenia. *Muscle Nerve* **42**, 283-288 (2010).
101. Nilsson, K. Septicaemia with Rickettsia helvetica in a patient with acute febrile illness, rash and myasthenia. *J Infect* **58**, 79-82 (2009).
102. O'Sullivan, S.S., Mullins, G.M., Neligan, A., McNamara, B. & Galvin, R.J. Acquired generalised neuromyotonia, cutaneous lupus erythematosus and alopecia areata in a patient with myasthenia gravis. *Clin Neurol Neurosurg* **109**, 374-375 (2007).
103. Ohnari, K., Okada, K., Higuchi, O., Matsuo, H. & Adachi, H. Late-onset Myasthenia Gravis Accompanied by Amyotrophic Lateral Sclerosis with Antibodies against the Acetylcholine Receptor and Low-density Lipoprotein Receptor-related Protein 4. *Intern Med* **57**, 3021-3024 (2018).
104. Okuyama, Y., Mizuno, T., Inoue, H. & Kimoto, K. Amyotrophic lateral sclerosis with anti-acetylcholine receptor antibody. *Intern Med* **36**, 312-315 (1997).
105. Oliveira, F., Schoeps, V., Sanvito, W. & Valerio, B. Gluten and Neuroimmunology. Rare association with Myasthenia Gravis and Literature Review. *Rev Assoc Med Bras (1992)* **64**, 311-314 (2018).

106. Oskarsson, B. & Ringel, S.P. Oculopharyngeal muscular dystrophy as a cause of progression of weakness in antibody positive myasthenia gravis. *Neuromuscul Disord* **23**, 316-318 (2013).
107. Panegyres, P.K., Squier, M., Mills, K.R. & Newsom-Davis, J. Acute myopathy associated with large parenteral dose of corticosteroid in myasthenia gravis. *J Neurol Neurosurg Psychiatry* **56**, 702-704 (1993).
108. Park, D.B., Dobson, J.V. & Losek, J.D. All that wheezes is not asthma: cognitive bias in pediatric emergency medical decision making. *Pediatr Emerg Care* **30**, 104-107 (2014).
109. Paul, B.S., Singh, G., Bansal, R.K. & Singla, M. Isaac's syndrome associated with myasthenia gravis and thymoma. *Indian J Med Sci* **64**, 320-324 (2010).
110. Peltier, A.C.*, et al.* Coexistent autoimmune autonomic ganglionopathy and myasthenia gravis associated with non-small-cell lung cancer. *Muscle Nerve* **41**, 416-419 (2010).
111. Pissarra, F.*, et al.* [Muscular diseases in hyperthyroidism]. *Acta Med Port* **8**, 501-504 (1995).
112. Plewnia, K.*, et al.* A rare association of myasthenia gravis and mitochondrial myopathy: a clinical, biochemical and morphologic study of one case. *J Submicrosc Cytol Pathol* **29**, 335-338 (1997).
113. Punga, A.R., Nygren, I., Askmark, H. & Stalberg, E.V. Monozygous twins with neuromuscular transmission defects at opposite sides of the motor endplate. *Acta Neurol Scand* **119**, 207-211 (2009).
114. Ramakrishnan, P. & Siddiqui, S. Myasthenia Gravis as a Cause of Failed Extubation Diagnosed by Diaphragmatic Ultrasound. *J Coll Physicians Surg Pak* **29**, 474-475 (2019).
115. Rehman, H.U. A 90 year old man with difficulty swallowing and proximal muscle weakness. *BMJ* **344**, e461 (2012).
116. Renard, D., Castelnovo, G. & Labauge, P. Distal myasthenia gravis. *Acta Neurol Belg* **108**, 107-108 (2008).
117. Rigamonti, A., Lauria, G., Piamarta, F., Fiumani, A. & Agostoni, E. Thymoma-associated myasthenia gravis without acetylcholine receptor antibodies. *J Neurol Sci* **302**, 112-113 (2011).
118. Robinson, R.O. & Hodgson, S. Congenital dystrophy and myasthenia gravis. *Neuropediatrics* **19**, 168 (1988).
119. Rodolico, C.*, et al.* Limb-girdle myasthenia: clinical, electrophysiological and morphological features in familial and autoimmune cases. *Neuromuscul Disord* **12**, 964-969 (2002).
120. Rodriguez Cruz, P.M.*, et al.* Congenital myopathies with secondary neuromuscular transmission defects; a case report and review of the literature. *Neuromuscul Disord* **24**, 1103-1110 (2014).
121. Rohde, D., Sliwka, U., Schweizer, K. & Jakse, G. Oculo-bulbar myasthenia gravis induced by cytokine treatment of a patient with metastasized renal cell carcinoma. *Eur J Clin Pharmacol* **50**, 471-473 (1996).
122. Romi, F.*, et al.* Thymectomy and antimuscle antibodies in nonthymomatous myasthenia gravis. *Ann N Y Acad Sci* **998**, 481-490 (2003).
123. Rubin, D.I. & Litchy, W.J. Severe, focal tibialis anterior and triceps brachii weakness in myasthenia gravis: a case report. *J Clin Neuromuscul Dis* **12**, 219-222 (2011).
124. Ruiz Torregrosa, P., Garcia Sevila, R. & Gaya Garcia-Manso, I. Myasthenia gravis and antisynthetase syndrome, an infrequent association. *Med Clin (Barc)* **157**, 38-39 (2021).
125. Rycroft, R.J., Valdimarsson, H., Bannister, L.H. & Wells, R.S. Chronic muco-cutaneous candidiasis of late onset, thymoma and myopathy. A report of four cases. *Clin Exp Dermatol* **1**, 59-74 (1976).

126. Sakuma, H., Shimazaki, S., Saito, H. & Ohuchi, M. [A patient with facioscapulohumeral muscular dystrophy accompanied by myasthenia gravis]. *Rinsho Shinkeigaku* **41**, 179-183 (2001).
127. Sanadze, A.G., Sidnev, D.V., Kasatkina, L.F., Dedaev, S.I. & Karganov, M. [Neuromyotonia and myasthenia in a patient with thymoma]. *Zh Nevrol Psikhiatr Im S S Korsakova* **110**, 101-103 (2010).
128. Sanford, D., MacDonald, M., Nicolle, M. & Xenocostas, A. Development of Myasthenia Gravis in a Patient with Chronic Myeloid Leukemia during Treatment with Nilotinib. *Hematol Rep* **6**, 5288 (2014).
129. Sansone, V., Saperstein, D.S., Barohn, R.J. & Meola, G. Concurrence of facioscapulohumeral muscular dystrophy and myasthenia gravis. *Muscle Nerve* **30**, 679-680 (2004).
130. Satoh, A., Tsujihata, M., Yoshimura, T., Mori, M. & Nagataki, S. Myasthenia gravis associated with Satoyoshi syndrome: muscle cramps, alopecia, and diarrhea. *Neurology* **33**, 1209-1211 (1983).
131. Sawa, N., Kataoka, H., Eura, N. & Ueno, S. Dropped head with positive intravenous edrophonium, progressing to myasthenia gravis. *BMJ Case Rep* **2013**(2013).
132. Sawanyawisuth, K., Tiamkao, S. & Pratipanawatr, T. Myasthenia gravis accompanied with hypokalemic periodic paralysis. *J Med Assoc Thai* **89**, 727-729 (2006).
133. Scheer, B.V., Valero-Burgos, E. & Costa, R. Myasthenia gravis and endurance exercise. *Am J Phys Med Rehabil* **91**, 725-727 (2012).
134. Schoser, B. Self-diagnosis of a triple trouble. *Neuromuscul Disord* **28**, 825-827 (2018).
135. Schoser, B., *et al.* Immune-mediated rippling muscle disease with myasthenia gravis: a report of seven patients with long-term follow-up in two. *Neuromuscul Disord* **19**, 223-228 (2009).
136. Schrijver-Levie, N.S., Boersma, B. & ten Houten, R. [Two children seriously weakened by myasthenia]. *Ned Tijdschr Geneeskd* **148**, 674-677 (2004).
137. Scola, R.H., *et al.* [Distal myasthenia gravis: case report]. *Arq Neuropsiquiatr* **61**, 119-120 (2003).
138. Selcen, D., *et al.* Impaired Synaptic Development, Maintenance, and Neuromuscular Transmission in LRP4-Related Myasthenia. *JAMA Neurol* **72**, 889-896 (2015).
139. Shah, L.K., *et al.* COVID-19 and Sepsis in an Atypical Case of Mixed Connective Tissue Disorder Presenting With a Myasthenic Crisis. *Cureus* **14**, e29092 (2022).
140. Singh, H., *et al.* Anaesthesia for a patient with Isaac's syndrome and myasthenia gravis. *Br J Anaesth* **103**, 460-461 (2009).
141. Singh, R. & Pentland, B. Myasthenia gravis masquerading as post-poliomyelitis syndrome. *J Rehabil Med* **38**, 136-137 (2006).
142. Sondes, S., Kennedy, J.M., Kishner, S., Lopez, F.A. & Anthony, L. Clinical case of the month. Severe progressive weakness in a 58-year-old man. Myasthenia gravis. *J La State Med Soc* **154**, 292-295; quiz 295 (2002).
143. Sousa, D.C., *et al.* Ophthalmoparesis and unilateral finger flexor muscle weakness in seronegative myasthenia gravis. *Can J Ophthalmol* **52**, e213-e216 (2017).
144. Spengos, K., Vassilopoulou, S., Christou, Y. & Manta, P. Sarcoidosis in a case of MuSK-positive myasthenia gravis. *Neuromuscul Disord* **18**, 890-891 (2008).
145. Spengos, K., *et al.* Dropped head syndrome as prominent clinical feature in MuSK-positive Myasthenia Gravis with thymus hyperplasia. *Neuromuscul Disord* **18**, 175-177 (2008).
146. Stortebecker, T.P. Signs of myositis in myasthenia gravis and in myopathy clinically resembling progressive muscular dystrophy. *Acta Med Scand* **151**, 452-464 (1955).
147. Strober, J., Cowan, M.J. & Horn, B.N. Allogeneic hematopoietic cell transplantation for refractory myasthenia gravis. *Arch Neurol* **66**, 659-661 (2009).

148. Sundar, U., *et al.* Experience with patients with anti-MUSK antibody positive myasthenia gravis. *J Assoc Physicians India* **58**, 640-642 (2010).
149. Takeda, T., *et al.* Acquired hemophilia A associated with myasthenia gravis and Isaacs' syndrome. *J Neurol Sci* **369**, 210-211 (2016).
150. Tanwani, L.K., Lohano, V., Ewart, R., Broadstone, V.L. & Mokshagundam, S.P. Myasthenia gravis in conjunction with Graves' disease: a diagnostic challenge. *Endocr Pract* **7**, 275-278 (2001).
151. Thilak, M.R., Prabhu, A.N., Rao, K.S. & Vasantbhai, M.J. Isolated bilateral triceps muscle weakness as a presenting complaint in myasthenia gravis: A review. *Neurol India* **67**, 566-568 (2019).
152. Tojo, M., Sakuragawa, N., Nonaka, I. & Sato, M. [An atypical form of juvenile myasthenia gravis associated with severe emaciation, muscle atrophy, ophthalmoplegia, bulbar signs and joint contracture]. *No To Hattatsu* **19**, 402-407 (1987).
153. Tsironis, T. & Catania, S. Reversible spontaneous EMG activity during myasthenic crisis: Two case reports. *eNeurologicalSci* **14**, 16-18 (2019).
154. Turker, H., *et al.* Hypothyroid myopathy with manifestations of Hoffman's syndrome and myasthenia gravis. *Thyroid* **18**, 259-262 (2008).
155. Uludag, I.F., Korucuk, M., Sener, U. & Zorlu, Y. Myasthenia gravis as a cause of head drop in Parkinson disease. *Neurologist* **17**, 144-146 (2011).
156. Vallet, B., Fourrier, F., Hurtevent, J.F., Parent, M. & Chopin, C. Myasthenia gravis and steroid-induced myopathy of the respiratory muscles. *Intensive Care Med* **18**, 424-426 (1992).
157. Van Parijs, V., Van den Bergh, P.Y. & Vincent, A. Neuromyotonia and myasthenia gravis without thymoma. *J Neurol Neurosurg Psychiatry* **73**, 344-345 (2002).
158. van Schaik, S.M., Kwa, V.I. & van der Kooi, A.J. Acquired rippling muscle disease associated with mild myasthenia gravis: a case report. *J Neurol* **256**, 1187-1188 (2009).
159. Vats, H.S., Richardson, S.K., Pulukurthy, S. & Olshansky, B. Pericarditis in myasthenia gravis. *Cardiol Rev* **12**, 134-137 (2004).
160. Verspyck, E., *et al.* Myasthenia gravis with polyhydramnios in the fetus of an asymptomatic mother. *Prenat Diagn* **13**, 539-542 (1993).
161. Vinagre, F., Santos, M.J. & da Silva, J.C. [Systemic lupus erythematosus and weakness]. *Acta Reumatol Port* **31**, 167-172 (2006).
162. Wakayama, Y., Ohbu, S. & Machida, H. Myasthenia gravis, muscle twitch, hyperhidrosis and limb pain associated with thymoma: proposal of possible new myasthenic syndrome. *Tohoku J Exp Med* **164**, 285-291 (1991).
163. Werneck, L.C., Bittencourt, P.C. & Novak, E.M. [Myasthenia gravis with the electrographic response of a myasthenic syndrome. Report of a case]. *Arq Neuropsiquiatr* **43**, 198-205 (1985).
164. Wielosz, E., Majdan, M., Jeleniewicz, R. & Mazurek, M. [Autoimmune diseases with the presence of anti-ku antibodies - analysis of three cases]. *Wiad Lek* **69**, 24-26 (2016).
165. Yaguchi, H., *et al.* Dropped head sign as the only symptom of myasthenia gravis. *Intern Med* **46**, 743-745 (2007).
166. Yamashita, S., *et al.* Myasthenia gravis complicated with primary aldosteronism and hypokalemic myopathy. *Intern Med* **48**, 1465-1469 (2009).
167. Yaskin, J.C., Hawthorne, H.R., Frobes, A.S. & Leopold, R.L. Thymectomy in treatment of myasthenia gravis: report of 3 cases. *Arch Neurol Psychiatry* **64**, 292-294 (1950).
168. Yasuda, M., *et al.* Mixed connective tissue disease presenting myasthenia gravis. *Intern Med* **32**, 633-637 (1993).

169. Zhang, J., Niu, S., Wang, Y. & Hu, W. Myasthenia gravis and Guillain-Barre cooccurrence syndrome. *Am J Emerg Med* **31**, 1264-1267 (2013).
170. Zouvelou, V., Stamboulis, E., Skriapa, L. & Tzartos, S.J. MuSK-Ab positive myasthenia: not always grave. *J Neurol Sci* **331**, 150-151 (2013).

• *Lack of definitive MG diagnosis (32 of these highlighted in yellow were included in a separate analysis due to presence of TET and IIM)*:

1. Aksu, A., Sin, C. & Yilmaz, B. Adult Tiger Man: A Case of Dermatomyositis Associated With Thymoma. *Clin Nucl Med* **47**, e448-e449 (2022).
2. Aquilina, N., Bugeja, V. & Zahra, C. A Case of Orbital Myositis Presenting With Dizziness. *Open Access Maced J Med Sci* **6**, 1278-1281 (2018).
3. Banwell, B.L., *et al.* Myopathy, myasthenic syndrome, and epidermolysis bullosa simplex due to plectin deficiency. *J Neuropathol Exp Neurol* **58**, 832-846 (1999).
4. Barre, M., Delaporte, E., Berbis, P. & Benzaquen, M. Severe dermatomyositis revealing a thymic carcinoma: Did rituximab delay the diagnosis? *Dermatol Ther* **33**, e14016 (2020).
5. Benedek, L. Pseudomyasthenisches syndrom bei polymyositis interstitialis chronica fibrosa. *Monatsschrift für Psychiatrie und Neurologie* **109**, 93-99 (1944).
6. Chen, B.H., Zhu, X.M., Xie, L. & Hu, H.Q. Immune-mediated necrotizing myopathy: Report of two cases. *World J Clin Cases* **11**, 3552-3559 (2023).
7. Cranney, A., Markman, S., Lach, B. & Karsh, J. Polymyositis in a patient with thymoma and T cell lymphocytosis. *J Rheumatol* **24**, 1413-1416 (1997).
8. D'Agostino, S., Avella, A. & Maddaluno, R. [Considerations on a case of the Fiedler type of acute interstitial myocarditis and of granulomatous myositis associated with a thvmic tumor]. *Rass Clin Ter* **61**, 118-126 (1962).
9. Dell'Amore, A., *et al.* Paraneoplastic dermatomyositis as presentation of thymic carcinoma. *Gen Thorac Cardiovasc Surg* **61**, 422-425 (2013).
10. Descamps, L. [Familial oculo-bulbo-facial myopathy with periodic myasthenia-like attacks]. *Acta Neurol Psychiatr Belg* **55**, 351-355 (1955).
11. Du, X., *et al.* Case Report: MDM4 Amplified in a Thymoma Patient With Autoimmune Enteropathy and Myocarditis. *Front Endocrinol (Lausanne)* **12**, 661316 (2021).
12. Du, Y.J., Liu, W.C., Chen, X. & Cheng, Y.J. [A case report of colchicine-induced myopathy in a patient with chronic kidney disease]. *Beijing Da Xue Xue Bao Yi Xue Ban* **53**, 1188-1190 (2021).
13. Fong, P.H., Wee, A., Chan, H.L. & Tan, Y.O. Primary thymic carcinoma and its association with dermatomyositis and pure red cell aplasia. *Int J Dermatol* **31**, 426-428 (1992).
14. Frith, J., Toller-Artis, E., Tcheurekdjian, H. & Hostoffer, R. Good syndrome and polymyositis. *Ann Allergy Asthma Immunol* **112**, 478 (2014).
15. Fu, Z., Chen, G., Chen, X. & Li, Q. 18F-FDG PET/CT in a Patient With Thymoma-Associated Paraneoplastic Polymyositis. *Clin Nucl Med* **45**, 148-150 (2020).
16. Funkhouser, J.W. Thymoma associated with myocarditis and the L.E.-cell phenomenon. Report of a case. *N Engl J Med* **264**, 34-36 (1961).
17. Gdynia, H.J., *et al.* [Diagnosis and differential diagnosis of granulomatous myositis]. *Nervenarzt* **79**, 470-474 (2008).
18. Giordano, A.S. & Haymond, J.L. Myasthenia gravis: a report of two cases with necropsy findings. *Am J Clin Pathol* **14**, 253-265 (1944).
19. Glennon, P.E., Petersen, M.E. & Sheppard, M.N. Fatal giant cell myocarditis after resection of thymoma. *Heart* **75**, 531-532 (1996).
20. Haen, S.P., *et al.* Choroidal metastases from thymic carcinoma during pregnancy: Case Report. *BMC Cancer* **15**, 972 (2015).

21. Iacovelli, R., *et al.* Dermatomyositis as first clinical appearance for a thymic epidermoid cell carcinoma. *Acta Biomed* **81**, 68-71 (2010).
22. Inoue, Y., True, L.D. & Martins, R.G. Thymic carcinoma associated with paraneoplastic polymyositis. *J Clin Oncol* **27**, e33-34 (2009).
23. Isobe, K., *et al.* [An autopsied case of giant cell myocarditis and myositis associated with invasive thymoma]. *Nihon Kokyuki Gakkai Zasshi* **48**, 432-438 (2010).
24. Karino, K., *et al.* Anti-TIF1gamma antibody predicted malignancy of thymic tumor with dermatomyositis as an "autoimmune tumor marker": A case report. *Medicine (Baltimore)* **97**, e13563 (2018).
25. Karippacheril, J.G., Shetty, R., Sagar, S.C. & Kamath, S.G. Myocarditis after thymoma resection, with left ventricular hypokinesia mimicking acute coronary syndrome. *J Anesth* **27**, 805-806 (2013).
26. Kilgallen, C.M., Jackson, E., Bankoff, M., Salomon, R.N. & Surks, H.K. A case of giant cell myocarditis and malignant thymoma: a postmortem diagnosis by needle biopsy. *Clin Cardiol* **21**, 48-51 (1998).
27. Langston, J.D., Wagman, G.F. & Dickenman, R.C. Granulomatous myocarditis and myositis associated with thymoma. *Arch Pathol* **68**, 367-373 (1959).
28. Le Marc'hadour, F., Martins Ramos, J., Pasquier, B., Pasquier, D. & Couderc, P. [Association of thymus carcinoma, Hashimoto's thyroiditis and polymyositis. Anatomoclinical case with autopsy findings]. *Ann Pathol* **9**, 355-359 (1989).
29. Lee, M., Kwon, G.Y., Kim, J.S. & Jeon, E.S. Giant cell myocarditis associated with Coxsackievirus infection. *J Am Coll Cardiol* **56**, e19 (2010).
30. Masuda, M., *et al.* [A case of dermatomyositis associated with thymic abnormalities]. *Ryumachi* **32**, 140-144; discussion 144-146 (1992).
31. Metthews, W.B. & Burne, J.C. The neurological aspects of dermatomyositis. *J Neurol Neurosurg Psychiatry* **16**, 49-55 (1953).
32. Meyer, T., Grumbach, I.M., Kreuzer, H. & Morguet, A.J. Giant cell myocarditis due to coxsackie B2 virus infection. *Cardiology* **88**, 296-299 (1997).
33. Mirg, S., Das, A., Pandit, A.K., Sharma, M.C. & Srivastava, A.K. Shrinking lung syndrome mimicking diaphragmatic palsy in systemic lupus erythematosus. *Pract Neurol* **24**, 313-315 (2024).
34. Morrissey, R.P., *et al.* Case of fulminant giant-cell myocarditis associated with polymyositis, treated with a biventricular assist device and subsequent heart transplantation. *Heart Lung* **40**, 340-345 (2011).
35. Munoz Malaga, A., Bautista Lorite, J., Lopez Dominguez, J.M. & Martinez Navarro, M.L. [Thymoma and muscular involvement. Differential diagnosis between polymyositis and myasthenia gravis]. *Med Clin (Barc)* **100**, 397 (1993).
36. Niakan, E., Bertorini, T.E., Acchiardo, S.R. & Werner, M.F. Procainamide-induced myasthenia-like weakness in a patient with peripheral neuropathy. *Arch Neurol* **38**, 378-379 (1981).
37. Pascuzzi, L., Dias, J.C., Cavalieri, M.J., Gagioti, S.M. & Melaragno Filho, R. [Ocular myopathy, Kiloh-Nevin type; study of a case with histochemical and ultrastructural changes]. *Arq Neuropsiquiatr* **37**, 424-434 (1979).
38. Pasetti Bombardella, M. & Boccato, P. [Histochemical and pathogenetic considerations on a case of thymoma without associated myasthenia]. *Riv Anat Patol Oncol* **18**, 533-548 (1960).
39. Pentz, W.H. Advanced heart block as a manifestation of a paraneoplastic syndrome from malignant thymoma. *Chest* **116**, 1135-1136 (1999).
40. Peppard, R., Byrne, E. & Dennett, X. Myopathy with fatiguability--myositis or myasthenia? *Clin Exp Neurol* **22**, 47-52 (1986).

41. Pyun, K.S., Kim, Y.H., Katzenstein, R.E. & Kikkawa, Y. Giant cell myocarditis. Light and electron microscopic study. *Arch Pathol* **90**, 181-188 (1970).
42. Rini, B.I. & Gajewski, T.F. Polymyositis with respiratory muscle weakness requiring mechanical ventilation in a patient with metastatic thymoma treated with octreotide. *Ann Oncol* **10**, 973-979 (1999).
43. Rowland, L.P., Lisak, R.P., Schotland, D.L., DeJesus, P.V. & Berg, P. Myasthenic myopathy and thymoma. *Neurology* **23**, 282-288 (1973).
44. Rundle, L.G. & Sparks, F.P. Thymoma and dermatomyositis. A disease entity. *Arch Pathol* **75**, 276-283 (1963).
45. Sakuma, H., Yoshida, H., Kasukawa, R., Satoh, N. & Yoshino, K. An autopsy case with Good's syndrome and dermatomyositis. *Clin Rheumatol* **4**, 196-201 (1985).
46. Souadjian, J.V., Howell, L.P. & Lambert, E.H. Thymoma with myopathy. Report of a case. *Minn Med* **52**, 595-596 (1969).
47. Sun, H.*, et al.* Case report: A patient with brachio-cervical inflammatory myopathy was misdiagnosed as flail arm syndrome. *Front Immunol* **15**, 1378130 (2024).
48. Svahn, J.*, et al.* Immune-Mediated Rippling Muscle Disease Associated With Thymoma and Anti-MURC/Cavin-4 Autoantibodies. *Neurol Neuroimmunol Neuroinflamm* **10**(2023).
49. Takahashi, F.*, et al.* Successful resection of dermatomyositis associated with thymic carcinoma: report of a case. *Surg Today* **38**, 245-248 (2008).
50. Waller, J.V., Shapiro, M. & Paltauf, R. Congestive heart failure in postmenopausal muscular dystrophy: myositis, myocarditis, thymoma. *Am Heart J* **53**, 479-484 (1957).
51. Yang, X.*, et al.* Successful treatment of thymic carcinoma with dermatomyositis and interstitial pneumonia: A case report. *Thorac Cancer* **10**, 2031-2034 (2019).

• *Lack of both definitive MG diagnosis and evidence for at least possible myositis*:

1. Agarwal, S., Lotze, T.E. & Woodbury, S.L. A 7-year-old child with chronic droopy eyes, weakness in head-neck control, and an abnormal gait. *Semin Pediatr Neurol* **21**, 111-113 (2014).
2. Alexakou, Z.*, et al.* Thymic Carcinoma With Multiple Paraneoplastic Disorders. *Am J Med Sci* **362**, 324-330 (2021).
3. Argente-Escrig, H.*, et al.* Plectin-related scapuloperoneal myopathy with treatment-responsive myasthenic syndrome. *Neuropathol Appl Neurobiol* **47**, 352-356 (2021).
4. Bhoopalan, S.V. & Jain, R. Case 2: Hypotonia and Muscle Weakness since Birth in a 2-year-old Boy. *Pediatr Rev* **38**, 531 (2017).
5. Boyle, K., McNaughten, B., Thompson, A. & Mullen, S. Child with acute weakness: don't forget the salts. *Arch Dis Child Educ Pract Ed* **107**, 21-23 (2022).
6. Chen, T. Clinical Reasoning: A 68-Year-Old Man With Proximal Weakness and Seizures. *Neurology* **97**, e423-e428 (2021).
7. Fan, Q., Gwathmey, K., Du, X., Seth, A. & Corse, A. Tubular aggregate myopathy causing progressive fatiguable weakness. *Pract Neurol* **24**, 137-140 (2024).
8. Fitzsimons, R.B., Gurwin, E.B. & Bird, A.C. Retinal vascular abnormalities in facioscapulohumeral muscular dystrophy. A general association with genetic and therapeutic implications. *Brain* **110 ( Pt 3)**, 631-648 (1987).
9. Harrison, P., Barton, J. & Winkel, A. Chronic mimics of myasthenia gravis: a retrospective case series. *Neuromuscul Disord* **33**, 250-256 (2023).
10. Jennekens, F.G.*, et al.* Deficiency of acetylcholine receptors in a case of end-plate acetylcholinesterase deficiency: a histochemical investigation. *Muscle Nerve* **15**, 63-72 (1992).

11. Lee, M.S., Kosmorsky, G.S., Cook, J.R., Barton, J.J. & Briemberg, H.R. My, what asthenia you have. *Surv Ophthalmol* **53**, 506-511 (2008).
12. Liewluck, T., Shen, X.M., Milone, M. & Engel, A.G. Endplate structure and parameters of neuromuscular transmission in sporadic centronuclear myopathy associated with myasthenia. *Neuromuscul Disord* **21**, 387-395 (2011).
13. Lin, N., Wang, J., Liao, W. & Wen, Y. Hypothyroid myopathy with periodic paralysis as the main symptom: a case report and literature review. *Ann Palliat Med* **9**, 3698-3704 (2020).
14. Liu, L.*, et al.* Case Report: Evidences of myasthenia and cerebellar atrophy in a chinese patient with novel compound heterozygous MSTO1 variants. *Front Genet* **13**, 947886 (2022).
15. Mahfoudhi, M. & Khamassi, K. [Amyopathic dermatomyositis revealing a thymic carcinoma]. *Pan Afr Med J* **21**, 292 (2015).
16. Manjunathan, S., Singh, K. & Saini, L. Multiple Pterygium Syndrome (Escobar Syndrome): A Rare Form of Prenatal Myasthenia Presenting With Arthrogryposis Multiplex Congenita. *Neurology* **103**, e209602 (2024).
17. Montes, L.F., Ceballos, R., Cooper, M.D., Bradley, M.N. & Bockman, D.E. Chronic mucocutaneous candidiasis, myositis, and thymoma. A new triad. *JAMA* **222**, 1619-1623 (1972).
18. Morgan-Hughes, J.A., Lecky, B.R., Landon, D.N. & Murray, N.M. Alterations in the number and affinity of junctional acetylcholine receptors in a myopathy with tubular aggregates. A newly recognized receptor defect. *Brain* **104**, 279-295 (1981).
19. Norris, F.H., Jr. & Panner, B.J. Hypothyroid myopathy. Clinical, electromyographical, and ultrastructural observations. *Arch Neurol* **14**, 574-589 (1966).
20. Poh, M.*, et al.* Postexercise reflex facilitation in Lambert-Eaton myasthenic syndrome. *Pract Neurol* **24**, 338-341 (2024).
21. Polavarapu, K.*, et al.* Partial loss of desmin expression due to a leaky splice site variant in the human DES gene is associated with neuromuscular transmission defects. *Neuromuscul Disord* **39**, 10-18 (2024).
22. Popova, L.M. & Moiseev, A.N. Mechanics of breathing during prolonged artificial ventilation. *Resuscitation* **8**, 29-41 (1980).
23. Roy, B. & Chahin, N. Clinical Reasoning: A 14-year-old boy with fatigue and episodic worsening of weakness. *Neurology* **88**, e96-e100 (2017).
24. Selcen, D.*, et al.* DPAGT1 myasthenia and myopathy: genetic, phenotypic, and expression studies. *Neurology* **82**, 1822-1830 (2014).
25. Stojic, A.S., Mekhail, T. & Tsao, B.E. Leg weakness in a 66-year-old woman: a common presentation of an uncommon disease. *Cleve Clin J Med* **74**, 23-26, 29-34 (2007).
26. Suarez, G.A. & Kelly, J.J., Jr. The dropped head syndrome. *Neurology* **42**, 1625-1627 (1992).
27. Torabi, T., Huttner, A., Nowak, R.J. & Roy, B. Clinical Reasoning: Progressive proximal weakness in a 56-year-old man with bone pain. *Neurology* **93**, 939-944 (2019).
28. Tsuchiya, T., Sano, A. & Kawashima, M. Paraneoplastic Dermatomyositis as a Potential Precursor to Thymic Carcinoma. *Ann Thorac Surg* **109**, e247-e249 (2020).
29. Verdelho, A., Bentes, C. & de Carvalho, M. [Two myopathy cases]. *Rev Neurol* **28**, 1059-1061 (1999).
30. Wang, Y., Wang, C.H. & Chen, H.C. Inflammatory Pseudotumour of Temporal Bone in a Multiple-Comorbidity Patient. *J Coll Physicians Surg Pak* **32**, S168-S170 (2022).

• *Insufficient diagnostic details for MG or myositis*:

1. Argiriou, M.*, et al.* Left ventricular rupture after double valve replacement in a patient with myocarditis due to myasthenia gravis : case report. *Cardiovasc J Afr* **24**, e5-7 (2013).

2. Fayssoil, A., *et al.* Percutaneous extracorporeal membrane oxygenation for cardiogenic shock due to acute fulminant myocarditis. *Ann Thorac Surg* **89**, 614-616 (2010).
3. HooKim, K., *et al.* IgG anti-cardiomyocyte antibodies in giant cell myocarditis. *Ann Clin Lab Sci* **38**, 83-87 (2008).
4. Joudinaud, T.M., *et al.* Fatal giant cell myocarditis after thymoma resection in myasthenia gravis. *J Thorac Cardiovasc Surg* **131**, 494-495 (2006).
5. Mendelow, H. & Genkins, G. Studies in myasthenia gravis: cardiac and associated pathology. *J Mt Sinai Hosp N Y* **21**, 218-225 (1954).
6. Mygland, A., Gilhus, N.E., Hofstad, H., Aarli, J.A. & Thunold, S. [Heart disease in myasthenia gravis]. *Tidsskr Nor Laegeforen* **110**, 1209-1211 (1990).
7. Saito, N., *et al.* A survival case of invasive thymoma accompanied by acute fulminant myocarditis. *Respirol Case Rep* **1**, 36-38 (2013).
8. Spalek, P., Schnorrer, M. & Cibulcik, F. [Simultaneous occurrence of acute myasthenia gravis and acute polymyositis in 3 patients and in 2 patients also associated with a thymoma]. *Rozhl Chir* **79**, 468-470 (2000).
9. van Haelst, P.L., Brugemann, J., Diercks, G.F., Suurmeijer, A. & van Veldhuisen, D.J. Serial right ventricular endomyocardial biopsy in rapid-onset severe heart failure due to giant cell myocarditis. *Cardiovasc Pathol* **15**, 228-230 (2006).
10. Yu, H., Ma, X., Tong, N., Zhou, Z. & Zhang, Y. Acute exudative paraneoplastic polymorphous vitelliform maculopathy in a patient with thymoma, myasthenia gravis, and polymyositis. *Eur J Ophthalmol* **32**, NP56-NP61 (2022).

• *Lack of case-level detail, diagnoses reported for separate cases within the same paper, duplicate studies*:

1. Adams, C., August, C.S., Maguire, H. & Sladky, J.T. Neuromuscular complications of bone marrow transplantation. *Pediatr Neurol* **12**, 58-61 (1995).
2. Albashir, S., Olansky, L. & Sasidhar, M. Progressive muscle weakness: More there than meets the eye. *Cleve Clin J Med* **78**, 385-391 (2011).
3. Argiriou, M., *et al.* Left ventricular rupture after double valve replacement in a patient with myocarditis due to myasthenia gravis. *Cardiovasc J Afr* **24**, e1-3 (2013).
4. Bhatt, J.R. & Pascuzzi, R.M. Neuromuscular disorders in clinical practice: case studies. *Neurol Clin* **24**, 233-265 (2006).
5. Cartwright, M.S., Jeffery, D.R., Nuss, G.R. & Donofrio, P.D. Statin-associated exacerbation of myasthenia gravis. *Neurology* **63**, 2188 (2004).
6. Engel, W.K., Lichter, A.S. & Dalakas, M.C. Splenic and total-body irradiation treatment of myasthenia gravis. *Ann N Y Acad Sci* **377**, 744-754 (1981).
7. Gekht, B.M., Gustainis, V.V., Kasatkina, L.F., Nikitin, S.S. & Nozdracheva, L.V. [Electromyographic and morphological analysis of the changes in the motor units in myasthenia, combined myasthenia with thymoma and polymyositis and terminal polyneuropathy with myasthenic syndrome]. *Zh Nevropatol Psikhiatr Im S S Korsakova* **81**, 1624-1632 (1981).
8. Ibi, T. & Sahashi, K. [Myasthenia gravis, giant cell polymyositis and cardiomyositis associated with thymoma]. *Ryoikibetsu Shokogun Shirizu*, 344-346 (2001).
9. Klehmet, J., Dudenhausen, J. & Meisel, A. [Course and treatment of myasthenia gravis during pregnancy]. *Nervenarzt* **81**, 956-962 (2010).
10. LoVecchio, F. & Jacobson, S. Approach to generalized weakness and peripheral neuromuscular disease. *Emerg Med Clin North Am* **15**, 605-623 (1997).

11. Mowzoon, N., Sussman, A. & Bradley, W.G. Mycophenolate (CellCept) treatment of myasthenia gravis, chronic inflammatory polyneuropathy and inclusion body myositis. *J Neurol Sci* **185**, 119-122 (2001).
12. Padua, L., *et al.* SFEMG in ocular myasthenia gravis diagnosis. *Clin Neurophysiol* **111**, 1203-1207 (2000).
13. Petiot, P., *et al.* [Dropped head syndrome: diagnostic discussion apropos of 3 cases]. *Rev Neurol (Paris)* **153**, 251-255 (1997).
14. Phanthumchinda, K., Sinswaiwong, S. & Jonpiputvanich, S. Chronic progressive external ophthalmoplegia. *J Med Assoc Thai* **80**, 791-794 (1997).
15. Ponseti, J.M., *et al.* Long-term results of tacrolimus in cyclosporine- and prednisone-dependent myasthenia gravis. *Neurology* **64**, 1641-1643 (2005).
16. Raghig, H., Young, G.B., Hammond, R. & Nicolle, M. A comparison of EMG and muscle biopsy in ICU weakness. *Neurocrit Care* **13**, 326-330 (2010).
17. Slucka, C. & Kujawa, H. [Electrocardiographic changes in some muscular diseases (excluding progressive dystrophy). Preliminary report]. *Neurol Neurochir Psychiatr Pol* **15**, 691-694 (1965).
18. Somnier, F.E., Skeie, G.O., Aarli, J.A. & Trojaborg, W. EMG evidence of myopathy and the occurrence of titin autoantibodies in patients with myasthenia gravis. *Eur J Neurol* **6**, 555-563 (1999).
19. Souadjian, J.V., Enriquez, P., Silverstein, M.N. & Pepin, J.M. The spectrum of diseases associated with thymoma. Coincidence or syndrome? *Arch Intern Med* **134**, 374-379 (1974).
20. Yamaguchi, K., Komori, T., Hirose, K. & Tanabe, H. [Clinical analysis of the complex repetitive discharge following M wave]. *Rinsho Shinkeigaku* **35**, 908-910 (1995).
21. Zhu, X.X. [Clinical analysis of 22 cases of hyperthyroidism complicated with myopathy--with pathological data in one case (author's transl)]. *Zhonghua Nei Ke Za Zhi* **20**, 743-746 (1981).

***B. Isolated IIM with TET***

• *Lack of evidence for the presence of TET*:

1. Wang, H., Li, H., Kai, C. & Deng, J. Polymyositis associated with hypothyroidism or hyperthyroidism: two cases and review of the literature. *Clin Rheumatol* **30**, 449-458 (2011).

• *Lack of evidence for at least possible myositis (excluding amyopathic DM)*:

1. Laperuta, P., *et al.* Extrathoracic recurrence of type A thymoma. *Int J Surg* **12 Suppl 1**, S16-18 (2014).
2. Mahfoudhi, M. & Khamassi, K. [Amyopathic dermatomyositis revealing a thymic carcinoma]. *Pan Afr Med J* **21**, 292 (2015).
3. Tsuchiya, T., Sano, A. & Kawashima, M. Paraneoplastic Dermatomyositis as a Potential Precursor to Thymic Carcinoma. *Ann Thorac Surg* **109**, e247-e249 (2020).

• *Definitive diagnosis of MG or subclinical MG*:

1. Ago, T., *et al.* Dermatomyositis associated with invasive thymoma. *Intern Med* **38**, 155-159 (1999).
2. Akatsuka, S. & Torigata, C. [Malignant thymoma associated with polymyositis--an autopsy case]. *Nihon Rinsho* **35**, 1788-1792 (1977).
3. Gidron, A., Quadrini, M., Dimov, N. & Argiris, A. Malignant thymoma associated with fatal myocarditis and polymyositis in a 32-year-old woman with a history of hairy cell leukemia. *Am J Clin Oncol* **29**, 213-214 (2006).
4. Katabami, S., *et al.* Polymyositis associated with thymoma and the subsequent development of pure red cell aplasia. *Intern Med* **34**, 569-573 (1995).
5. Kelly, R.J., Browne, S.K., Rajan, A. & Giaccone, G. Thymoma-associated paraneoplastic polymyositis. *J Clin Oncol* **28**, e378 (2010).
6. Koul, D., *et al.* Fulminant giant cell myocarditis and cardiogenic shock: an unusual presentation of malignant thymoma. *Cardiol Res Pract* **2010**, 185896 (2010).

#### *C. ICI-Induced Myositis and/or MG*

• <u>*Lack of either ICI-induced MG (including MG-like syndrome) or evidence for at least possible myositis (excluding DM without muscle involvement)*</u>:

1. Abdallah, A.O., *et al.* Ipilimumab-induced necrotic myelopathy in a patient with metastatic melanoma: A case report and review of literature. *J Oncol Pharm Pract* **22**, 537-542 (2016).
2. Abushalha, K., Abulaimoun, S. & Silberstein, P.T. So slow, so fast, a case of nivolumab-induced hypothyroidism with subsequent rhabdomyolysis. *Immunotherapy* **12**, 625-628 (2020).
3. Adachi, Y., *et al.* Reduced doses of dabrafenib and trametinib combination therapy for BRAF V600E-mutant non-small cell lung cancer prevent rhabdomyolysis and maintain tumor shrinkage: a case report. *BMC Cancer* **20**, 156 (2020).
4. Agrawal, K. & Agrawal, N. Lambert-Eaton Myasthenic Syndrome Secondary to Nivolumab and Ipilimumab in a Patient with Small-Cell Lung Cancer. *Case Rep Neurol Med* **2019**, 5353202 (2019).
5. Alhammad, R.M., *et al.* Brachial Plexus Neuritis Associated With Anti-Programmed Cell Death-1 Antibodies: Report of 2 Cases. *Mayo Clin Proc Innov Qual Outcomes* **1**, 192-197 (2017).
6. Alkharashi, M.S., Al-Essa, R.S., Otaif, W. & Algorashi, I. Corneal Perforation in a Patient Treated with Atezolizumab-Bevacizumab Combination Therapy for Unresectable Hepatocellular Carcinoma. *Am J Case Rep* **24**, e940688 (2023).
7. Arangalage, D., *et al.* Acute cardiac manifestations under immune checkpoint inhibitors-beware of the obvious: a case report. *Eur Heart J Case Rep* **5**, ytab262 (2021).
8. Barbacki, A., Maliha, P.G., Hudson, M. & Small, D. A case of severe Pembrolizumab-induced neutropenia. *Anticancer Drugs* **29**, 817-819 (2018).
9. Bell, P.T., Beaton, T., Terrill, M., Gillis, D. & Goddard, J. Anti-synthetase syndrome associated interstitial lung disease after combination dual immune checkpoint inhibition. *Respirol Case Rep* **11**, e01115 (2023).
10. Benassaia, E., Vallet, A., Rouleau, E., Ederhy, S. & Robert, C. Troponin increase during immunotherapy: Not always myocarditis. *Eur J Cancer* **157**, 424-427 (2021).
11. Boutros, A., Vera, L., Gatto, F., Fornarini, G. & Zanardi, E. Case report: Pembrolizumab plus Axitinib related hypothyroid myopathy in two kidney cancer patients. *Front Oncol* **12**, 1048526 (2022).
12. Bruno, F., *et al.* Pembrolizumab-Induced Isolated Cranial Neuropathy: A Rare Case Report and Review of Literature. *Front Neurol* **12**, 669493 (2021).
13. Cafuir, L., Lawson, D., Desai, N., Kesner, V. & Voloschin, A. Inflammatory demyelinating polyneuropathy versus leptomeningeal disease following Ipilimumab. *J Immunother Cancer* **6**, 11 (2018).
14. Calabrese, C., *et al.* Polymyalgia rheumatica-like syndrome from checkpoint inhibitor therapy: case series and systematic review of the literature. *RMD Open* **5**, e000906 (2019).
15. Cao, C., *et al.* A Muscle-Invasive Bladder Cancer Patient With High Tumor Mutational Burden and RB1 Mutation Achieved Bladder Preservation Following Chemotherapy Combined With Immunotherapy: A Case Report. *Front Immunol* **12**, 684879 (2021).
16. Chae, J. & Peikert, T. Myasthenia gravis crisis complicating anti-PD-L1 cancer immunotherapy. *Crit. Care Med* **46**, 280 (2018).
17. Cheng, Y., Nie, L., Ma, W. & Zheng, B. Early Onset Acute Coronary Artery Occlusion After Pembrolizumab in Advanced Non-Small Cell Lung Cancer: A Case Report. *Cardiovasc Toxicol* **21**, 683-686 (2021).

18. Clark, J.I., Bufalino, S., Singh, S. & Borys, E. Rhabdomyolysis during high dose interleukin-2 treatment of metastatic melanoma after sequential immunotherapies: a case report. *J Immunother Cancer* **6**, 53 (2018).
19. Cobarro, L., *et al.* Immune Checkpoint Inhibitor-Related Stress Cardiomyopathy: Differential Diagnosis and Key Role of Cardiac Imaging. *JACC Case Rep* **16**, 101881 (2023).
20. Daoussis, D., Kraniotis, P., Liossis, S.N. & Solomou, A. Immune checkpoint inhibitor-induced myo-fasciitis. *Rheumatology (Oxford)* **56**, 2161 (2017).
21. Desikan, S.P., Varghese, R., Kamoga, R. & Desikan, R. Acute hyponatremia from immune checkpoint inhibitor therapy for non-small cell lung cancer. *Postgrad Med J* **96**, 570-571 (2020).
22. Ederhy, S., *et al.* Takotsubo-Like Syndrome in Cancer Patients Treated With Immune Checkpoint Inhibitors. *JACC Cardiovasc Imaging* **11**, 1187-1190 (2018).
23. Eisenbud, L., Ejadi, S. & Mar, N. Development of carpal tunnel syndrome in association with checkpoint inhibitors. *J Oncol Pharm Pract* **27**, 764-765 (2021).
24. Fellner, A., *et al.* Neurologic complications of immune checkpoint inhibitors. *J Neurooncol* **137**, 601-609 (2018).
25. Gaibor, C., Das, R. & Reddy, V. Pembrolizumab-Induced Hypothyroidism: A Case Report. *Cureus* **15**, e41889 (2023).
26. Gill, A.J., Gandhy, S. & Lancaster, E. Nivolumab-associated Lambert-Eaton myasthenic syndrome and cerebellar dysfunction in a patient with a neuroendocrine tumor. *Muscle Nerve* **63**, E18-E21 (2021).
27. Goldstein, B.L., Gedmintas, L. & Todd, D.J. Drug-associated polymyalgia rheumatica/giant cell arteritis occurring in two patients after treatment with ipilimumab, an antagonist of ctla-4. *Arthritis Rheumatol* **66**, 768-769 (2014).
28. Gra, M., *et al.* Brief Communication: Lambert-Eaton Myasthenic Paraneoplastic Syndrome Associated With Merkel Cell Carcinoma Successfully Treated by Immune Checkpoint Inhibitors: 2 Cases. *J Immunother* **46**, 276-278 (2023).
29. Hara, S., *et al.* [A Case of Metastatic Ureteral Cancer Treated with Pembrolizumab without Relapse of Ocular Myasthenia Gravis]. *Hinyokika Kiyo* **68**, 377-383 (2022).
30. Hasegawa, T., *et al.* Nivolumab-related severe thrombocytopenia in a patient with relapsed lung adenocarcinoma: a case report and review of the literature. *J Med Case Rep* **13**, 316 (2019).
31. Hsu, C.Y., Su, Y.W. & Chen, S.C. Sick sinus syndrome associated with anti-programmed cell death-1. *J Immunother Cancer* **6**, 72 (2018).
32. Ibrahim, T., Adam, C., Routier, E., Slama, A. & Robert, C. Mitochondrial myopathy associated with anti-programmed cell death 1 therapy. *Eur J Cancer* **110**, 71-73 (2019).
33. Ichihara, S., *et al.* Immune checkpoint inhibitor-related pneumonitis with atypical radiologic features in a patient with anti-aminoacyl-tRNA synthetase antibody. *Respir Med Case Rep* **41**, 101797 (2023).
34. Ida, M., *et al.* Subtle-but-smouldering myocardial injury after immune checkpoint inhibitor treatment accompanied by amyloid deposits. *ESC Heart Fail* **9**, 2027-2031 (2022).
35. Imai, Y., Tanaka, M., Fujii, R., Uchitani, K. & Okazaki, K. [Effectiveness of a Low-dose Corticosteroid in a Patient with Polymyalgia Rheumatica Associated with Nivolumab Treatment]. *Yakugaku Zasshi* **139**, 491-495 (2019).
36. Irimada, M., *et al.* Severe rhabdomyolysis developing in an advanced melanoma patient treated by pembrolizumab followed by dabrafenib trametinib combined therapy. *J Dermatol* **46**, e256-e258 (2019).

37. Ishii, A., Yokoyama, M., Tsuji, H., Fujii, Y. & Tamaoka, A. Pembrolizumab treatment of metastatic urothelial cancer without exacerbating myasthenia gravis. *eNeurologicalSci* **19**, 100236 (2020).
38. Ishikawa, M. & Oashi, K. Case of hypophysitis caused by nivolumab. *J Dermatol* **44**, 109-110 (2017).
39. Iskandar, A., Hwang, A. & Dasanu, C.A. Polymyalgia rheumatica due to pembrolizumab therapy. *J Oncol Pharm Pract* **25**, 1282-1284 (2019).
40. Izzedine, H., *et al.* Kidney injuries related to ipilimumab. *Invest New Drugs* **32**, 769-773 (2014).
41. Jaffer, M., *et al.* Immunotherapy Induced Myasthenic-Like Syndrome in a Metastatic Melanoma Patient With Amyotrophic Lateral Sclerosis. *Clin Med Insights Oncol* **14**, 1179554920978024 (2020).
42. Jiwa, N.S., Lawrence, D.P. & Guidon, A.C. Dual hereditary and immune-mediated neuromuscular diagnoses after cancer immunotherapy. *Muscle Nerve* **63**, E21-E24 (2021).
43. Johnson, E.D., Kerrigan, K., Butler, K. & Patel, S.B. Nivolumab-induced hypothyoidism with consequent hypothyroid related myopathy. *J Oncol Pharm Pract* **26**, 224-227 (2020).
44. Jordan, B., Zierz, S., Weber, T. & Jordan, K. Successful use of an immune checkpoint inhibitor in a patient with myasthenia gravis in remission. *Muscle Nerve* **60**, E7-E8 (2019).
45. Kawataki, M., Nakanishi, Y., Yokoyama, T. & Ishida, T. Hypothyroidism as an immune-related adverse event caused by atezolizumab in a patient with muscle spasms: a case report. *Respir Med Case Rep* **36**, 101585 (2022).
46. Khan, A., Riaz, S. & Carhart, R., Jr. Pembrolizumab-Induced Mobitz Type 2 Second-Degree Atrioventricular Block. *Case Rep Cardiol* **2020**, 8428210 (2020).
47. Kim, K.H., Baek, Y.H., Kang, Y.W., Yoon, B.A. & Moon, S.Y. A Case of Transverse Myelitis Following Treatment with Atezolizumab for Advanced Hepatocellular Carcinoma. *Korean J Gastroenterol* **82**, 35-39 (2023).
48. Kim, S.Y., Kim, D.K., Choi, S.Y. & Chung, C. Comparative analysis of immunotherapy responses in small cell lung cancer patients with dermatomyositis. *Thorac Cancer* **15**, 672-677 (2024).
49. Kim, Y., Park, D., Choi, S.Y. & Chung, C. Immune checkpoint inhibitor therapy in a patient with small cell lung cancer and anti-transcriptional intermediary factor 1-gamma antibody-positive dermatomyositis: A case report. *Thorac Cancer* **13**, 2808-2811 (2022).
50. Kita, T., *et al.* Nivolumab-Induced Polymyalgia Rheumatica in a Patient with Lung Adenocarcinoma. *Am J Med Sci* **362**, 321-323 (2021).
51. Kobak, S. Pembrolizumab-Induced Seronegative Arthritis and Fasciitis in a Patient with Lung Adenocarcinoma. *Curr Drug Saf* **14**, 225-229 (2019).
52. Kodama, S., Kurose, K., Mukai, T. & Morita, Y. Nivolumab-induced polyarthritis. *BMJ Case Rep* **2017**(2017).
53. Kosche, C., Owen, J.L. & Choi, J.N. Widespread subacute cutaneous lupus erythematosus in a patient receiving checkpoint inhibitor immunotherapy with ipilimumab and nivolumab. *Dermatol Online J* **25**(2019).
54. Krusche, M., *et al.* [Myofasciitis under nivolumab treatment]. *Z Rheumatol* **80**, 884-888 (2021).
55. Kunii, E., *et al.* Lambert-Eaton Myasthenic Syndrome Caused by Atezolizumab in a Patient with Small-cell Lung Cancer. *Intern Med* **61**, 1739-1742 (2022).
56. Kuswanto, W.F., *et al.* Rheumatologic symptoms in oncologic patients on PD-1 inhibitors. *Semin Arthritis Rheum* **47**, 907-910 (2018).
57. Le Bras, P., Plard, L., Le Gouill, C., Piriou, N. & Touchefeu, Y. Transthyretin Cardiac Amyloidosis Mimicking Immune Checkpoint-Induced Myocarditis in a Patient Treated with

Atezolizumab and Bevacizumab for Advanced Hepatocellular Carcinoma: A Case Report. *Case Rep Oncol* **15**, 967-973 (2022).
58. Lee, J.C., Al-Humimat, G. & Kooner, K.S. Acute Bilateral Uveitis, Hypotony, and Cataracts Associated with Ipilimumab and Nivolumab Therapy: Optical Coherence Tomography Angiography Findings. *Case Rep Ophthalmol* **11**, 606-611 (2020).
59. Lee, J.H.*, et al.* Lambert-Eaton myasthenic syndrome (LEMS) in a patient with lung cancer under treatment with pembrolizumab: a case study. *J Chemother* **35**, 275-280 (2023).
60. Lobo, Y., Good, P. & Murphy, F. Polymyalgia rheumatica in a melanoma patient 11 months after completion of immunotherapy with nivolumab. *Cancer Rep (Hoboken)* **3**, e1244 (2020).
61. Loricera, J.*, et al.* Subclinical aortitis after starting nivolumab in a patient with metastatic melanoma. A case of drug-associated aortitis? *Clin Exp Rheumatol* **36 Suppl 111**, 171 (2018).
62. Machiyama, H. & Minami, S. Durvalumab for Extensive-Stage of Small-Cell Lung Cancer With Lambert-Eaton Myasthenic Syndrome. *J Med Cases* **14**, 71-75 (2023).
63. Mackintosh, D., Islam, M.F., Ng, J. & Basham, J. Immune checkpoint inhibitor use in antisynthetase syndrome. *Asia Pac J Clin Oncol* **15**, 266-269 (2019).
64. Mahmood, S.S.*, et al.* Myocarditis with tremelimumab plus durvalumab combination therapy for endometrial cancer: A case report. *Gynecol Oncol Rep* **25**, 74-77 (2018).
65. Maniu, C., Kobe, C., Schlaak, M., Mauch, C. & Eming, S.A. Polymyalgia rheumatica occurring during treatment with ipilimumab. *Eur J Dermatol* **26**, 513-514 (2016).
66. Masson, R., Manthripragada, G., Liu, R., Tavakoli, J. & Mok, K. Possible Precipitation of Acute Coronary Syndrome with Immune Checkpoint Blockade: A Case Report. *Perm J* **24**, 1 (2020).
67. Mazzarella, L.*, et al.* Evidence for interleukin 17 involvement in severe immune-related neuroendocrine toxicity. *Eur J Cancer* **141**, 218-224 (2020).
68. McElnea, E., Ni Mhealoid, A., Moran, S., Kelly, R. & Fulcher, T. Thyroid-like ophthalmopathy in a euthyroid patient receiving Ipilimumab. *Orbit* **33**, 424-427 (2014).
69. Mencel, J.*, et al.* Thymic hyperplasia following double immune checkpoint inhibitor therapy in two patients with stage IV melanoma. *Asia Pac J Clin Oncol* **15**, 383-386 (2019).
70. Min, L. & Hodi, F.S. Anti-PD1 following ipilimumab for mucosal melanoma: durable tumor response associated with severe hypothyroidism and rhabdomyolysis. *Cancer Immunol Res* **2**, 15-18 (2014).
71. Moradi, L.A., Clark, C.A., Schneider, C.S., Deshane, A.S. & Dobelbower, M.C. Durable Metastatic Melanoma Remission Following Pembrolizumab and Radiotherapy: A Case Report of Prophylactic Immunosuppression in a Patient with Myasthenia Gravis and Immune-Mediated Colitis. *Front Immunol* **12**, 788499 (2021).
72. Murakami, S.*, et al.* Tenosynovitis Induced by an Immune Checkpoint Inhibitor: A Case Report and Literature Review. *Intern Med* **58**, 2839-2843 (2019).
73. Muto, Y.*, et al.* Success of rechallenging dabrafenib and trametinib combination therapy after trametinib-induced rhabdomyolysis: a case report. *Melanoma Res* **28**, 151-154 (2018).
74. Nakamagoe, K.*, et al.* Polymyalgia rheumatica in a melanoma patient due to nivolumab treatment. *J Cancer Res Clin Oncol* **143**, 1357-1358 (2017).
75. Nakatani, Y.*, et al.* Lambert-Eaton Myasthenic Syndrome Caused by Nivolumab in a Patient with Squamous Cell Lung Cancer. *Case Rep Neurol* **10**, 346-352 (2018).
76. Nguyen, C.B., Su, C.T., Morgan, M. & Alva, A.S. Case report: Immune-mediated meibomian gland dysfunction following pembrolizumab therapy for advanced urothelial carcinoma. *Front Oncol* **12**, 1000023 (2022).
77. Oldfield, K., Jayasinghe, R., Niranjan, S. & Chadha, S. Immune checkpoint inhibitor-induced takotsubo syndrome and diabetic ketoacidosis: rare reactions. *BMJ Case Rep* **14**(2021).

78. Ouyang, Z.M.*, et al.* [Immune checkpoint inhibitor-induced eosinophilic fasciitis: a case report and literature review]. *Zhonghua Nei Ke Za Zhi* **62**, 182-187 (2023).
79. Papavasileiou, E., Prasad, S., Freitag, S.K., Sobrin, L. & Lobo, A.M. Ipilimumab-induced Ocular and Orbital Inflammation--A Case Series and Review of the Literature. *Ocul Immunol Inflamm* **24**, 140-146 (2016).
80. Park, C. & Kim, K.T. Demyelinating polyneuropathy combined with brachial plexopathy after nivolumab therapy for hodgkin lymphoma: a case report. *BMC Neurol* **23**, 130 (2023).
81. Perrotta, F.M., Scriffignano, S., Fatica, M., Specchia, M. & Lubrano, E. Case report of polymyalgia rheumatica in a male patient with three different neoplasms treated with pembrolizumab. *Reumatismo* **72**, 178-181 (2020).
82. Polak, P., Speldova, J., Bratova, M., Zavrelova, J. & Penka, M. [Pembrolizumab-induced hypothyreosis and subcutaneous bleeding]. *Vnitr Lek* **67**, 175-179 (2021).
83. Pourhassan, H.Z., Tryon, D., Schaeffer, B., Mirshahidi, H. & Wong, J. Autoimmune rhabdomyolysis and a multiorgan display of PD-1 inhibitor induced immune related adverse events during treatment of metastatic melanoma. *Exp Hematol Oncol* **8**, 20 (2019).
84. Robilliard, B., Arnaud, E., Gastaud, L. & Broner, J. A case of pembrolizumab-induced autoimmune haemolytic anaemia with polymyalgia rheumatica. *Eur J Cancer* **103**, 281-283 (2018).
85. Sakaguchi, T.*, et al.* An Extensive-stage Small-cell Lung Cancer Case With Preexisting Lambert-Eaton Myasthenic Syndrome Successfully Treated With an Immune Checkpoint Inhibitor. *Clin Lung Cancer* **23**, e273-e275 (2022).
86. Saruwatari, K.*, et al.* The Risks and Benefits of Immune Checkpoint Blockade in Anti-AChR Antibody-Seropositive Non-Small Cell Lung Cancer Patients. *Cancers (Basel)* **11**(2019).
87. Sato, I., Nakaya, N., Obara, Y., Ueno, S. & Nakajima, H. [Advanced Gastric Cancer with Tumor Shrinkage Persisting after the Discontinuation of Nivolumab-A Case Report]. *Gan To Kagaku Ryoho* **47**, 1715-1717 (2020).
88. Savarapu, P., Abdelazeem, B., Isa, S., Kesari, K. & Kunadi, A. Ipilimumab and Nivolumab induced ventricular tachycardia in a patient with metastatic renal cell carcinoma. *J Community Hosp Intern Med Perspect* **11**, 874-876 (2021).
89. Schmidt, T.*, et al.* Case Report: Pseudomeningeosis and Demyelinating Metastasis-Like Lesions From Checkpoint Inhibitor Therapy in Malignant Melanoma. *Front Oncol* **11**, 637185 (2021).
90. Schwab, A.*, et al.* Pembrolizumab-Induced Myasthenia Gravis and Myositis: Literature Review on Neurological Toxicities of Programmed Death Protein 1 Inhibitors. *J Med Cases* **13**, 530-535 (2022).
91. Sepulveda, M.*, et al.* Motor polyradiculopathy during pembrolizumab treatment of metastatic melanoma. *Muscle Nerve* **56**, E162-E167 (2017).
92. Takigawa, Y.*, et al.* Lambert-Eaton Myasthenic Syndrome Recurrence Induced by Pembrolizumab in a Patient with Non-small-cell Lung Cancer. *Intern Med* **62**, 1055-1058 (2023).
93. Trachtenberg, B., Hussain, F., Mukherjee, A., Araujo-Gutierrez, R. & Pingali, S.R.K. Immune Checkpoint Inhibitor-Related Cardiotoxicity. *Methodist Debakey Cardiovasc J* **14**, e1-e4 (2018).
94. Trevisani, F.*, et al.* Renal function outcomes in patients with muscle-invasive bladder cancer treated with neoadjuvant pembrolizumab and radical cystectomy in the PURE-01 study. *Int J Cancer* **149**, 186-190 (2021).

95. Tu, L., Liu, J., Li, Z., Liu, Y. & Luo, F. Early detection and management of immune-related myocarditis: Experience from a case with advanced squamous cell lung carcinoma. *Eur J Cancer* **131**, 5-8 (2020).
96. van Binsbergen, W.H., *et al.* [Rheumatic adverse events due to immune checkpoint inhibitors]. *Ned Tijdschr Geneeskd* **167**(2023).
97. Vartanov, A., *et al.* Immunotherapy-associated complete heart block in a patient with NSCLC: A case report and literature review. *Respir Med Case Rep* **33**, 101390 (2021).
98. Wang, L., *et al.* Case report: Significant response to PD-L1 inhibitor after resistance to PD-1 inhibitor in an advanced alpha-fetoprotein-positive gastric cancer. *Front Oncol* **12**, 962126 (2022).
99. Williams, S., Liang, C., Guminski, A., Hruby, G. & Chan, D. Outcome of patient with myasthenia gravis with the use of immunotherapy in metastatic Merkel cell carcinoma. *Oxf Med Case Reports* **2022**, omac012 (2022).
100. Yamagata, N., Michizaki, H., Komatsu, S., Kobayashi, Y. & Machida, A. Pembrolizumab-induced radiation recall myopathy with fasciopathy: A case report. *Muscle Nerve* **68**, E41-E43 (2023).
101. Yamazoe, M., Hatakeyama, T., Furukawa, K., Kato, K. & Horiuchi, K. Atezolizumab-Induced Lambert-Eaton Myasthenic Syndrome in a Patient With Small-Cell Lung Cancer. *Cureus* **15**, e33557 (2023).
102. Yu, W.Y., North, J.P., McCalmont, T.H. & Shinkai, K. Wong-type dermatomyositis during anti-PD-1 therapy. *JAAD Case Rep* **4**, 1049-1051 (2018).
103. Yuen, C., *et al.* Severe Relapse of Vaccine-Induced Guillain-Barre Syndrome After Treatment With Nivolumab. *J Clin Neuromuscul Dis* **20**, 194-199 (2019).
104. Zampeli, E. & Zervas, E. Eosinophilic Fasciitis following Checkpoint Inhibitor Therapy with Pembrolizumab. *Mediterr J Rheumatol* **32**, 376-377 (2021).
105. Zaremba, A., *et al.* Metastatic Merkel cell carcinoma and myasthenia gravis: contraindication for therapy with immune checkpoint inhibitors? *J Immunother Cancer* **7**, 141 (2019).
106. Zekic, T. & Benic, M.S. Anti-programmed death-1 inhibitor nivolumab-induced immune-related adverse events: hepatitis, renal insufficiency, myositis, vitiligo, and hypothyroidism: a case-based review. *Rheumatol Int* **43**, 559-565 (2023).
107. Zhang, X., Gao, B.X., Guo, C.Y. & Su, T. A 71-year-old male with a life-threatening recurrence of hemolytic anemia, thrombocytopenia, and acute kidney injury after pembrolizumab therapy: a case report. *BMC Geriatr* **23**, 478 (2023).
108. Zhao, L.Z., Liu, G., Li, Q.F., Chen, G. & Jin, G.W. A case of carrelizumab-associated immune myocarditis. *Asian J Surg* **45**, 496-497 (2022).

• <u>*Lack of ICI treatment*</u>:

1. Arabshahi, B., Silverman, R.A., Jones, O.Y. & Rider, L.G. Abatacept and sodium thiosulfate for treatment of recalcitrant juvenile dermatomyositis complicated by ulceration and calcinosis. *J Pediatr* **160**, 520-522 (2012).
2. Aurensanz-Clemente, E., *et al.* Acute myocarditis with transient myocardial thickening in two oncologic patients treated with anti-GD2 immunotherapy. *ESC Heart Fail* **10**, 2090-2093 (2023).
3. Cho, Y., *et al.* Diagnostic Dilemma of Paraneoplastic Rheumatic Disorders: Case Series and Narrative Review. *Cureus* **13**, e19993 (2021).
4. Demichelis, C., *et al.* Neuromuscular complications following targeted therapy in cancer patients: beyond the immune checkpoint inhibitors. Case reports and review of the literature. *Neurol Sci* **42**, 1405-1409 (2021).

5. Doughty, C.T. & Amato, A.A. Toxic Myopathies. *Continuum (Minneap Minn)* **25**, 1712-1731 (2019).
6. Gauci, M.L.*, et al.* Focal necrotizing myopathy with 'dropped-head syndrome' induced by cobimetinib in metastatic melanoma. *Melanoma Res* **27**, 511-515 (2017).
7. Hegde, M.*, et al.* Tumor response and endogenous immune reactivity after administration of HER2 CAR T cells in a child with metastatic rhabdomyosarcoma. *Nat Commun* **11**, 3549 (2020).
8. Hunt, S.V., Garrott, H.M., Williams, M.E. & Ford, R.L. Bilateral Paraneoplastic Orbital Myositis Associated With Malignant Melanoma and Multiple Myeloma. *Ophthalmic Plast Reconstr Surg* **38**, e72-e75 (2022).
9. Kawashiri, S.Y.*, et al.* Usefulness of ultrasonography-proven tenosynovitis to monitor disease activity of a patient with very early rheumatoid arthritis treated by abatacept. *Mod Rheumatol* **23**, 582-586 (2013).
10. Kerola, A.M. & Kauppi, M.J. Abatacept as a successful therapy for myositis-a case-based review. *Clin Rheumatol* **34**, 609-612 (2015).
11. Koizumi, H.*, et al.* A case of juvenile amyopathic dermatomyositis with anti-transcription intermediary factor 1-alpha antibody showing negative anti-TIF1-gamma ELISA results: Comment on "Case of pembrolizumab-induced dermatomyositis with anti-transcription intermediary factor 1-gamma antibody". *J Dermatol* **50**, e39-e40 (2023).
12. Kwan, J.M.*, et al.* Mogamulizumab-Associated Acute Myocarditis in a Patient With T-Cell Lymphoma. *JACC Case Rep* **3**, 1018-1023 (2021).
13. Lanis, A.*, et al.* Nodular Regenerative Hyperplasia of the liver in Juvenile Dermatomyositis. *Pediatr Rheumatol Online J* **20**, 30 (2022).
14. Maeshima, K.*, et al.* Successful treatment of refractory anti-signal recognition particle myopathy using abatacept. *Rheumatology (Oxford)* **53**, 379-380 (2014).
15. Musuruana, J.L. & Cavallasca, J.A. Abatacept for treatment of refractory polymyositis. *Joint Bone Spine* **78**, 431-432 (2011).
16. Pejic, M., Shifman, M., Rose, T. & Jeong, D. Chronic Anthracycline-related Myocarditis Presenting as Diffuse Myocardial Calcification. *J Clin Imaging Sci* **9**, 47 (2019).
17. Rodziewicz, M. & Kiely, P. The successful use of subcutaneous abatacept in refractory anti-human transcriptional intermediary factor 1-gamma dermatomyositis skin and oesphagopharyngeal disease. *Rheumatology (Oxford)* **57**, 1866-1867 (2018).
18. Sakamoto, T., Saito, Y., Takekuma, Y., Kikuchi, E. & Sugawara, M. Gefitinib-induced Myositis: A Novel Case Report. *Yakugaku Zasshi* **143**, 617-620 (2023).
19. Slean, G.R. & Silkiss, R.Z. Lenalidomide-Associated Thyroid-Related Eyelid Retraction. *Ophthalmic Plast Reconstr Surg* **34**, e46-e48 (2018).
20. Takizawa, T.*, et al.* New onset of myasthenia gravis after intravesical Bacillus Calmette-Guerin: A case report and literature review. *Medicine (Baltimore)* **96**, e8757 (2017).
21. Tjarnlund, A.*, et al.* Abatacept in the treatment of adult dermatomyositis and polymyositis: a randomised, phase IIb treatment delayed-start trial. *Ann Rheum Dis* **77**, 55-62 (2018).
22. Zheng, Y.*, et al.* Myasthenia gravis associated with renal cell carcinoma: a paraneoplastic syndrome or just a coincidence. *BMC Neurol* **21**, 277 (2021).
23. Ziogas, D.C.*, et al.* Neuromuscular Complications of Targeted Anticancer Agents: Can Tyrosine Kinase Inhibitors Induce Myasthenia Gravis? Getting Answers From a Case Report up to a Systematic Review. *Front Oncol* **11**, 727010 (2021).

• *Lack of case-level detail*:

1. Altan, M., *et al.* Nivolumab and ipilimumab with concurrent stereotactic radiosurgery for intracranial metastases from non-small cell lung cancer: analysis of the safety cohort for non-randomized, open-label, phase I/II trial. *J Immunother Cancer* **11**(2023).
2. Antonia, S., *et al.* Safety and antitumour activity of durvalumab plus tremelimumab in non-small cell lung cancer: a multicentre, phase 1b study. *Lancet Oncol* **17**, 299-308 (2016).
3. Antonia, S.J., *et al.* Nivolumab alone and nivolumab plus ipilimumab in recurrent small-cell lung cancer (CheckMate 032): a multicentre, open-label, phase 1/2 trial. *Lancet Oncol* **17**, 883-895 (2016).
4. Balar, A.V., *et al.* First-line pembrolizumab in cisplatin-ineligible patients with locally advanced and unresectable or metastatic urothelial cancer (KEYNOTE-052): a multicentre, single-arm, phase 2 study. *Lancet Oncol* **18**, 1483-1492 (2017).
5. Barlesi, F., *et al.* Avelumab versus docetaxel in patients with platinum-treated advanced non-small-cell lung cancer (JAVELIN Lung 200): an open-label, randomised, phase 3 study. *Lancet Oncol* **19**, 1468-1479 (2018).
6. Chen, R., *et al.* Case Report: Cardiac Toxicity Associated With Immune Checkpoint Inhibitors. *Front Cardiovasc Med* **8**, 727445 (2021).
7. Cho, J., *et al.* Pembrolizumab for Patients With Refractory or Relapsed Thymic Epithelial Tumor: An Open-Label Phase II Trial. *J Clin Oncol* **37**, 2162-2170 (2019).
8. Choueiri, T.K., *et al.* Preliminary results for avelumab plus axitinib as first-line therapy in patients with advanced clear-cell renal-cell carcinoma (JAVELIN Renal 100): an open-label, dose-finding and dose-expansion, phase 1b trial. *Lancet Oncol* **19**, 451-460 (2018).
9. Chung, V., *et al.* Evaluation of safety and efficacy of p53MVA vaccine combined with pembrolizumab in patients with advanced solid cancers. *Clin Transl Oncol* **21**, 363-372 (2019).
10. Conforti, F., *et al.* Avelumab plus axitinib in unresectable or metastatic type B3 thymomas and thymic carcinomas (CAVEATT): a single-arm, multicentre, phase 2 trial. *Lancet Oncol* **23**, 1287-1296 (2022).
11. Eggermont, A.M., *et al.* Adjuvant ipilimumab versus placebo after complete resection of high-risk stage III melanoma (EORTC 18071): a randomised, double-blind, phase 3 trial. *Lancet Oncol* **16**, 522-530 (2015).
12. Eggermont, A.M.M., *et al.* Adjuvant Pembrolizumab versus Placebo in Resected Stage III Melanoma. *N Engl J Med* **378**, 1789-1801 (2018).
13. Fujiwara, Y., *et al.* Phase I Study of Tremelimumab Monotherapy or in Combination With Durvalumab in Japanese Patients With Advanced Solid Tumors or Malignant Mesothelioma. *Oncologist* **27**, e703-e722 (2022).
14. Giaccone, G., *et al.* Pembrolizumab in patients with thymic carcinoma: a single-arm, single-centre, phase 2 study. *Lancet Oncol* **19**, 347-355 (2018).
15. Girard, N., *et al.* Efficacy and safety of nivolumab for patients with pre-treated type B3 thymoma and thymic carcinoma: results from the EORTC-ETOP NIVOTHYM phase II trial. *ESMO Open* **8**, 101576 (2023).
16. Graff, J.N., *et al.* Early evidence of anti-PD-1 activity in enzalutamide-resistant prostate cancer. *Oncotarget* **7**, 52810-52817 (2016).
17. Heery, C.R., *et al.* Avelumab for metastatic or locally advanced previously treated solid tumours (JAVELIN Solid Tumor): a phase 1a, multicohort, dose-escalation trial. *Lancet Oncol* **18**, 587-598 (2017).
18. Juergens, R.A., *et al.* A phase IB study of durvalumab with or without tremelimumab and platinum-doublet chemotherapy in advanced solid tumours: Canadian Cancer Trials Group Study IND226. *Lung Cancer* **143**, 1-11 (2020).

19. Long, G.V., *et al.* Standard-Dose Pembrolizumab Plus Alternate-Dose Ipilimumab in Advanced Melanoma: KEYNOTE-029 Cohort 1C, a Phase 2 Randomized Study of Two Dosing Schedules. *Clin Cancer Res* **27**, 5280-5288 (2021).
20. Maio, M., *et al.* Pembrolizumab in microsatellite instability high or mismatch repair deficient cancers: updated analysis from the phase II KEYNOTE-158 study. *Ann Oncol* **33**, 929-938 (2022).
21. Mateos, M.V., *et al.* Pembrolizumab plus pomalidomide and dexamethasone for patients with relapsed or refractory multiple myeloma (KEYNOTE-183): a randomised, open-label, phase 3 trial. *Lancet Haematol* **6**, e459-e469 (2019).
22. Monge, C., *et al.* Phase I/II study of PexaVec in combination with immune checkpoint inhibition in refractory metastatic colorectal cancer. *J Immunother Cancer* **11**(2023).
23. O'Brien, M., *et al.* Pembrolizumab versus placebo as adjuvant therapy for completely resected stage IB-IIIA non-small-cell lung cancer (PEARLS/KEYNOTE-091): an interim analysis of a randomised, triple-blind, phase 3 trial. *Lancet Oncol* **23**, 1274-1286 (2022).
24. Peleg Hasson, S., *et al.* Re-introducing immunotherapy in patients surviving immune checkpoint inhibitors-mediated myocarditis. *Clin Res Cardiol* **110**, 50-60 (2021).
25. Pignata, S., *et al.* Carboplatin and paclitaxel plus avelumab compared with carboplatin and paclitaxel in advanced or recurrent endometrial cancer (MITO END-3): a multicentre, open-label, randomised, controlled, phase 2 trial. *Lancet Oncol* **24**, 286-296 (2023).
26. Postel-Vinay, S., *et al.* First-in-human phase I study of the OX40 agonist GSK3174998 with or without pembrolizumab in patients with selected advanced solid tumors (ENGAGE-1). *J Immunother Cancer* **11**(2023).
27. Sanchez-Conde, M., *et al.* Pembrolizumab in combination with tocilizumab in high-risk hospitalized patients with COVID-19 (COPERNICO): A randomized proof-of-concept phase II study. *Int J Infect Dis* **123**, 97-103 (2022).
28. Sanderson, K., *et al.* Autoimmunity in a phase I trial of a fully human anti-cytotoxic T-lymphocyte antigen-4 monoclonal antibody with multiple melanoma peptides and Montanide ISA 51 for patients with resected stages III and IV melanoma. *J Clin Oncol* **23**, 741-750 (2005).
29. Sonpavde, G., *et al.* ENERGIZE: a Phase III study of neoadjuvant chemotherapy alone or with nivolumab with/without linrodostat mesylate for muscle-invasive bladder cancer. *Future Oncol* **16**, 4359-4368 (2020).
30. Suzuki, S., *et al.* Nivolumab-related myasthenia gravis with myositis and myocarditis in Japan. *Neurology* **89**, 1127-1134 (2017).
31. Szabados, B., *et al.* Final Results of Neoadjuvant Atezolizumab in Cisplatin-ineligible Patients with Muscle-invasive Urothelial Cancer of the Bladder. *Eur Urol* **82**, 212-222 (2022).
32. Tang, Q., *et al.* Effect of CTLA4-Ig (abatacept) treatment on T cells and B cells in peripheral blood of patients with polymyositis and dermatomyositis. *Scand J Immunol* **89**, e12732 (2019).
33. Taniguchi, Y., *et al.* A Randomized Comparison of Nivolumab versus Nivolumab + Docetaxel for Previously Treated Advanced or Recurrent ICI-Naive Non-Small Cell Lung Cancer: TORG1630. *Clin Cancer Res* **28**, 4402-4409 (2022).
34. Tawbi, H.A., *et al.* Combined Nivolumab and Ipilimumab in Melanoma Metastatic to the Brain. *N Engl J Med* **379**, 722-730 (2018).
35. Tawbi, H.A., *et al.* Long-term outcomes of patients with active melanoma brain metastases treated with combination nivolumab plus ipilimumab (CheckMate 204): final results of an open-label, multicentre, phase 2 study. *Lancet Oncol* **22**, 1692-1704 (2021).

36. Tong, T.M.L., *et al.* Combining Melphalan Percutaneous Hepatic Perfusion with Ipilimumab Plus Nivolumab in Advanced Uveal Melanoma: First Safety and Efficacy Data from the Phase Ib Part of the Chopin Trial. *Cardiovasc Intervent Radiol* **46**, 350-359 (2023).
37. Ueno, M., *et al.* Nivolumab alone or in combination with cisplatin plus gemcitabine in Japanese patients with unresectable or recurrent biliary tract cancer: a non-randomised, multicentre, open-label, phase 1 study. *Lancet Gastroenterol Hepatol* **4**, 611-621 (2019).
38. Usmani, S.Z., *et al.* Pembrolizumab plus lenalidomide and dexamethasone for patients with treatment-naive multiple myeloma (KEYNOTE-185): a randomised, open-label, phase 3 trial. *Lancet Haematol* **6**, e448-e458 (2019).

• *Review or systematic review without original patient data*:

1. Ao, Y.Q., *et al.* Immunotherapy of thymic epithelial tumors: molecular understandings and clinical perspectives. *Mol Cancer* **22**, 70 (2023).
2. Atallah-Yunes, S.A., Kadado, A.J., Kaufman, G.P. & Hernandez-Montfort, J. Immune checkpoint inhibitor therapy and myocarditis: a systematic review of reported cases. *J Cancer Res Clin Oncol* **145**, 1527-1557 (2019).
3. Ghosh, N., *et al.* Checkpoint Inhibitor-Associated Arthritis: A Systematic Review of Case Reports and Case Series. *J Clin Rheumatol* **27**, e317-e322 (2021).
4. Guidon, A.C. Lambert-Eaton Myasthenic Syndrome, Botulism, and Immune Checkpoint Inhibitor-Related Myasthenia Gravis. *Continuum (Minneap Minn)* **25**, 1785-1806 (2019).
5. Johansen, A., Christensen, S.J., Scheie, D., Hojgaard, J.L.S. & Kondziella, D. Neuromuscular adverse events associated with anti-PD-1 monoclonal antibodies: Systematic review. *Neurology* **92**, 663-674 (2019).
6. Remon, J., *et al.* Immune checkpoint blockers in patients with unresectable or metastatic thymic epithelial tumours: A meta-analysis. *Eur J Cancer* **180**, 117-124 (2023).
7. Takamatsu, K., *et al.* Immune checkpoint inhibitors in the onset of myasthenia gravis with hyperCKemia. *Ann Clin Transl Neurol* **5**, 1421-1427 (2018).
8. Wang, C., *et al.* Immune checkpoint inhibitor-associated myocarditis: a systematic analysis of case reports. *Front Immunol* **14**, 1275254 (2023).

***D. Thymoma Spontaneous Regression***

• *Thymoma did not regress*:

1. Harada, S., *et al.* A Case of Hypogamma-globulinemia with Thymoma (Good's Syndrome) Followed for 8 Years. *Jpn J Thorac Dis* **32**, 511-517 (1994).
2. Idogawa, M., Hinoda, Y., Hayashi, T., Ishida, T. & Imai, K. A Case of Thymoma and Hypogammaglobulinemia(Good's syndrome) with Lichen planus. *Jpn J Clin Immunol* **22**, 137-143 (1999).
3. Iorio, R., Evoli, A., Lauriola, L. & Batocchi, A.P. A B3 type-thymoma in a 7-year-old child with myasthenia gravis. *J Thorac Oncol* **7**, 937-938 (2012).
4. Kameyama, A., *et al.* Intrathoracic Recurrent Thymoma Exhibiting Rapid Progression After Resection. *Jpn J Lung Cancer* **55**, 42-47 (2015).
5. Kataoka, Y., *et al.* A case of thymoma with extensive necrosis. *J Jpn Assoc Chest Surg* **29**, 627-631 (2015).
6. Katsumata, Y., *et al.* Thymoma with Extensive Necrosis: A Case Report and Literature Review. *Surg Case Rep* **11**(2025).
7. Kitami, A., *et al.* Stage IV A thymoma associated with hypogammaglobulinemia. *J Jpn Assoc Chest Surg* **23**, 812-815 (2009).
8. Lee, N., *et al.* Invasive thymoma with chief complaints of a hemoptysis. *J Jpn Assoc Chest Surg* **5**, 481-486 (1991).
9. Li, X., Wang, M. & Sun, D. Sclerosing thymoma: A rare case report and brief review of literature. *Medicine (Baltimore)* **97**, e0520 (2018).
10. Oyamada, Y., *et al.* A case report of acquired hypogammaglobulinemia with thymoma (Good syndrome). *Jpn J Clin Immunol* **14**, 454-462 (1991).
11. Saito, R., Isogami, K., Fujimura, S. & Ookuda, K. OKT4 epitope deficiency in a patient with thymoma and hypogammaglobulinemia. *Jpn J Thorac Dis* **28**, 1120-1124 (1990).
12. Takada, J. & Inoue, Y. A Case of Cystic Thymoma with Extensive Necrosis Detected in an Asymptomatic Patient. *J Jpn Surg Assoc* **85**, 244-249 (2024).
13. Takasaki, C., Ishibashi, H., Fujiwara, N., Akashi, T. & Okubo, K. A case of thymoma with chest pain caused by bleeding necrosis. *J Jpn Assoc Chest Surg* **26**, 629-632 (2012).
14. Tsukahara, H., Miyauchi, Y., Kimura, N., Koizumi, R. & Hara, H. ３日間で急速に増大し広範な壊死を認めた胸腺 腫の１例 [A case of thymoma that rapidly enlarged over three days and showed extensive necrosis]. *J Jpn Surg Assoc* **84**, S418 (2023).
15. Uno, Y., Mizuguchi, T., Nakao, K. & Kosaka, M. 'Immunodeficiency with thymoma' associated with leukocytopenia. *Jpn J Clin Immunol* **15**, 177-183 (1992).
16. Zouvelou, V. When Thymomatous Myasthenia Gravis Is Not Grave. *J Clin Neuromuscul Dis* **21**, 124-125 (2019).

• *Other types of thymic tumor*:

1. Endo, K., Kato, M. & Sano, M. [Thymic Cancer Associated with Spontaneous Regression of Multilocular Thymic Cysts]. *Kyobu Geka* **75**, 617-621 (2022).
2. Fujiwara, T., Araki, K., Nishikawa, H., Kotani, K. & Matsuura, M. A case of thymic cancer with spontaneous regression detected on examination for thoracic injury. *J Jpn Assoc Chest Surg* **29**, 605-609 (2015).
3. Mizuno, K., Urabe, N. & Uematsu, S. A case of thymic cancer with spontaneous regression. *J Jpn Assoc Chest Surg* **30**, 550-554 (2016).
4. Naruke, Y., *et al.* A rare case of thymic carcinoid detected spontaneous regression. *J Jpn Assoc Chest Surg* **11**, 575-578 (1997).

5. Yamada, M., Abiko, M., Fuyama, S., Katoh, H. & Endoh, M. A case of thymic atypical carcinoid with spontaneous regression and metastasis within the thymus. *J Jpn Assoc Chest Surg* **31**, 103-108 (2017).

• _Patient received steroid, non-steroidal immuno-suppressant, or IVIG during regression period_:

1. Arai, H., *et al.* A cases of myasthenia gravis with a thymoma that disappeared following preoperative steroid therapy. *J Jpn Surg Assoc* **69**, 2189-2192 (2008).
2. Barratt, S., Puthucheary, Z.A. & Plummeridge, M. Complete regression of a thymoma to glucocorticoids, commenced for palliation of symptoms. *Eur J Cardiothorac Surg* **31**, 1142-1143 (2007).
3. Elaamadi, W., *et al.* Unexpected preoperative regression of a thymoma: a case report. *Adv Thorac Dis* **4**, 1-3 (2022).
4. Jiang, W. & Yu, Q. Case Report of Thymoma Tumor Reduction Following Plasmapheresis. *Medicine (Baltimore)* **94**, e2173 (2015).
5. Modrego, P.J. & Arribas, J. Spontaneous resolution of a mediastinal mass in a woman with myasthenia gravis. *Neurologia (Engl Ed)* **33**, 556-557 (2018).
6. Onuki, T., *et al.* Thymectomy during Myasthenic Crisis under Artificial Respiration. *Ann Thorac Cardiovasc Surg* **25**, 215-218 (2018).
7. Tombul, İ., *et al.* Preoperative spontaneous regression of type B2 thymoma in a patient with myasthenia gravis. *J Pulmonol Intens Care* **1**, 73-75 (2023).

• _Patient received unspecified treatment for MG_:

1. Kuo, T. Sclerosing thymoma--a possible phenomenon of regression. *Histopathology* **25**, 289-291 (1994).

• _Duplicate case_:

1. Furuya, K., *et al.* 経過中に自然縮小した胸腺腫の 1 例 [A case of thymoma that underwent spontaneous regression]. *Jpn J Lung Cancer* **54**, 166 (2014).
2. Hayakawa, M. & Tomita, E. 炎症所見を伴う急速増大後に自然縮小を示した胸腺腫の 1 例 [A case of thymoma showing spontaneous regression after rapid enlargement with inflammatory findings]. *J Jpn Assoc Chest Surg* **27**, S11 (P86-01) (2013).
3. Hori, D., Nakano, T., Tsubochi, H., Endo, S. & Sohara, Y. 自然縮小した胸腺腫の 1 手術 [Surgical case of a thymoma that underwent spontaneous regression]. in *日本胸部外科学会関東甲信越地方会要旨集*, Vol. 144 20 (2007).
4. Kasai, Y., *et al.* 短期間に増大と縮小を繰り返した胸腺腫の 1 例 [A case of thymoma that repeatedly enlarged and regressed over a short period]. *J Jpn Assoc Chest Surg* **24**, 613 (2010).
5. Tokunaga, T., *et al.* 経過中に自然縮小した壊死性胸腺腫の 1 例 [A case of necrotic thymoma that spontaneously regressed during its clinical course]. *Jpn J Lung Cancer* **64**, 961 (2024).
6. 成毛佳樹, *et al.* 自然縮小をみた胸腺カルチノイドの一例 [A case of thymic carcinoid tumor with spontaneous regression]. *J Jpn Assoc Chest Surg* **8**, 363 (D397) (1994).

### Appendix 4: Subclinical Inflammatory Myopathies in MG

Scientists have long acknowledged the frequent coexistence of a distinct, asymptomatic myopathic disorder in a subgroup of MG patients. EMG analyses reveal myopathic changes, including shortened mean motor unit potential (MUP) duration or increased polyphasic MUPs, in approximal 20% of MG patients, which were positively correlated with the presence of StrAbs.[1,2] The myopathy is not limited to electrophysiological abnormalities, as histopathological analyses show muscle alterations resembling those seen in IIM.

Many patients with MG exhibit focal collections of mononuclear cells, primarily composed of lymphocytes with a few macrophages, associated with muscle fiber necrosis and regeneration in skeletal and/or cardiac muscles, despite the absence of overt clinical IIM.[3,4] These inflammatory foci, known as lymphorrhages, occur most frequently in extraocular muscles, followed by the diaphragm and other skeletal muscles. They likely represent low-grade, chronic, multifocal muscle inflammation rather than a widespread muscle devastation.[5] Reported in 23–77% of MG patients depending on patient selection and sampling methodology, lymphorrhages are detected at particularly high frequencies in autopsy studies,[5-12] reflecting the greater sensitivity of post-mortem examination for identifying scattered focal lesions compared with a single, randomly targeted biopsy.[4,5] Lymphorrhage is frequently associated with thymoma but also occurs in non-thymomatous MG, albeit typically to a lesser extent.[3,5,13-19] A close correlation with positive StrAbs has likewise been documented.[14]

Despite their histological similarities to IIM, the infiltrated T lymphocytes observed in MG differ phenotypically from those in IIM. These cells express CD45RA, a marker typically associated with naïve T cells, even though they cluster around muscle fibers with MHC-I overexpression at inflammatory foci.[15] This pattern suggests that they may represent antigen-primed T cells that retain or reacquire a naïve-like phenotype. Notably, CD45RA is not an exclusive marker of naïve T cells, as a substantial subset of long-lived, antigen-experienced $CD8^+$ and $CD4^+$ memory T cells can re-express CD45RA, a phenotype that can persist for years after antigen exposure.[20,21] These infiltrating cells lose CD45RA expression when patients later develop clinically overt myositis, although the mechanisms underlying this phenotypic transition remain unknown.[15,22]

Signs of cardiac disease without an identifiable cardiovascular etiology have been reported in 16% MG patients, with much higher frequencies among those with thymoma.[17,23] In these individuals, focal cellular infiltrates have been identified in both cardiac and skeletal muscle. Collectively, these findings support the presence of a subclinical form of StrAb-associated myositis in a subset of MG patients, especially those with a thymoma. Such smoldering myositis likely contributes, at least in part, to the more severe myasthenic symptoms and poorer prognosis observed in thymoma-associated MG.[24]

Although subclinical StrAb-associated myositis is observed in a substantial proportion of MG patients, it rarely progresses to overt disease.[22,25,26] Most individuals remain asymptomatic for much of their lives, suggesting the existence of protective mechanisms that restrain muscle inflammation. In skeletal muscle biopsies of patients with concurrent MG and StrAb-associated myositis, infiltrated

CD8+ T cells overexpress PD-1 and often cluster near necrotic myofibers.[22] In response to inflammation, muscle fibers not only overexpress MHC molecules but also upregulate PD-L1, frequently adjacent to PD-1+ T cells. Some inflammatory cells in endomysial infiltrates also express CTLA-4. However, PD-L1 expression on muscle fibers in MG has not been systematically studied.

Evidence from murine models indicates the PD-1 pathway plays a major role in protecting cardiac muscle from T cell–mediated injury.[27,28] By analogy, in patients with subclinical StrAb-associated myositis, both PD-1 and CTLA-4 pathways likely mitigate autoimmune damage and maintain peripheral tolerance by exhausting striated-muscle-specific CD8+ T cells and inhibiting their proliferation. Given the strong association between StrAb-associated myositis and thymoma, this hypothesis predicts that down-regulation of these pathways is highly likely to provoke flares of previously silent myositis in thymoma patients. Consistent with this, individuals with TETs treated with ICIs exhibit a 10- to 30-fold higher risk of developing myositis or myocarditis than those with other cancers.[29] Moreover, patients with thymoma are significantly more likely to develop ICI-induced myositis or myocarditis than those with thymic carcinoma. The presence of anti-AChR antibodies is also associated with ICI-induced myositis in both thymoma and non-thymoma malignancies.[29,30] Together, these observations support the crucial protective role of the PD-1 and CTLA-4 pathways in restraining StrAb-associated myositis.

**References**

1. Somnier, F.E., Skeie, G.O., Aarli, J.A. & Trojaborg, W. EMG evidence of myopathy and the occurrence of titin autoantibodies in patients with myasthenia gravis. *Eur J Neurol* **6**, 555-563 (1999).
2. Somnier, F.E. & Trojaborg, W. Neurophysiological evaluation in myasthenia gravis. A comprehensive study of a complete patient population. *Electroencephalogr Clin Neurophysiol* **89**, 73-87 (1993).
3. Russell, D.S. Histological changes in the striped muscles in myasthenia gravis. *J Pathol Bacteriol* **65**, 279-289 (1953).
4. Buzzard, E.F. The clinical history and postmortem examination of five cases of myasthenia gravis. *Brain* **28**, 438-483 (1905).
5. Araki, S. [Muscle lesions in myasthenia gravis]. *Shinkei Kenkyu No Shimpo* **14**, 492-493 (1970).
6. Engel, W.K. & McFarlin, D.E. Muscle lesions in myasthenia gravis. Discussion. *Ann N Y Acad Sci* **135**, 68-78 (1966).
7. Fenichel, G.M. Muscle lesions in myasthenia gravis. *Ann N Y Acad Sci* **135**, 60-67 (1966).
8. Fenichel, G.M. & Shy, G.M. Muscle Biopsy Experience in Myasthenia Gravis. *Arch Neurol* **9**, 237-243 (1963).
9. Werneck, L.C. [The muscular lesion in myasthenia gravis: study of 17 cases with muscular histochemistry]. *Arq Neuropsiquiatr* **40**, 67-76 (1982).
10. Nakano, S. & Engel, A.G. Myasthenia gravis: quantitative immunocytochemical analysis of inflammatory cells and detection of complement membrane attack complex at the end-plate in 30 patients. *Neurology* **43**, 1167-1172 (1993).
11. Johns, T.R., Crowley, W.J., Miller, J.Q. & Campa, J.F. The syndrome of myasthenia and polymyositis with comments on therapy. *Ann N Y Acad Sci* **183**, 64-71 (1971).
12. Rowland, L.P., Hoefer, P.F., Aranow, H., Jr. & Merritt, H.H. Fatalities in myasthenia gravis; a review of 39 cases with 26 autopsies. *Neurology* **6**, 307-326 (1956).

13. Zamecnik, J., *et al.* Muscle lymphocytic infiltrates in thymoma-associated myasthenia gravis are phenotypically different from those in polymyositis. *Neuromuscul Disord* **17**, 935-942 (2007).
14. Long, H.M., *et al.* MHC II tetramers visualize human CD4+ T cell responses to Epstein-Barr virus infection and demonstrate atypical kinetics of the nuclear antigen EBNA1 response. *J Exp Med* **210**, 933-949 (2013).
15. Miller, J.D., *et al.* Human effector and memory CD8+ T cell responses to smallpox and yellow fever vaccines. *Immunity* **28**, 710-722 (2008).
16. Uchio, N., *et al.* Inflammatory myopathy with myasthenia gravis: Thymoma association and polymyositis pathology. *Neurol Neuroimmunol Neuroinflamm* **6**, e535 (2019).
17. Genkins, G., Mendelow, H., Zobel, H.J. & Osserman, K.E. Myasthenia Gravis Analysis of 31 Consecutive Post-Mortem Examinations. in *Myasthenia Gravis: The Second International Symposium* (eds. Viets, H.R. & Schwab, R.S.) 519-530 (Charles C Thomas, Springfield, IL, 1959).
18. Oosterhuis, H.J., Bethlem, J. & Feltkamp, T.E. Muscle pathology, thymoma, and immunological abnormalities in patients with myasthenia gravis. *J Neurol Neurosurg Psychiatry* **31**, 460-463 (1968).
19. Oosterhuis, H. & Bethlem, J. Neurogenic muscle involvement in myasthenia gravis. A clinical and histopathological study. *J Neurol Neurosurg Psychiatry* **36**, 244-254 (1973).
20. Mendelow, H. & Genkins, G. Studies in myasthenia gravis: cardiac and associated pathology. *J Mt Sinai Hosp N Y* **21**, 218-225 (1954).
21. Norris, E.H. The thymoma and thymic hyperplasia in myasthenia gravis with observations on the general pathology. *Am. J. Cancer* **27**, 421–433 (1936).
22. Brem, J. & Wechsler, H.F. Myasthenia gravis associated with thymoma: report of two cases with autopsy. *Arch. Int. Med.* **54**, 901-915 (1934).
23. Hofstad, H., Ohm, O.J., Mork, S.J. & Aarli, J.A. Heart disease in myasthenia gravis. *Acta Neurol Scand* **70**, 176-184 (1984).
24. Alvarez-Velasco, R., *et al.* Clinical characteristics and outcomes of thymoma-associated myasthenia gravis. *Eur J Neurol* **28**, 2083-2091 (2021).
25. Garibaldi, M., *et al.* Muscle involvement in myasthenia gravis: Expanding the clinical spectrum of Myasthenia-Myositis association from a large cohort of patients. *Autoimmun Rev* **19**, 102498 (2020).
26. Suzuki, S., *et al.* Autoimmune targets of heart and skeletal muscles in myasthenia gravis. *Arch Neurol* **66**, 1334-1338 (2009).
27. Tarrio, M.L., Grabie, N., Bu, D.X., Sharpe, A.H. & Lichtman, A.H. PD-1 protects against inflammation and myocyte damage in T cell-mediated myocarditis. *J Immunol* **188**, 4876-4884 (2012).
28. Wang, J., *et al.* PD-1 deficiency results in the development of fatal myocarditis in MRL mice. *Int Immunol* **22**, 443-452 (2010).
29. Fenioux, C., *et al.* Thymus alterations and susceptibility to immune checkpoint inhibitor myocarditis. *Nat Med* **29**, 3100-3110 (2023).
30. Mammen, A.L., *et al.* Pre-existing antiacetylcholine receptor autoantibodies and B cell lymphopaenia are associated with the development of myositis in patients with thymoma treated with avelumab, an immune checkpoint inhibitor targeting programmed death-ligand 1. *Ann Rheum Dis* **78**, 150-152 (2019).

### Appendix 5: Thymoma Spontaneous Regression

The strong association between StrAb-associated MG-myositis overlap syndrome and thymoma hints a key etiological role for thymoma in this condition. However, as shown in Table 1, 40% of patients with the overlap syndrome had no detectable thymoma. Consistent with this, lymphorrhages and StrAbs have also been reported in a subset of nonthymomatous MG patients.[1-5] These StrAb-positive individuals exhibited genetic and cellular features closely resembling those seen in thymoma-associated MG, including shared allelic variants at the TNF, FCGR2A, and IL10 loci,[6,7] and an expansion of naïve-like $CD8^+CD45RA^+$ T cells within the T-cell receptor (TCR) Vβ subset.[8] These parallels suggest that StrAb-associated myositis can arise even in the absence of thymoma.

However, it remains theoretically possible that some nonthymomatous patients with StrAb-positive overlap syndrome may have already rejected occult thymomas prior to diagnosis. Extensive necrotic changes in thymomas is well documented,[9,10] with 4-25% showing hemorrhagic, cystic, or necrotic alterations.[11,12] In contrast to other neoplasms, in which these histopathologic features often indicate aggressive or malignant behavior, thymomas with these changes are usually early stage and tend to follow a benign clinical course. Furthermore, many necrotic thymomas have been reported to undergo spontaneous regression. To characterize thymomas exhibiting spontaneous regression, we performed a literature search of the PubMed, J-STAGE, and J-Global databases from inception to July 23, 2025 (Supplementary Fig. 1). A total of 213 reports were identified, of which 29 duplicates were removed. After title and abstract screening, 66 articles were selected for full-text review, and 16 additional articles were identified through reference screening. Following full-text assessment, 47 articles comprising 56 unique patients met inclusion criteria and were included in the final analysis (Supplementary Appendix 2F).

Among these cases, 41% were female and 59% were male, with a median age of 46 years, which is younger than the median age reported in large cohort studies (Supplementary Table 6).[13,14] Local symptoms, such as fever, cough, chest pain, or back pain, led to thymoma detection in 78% of patients. The remaining cases were detected incidentally on chest radiographs obtained during routine exams or for unrelated evaluations, including for MG. In this cohort, initial tumor sizes tended to be larger than those typically reported in the general thymoma population, with a mean size of 6.7 cm in maximum diameter, which may explain the higher frequency of local symptoms.[15] In line with this, asymptomatic cases, accounting for 14% of the cohort, had significantly smaller tumors than symptomatic cases (mean ± SD: 4.9 ± 1.9 cm versus 7.0 ± 2.6 cm, $p$=0.03). It is therefore possible that additional asymptomatic thymomas undergoing spontaneous regression, especially small tumors, were not recognized.

Histological subtypes of the resected tumors were classified as WHO type B in 88% of cases, with half of these further categorized as B2, a subtype strongly associated with MG.[16,17] Despite the presence of aggressive histologic types and relatively large tumor sizes, 92% of thymomas exhibiting spontaneous regression were early-stage tumors (Masaoka stage I or II). Follow-up information was available for 29 patients, with a median follow-up of 12 months (IQR, 7.5-24). Only one patient experienced recurrence. Clinical MG developed in 11% of cases, either concurrently or subsequently. Anti-AChR antibody testing was reported in 19 patients, of whom 7 (37%) tested positive; among these, 3 had concomitant MG. These findings demonstrate that spontaneous thymoma regression can occur in patients with thymomatous MG.

Extensive areas of necrosis were present in 84% of thymomas, while hemorrhage, cyst, and fibrosis were identified in 21%, 36%, and 34% of cases, respectively. Spontaneous thymoma regression has been attributed to tumor necrosis, although the underlying cause remains unclear. Circulatory disturbances and ischemic necrosis have been proposed as potential contributing factors, with regression occurring as necrotic and hemorrhagic components are resorbed. In some cases, the residual necrotic thymoma was microscopic and undetectable on radiologic imaging.[18,19] The estimated mean tumor halving-time was 0.46 month (Supplementary Fig. 2), leaving only a narrow window in which rapidly regressing thymomas can be detected before complete disappearance. This limited detection period likely contributes to the rarity of reported cases in the literature.

Alternatively, some patients initially classified as nonthymomatous may have harbored microscopic thymomas that were not detectable on computed tomography. Such lesions may have escaped post-operative histological evaluation due to sampling limitations, or they may have regressed in patients who did not undergo thymectomy. In the latter scenario, regression may occur spontaneously or in response to corticosteroid therapy administered for MG,[19,20] as corticosteroids are known to induce thymoma regression in some cases.[21] Supporting this hypothesis, thymomas have occasionally been detected years after an initial diagnosis of nonthymomatous MG in both thymectomized and non-thymectomized patients.[22-29] In most such cases, available data are insufficient to determine whether these tumors represented local recurrence, dissemination, or new primary tumors arising from thymic remnants or ectopic thymic tissue. However, positive StrAb titers at the time of MG diagnosis have been reported in some patients, suggesting the possible presence of an occult microscopic thymoma at disease onset.[22,30] Collectively, these observations highlight the potential underdiagnosis of thymoma in MG patients, particularly those who are StrAb-positive.

**References**

1. Genkins, G., Mendelow, H., Zobel, H.J. & Osserman, K.E. Myasthenia Gravis Analysis of 31 Consecutive Post-Mortem Examinations. in *Myasthenia Gravis: The Second International Symposium* (eds. Viets, H.R. & Schwab, R.S.) 519-530 (Charles C Thomas, Springfield, IL, 1959).
2. Oosterhuis, H. & Bethlem, J. Neurogenic muscle involvement in myasthenia gravis. A clinical and histopathological study. *J Neurol Neurosurg Psychiatry* **36**, 244-254 (1973).
3. Oosterhuis, H.J., Bethlem, J. & Feltkamp, T.E. Muscle pathology, thymoma, and immunological abnormalities in patients with myasthenia gravis. *J Neurol Neurosurg Psychiatry* **31**, 460-463 (1968).
4. Romi, F., Skeie, G.O., Aarli, J.A. & Gilhus, N.E. Muscle autoantibodies in subgroups of myasthenia gravis patients. *J Neurol* **247**, 369-375 (2000).
5. Yamamoto, A.M., *et al.* Anti-titin antibodies in myasthenia gravis: tight association with thymoma and heterogeneity of nonthymoma patients. *Arch Neurol* **58**, 885-890 (2001).
6. Alseth, E.H., Nakkestad, H.L., Aarseth, J., Gilhus, N.E. & Skeie, G.O. Interleukin-10 promoter polymorphisms in myasthenia gravis. *J Neuroimmunol* **210**, 63-66 (2009).

7. Amdahl, C., Alseth, E.H., Gilhus, N.E., Nakkestad, H.L. & Skeie, G.O. Polygenic disease associations in thymomatous myasthenia gravis. *Arch Neurol* **64**, 1729-1733 (2007).
8. Tackenberg, B.*, et al.* Expanded TCR Vbeta subsets of CD8(+) T-cells in late-onset myasthenia gravis: novel parallels with thymoma patients. *J Neuroimmunol* **216**, 85-91 (2009).
9. Katsumata, Y.*, et al.* Thymoma with Extensive Necrosis: A Case Report and Literature Review. *Surg Case Rep* **11**(2025).
10. Tokunaga, T., Ose, N., Nagata, H., Morii, E. & Shintani, Y. Necrotic Thymoma Discovered Due to Subjective Symptoms: A Report of Three Cases. *Surg Case Rep* **11**(2025).
11. Moran, C.A. & Suster, S. Thymoma with prominent cystic and hemorrhagic changes and areas of necrosis and infarction: a clinicopathologic study of 25 cases. *Am J Surg Pathol* **25**, 1086-1090 (2001).
12. Rosai, J. & Levine, G.D. Thymoma. Tumors of the thymus. in *Atlas of Tumor Pathology, Fasc. 13, ser. 2* (ed. Firminger, H.I.) 43-49 (Armed Forces Institute of Pathology, Washington, 1976).
13. Bian, D.*, et al.* Thymoma size significantly affects the survival, metastasis and effectiveness of adjuvant therapies: a population based study. *Oncotarget* **9**, 12273-12283 (2018).
14. Okumura, M.*, et al.* Tumour size determines both recurrence-free survival and disease-specific survival after surgical treatment for thymoma. *Eur J Cardiothorac Surg* **56**, 174-181 (2019).
15. Moran, C.A. & Suster, S. "Ancient" (sclerosing) thymomas: a clinicopathologic study of 10 cases. *Am J Clin Pathol* **121**, 867-871 (2004).
16. Yasumizu, Y.*, et al.* Myasthenia gravis-specific aberrant neuromuscular gene expression by medullary thymic epithelial cells in thymoma. *Nat Commun* **13**, 4230 (2022).
17. Okumura, M.*, et al.* The World Health Organization histologic classification system reflects the oncologic behavior of thymoma: a clinical study of 273 patients. *Cancer* **94**, 624-632 (2002).
18. Huang, T.W.*, et al.* Spontaneous regression of a mediastinal thymoma. *J Thorac Cardiovasc Surg* **137**, 1277-1278 (2009).
19. Kuo, T. Sclerosing thymoma--a possible phenomenon of regression. *Histopathology* **25**, 289-291 (1994).
20. Modrego, P.J. & Arribas, J. Spontaneous resolution of a mediastinal mass in a woman with myasthenia gravis. *Neurologia (Engl Ed)* **33**, 556-557 (2018).
21. Kobayashi, Y.*, et al.* Preoperative steroid pulse therapy for invasive thymoma: clinical experience and mechanism of action. *Cancer* **106**, 1901-1907 (2006).
22. Husain, F., Ryan, N.J., Hogan, G.R. & Gonzalez, E. Occurrence of invasive thymoma after thymectomy for myasthenia gravis: report of a case. *Neurology* **40**, 170-171 (1990).
23. Santos, E.*, et al.* Signs heralding appearance of thymomas after extended thymectomy for myasthenia gravis. *Neurol Clin Pract* **9**, 48-52 (2019).
24. Toker, A., Tanju, S., Ozluk, Y. & Serdaroglu, P. Thymoma appearing 10 years after an extended thymectomy for myasthenia gravis. *Eur J Cardiothorac Surg* **33**, 1155-1156 (2008).
25. Hirabayashi, H., Ohta, M., Okumura, M. & Matsuda, H. Appearance of thymoma 15 years after extended thymectomy for myasthenia gravis without thymoma. *Eur J Cardiothorac Surg* **22**, 479-481 (2002).
26. Zouvelou, V., Velonakis, G., Rontogianni, D. & Zisis, C. Appearance of thymoma 5 years after thymectomy for nonthymomatous myasthenia gravis. *J Clin Neuromuscul Dis* **16**, 42-43 (2014).
27. Sugawara, M.*, et al.* Long-term follow up of thymus in patients with myasthenia gravis. *J Neuroimmunol* **221**, 121-124 (2010).

28. Hengstman, G.J., Drost, G., Wagenaar, M. & van Engelen, B.G. Persistent increased risk for thymoma in myasthenia gravis associated with myositis. *Muscle Nerve* **34**, 251-252 (2006).
29. Reznik, M. [Two cases of myasthenic syndrome with thymoma, polymyositis, myocarditis, and thyroiditis]. *J Neurol Sci* **22**, 341-351 (1974).
30. van de Warrenburg, B.P., *et al*. Concomitant dermatomyositis and myasthenia gravis presenting with respiratory insufficiency. *Muscle Nerve* **25**, 293-296 (2002).

### Appendix 6: Supplementary Tables and Figures

**Supplementary Table 1.** Comparison between 31 cases from three single-center studies and 175 cases from case reports and case series.

| Characteristic | Single-center studies (n = 31) | Case reports/series (n = 175) | p-value |
|---|---|---|---|
| Male, % (SD) | 35.5 (8.6) | 39.4 (3.7) | 0.69 |
| Age at MG onset, mean (SD) (yrs) | 53.0 (13.1) | 50.0 (17.9) | 0.37 |
| Age at myositis onset, mean (SD) (yrs) | 56.2 (12.1) | 51.7 (17.8) | 0.18 |
| EOMG (<50 years old), % (SD) | 41.9 (8.9) | 47.7 (3.8) | 0.55 |
| Anti-AChR-Positive (after 1979) [a], % (SD) | 96.8 (3.2) | 94.8 (1.9) | 0.70 |
| Occurrence of TET, % (SD) | 72.4 (8.3) | 55.2 (3.9) | 0.09 |
| Cardiac Involvement, % (SD) | 19.4 (7.1) | 24.6 (3.3) | 0.53 |
| Simultaneous presence of MG & IIM, % (SD) | 55.2 (9.2) | 72.4 (3.5) | 0.07 |
| Subsequent Presence of IIM, % (SD) | 37.9 (9.0) | 27.0 (3.5) | 0.18 |

[a] Anti-AChR antibody test was widely used as the diagnostic marker for MG since 1980s.

**Supplementary Table 2.** Detection of StrAbs in classic myositis

| Serial | Case # | Striational Antigen | Striational Antibodies (+/-) | Detection Method [a] | Reference [b] |
|---|---|---|---|---|---|
| 1 | 35 | Beef skeletal muscle | 0/35 | IFA | Strauss et al. 1965 |
| 2 | 20 | Unspecified | 2/20 | IFA | Wada et al. 1983 |
| 3 | 13 | Rat skeletal muscle & human muscle extract | 0/13 | IFA & ELISA | Cikes et al. 1988 |
| 4 | 2 | Human cardiac muscle | 0/2 | ELISA | Mygland et al. 1991 |
| 5 | 2 | Ryr | 0/2 | Western blot | Mygland et al. 1992 |
| 6 | 2 | Titin | 0/2 | ELISA | Gautel et al. 1993 |
| 7 | Unspecified | Titin | Undetected | ELISA | Somnier et al. 1999 |
| 8 | 30 | Kv1.4 | 0/30 | RIPA | Suzuki et al. 2005 |
| 9 | 18 | Titin | 0/18 | RIPA | Stergiou et al. 2016 |
| 10 [c] | 60 | Titin<br>Ryr<br>Kv1.4 | 0/60<br>Unspecified<br>7/60 | Cytometric cell-based assay | Kufukihara et al. 2019 |

[a] IFA, indirect immunofluorescence assay; ELISA, enzyme-linked immunosorbent assay; RIPA, radioimmunoprecipitation assay.

[b] Relevant references are provided in Supplementary Appendix 2B.

[c] The authors reported that 61% of the disease control patients (including 30 with anti-signal recognition particle IMNM, 30 with IBM, and 30 with Duchenne muscular dystrophy) and 23% of 30 healthy controls tested positive for anti-RyR using a cytometric cell-based assay. This contrasts with results obtained by ELISA, in which none of the 90 healthy controls or 10 patients with muscular dystrophy reacted with RyR.[5] These discrepancies may reflect false-positive results due to the limited specificity of the cytometric cell-based assay. Similarly, anti-Kv1.4 antibodies were weakly positive in 12% of patients with idiopathic inflammatory myopathies (IIM), 7% with muscular dystrophy, and 3% of healthy controls, suggesting the possibility of nonspecific serum antibody binding.

**Supplementary Table 3.** Characteristics and outcomes of patients with IIM associated with TETs without concurrent MG (N = 48)[a]

| Characteristic | Thymic carcinoma | Thymoma | Adj-OR (95% CI) [b] | *p* value |
|---|---|---|---|---|
| **Total – N** | 16 | 32 | | |
| **Female – n (%)** | 5 (31) | 13 (42) | 1.7 (0.5-6.4) | 0.44 |
| **Age at IIM Onset – median (IQR), years** | 58 (46-66) | 53 (43-64) | n/a | 0.86 |
| **IIM Subtypes – n (%)** | | | n/a | <0.0001 |
| DM | 11 (69) | 3 (9) | | |
| GCM | 1 (6) | 13 (41) | | |
| PM | 4 (25) | 16 (50) | | |
| **Pathological Features – n/N (%)** | | | | |
| MG-like ocular symptoms | 1/16 (6) | 5/32 (16) | 2.6 (0.3-25.2) | 0.41 |
| Elevated CK levels | 12/14 (86) | 17/18 (94) | 3.4 (0.3-45.7) | 0.35 |
| Peak CK level – median (IQR), U/L | 1379 (273-7906) | 846 (363-3212) | n/a | 0.90 |
| Cardiac involvement | 0/16 (0) | 20/32 (63) | n/a | n/a |
| **Autoantibodies – n/N (%)** | | | | |
| Anti-AChR$^+$ – n/N (%) | 0/1 (0) | 0/7 (0) | n/a | n/a |
| StrAb$^+$ – n/N (%) | n/a | 5/7 (71) | n/a | n/a |
| MSA$^+$ – n/N (%) | 5/8 (63) | 0/4 (0) | n/a | n/a |
| **Treatment – n/N (%)** | | | | |
| Corticosteroids | 11/16 (69) | 23/32 (72) | 1.1 (0.3-4.7) | 0.91 |
| Immunomodulators | 5/16 (31) | 9/32 (28) | 0.9 (0.2-3.7) | 0.85 |
| Respiratory support | 2/16 (25) | 11/32 (34) | 3.7 (0.7-20.0) | 0.13 |
| Cardiovascular interventions | 2/16 (12) | 13/32 (41) | 4.7 (0.9-24.6) | 0.07 |
| **Outcome – n/N (%)** | | | n/a | 0.0004 |
| Remission | 3/15 (20) | 0/32 (0) | | |
| Pharmacological Remission | 3/15 (20) | 3/32 (9) | | |
| Minimal Manifestations | 8/15 (53) | 10/32 (31) | | |
| Death | 1/15 (7) | 19/32 (59) | | |

[a] A case in which the thymic tumor had not been classified by histopathology was excluded. In addition, we identified five cases of isolated myositis associated with the presence of StrAbs, without concurrent TETs or a confirmed diagnosis of MG, which were excluded from this analysis (Supplementary Appendix 2D).

[b] ORs and 95% CIs were adjusted for sex and age, with corresponding *p*-values reported.

**Supplementary Table 4.** Patient characteristics of the ICI-induced myocarditis reported in our dataset versus in Fenioux et al. 2023.

| | This study | | Fenioux et al. 2023 | | *p* value[a] |
|---|---|---|---|---|---|
| | **TET** | **Other cancers** | **TET** | **Other cancers** | |
| **Total** | 25 | 275 | 28 | 767 | |
| **Female – n/N (%)** | 14/25 (56) | 98/272 (36) | 14/27 (52) | 252/764 (33) | 0.99 |
| **Age at onset – Median (IQR), years** | 48 (45-58) | 69 (63-75) | 53 (43-64) | 70 (62-77) | n/a |
| **Prior history of MG – n (%)** | 10 (40) | 3 (1) | 2 (8) | 3 (0.4) | 0.44 |
| **ICI treatment – n/N (%)** | | | | | |
| Anti-PD-1/PD-L1 | 25/25 (100) | 202/275 (73) | 26/27 (96) | 587/759 (77) | 0.62 |
| Anti-CTLA-4 | 0/25 (0) | 9/275 (3) | 0/27 (0) | 13/759 (2) | 0.97 |
| Anti-CTLA-4 and PD-1/PD-L1 | 0/25 (0) | 63/275 (23) | 1/27 (4) | 155/759 (20) | 0.65 |
| **Associated irAEs – n/N (%)** | | | | | |
| MG-like syndrome | 20/25 (80) | 76/275 (28) | 18/28 (64) | 157/767 (20) | 0.47 |
| Myositis/rhabdomyolysis | 19/25 (76) | 155/275 (56) | 21/28 (75) | 280/767 (37) | 0.15 |
| **Myocarditis** | | | | | |
| Diagnostic certainty – n/N (%) | | | | | |
| Definite | 9/24 (38) | 149/273 (55) | 9/22 (41) | 221/696 (32) | 0.08 |
| Probable | 1/24 (4) | 26/273 (10) | 4/22 (18) | 262/696 (38) | 0.83 |
| Possible | 14/24 (58) | 98/273 (36) | 7/22 (32) | 204/696 (29) | 0.20 |
| Peak troponin – Median (IQR) [b] | 290 (132-452) | 99 (23-240) | 262 (66-399) | 36 (8-117) | n/a |
| Peak CK – Median (IQR) [b] | 45 (17-68) | 15 (6-37) | 24 (9-56) | 3 (1-13) | n/a |
| **Treatment and outcomes – n/N (%)** | | | | | |
| Corticosteroids | 25/25 (100) | 263/275 (96) | 27/28 (96) | 625/767 (82) | 0.92 |
| Abatacept | 1/25 (4) | 11/275 (4) | 5/28 (18) | 82/767 (11) | 0.55 |
| Intravenous immunoglobulin | 17/25 (68) | 96/275 (35) | 13/28 (46) | 71/767 (9) | 0.0009 |
| Plasmapheresis | 6/25 (24) | 48/275 (17) | 7/28 (25) | 82/767 (11) | 0.36 |
| irAE-related death | 11/25 (44) | 91/274 (33) | 10/28 (36) | 122/738 (17) | 0.20 |

[a] The Breslow–Day test was used to evaluate the homogeneity of odds ratios in a 2 × 2 × 2 contingency table. The analysis was performed using the DescTools package in R. The test was not adjusted for age or sex, as the dataset from Fenioux et al. (2023) was not available for stratified analysis.

[b] Both the peak troponin level and peak creatine kinase level have been normalized to the upper normal limit.

**Supplementary Table 5.** Differentiation of ICI-induced DM from Other Myositis-Spectrum Conditions (N = 462)

| | ICI-DM | ICI-PM [a] | Adj-OR (95% CI) [b] | *p* value |
|---|---|---|---|---|
| **Female – n/N (%)** | 8/30 (27) | 154/428 (36) | 1.5 (0.7-3.6) | 0.33 |
| **Age at Onset – median (IQR), years** | 70 (57-74) | 69 (60-76) | n/a | 0.77 |
| **Cancer Type – n/N (%)** [c] | | | n/a | 0.02 |
| TET | 0/30 (0) | 28/431 (6) | | |
| Lung cancer | 14/30 (47) | 109/431 (25) | | |
| Melanoma | 7/30 (23) | 112/431 (26) | | |
| Other cancer | 9/30 (30) | 182/431 (42) | | |
| **ICI treatment – n/N (%)** [d] | | | n/a | 0.46 |
| Anti-PD-1/PD-L1 | 24/30 (80) | 331/430 (77) | | |
| Anti-CTLA-4 | 2/30 (7) | 15/430 (3) | | |
| Anti-CTLA-4 and PD-1/PD-L1 | 4/30 (13) | 84/430 (20) | | |
| **Time to onset of irAEs –mean ± SD, days** | 33.3 ± 39.1 | 48.9 ± 92.5 | n/a | 0.40 |
| **Exacerbation of pre-existing IIM – n/N (%)** | 8/30 (27) | 4/432 (1) | 0.02 (0-0.1) | <0.0001 |
| **Pathological Features – n/N (%)** | | | | |
| MG-like ocular symptoms | 2/30 (7) | 193/430 (45) | 10.9 (2.6-46.8) | 0.001 |
| Concurrent MG-like syndrome | 2/30 (7) | 161/432 (37) | 8.0 (1.9-34.3) | 0.005 |
| Concurrent myocarditis | 1/30 (3) | 299/432 (69) | 64.0(8.6-475.9) | <0.0001 |
| Peak CK level – median (IQR), U/L | 1571 (504-4145) | 3056 (1284-7311) | n/a | 0.14 |
| **Autoantibodies – n/N (%)** | | | | |
| Anti-AChR$^{+}$ | 1/7 (14) | 83/203 (41) | 4.2 (0.5-35.7) | 0.19 |
| MSA$^{+}$ | 19/26 (73) | 7/123 (6) | 0.01 (0-0.1) | <0.0001 |
| StrAb$^{+}$ | 0/1 (0) | 60/83 (72) | n/a | n/a |
| **Outcome – n/N (%)** | | | n/a | 0.12 |
| Remission | 2/30 (7) | 48/428 (11) | | |
| Pharmacological remission | 5/30 (17) | 55/428 (13) | | |
| Minimal manifestations | 17/30 (57) | 188/428 (44) | | |
| No improvement | 2/30 (7) | 6/428 (1) | | |
| irAE-related death & hospice care [e] | 4/30 (13) | 131/428 (31) | | |

[a] In this subgroup, 22 cases were identified as IMNM based on characteristic histopathological features. However, unlike typical IMNM, half of these cases were associated with MG-like syndrome, of which 64% had a confirmed diagnosis of MG. Conventional MSAs were detected in only one case, while 78% of tested cases were positive for StrAbs. We have previously shown that StrAb-associated myositis can be misdiagnosed as seronegative IMNM owing to overlapping histopathological characteristics (Table 2). Therefore, these cases were consolidated with the broader group for subsequent analysis.

[b] ORs and 95% CIs were adjusted for sex and age, with corresponding *p*-values reported.

[c] TET: thymoma and thymic carcinoma; lung cancer: non-small cell lung cancer, small-cell lung cancer, and neuroendocrine lung cancer; other cancers include: bladder cancer, prostate cancer, renal cancer, renal pelvic cancer, upper tract urothelial cancer, urachal carcer; ureter cancer, urinary tract cancer; urothelial cancer, bile duct cancer, colorectal cancer, gastric cancer, esophageal cancer, esophagogastric junction cancer, liver cancer, lower esophageal and gastric cardia cancer, rectal cancer, pancreatic cancer; rectal endocrine cancer, sarcoma, mesothelioma, Hodgkin lymphoma, non-Hodgkin lymphoma, brain cancer, breast cancer, chordoma, eye cancer, head and neck cancer, endometrial cancer, vaginal cancer, and ovarian cancer.

[d] PD-1, programmed cell death 1; PD-L1, programmed cell death-ligand 1; CTLA-4, cytotoxic T-lymphocyte-associated protein 4.

[e] Patients whose deaths were attributed to cancer progression were excluded from the 'death and hospice care' outcome category and instead classified according to the prognosis of their irAEs prior to death. Eight patients who were

transitioned to hospice care and subsequently lost to follow-up were consolidated into the 'death and hospice care' category, which was used to represent the worst observed outcome in our cohort.

**Supplementary Table 6.** Clinical features and prognosis of patients with spontaneously regressed thymoma (N = 56).

| | |
|---|---|
| **Age – Median (IQR), years** | 46 (32-58) |
| **Female – n/N (%)** | 23/56 (41) |
| **Symptomatic finding – n/N (%)** | 43/55 (78) |
| **Pleural effusion – n/N (%)** | 36/56 (64) |
| **Initial tumor size – Mean ± SD, cm** | 6.7 ± 2.6 |
| **WHO subtype – n/N (%)** | |
| A/AB | 6/51 (12) |
| B1 | 13/51 (25) |
| B2 | 23/51 (45) |
| B3 | 9/51 (18) |
| **Masaoka Stage – n/N (%)** | |
| Stage I | 19/36 (53) |
| Stage II | 14/36 (39) |
| Stage III | 3/36 (8) |
| **Histopathological features – n/N (%)** | |
| Hemorrhage | 12/56 (21) |
| Cyst | 20/56 (36) |
| Necrosis | 47/56 (84) |
| Fibrosis | 19/56 (34) |
| **MG – n/N (%)** | |
| MG symptoms | 6/56 (11) |
| Anti-AChR$^+$ | 7/19 (37) |
| **Follow-up – Median (IQR), months** | 12.0 (7.5-24.0) |
| **Recurrence/metastasis – n/N (%)** | 1/29 (3) |

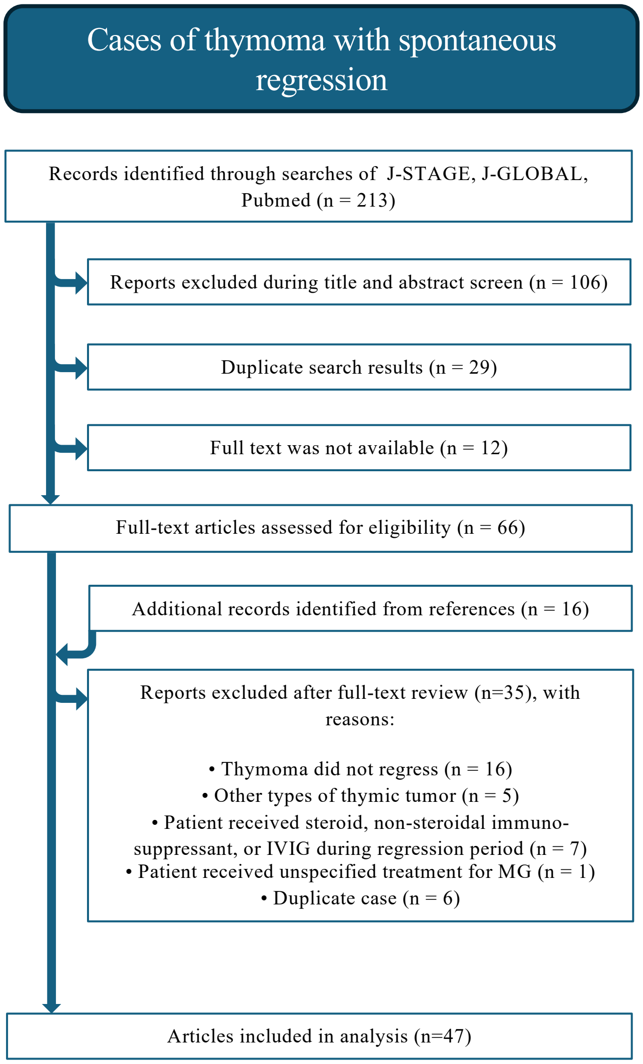

**Supplementary Figure 1: Flowchart illustrating the literature search and selection process for reports of spontaneous regression of thymoma.**

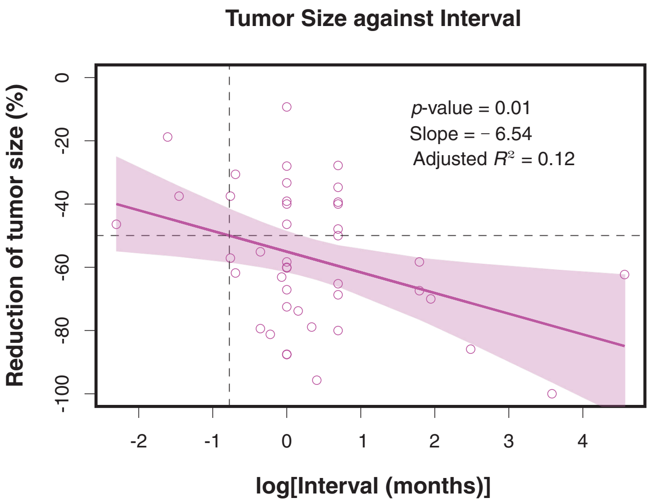

**Supplementary Figure 2: Scatter plot with linear regression and 95% confidence interval showing the association between tumor size reduction and the interval between imaging.**
